# Investigating SARS-CoV-2 infection and the health and psychosocial impact of the COVID-19 pandemic in the Canadian CHILD Cohort: study methodology and cohort profile

**DOI:** 10.1101/2022.10.19.22281242

**Authors:** Rilwan Azeez, Larisa Lotoski, Aimée Dubeau, Natalie Rodriguez, Myrtha E. Reyna, Tyler Freitas, Stephanie Goguen, Maria Medeleanu, Geoffrey L. Winsor, Fiona S.L. Brinkman, Emily E. Cameron, Leslie Roos, Elinor Simons, Theo J. Moraes, Piush J. Mandhane, Stuart E. Turvey, Shelly Bolotin, Kim Wright, Deborah McNeil, David M. Patrick, Jared Bullard, Marc-André Langlois, Corey R. Arnold, Yannick Galipeau, Martin Pelchat, Natasha Doucas, Padmaja Subbarao, Meghan B. Azad

## Abstract

**Background:** The COVID-19 pandemic is affecting all Canadian families, with some impacted differently than others. Our study aims to: 1) determine the prevalence and transmission of SARS-CoV-2 infection among Canadian families, 2) identify predictors of infection susceptibility and severity of SARS-CoV-2 and 3) identify health and psychosocial impacts of the COVID-19 pandemic.

**Methods:** This study builds upon the CHILD Cohort Study, an ongoing multi-ethnic general population prospective cohort consisting of 3454 Canadian families with children born in Vancouver, Edmonton, Manitoba, and Toronto between 2009-12. During the pandemic, 1462 CHILD households (5378 individuals) consented to participate in the CHILD COVID-19 Add-On Study involving: (1) brief biweekly surveys about COVID-19 symptoms and testing; (2) quarterly questionnaires assessing COVID-19 exposure, testing and vaccination status, physical and mental health, and pandemic-driven life changes; (3) in-home biological sampling kits to collect blood and stool. Mean ages were 9 years (range 0-17) for children and 43 years (range 18-85) for adults. Prevalence of SARS-CoV-2 infection will be estimated from survey data and confirmed through serology testing. We will combine these new data with a wealth of pre-pandemic CHILD data and use multivariate modelling and machine learning methods to identify risk and resilience factors for susceptibility and severity to the direct and indirect effects of the pandemic.

**Interpretation:** Our short-term findings will inform key stakeholders and knowledge users to shape current and future pandemic responses. Additionally, this study provides a unique resource to study the long-term impacts of the pandemic as the CHILD Cohort Study continues.

## Introduction

SARS-CoV-2 infection results in a broad range of clinical phenotypes, from asymptomatic infection to severe disease and death^1,2^. Pre-existing medical conditions, socioeconomic disadvantage, occupation, gender, and race/ethnicity have been associated with higher SARS-CoV-2 infection rate and disease severity in adults^3–6^. However, much of the biological variation in this new disease remains unexplained, and more research is needed to understand why children are less likely than adults to develop severe COVID-19^7–9^. Identifying these divergent outcomes in adults and children will help inform prevention and treatment strategies for people of all ages^10^. Additionally, understanding the persistence and biology of antibody responses following infection or vaccination will help inform public health responses to the ongoing pandemic.

In an effort to control this pandemic, public health measures have varied across Canada both geographically and over time. COVID-19 vaccination and non-pharmaceutical interventions, such as masking and school closures, have helped slow the spread of COVID-19 in Canada and beyond^3,11,12^. However, these measures have had consequences to the economy, mental health, and emotional and physical wellbeing of family members. They may also amplify inequities experienced by marginalized and vulnerable populations^12^. It is therefore important to assess how pandemic control measures differentially affect the short and long-term physical and mental health of Canadian families.

The CHILD Cohort Study (www.childstudy.ca), an ongoing birth cohort of 3454 families, offers a unique opportunity to study COVID-19 as CHILD has collected a wealth of sociodemographic, health and biological data since 2009. The CHILD COVID-19 Add-On Study **(Figure 1)** will investigate (1) symptomatic and asymptomatic SARS-CoV-2 infection prevalence, transmission, and immunity; (2) predictors of SARS-CoV-2 infection susceptibility and severity; (3) prevalence and predictors of health and psychosocial impacts of the COVID-19 pandemic on CHILD families; and (4) individual and social determinants of COVID-19 vaccine uptake or hesitancy. Further, it will provide a lasting resource to study the long-term impacts of the pandemic as the CHILD Cohort Study continues.

**Figure 1.**
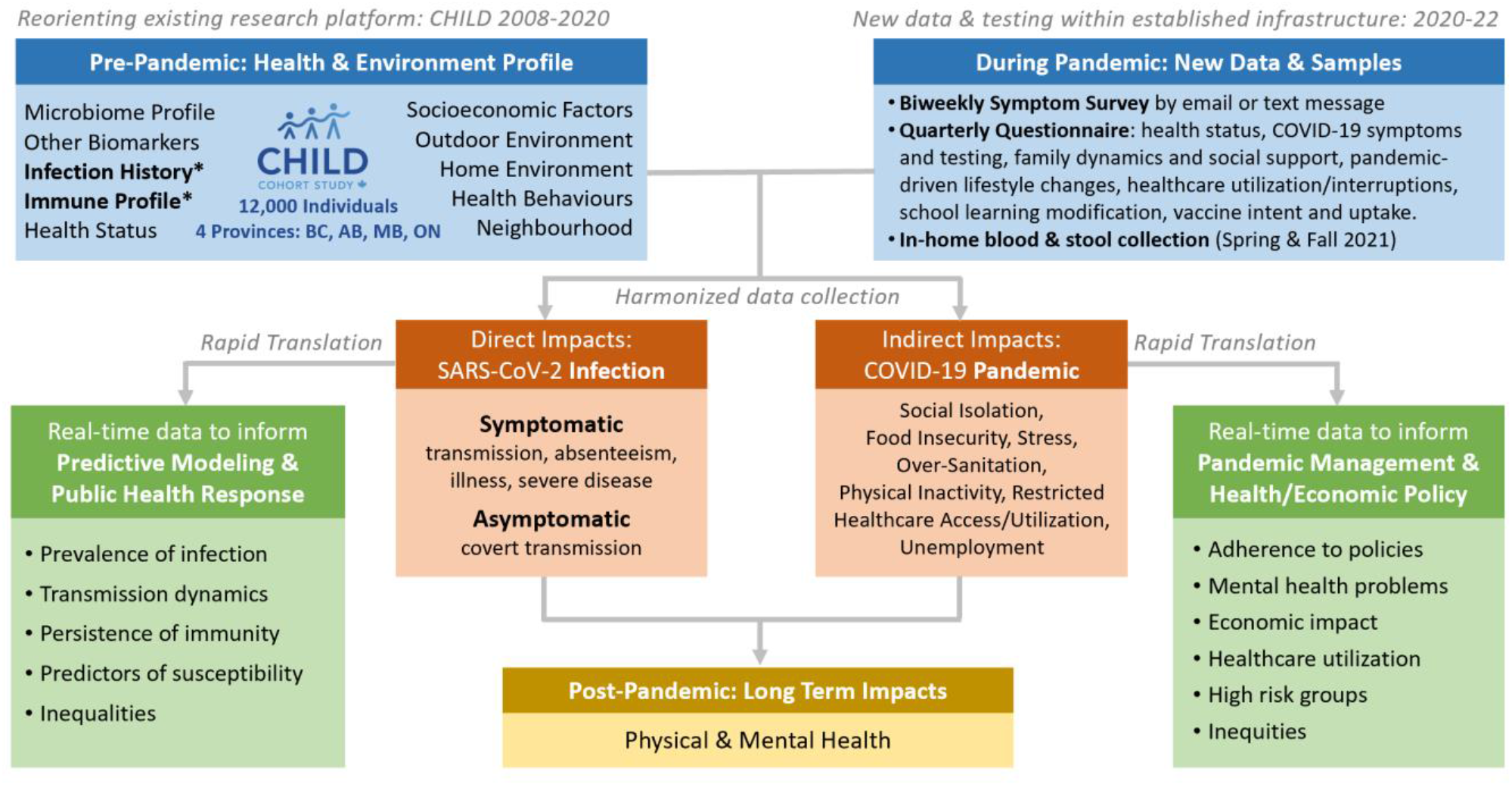
Overview of the CHILD COVID-19 Add-on Study. CHILD COVID-19 Add-On Study, a sub study of CHILD, was developed in response to the COVID-19 pandemic. The CHILD COVID-19 Study will leverage the rich pre-pandemic CHILD dataset with the newly collected pandemic dataset to study the direct effects of SARS-CoV-2 infection and the indirect effects of the COVID-19 pandemic among study participants and their households across 4 provinces in Canada. Data will be collected for the CHILD COVID-19 Add-On study through (1) biweekly surveys to assess COVID-19 symptoms and testing, (2) quarterly questionnaires on health care utilization, lifestyle, employment status, mental and physical health during the pandemic, and (3) in-home stool and finger-prick blood samples.

## Methods and Study Population

### Study design and population

This is a prospective longitudinal study embedded in the existing CHILD cohort. CHILD is a general population cohort that recruited 3454 families (∼10,000 individuals) with children born in British Columbia, Alberta, Manitoba, and Ontario between 2009-12 (90.7% retention to date)^13^. Eligible participants for the COVID Add-On Study included all parents and children enrolled in the CHILD Cohort Study plus all other individuals living in the same household (n= 3032 eligible households) **(Figure 2)**. This COVID Add-On Study involves (1) brief biweekly surveys about COVID-19 symptoms and testing; (2) quarterly questionnaires assessing COVID-19 exposure and vaccination status and pandemic-driven life changes; (3) in-home collection of blood for SARS-CoV-2 IgG serology, biomarker analysis and (optionally) genetic analysis, and stool for microbiome analysis.

**Figure 2.**
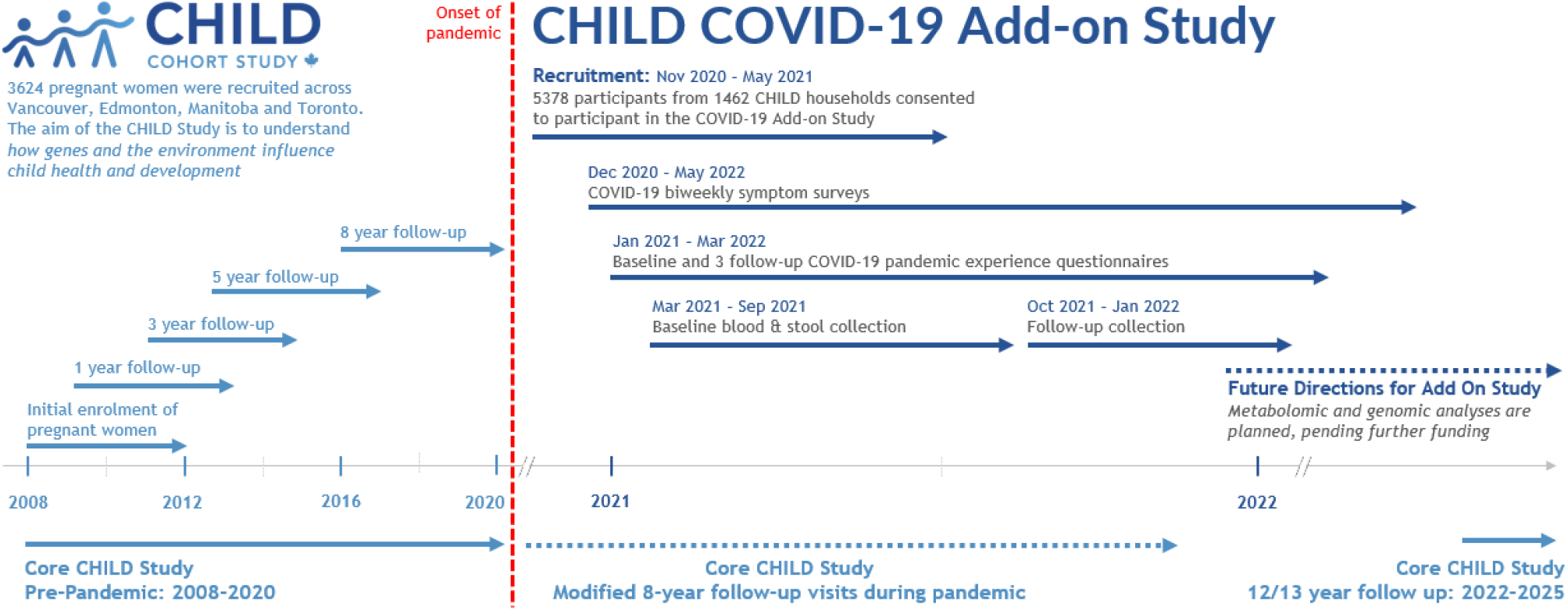
Timeline for the core CHILD Cohort Study and CHILD-COVID-19 Add-on Study. The CHILD Cohort Study, launched in 2008, is an ongoing prospective, population-based cohort study involving 3624 families recruited from Vancouver, Edmonton, Manitoba, and Toronto. The CHILD COVID-19 Add-on is embedded in the CHILD Study and was designed in 2020 to understand how the pandemic affects CHILD families.

### Consent, recruitment, and retention

Eligible participants were invited to take part in this study during in-person CHILD Cohort Study clinic visits or by phone, video call or e-mail, between November 2020 and May 2021. E-mail invitations included a summary of the study, a YouTube video (https://www.youtube.com/watch?v=lEzbn7HYiQw) describing the study’ s purpose and participation requirements, and instructions on how to arrange a virtual consent appointment with research staff. Participants attending clinic visits were provided with paper consent forms. Otherwise, consent was obtained virtually during a videoconference appointment with study staff (<u>2021 Zoom Video Communications, Inc.</u>) using the web browser-based Research Electronic Data Capture (REDCap) consent module^14,15^. Child participant assent (as defined provincially by age) or consent was obtained in the same manner in the presence of their caregiver. Participants were invited to additionally provide consent for three optional activities: (1) whole genome sequencing (65% consented), (2) future undefined analyses on their biological samples (69%) and (3) analysis of previously collected biological samples for future unknown research studies (59%). To encourage retention, we provided gift cards ($25/household for completion of the baseline survey, and again for return of biological samples) and returned SARS-CoV-2 serology results to participants.

### Recruited population and early pandemic experiences at enrolment

The final CHILD COVID-19 Add-On Study population **(Table 1)** included 5378 participants from 1462 households (mean 3.7 participants per household; IQR 3-4; range 1-12), of which 3848 were original CHILD participants (1431 mothers, 1015 fathers and 1402 children). The remaining 1530 participants were siblings (n=1427) or other household members (n=103). Almost half (48%) of eligible CHILD households consented to participate in the CHILD COVID-19 Add-On Study. A total of 370 (12% of eligible) households actively declined to participate, while the remainder were unresponsive (40%, n=1205). Compared to non-participating CHILD families, those participating in the COVID-19 Add On Study tended to have higher education and household income, and larger households **(Table S1)**. Participants were distributed across the Manitoba (34%), Vancouver (27%), Toronto (22%) and Edmonton (17%) study sites. Among children enrolled (n=2802, 52% female) the mean age was 9 years (SD 2.7, range 0-17), while among adults (n=2576, 58% female), the mean age was 43 years (SD 6.5, range 18-85) **(Figure 3)**. Participants predominantly identified as having European ancestral origins (77%) and the majority (68%) of adults had a university degree.

**Table 1.**
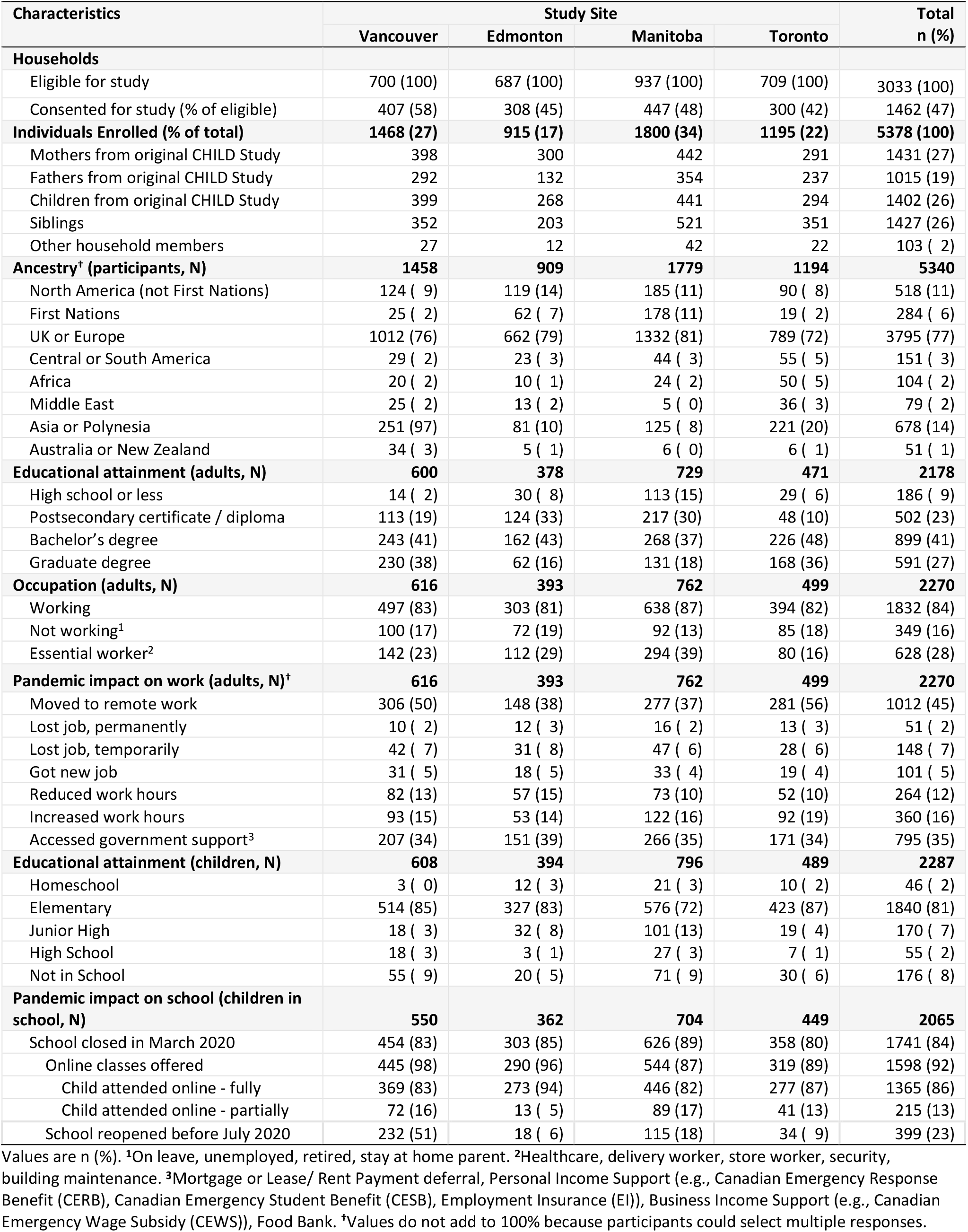
Characteristics of CHILD COVID-19 Add-On Study participants at baseline (Jan-Jun, 2021).

| Characteristics | Study Site |  |  |  | Total<br>n (%) |
| --- | --- | --- | --- | --- | --- |
|  | Vancouver | Edmonton | Manitoba | Toronto |  |
| **Households** |  |  |  |  |  |
| Eligible for study | 700 (100) | 687 (100) | 937 (100) | 709 (100) | 3033 (100) |
| Consented for study (% of eligible) | 407 (58) | 308 (45) | 447 (48) | 300 (42) | 1462 (47) |
| **Individuals Enrolled (% of total)** | **1468 (27)** | **915 (17)** | **1800 (34)** | **1195 (22)** | **5378 (100)** |
| Mothers from original CHILD Study | 398 | 300 | 442 | 291 | 1431 (27) |
| Fathers from original CHILD Study | 292 | 132 | 354 | 237 | 1015 (19) |
| Children from original CHILD Study | 399 | 268 | 441 | 294 | 1402 (26) |
| Siblings | 352 | 203 | 521 | 351 | 1427 (26) |
| Other household members | 27 | 12 | 42 | 22 | 103 ( 2) |
| **Ancestry<sup>†</sup> (participants, N)** | **1458** | **909** | **1779** | **1194** | **5340** |
| North America (not First Nations) | 124 ( 9) | 119 (14) | 185 (11) | 90 ( 8) | 518 (11) |
| First Nations | 25 ( 2) | 62 ( 7) | 178 (11) | 19 ( 2) | 284 ( 6) |
| UK or Europe | 1012 (76) | 662 (79) | 1332 (81) | 789 (72) | 3795 (77) |
| Central or South America | 29 ( 2) | 23 ( 3) | 44 ( 3) | 55 ( 5) | 151 ( 3) |
| Africa | 20 ( 2) | 10 ( 1) | 24 ( 2) | 50 ( 5) | 104 ( 2) |
| Middle East | 25 ( 2) | 13 ( 2) | 5 ( 0) | 36 ( 3) | 79 ( 2) |
| Asia or Polynesia | 251 (97) | 81 (10) | 125 ( 8) | 221 (20) | 678 (14) |
| Australia or New Zealand | 34 ( 3) | 5 ( 1) | 6 ( 0) | 6 ( 1) | 51 ( 1) |
| **Educational attainment (adults, N)** | **600** | **378** | **729** | **471** | **2178** |
| High school or less | 14 ( 2) | 30 ( 8) | 113 (15) | 29 ( 6) | 186 ( 9) |
| Postsecondary certificate / diploma | 113 (19) | 124 (33) | 217 (30) | 48 (10) | 502 (23) |
| Bachelor's degree | 243 (41) | 162 (43) | 268 (37) | 226 (48) | 899 (41) |
| Graduate degree | 230 (38) | 62 (16) | 131 (18) | 168 (36) | 591 (27) |
| **Occupation (adults, N)** | **616** | **393** | **762** | **499** | **2270** |
| Working | 497 (83) | 303 (81) | 638 (87) | 394 (82) | 1832 (84) |
| Not working<sup>1</sup> | 100 (17) | 72 (19) | 92 (13) | 85 (18) | 349 (16) |
| Essential worker<sup>2</sup> | 142 (23) | 112 (29) | 294 (39) | 80 (16) | 628 (28) |
| **Pandemic impact on work (adults, N)<sup>†</sup>** | **616** | **393** | **762** | **499** | **2270** |
| Moved to remote work | 306 (50) | 148 (38) | 277 (37) | 281 (56) | 1012 (45) |
| Lost job, permanently | 10 ( 2) | 12 ( 3) | 16 ( 2) | 13 ( 3) | 51 ( 2) |
| Lost job, temporarily | 42 ( 7) | 31 ( 8) | 47 ( 6) | 28 ( 6) | 148 ( 7) |
| Got new job | 31 ( 5) | 18 ( 5) | 33 ( 4) | 19 ( 4) | 101 ( 5) |
| Reduced work hours | 82 (13) | 57 (15) | 73 (10) | 52 (10) | 264 (12) |
| Increased work hours | 93 (15) | 53 (14) | 122 (16) | 92 (19) | 360 (16) |
| Accessed government support<sup>3</sup> | 207 (34) | 151 (39) | 266 (35) | 171 (34) | 795 (35) |
| **Educational attainment (children, N)** | **608** | **394** | **796** | **489** | **2287** |
| Homeschool | 3 ( 0) | 12 ( 3) | 21 ( 3) | 10 ( 2) | 46 ( 2) |
| Elementary | 514 (85) | 327 (83) | 576 (72) | 423 (87) | 1840 (81) |
| Junior High | 18 ( 3) | 32 ( 8) | 101 (13) | 19 ( 4) | 170 ( 7) |
| High School | 18 ( 3) | 3 ( 1) | 27 ( 3) | 7 ( 1) | 55 ( 2) |
| Not in School | 55 ( 9) | 20 ( 5) | 71 ( 9) | 30 ( 6) | 176 ( 8) |
| **Pandemic impact on school (children in school, N)** | **550** | **362** | **704** | **449** | **2065** |
| School closed in March 2020 | 454 (83) | 303 (85) | 626 (89) | 358 (80) | 1741 (84) |
| Online classes offered | 445 (98) | 290 (96) | 544 (87) | 319 (89) | 1598 (92) |
| Child attended online - fully | 369 (83) | 273 (94) | 446 (82) | 277 (87) | 1365 (86) |
| Child attended online - partially | 72 (16) | 13 ( 5) | 89 (17) | 41 (13) | 215 (13) |
| School reopened before July 2020 | 232 (51) | 18 ( 6) | 115 (18) | 34 ( 9) | 399 (23) |

Values are n (%). <sup>1</sup>On leave, unemployed, retired, stay at home parent. <sup>2</sup>Healthcare, delivery worker, store worker, security, building maintenance. <sup>3</sup>Mortgage or Lease/ Rent Payment deferral, Personal Income Support (e.g., Canadian Emergency Response Benefit (CERB), Canadian Emergency Student Benefit (CESB), Employment Insurance (EI)), Business Income Support (e.g., Canadian Emergency Wage Subsidy (CEWS)), Food Bank. <sup>†</sup>Values do not add to 100% because participants could select multiple responses.

**Figure 3.**
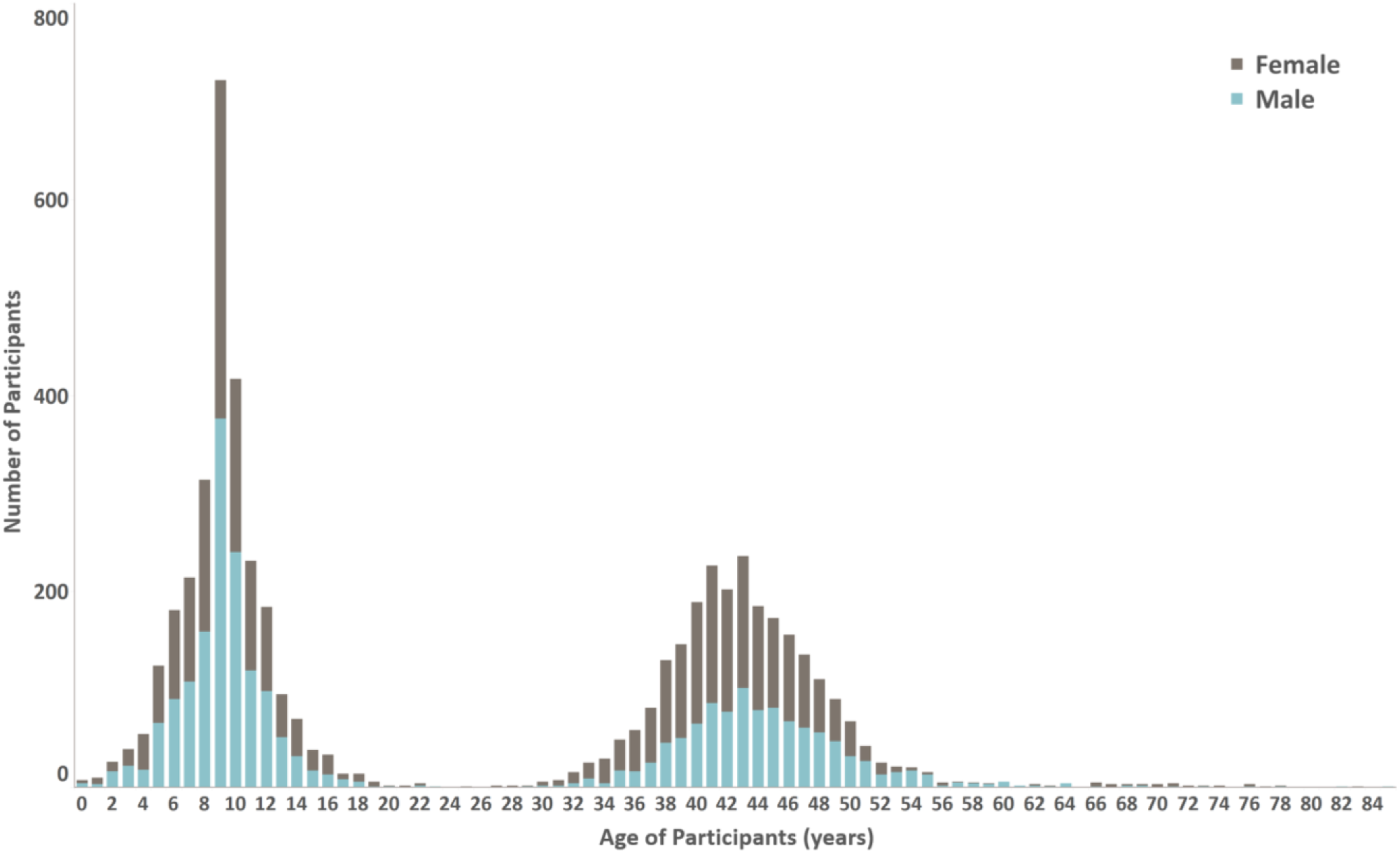
Age and sex distribution of CHILD-COVID-19 Add On Study participants.

At enrollment, 28% of adults were essential workers (e.g., healthcare, delivery, store, security and building maintenance), ranging from 16% in Toronto to 39% in Manitoba. During the first wave of the pandemic, 45% of working adults moved to remote work, 9% lost their job (permanently or temporarily) and 35% accessed government supports (e.g., loan deferrals, personal or business income support). The majority of children (84%) experienced school closures during spring 2020 (range: 80-89% across study sites), of which 23% returned to in-person classes before the end of the academic year in June 2020 (ranging from 6% in Edmonton to 51% in Vancouver). Most children who experienced school closures were offered (92%) and attended (85% fully and 14% partially) online classes **(Table 1)**.

### Biweekly surveys

Study participants’ COVID-19 symptoms, exposure and diagnosis were captured through a biweekly symptom survey **(Appendix 1)**. Beginning in December 2020, one designated participant per household was prompted biweekly by text message or e-mail (according to their preference) with three COVID-19 screening questions asking if anyone in their household had symptoms of, was suspected of having, or was tested for COVID-19. Participants responding ‘ yes’ to any of these questions were sent a follow-up COVID-19 symptom survey by e-mail to collect individual-level information on specific COVID-19 symptoms and testing, travel, and exposure history, as well as clinical or public health action taken after a positive COVID-19 test result. Completion rates for the biweekly surveys ranged from 70% in the Edmonton site to 88% in the Winnipeg site and generally were consistent over time.

### Quarterly questionnaires

Questionnaires **(Appendix 1)** were delivered online using REDCap at enrolment and approximately every 3 months thereafter **(Figure 2)**. They were adapted from other Canadian COVID-19 studies, and addressed the following topics: individual demographic characteristics, employment status, COVID-19 pandemic government support use, mental and physical health, school closures, adherence to non-pharmaceutical public health measures, screen time and use of technology for school/work or other purposes, COVID-19 symptoms, testing and diagnosis, travel history and COVID-19 vaccination perceptions and status. There were three versions of each questionnaire, designed for completion by children (about themselves), adults (about themselves), or parents/caregivers (about their children). Individuals able to provide consent on their own behalf were asked to complete the adult questionnaire. All remaining participants (i.e., children enrolled through assent or caregiver consent) were asked to complete the child questionnaire. Caregivers of participating children were also asked to complete a parent questionnaire for each child enrolled in their household. Completion rates for the quarterly questionnaires ranged from 85% (n=6287) at baseline to 44% (n=3248) at the final follow-up **(Table 2)**. In general, the sociodemographic characteristics of participants who completed the final follow-up survey (51% of adults and 49% of children) did not differ much from those who did not. However, participants who did not complete the final survey were slightly more likely to be essential workers and have a lower level of formal education (**Table S2**).

**Table 2.** Questionnaire completion and biological sample collection among 1462 households participating in the CHILD COVID-19 Add-On Study. N=5378 children and adults.

| Questionnaires | Baseline | Follow Up 1 | Follow Up 2 | Follow Up 3 |
| --- | --- | --- | --- | --- |
| Collection period | Jan-Jun, 2021 | Aug-Sep, 2021 | Oct-Dec, 2021 | Jan-Mar, 2022 |
| **Completion rate by participant type** |  |  |  |  |
| Child self-report | 1643/2064 (80) | 987/2064 (48) | 880/2064 (43) | 794/2064 (38) |
| Parent on behalf of child | 2336/2766 (84) | 1527/2766 (55) | 1275/2766 (46) | 1202/2766 (43) |
| Adult self-report | 2308/2576 (90) | 1563/2576 (61) | 1370/2576 (53) | 1252/2576 (49) |
| **Overall completion rate** | **6287/7406 (85)** | **4077/7406 (55)** | **3525/7406 (48)** | **3248/7406 (44)** |
| **Biological Samples** | **Phase A (N=5326)** |  | **Phase B (N=3219)<sup>1</sup>** |  |
| Collection period | Mar-Sep, 2021 |  | Oct-Dec, 2021 |  |
| **Sample Type** | **Stool** | **Blood** | **Stool** | **Blood** |
| Children | 1445/2738 (53) | 1462/2738 (53) | 823/1648 (50) | 847/1648 (51) |
| Adults | 1506/2588 (58) | 1567/2588 (63) | 969/1571 (62) | 1022/1571 (65) |
| **Total** | **2951/5326 (55)** | **3029/5326 (57)** | **1792/3219 (56)** | **1869/3219 (58)** |

Values are n/N (%).
<sup>1</sup>Households who returned biological samples in Phase A were invited to participate in Phase B.

**Table 3.** CHILD COVID-19 Add-On Study Questionnaires

| Questionnaire Domains & Contents | Questionnaire & Date of Data Collection |  |  |  |  |
| --- | --- | --- | --- | --- | --- |
|  | Baseline | Follow-up 1 | Follow-up 2 | Follow-up 3 | Biweekly |
|  | Jan-Jun 2021 | Aug-Sep 2021 | Oct-Dec 2021 | Jan-Mar 2022 |  |
| **COVID-19 exposure/health status** |  |  |  |  |  |
| Physical and mental health | x | x | x | x |  |
| COVID-19 exposure, symptoms, and testing | x | x | x | x | x |
| COVID-19 diagnosis and outcomes | x | x | x | x | x |
| Post-COVID (Long-COVID) symptoms |  |  |  | x |  |
| **Healthcare access and pandemic driven lifestyle changes** |  |  |  |  |  |
| Impact on healthcare access & delivery | x | x | x | x |  |
| Impact on travel, work, and childcare | x | x | x | x |  |
| Adherence to non pharmaceutical public health measures | x | x | x | x |  |
| Financial stability during the pandemic | x | x | x | x |  |
| Eating habit, sleep and physical activity | x | x | x | x |  |
| Government income support/benefits | x | x | x |  |  |
| Family and friends' connection/relationship | x | x | x |  |  |
| Screen time and social media use | x | x | x |  |  |
| Alcohol and substance use during the pandemic | x | x | x |  |  |
| Disruptions and modifications to school learning | x | x |  |  |  |
| Family routines and dynamics |  | x | x |  |  |
| Perceptions towards returning to in-person learning |  |  | x |  |  |
| **COVID-19 perceptions** |  |  |  |  |  |
| Pandemic induced worry, stress, anxiety, and loneliness | x | x | x | x |  |
| Stress and coping during the pandemic | x | x | x | x |  |
| Pandemic driven positive changes | x | x | x |  |  |
| Access to, and usage of mental health services |  | x | x | x |  |
| Social support during the pandemic |  | x | x |  |  |
| **Demographic information** |  |  |  |  |  |
| Family and household structure | x | x | x | x |  |
| Occupation | x | x | x |  |  |
| Essential worker in household | x | x | x |  |  |
| Geographic ancestral origins | x |  |  |  |  |
| Level of formal education | x |  |  |  |  |
| **Vaccination & medical history** |  |  |  |  |  |
| COVID-19 and flu vaccine uptake | x | x | x | x |  |
| COVID-19 vaccine hesitancy with reasons |  | x | x | x |  |
| COVID-19 vaccine side effects |  | x | x | x |  |
| Intent to receive COVID-19 vaccine |  | x | x | x |  |
| Vaccine perceptions and attitudes |  | x | x |  |  |
| COVID-19 vaccine information sources |  |  |  | x |  |
| Body weight and height |  |  | x | x |  |
| Current and past medical conditions | x |  |  |  |  |

'x' denotes that question was asked in the survey.

### In-home biological sample collection and SARS-CoV-2 serology

Blood and stool samples were collected twice (Spring and Fall/Winter 2021) using in-home sampling kits sent to each household along with a pre-paid return shipment box. Participants were asked to collect all samples within a 48-hour period of receipt. Sample kits were prepared by DaklaPack company (Moonachie, NJ, USA) and included all materials required to collect blood and stool, including an instructional pamphlet **(Appendix 2)**, 2-Ply 70% alcohol swabs, 2”x2” 8-Ply sterile gauze sponges (VWR CA95041-740), bandages, a registration card and specimen bag containing a Desco Humidity Indicator Cards (VWR, 89131-360) and MiniPax absorbent desiccant packets (Sigma-Aldrich, Z163570). Sample packages were returned to the Clinical Research Laboratory and Biobank (CRLB, Hamilton, ON) by airmail. Collection rates for biological samples were 56% in the first wave and 57% in the second wave of collection **(Table 2)**.

#### Blood Collection

Blood specimens were collected using the Dried Blood Spot (DBS) method and Mitra Blood Collection devices. DBS were collected using a Whatman 903 protein saver 5-spot card (VWR Int, Radnor, PA, USA). Returned sample bags containing DBS collection cards, desiccant pouches and humidity cards were sealed in a second polybag and stored at 4°C for SARS-CoV-2 serological analysis using automated chemiluminescent ELISAs to detect anti-spike and anti-nucleoprotein antibodies as described by Cholette F., et al. 2021^16^. COVID-19 antigens and anti-hIgG#5-HRP fusion antibody were generously provided by Dr. Yves Durocher, National Research Council of Canada. Cutoffs for seropositivity used were based on False Discovery Rates (FDRs) in prepandemic cohorts of 2% for Spike and both 1% and 10% for Nucleoprotein. Preserved blood was also collected using the Mitra Clamshell Devices containing two samplers (Neoteryx, Torrance, CA, USA). Upon receipt, the clamshell casing was removed, samplers were loaded into a 96-autorack, placed into a polybag containing a desiccant pouch and stored at −80°C until further use. Metabolomic and genomic analyses are planned, pending further funding.

#### Stool Collection

Stool samples were collected by participants using two cotton tipped swabs. Fecal swabs were then suspended in a capped collection tube containing ethanol for transport. Upon receipt, the shafts of the fecal swabs were cut off and the specimens were cryopreserved. Microbiome analyses are planned, pending further funding.

### Statistical analysis plan

#### Aim 1: Estimate SARS-CoV-2 infection, transmission, severity, and immunity

Prevalence and severity of infection will be estimated from self-reported biweekly reports, quarterly questionnaire testing information, and serological testing. Severity will be classified as asymptomatic, mild, or severe using WHO criteria^19^. Transmission will be assessed using multilevel modelling (by household and study site). Persistence of SARS-CoV-2-induced immunity will be determined from longitudinal serology testing for SARS-CoV-2 antibodies over two time frames (Spring and Fall 2021).

#### Aim 2: Identify predictors of SARS-CoV-2 infection susceptibility and severity

We will combine new data from the CHILD COVID-19 Add On Study with existing pre-pandemic CHILD data to predict SARS-CoV-2 infection susceptibility (incidence) and severity. We hypothesize that infection susceptibility will be associated with (a) physical interactions with (or distancing from) other individuals in the community during the pandemic, (b) sociodemographic characteristics including sex, age, occupation, socioeconomic status, and household size (c) pre-existing medical conditions including obesity and asthma, (d) neighbourhood characteristics including air pollution exposure, housing type, green and blue spaces, and (e) pre-pandemic immune biomarker and genetic profiles, viral infection history (including pre-existing coronavirus serology), respiratory health (wheezing, lung function), and health-related lifestyle factors (e.g., diet quality and physical activity). Associations will be tested in multivariate regression models with a three-category outcome (symptomatic, asymptomatic or no infection), random forest analysis, gradient boosting, recursive feature elimination and mutual information method. If sufficient cases are identified, we will also evaluate infection severity.

#### Aim 3: Understand the impact of COVID-19 pandemic on mental and physical health of Canadian families using a health equity lens

We will quantify prevalence of and changes between pre-pandemic and during-pandemic physical and mental health functioning (not directly related to SARS-CoV-2 infection). Prediction models, as described for Aim 2, will be used to identify factors that predict risk for (or protection against) poor pandemic-related mental and physical health functioning. We hypothesize that protective factors will include (a) maintaining social connections, physical activity, healthy eating behaviours, and adequate sleep during the pandemic, (b) higher pre-pandemic mental wellbeing, and (c) continued employment throughout the pandemic.

### Integrated knowledge translation and participant engagement

The CHILD COVID-19 Add-On Study adopts an integrated knowledge translation approach. To accelerate the availability and translation of high quality, real-time evidence, we included key stakeholders on our team to develop the study and translate and disseminate results. Our team includes members of the CHILD Parent Advisory Committee and Knowledge Users from public health authorities at the national (Public Health Agency of Canada) and provincial levels (BC Centre for Disease Control, Alberta Health Services, Manitoba Shared Health, Public Health Ontario) who meet monthly with study investigators to share data in real-time and adapt procedures if necessary (e.g., modify symptom surveys, questionnaires, and serology testing methods) to ensure that our results can be rapidly translated into policy and practice. Study investigators also meet monthly with the Canadian Covid Immunity Task Force Pediatric Working Group to share experiences and expertise, develop harmonized protocols, and foster research collaborations. A Rapid Results website (https://childstudy.ca/covid-rapid-results/) was developed to facilitate rapid widespread translation of study results to participants and stakeholders. To maximize recruitment, retention and relevance to families, the CHILD Parent Advisory Committee was engaged to co-develop the study design, grant application, recruitment strategies and questionnaires.

In addition to traditional dissemination of results through open access publication in peer-reviewed journals, we will use our website (<u>childstudy.ca/covid-rapid-results</u>) and social media (Twitter) to rapidly disseminate time-sensitive findings that are relevant to stakeholders and knowledge users, including provincial public health organizations and non-profit organizations (e.g., Children First Canada). We will also engage the CHILD Knowledge Mobilization Stakeholder Advisory Committee, which includes stakeholders from government, industry, patient organizations, clinical societies, child health-focused organizations, parenting-focused communications platforms, and CHILD parents.

### Ethics approval and consent to participate

This study was approved by Research ethics boards at the University of British Columbia (H20-02324), University of Alberta (Pro00102524), University of Manitoba (HS24250), The Hospital for Sick Children (1000071220) and McMaster University (1108).

### Interpretation

Our study will provide timely data on SARS-CoV-2 infection and transmission dynamics, and the mental and physical health of Canadian families during the pandemic. Importantly, these new data will be linked to a wealth of existing data from the ongoing CHILD Cohort Study where, over more than a decade of participation, CHILD families have attended repeated clinical assessments; contributed longitudinal data on their health, environment, and lifestyle; and donated biological samples for genetic and biomarker analysis. This unique data resource will allow us to determine which individuals and families are particularly susceptible or resilient to COVID-19 and the indirect psychosocial impacts of the pandemic, and then identify risk and protective factors for these outcomes. This research will help us understand how the pandemic is disrupting social connections and health behaviours, how families are coping, and which families would benefit most from targeted social supports and programming. In addition, because it is nested within an ongoing longitudinal cohort, our study has the potential to investigate long-term sequelae of COVID-19 (Long COVID).

### Limitations and pandemic-related challenges

While it offers a powerful opportunity to leverage existing research infrastructure and pre-pandemic data, nesting our study within the ongoing CHILD Cohort Study also presents challenges. For example, participant burden is a paramount concern and must be considered within the context of the ongoing CHILD Cohort Study. Additionally, generalizability may be limited by selection bias because socioeconomic status tends to be higher among CHILD families than the general population. Other study limitations are related to unanticipated challenges encountered during the pandemic **(Figure 4)**. For example, we were unable to collect data during a critical period of the pandemic (summer 2020) because ethical approvals were delayed by the introduction of many novel study elements (e.g., virtual consent by video conference and in-home sample collection). Pandemic-driven supply chain disruptions made acquiring biological sample kit components (e.g., gauze, bandages) exceedingly difficult, leading to delays in sample collection. Despite being used in the USA for serology testing, the Mitra Clamshell devices were not approved by Health Canada in time for study deployment. Standard DBS methods were used instead, requiring a more painful finger prick and larger blood volume which negatively impacted retention. The frequent biweekly questionnaires and lengthy adult and parent quarterly questionnaires created a large burden on the designated household participant responsible for providing responses for themselves and all household members. Collectively, the high burden placed on participants led to study withdrawal in a subset of families.

**Figure 4.**
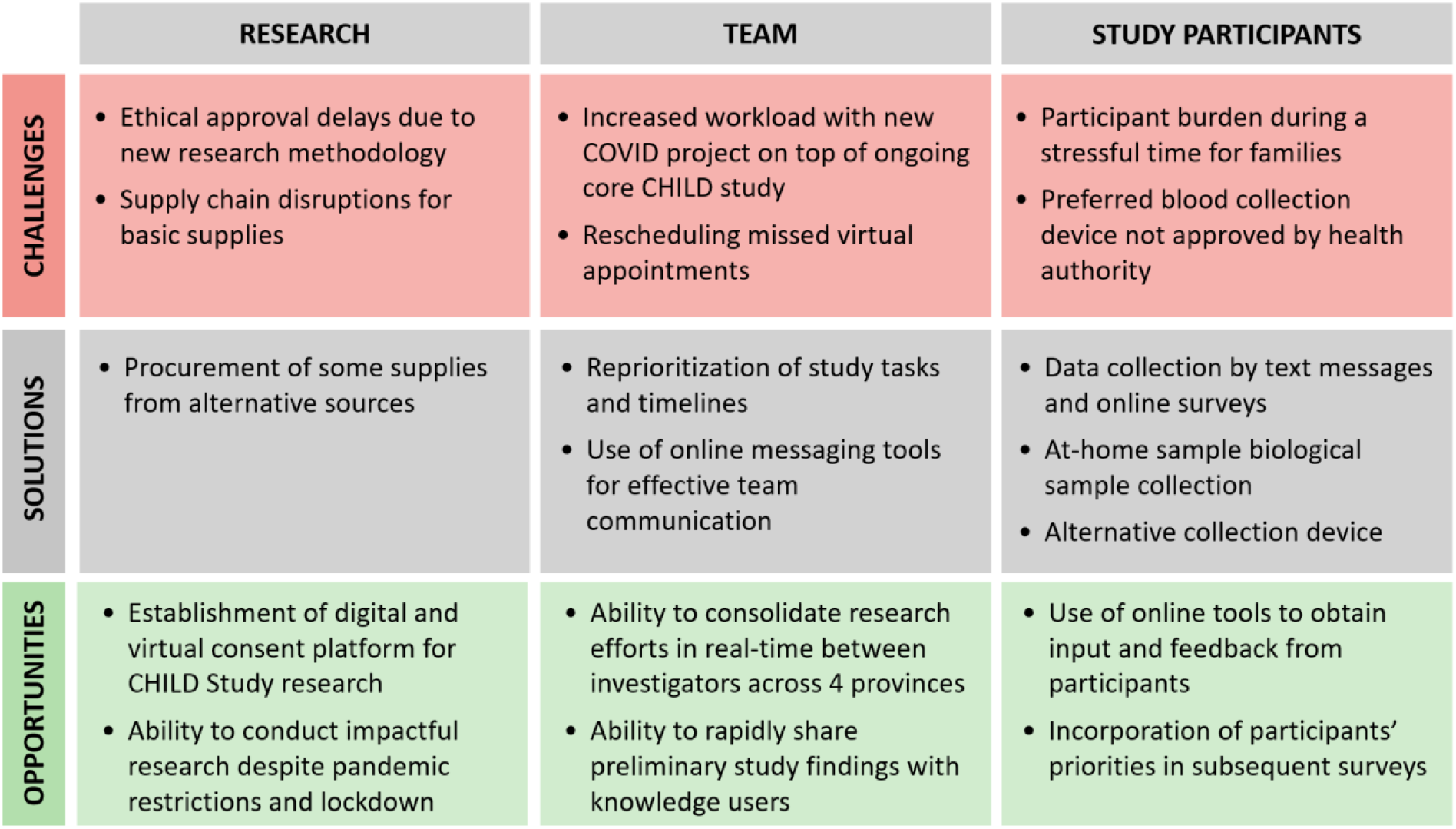
CHILD COVID-19 Add-on Study pandemic-related research challenges, solutions, and opportunities.

## Conclusion

To our knowledge, there is no other multi-site pediatric cohort with rich longitudinal data and recent pre-pandemic biosamples available to investigate how current and pre-pandemic health status, immune phenotypes and biomarkers can interactively predict susceptibility and resilience to COVID-19 and the psychosocial impacts of the pandemic. The CHILD Cohort offers a unique and powerful opportunity to study the clinical features and long-term sequelae of COVID-19, assess viral transmission within households, understand COVID-19 vaccine uptake/hesitancy, and investigate the psychosocial impact of the pandemic and its management among Canadian children and their families.

## Supporting information

STROBE statement

## Data Availability

All data produced in the present study are available upon reasonable request to the authors.

https://childstudy.ca/for-researchers/study-data/.

## Abbreviations

ACE2: Angiotensin-converting Enzyme 2
BC: British Columbia
CANCOGEN: Canadian COVID Genomics Network
CANUE: Canadian Urban Environmental Health Research Consortium
CHILD: Canadian Healthy Infant Longitudinal Development
CINECA: Common Infrastructure for National Cohorts in Europe, Canada, and Africa
COVID-19: Coronavirus disease 2019
CRISIS: CoRonavIruS Health Impact Survey
DBS: Dried Blood Spots
ELISA: Enzyme-linked Immunosorbent Assay
IgG: Immunoglobulin G
NCC: National Coordinating Centre
NGOs: Non-Governmental Organizations
REDCap: Research Electronic Data Capture
SARS-CoV-2: Severe Acute Respiratory Syndrome Coronavirus 2
SNP: Single Nucleotide Polymorphisms
UNESCO: United Nations Educational, Scientific and Cultural Organization
UNICEF: United Nations Children’ s Fund
WHO: World Health Organization

## Declarations

## Acknowledgements

We are grateful to all the families who took part in this CHILD COVID-19 Add-on Study and the entire CHILD Study team, which includes study site coordinators, research assistants, nurses, computer and laboratory technicians, clerical workers, research scientists, volunteers, managers, and receptionists at the following institutions: McMaster University, University of Manitoba, University of Alberta, SickKids, and University of British Columbia. We acknowledge Jay Onysko (Public Health Agency of Canada) who served on the CHILD COVID-19 Add-on Study Knowledge User Committee. We thank the main CHILD Cohort Study participant families for their dedication and commitment to advancing health research.

## Authors’ contributions

RA contributed to conceptualization, writing the original draft, formal analysis and reviewing and editing the final version. LL contributed to conceptualization, writing the original draft, project administration and supervision, formal analysis, and reviewing and editing the final version. MER contributed to formal analysis and reviewing the final version. MM contributed to writing the original draft and editing and reviewing the final version. AD contributed to conceptualization, project administration, and data curation. TF contributed to project administration and data curation. GW and SG contributed to data curation and formal analysis. FB contributed to conceptualization, funding acquisition, data curation, formal analysis and reviewing and editing the final version. ML, CA, YG and MP contributed to serological analysis, editing and reviewing the final version. SB, DM, KW, JB, and DP, contributed to conceptualization, funding acquisition, and reviewing and editing the final version. ES, TJM, TES, PJM and PS contributed to conceptualization, funding acquisition, and reviewing and editing the final version. EC and LR contributed to questionnaire review and revision, formal analysis, reviewing, and editing the final version. NR contributed to conceptualization, funding acquisition, project administration and reviewing the final version. MBA contributed to conceptualization, writing the original draft, funding acquisition, project administration and supervision, formal analysis, methodology and reviewing and editing the final version.

## Data availability

Existing CHILD researchers and CHILD-COVID Knowledge Users will be given rapid access to study data on request. New researchers interested in developing or collaborating on a project using CHILD data are encouraged to contact the study’ s National Coordinating Centre for a formal request. Details for data request and access are outlined on the CHILD website: https://childstudy.ca/for-researchers/study-data/.

## Supplementary Material

**Table S1.** Comparison of sociodemographic characteristics of CHILD Cohort Study participants to CHILD COVID-19 Add-on Study participants.

| Variable | All CHILD households† | CHILD households involved in COVID-19 Study | CHILD households not involved in COVID-19 Study | Chi-square *p*-value‡ |
| --- | --- | --- | --- | --- |
| **Total participants, N** | **3263** | **1386** | **1877** |  |
| **Study centre at enrollment** |  |  |  |  |
| Vancouver | 731 (21) | 378 (27) | 353 (19) | <0.001* |
| Edmonton | 767 (24) | 296 (21) | 471 (25) |  |
| Manitoba | 994 (31) | 427 (31) | 567 (30) |  |
| Toronto | 771 (24) | 285 (21) | 486 (26) |  |
| **Parents' marital status (participants, N)** | **3184** | **1379** | **1805** |  |
| Single | 169 ( 5) | 52 ( 4) | 117 ( 7) | 0.003* |
| Married/common law | 2989 (94) | 1316 (95) | 1673 (92) |  |
| Divorced/separated/widowed | 26 ( 1) | 11 ( 1) | 15 ( 1) |  |
| **Mother's race (participants, N)** | **3207** | **1379** | **1828** |  |
| European/White | 2339 (73) | 1067 (78) | 1272 (70) | <0.001* |
| First Nations | 141 ( 4) | 47 ( 3) | 94 ( 5) |  |
| Asian | 505 (16) | 191 (14) | 314 (17) |  |
| Other | 222 ( 7) | 74 ( 5) | 148 ( 8) |  |
| **Father's race (participants, N)** | **3218** | **1383** | **1835** |  |
| European/White | 2377 (74) | 1065 (78) | 1312 (71) | <0.001* |
| First Nations | 128 ( 4) | 58 ( 4) | 70 ( 4) |  |
| Asian | 442 (14) | 171 (12) | 271 (15) |  |
| Other | 271 ( 8) | 89 ( 6) | 182 (10) |  |
| **Mother's age (Mean ± SD)** | **37.9 ± 4.5** | **38.1 ± 4.1** | **37.6 ± 4.8** | 0.01* |
| **Father's age (Mean ± SD)** | **39.8 ± 5.3** | **39.9 ± 5.0** | **39.7 ± 5.7** | 0.38 |
| **Mother's education (participants, N)** | **3136** | **1361** | **1775** |  |
| High school or less | 276 ( 9) | 59 ( 4) | 217 (12) | <0.001* |
| Postsecondary certificate/diploma | 655 (21) | 283 (21) | 372 (21) |  |
| Bachelor's degree | 1605 (51) | 725 (53) | 880 (50) |  |
| Graduate degree | 600 (19) | 294 (22) | 306 (17) |  |
| **Father's education (participants, N)** | **3104** | **1347** | **1757** |  |
| High school or less | 475 (15) | 145 (11) | 330 (19) | <0.001* |
| Postsecondary certificate/ diploma | 813 (26) | 345 (26) | 468 (26) |  |
| Bachelor's degree | 1322 (43) | 639 (47) | 683 (39) |  |
| Graduate degree | 494 (16) | 218 (16) | 276 (16) |  |
| **Household income (households, N)** | **2890** | **1288** | **1602** |  |
| < \$30,000 | 127 ( 4) | 31 ( 2) | 96 ( 6) | <0.001* |
| \$30,000-\$59,999 | 352 (12) | 110 ( 9) | 242 (15) | |
| \$60,000-\$99,999 | 667 (23) | 298 (23) | 369 (23) | |
| \$100,000-\$149,999 | 822 (29) | 388 (30) | 434 (27) | |
| ≥ \$150,000 | 922 (32) | 461 (36) | 461 (29) | |
| **Household size (households, N)** | **3121** | **1373** | **1748** |  |
| 1-3 | 1046 (34) | 335 (24) | 711 (41) | <0.001* |
| 4-6 | 1965 (63) | 990 (72) | 975 (56) |  |
| >6 | 110 ( 3) | 48 ( 4) | 62 ( 3) |  |

Values are n (%) or mean ± standard deviation, for households with non-missing data for each variable. Values reflect most recently available data up to 5-year CHILD study visit (completed in 2018-2020), unless otherwise stated. "Enrollment" refers to the original CHILD cohort study, undertaken in 2009-12.
\* $p < 0.05$ . †General cohort (excludes Vanguard cohort of the CHILD study). ‡Chi-square *p*-value for comparison of CHILD participants involved vs not involved in CHILD COVID-19 Add-on Study.

**Table S2.** Sociodemographic characteristics of CHILD COVID-19 Add-on Study participants who did vs. did not complete the final follow-up questionnaire.

|  | Enrolled and completed final questionnaire | Enrolled but did not complete final questionnaire | Chi-square *p*-value |
| --- | --- | --- | --- |
| **Total participants, N** | **2388** | **2256** |  |
| **Participants** |  |  |  |
| Adults | 1216 (51) | 1065 (47) | 0.01* |
| Children | 1172 (49) | 1191 (53) |  |
| **Ancestry<sup>†</sup>** | **2387** | **2232** |  |
| North American (not First Nation) | 252 (11) | 266 (12) | 0.16 |
| First Nation | 138 ( 6) | 146 ( 7) | 0.31 |
| UK or Europe | 1985 (83) | 1810 (81) | 0.07 |
| Central or South America | 57 ( 2) | 94 ( 4) | 0.001* |
| Africa | 41 ( 2) | 63 ( 3) | 0.02* |
| Middle East | 21 ( 1) | 58 ( 3) | <0.001* |
| Asia or Polynesia | 394 (17) | 284 (13) | <0.001* |
| Australia or New Zealand | 29 ( 1) | 22 ( 1) | 0.55 |
| **Education attainment (adults, N)** | **1197** | **992** |  |
| High school or less | 85 ( 7) | 101 (10) | <0.001* |
| Post-secondary certificate/diploma | 268 (22) | 237 (24) |  |
| Bachelor's degree | 510 (43) | 392 (40) |  |
| Graduate degree | 334 (28) | 262 (26) |  |
| **Occupation (adults, N)** | **1164** | **975** |  |
| Working | 990 (85) | 842 (86) | 0.28 |
| Not Working<sup>1</sup> | 174 (15) | 133 (14) |  |
| **Occupation type (adults, N)** | **1216** | **1065** |  |
| Essential worker<sup>2</sup> | 341 (28) | 295 (28) | 0.25 |
| **Pandemic impact on work (adults, N)<sup>†</sup>** | **1215** | **1045** |  |
| Moved to remote work | 566 (47) | 446 (43) | 0.01* |
| Lost job, permanently | 21 ( 2) | 30 ( 3) | 0.05* |
| Lost job, temporarily | 74 ( 6) | 74 ( 7) | 0.15 |
| Got new job | 49 ( 4) | 52 ( 5) | 0.14 |
| Reduced work hours | 135 (11) | 129 (12) | 0.17 |
| Increased work hours | 186 (15) | 174 (17) | 0.18 |
| Increased risk of COVID from job | 288 (24) | 246 (23) | 0.17 |
| Accessed government support<sup>3</sup> | 399 (33) | 396 (37) | 0.02* |
| **Educational attainment (children, N)** | **1170** | **1126** |  |
| Home school | 29 ( 3) | 17 ( 1) | 0.05 |
| Elementary school | 943 (80) | 897 (80) |  |
| Junior high | 80 ( 7) | 91 ( 8) |  |
| High school | 23 ( 2) | 40 ( 4) |  |
| Not in school | 95 ( 8) | 81 ( 7) |  |
| **Pandemic impact on school (children in school, N)** | **1046** | **1028** |  |
| School closed in March 2020 | 878 (84) | 863 (84) | 0.50 |
| *Of schools that closed,* |  |  |  |
| Online classes offered | 814 (93) | 784 (91) | 0.13 |
| Online classes attended - fully | 708 (88) | 657 (85) | 0.14 |
| Online classes attended - partially | 96 (12) | 119 (15) | 0.06 |
| School reopened before July 2020 | 206 (24) | 200 (23) | 0.82 |

Values are n (%) for participants with non-missing data for each variable. Values are derived from the baseline questionnaires completed between Jan – Jun 2021.
<sup>1</sup>On leave, unemployed, retired, stay at home parent. <sup>2</sup>Healthcare, delivery worker, store worker, security, building maintenance.
<sup>3</sup>Mortgage or Lease/ Rent Payment deferral, Personal Income Support (e.g., Canadian Emergency Response Benefit (CERB), Canadian Emergency Student Benefit (CESB), Employment Insurance (EI)), Business Income Support (e.g., Canadian Emergency Wage Subsidy (CEWS)), Food Bank. \* *p* < 0.05. <sup>†</sup>Values do not add to 100% because participants could select multiple responses.

Appendix 1 – CHILD COVID-19 Add-On Study Data Collection Tools

Appendix 2 - CHILD COVID-19 Add-On Study Sample Collection Instructions

