## Appendix 1. for "Investigating SARS-CoV-2 infection and the health and psychosocial impact of the COVID-19 pandemic in the Canadian CHILD Cohort: study methodology and cohort profile"

### Child Self Report

We are interested in finding about how you are feeling and what you are thinking about during the Coronavirus (COVID-19) pandemic.

COVID-19 is a new type of illness affecting many people in Canada. The virus that causes COVID-19 is related to the common cold virus. Some people affected by the virus may have symptoms that can be mild or severe, similar to a cold or flu. Other people with COVID-19 may not have any symptoms at all but they can spread the virus to others.

It is important for scientists to learn more about COVID-19. You can help us by answering this survey. Do the best you can to answer each question. This survey should take about 10 minutes to complete.

If you have questions or want help filling the survey out, please ask a family member to help you.

**1.** Before the COVID-19 crisis in your area, how would you rate your overall physical health (how healthy your body is)?

- ○ Excellent
- ○ Very Good
- ○ Good
- ○ Fair
- ○ Poor

**2.** Before the COVID-19 crisis in your area, how would you rate your overall mental/emotional health (the way you think and feel)?

- ○ Excellent
- ○ Very Good
- ○ Good
- ○ Fair
- ○ Poor

**3.** Since March 15th 2020, have you had any of the following symptoms? (Select all that apply)

- ☐ Cough
- ☐ Shortness of breath
- ☐ Difficulty breathing
- ☐ Fever
- ☐ Chills
- ☐ Muscle or body aches
- ☐ Headache
- ☐ Sore throat
- ☐ New loss of sense of smell or taste
- ☐ Runny nose
- ☐ Other, Specify ____________________
- ☐ None of the above

**4.** How much are you asking questions, reading, or talking about COVID-19?

- ○ Never
- ○ A few times a month
- ○ Weekly
- ○ A few times a week
- ○ Daily

**5.** Has the COVID-19 crisis in your area led to any POSITIVE (GOOD) changes in your life?

- ○ Yes
- ○ No (Go to Question 6)

**5.1** Please specify these POSITIVE (GOOD) changes in your life (Select all that apply)

- ☐ More time with family
- ☐ More time with friends
- ☐ More exercising
- ☐ Healthier eating
- ☐ New hobbies
- ☐ Other, specify ____________________

#### LIFE CHANGES DUE TO THE COVID-19 CRISIS

**6.** During the PAST TWO WEEKS how many people (who do not live with you) have you talked to in-person?

____________________
(In-person means that the person is in the same place as you (NOT on the phone or video chat))

**7.** During the PAST TWO WEEKS, how many times did you go outside of your home (e.g., going to stores, parks, etc.)?

- ○ Not at all
- ○ 1-2 days per week
- ○ 3-4 days per week
- ○ 5-6 days per week
- ○ Every day

**8.** During the PAST TWO WEEKS, how stressful have the restrictions on leaving home been for you?

- ○ Not stressful
- ○ A little bit stressful
- ○ Stressful
- ○ Very stressful
- ○ Extremely stressful
- ○ There are no restrictions on leaving home

**9.** During the PAST TWO WEEKS, how difficult has it been for you to follow the recommendations for keeping away from close contact with people who you don't live with?

- ○ Not difficult
- ○ A little bit difficult
- ○ Difficult
- ○ Very difficult
- ○ Extremely difficult
- ○ There are no recommendations for keeping away from close contact with people who I don't live with

**10.** During the PAST TWO WEEKS, has the quality of the relationships between you and members of your family changed?

- ○ A lot better
- ○ A little better
- ○ About the same (Go to Question 11)
- ○ A little worse
- ○ A lot worse

**10.1** During the PAST TWO WEEKS, how stressful have these changes in relationships between you and members of your family been for you?

- ○ Not stressful
- ○ A little bit stressful
- ○ Stressful
- ○ Very stressful
- ○ Extremely stressful

**11.** During the PAST TWO WEEKS, has the quality of your relationships with your friends changed?

- ○ A lot better
- ○ A little better
- ○ About the same (Go to Question 12)
- ○ A little worse
- ○ A lot worse

**11.1** During the PAST TWO WEEKS, how stressful have these changes in your relationships with your friends been for you?

- ○ Not stressful
- ○ A little bit stressful
- ○ Stressful
- ○ Very stressful
- ○ Extremely stressful

**12.** During the PAST TWO WEEKS, how difficult has the cancellation of important events in your life (such as sporting events, school trips or vacations, etc.) been for you?

- ○ Not difficult
- ○ A little bit difficult
- ○ Difficult
- ○ Very difficult
- ○ Extremely difficult
- ○ There hasn't been any cancellation of important events

**13.** How hopeful are you that the COVID-19 crisis in your area will end soon?

- ○ Extremely hopeful
- ○ Very hopeful
- ○ Hopeful
- ○ A little bit hopeful
- ○ Not at all hopeful
- ○ There is no COVID-19 crisis in my area

#### DAILY BEHAVIOURS (PAST TWO WEEKS)

**14.** During the PAST TWO WEEKS on average, what time did you go to bed on WEEKDAYS?

- ○ Before 8 pm
- ○ 8 pm - 10 pm
- ○ 10 pm - 12 am (Midnight)
- ○ After 12 am (Midnight)

**15.** During the PAST TWO WEEKS on average, what time did you go to bed on WEEKENDS?

- ○ Before 8 pm
- ○ 8 pm - 10 pm
- ○ 10 pm - 12 am (Midnight)
- ○ After 12 am (Midnight)

**16.** During the PAST TWO WEEKS on average, how many hours per night did you sleep on WEEKDAYS?

- ○ Less than 6 hours
- ○ 6-8 hours
- ○ 8-10 hours
- ○ Greater than 10 hours

**17.** During the PAST TWO WEEKS on average, how many hours per night did you sleep on WEEKENDS?

- ○ Less than 6 hours
- ○ 6-8 hours
- ○ 8-10 hours
- ○ Greater than 10 hours

**18.** During the PAST TWO WEEKS, how many days per week did you exercise (e.g., increased heart rate, breathing) for at least 30 minutes?

- ○ None
- ○ 1-2 days
- ○ 3-4 days
- ○ 5-6 days
- ○ Daily

**19.** During the PAST TWO WEEKS, how many days per week did you spend time outdoors?

- ○ None
- ○ 1-2 days
- ○ 3-4 days
- ○ 5-6 days
- ○ Daily

#### EMOTIONS/ WORRIES

**During the PAST TWO WEEKS…**

| | Not at all worried | Slightly worried | Moderately worried | Very worried | Extremely worried |
|---|---|---|---|---|---|
| **20.** How worried have you been about being infected by the virus that causes COVID-19? | ○ | ○ | ○ | ○ | ○ |
| **21.** How worried have you been about friends or family being infected by the virus that causes COVID-19? | ○ | ○ | ○ | ○ | ○ |
| **22.** How worried have you been about your physical health (how healthy your body is) being influenced by COVID-19? | ○ | ○ | ○ | ○ | ○ |
| **23.** How worried have you been about your mental/emotional health (how you think and feel) being influenced by COVID-19? | ○ | ○ | ○ | ○ | ○ |
| **24.** How worried were you generally? | ○ | ○ | ○ | ○ | ○ |

**25.** During the PAST TWO WEEKS, how happy versus sad were you?

- ○ Very happy/cheerful
- ○ Moderately happy/cheerful
- ○ Neutral
- ○ Moderately sad/depressed/unhappy
- ○ Very sad/depressed/unhappy

**26.** During the PAST TWO WEEKS, how relaxed versus anxious were you?

- ○ Very relaxed/calm
- ○ Moderately relaxed/calm
- ○ Neutral
- ○ Moderately nervous/anxious
- ○ Very nervous/anxious

**27.** During the PAST TWO WEEKS, how fidgety or restless were you?

- ○ Not fidgety/restless at all
- ○ Slightly fidgety/restless
- ○ Moderately fidgety/restless
- ○ Very fidgety/restless
- ○ Extremely fidgety/restless

**28.** During the PAST TWO WEEKS, how tired were you?

- ○ Not tired at all
- ○ Slightly tired
- ○ Moderately tired
- ○ Very tired
- ○ Extremely tired

**29.** During the PAST TWO WEEKS, how well were you able to concentrate or focus?

- ○ Very focused/attentive
- ○ Moderately focused/attentive
- ○ Neutral
- ○ Moderately unfocused/distracted
- ○ Very unfocused/distracted

**30.** During the PAST TWO WEEKS, how irritable or easily angered were you?

- Not irritable or easily angered at all
- Slightly irritable or easily angered
- Moderately irritable or easily angered
- Very irritable or easily angered
- Extremely irritable or easily angered

**31.** During the PAST TWO WEEKS, how lonely were you?

- Not lonely at all
- Slightly lonely
- Moderately lonely
- Very lonely
- Extremely lonely

#### MEDIA USE (PAST TWO WEEKS)

**32.** During the PAST TWO WEEKS, how much time per day did you spend watching TV or digital media (e.g., Netflix, YouTube, web surfing)?

(Include time for educational purposes, including school work)

- No TV or digital media
- Under 1 hour
- 1-3 hours
- 4-6 hours
- More than 6 hours

**33.** During the PAST TWO WEEKS, how much time per day did you spend using social media (e.g., Facebook, Instagram, Snapchat, Twitter, TikTok)?

(Include time for educational purposes, including school work)

- No social media
- Under 1 hour
- 1-3 hours
- 4-6 hours
- More than 6 hours

**34.** During the PAST TWO WEEKS, how much time per day did you spend playing video games?

- No video games
- Under 1 hour
- 1-3 hours
- 4-6 hours
- More than 6 hours

**35.** Please share any comments, suggestions or clarifications that you have related to this survey

______________________________

**36.** Did **you** complete this survey?

Note: **You** refers to the Child participant in the study

- Yes - by myself
- Yes - partially or with help from someone else
- No - someone else completed this survey on their behalf

**37.** Today's Date

(DD-MM-YYYY)

________________________

### Parental Report

**Study ID:**

We are interested in finding out about how your child is feeling and what they are thinking about during the Coronavirus (COVID-19) pandemic.

COVID-19 is a new type of illness affecting many people in Canada. The virus that causes COVID-19 is related to the common cold virus. Some people affected by the virus may have symptoms that can be mild or severe, similar to a cold or flu. Other people with COVID-19 may not have any symptoms at all but they can spread the virus to others.

It is important for scientists to learn more about COVID-19. You can help us by answering this survey about your child. Do the best you can to answer each question. This survey should take about 10 minutes to complete.

If you have questions or want help filling the survey out, please contact your site's Coordinator.

#### COVID-19 HEALTH/EXPOSURE STATUS

**1.** How would you rate the child's overall physical health before the COVID-19 crisis in the child's area?

- ○ Excellent
- ○ Very Good
- ○ Good
- ○ Fair
- ○ Poor

**2.** How would you rate the child's overall mental/emotional health before the COVID-19 crisis in the child's area?

- ○ Excellent
- ○ Very Good
- ○ Good
- ○ Fair
- ○ Poor

**3.** In what ways has the COVID-19 outbreak affected the child's overall healthcare? (Select all that apply)

- ☐ The child did not go to healthcare appointments because I was concerned about entering the healthcare provider's office
- ☐ The child's healthcare provider cancelled appointments
- ☐ The child's healthcare provider changed to phone or online visits
- ☐ The child's healthcare provider told him/her to self-isolate or quarantine
- ☐ None of these apply

**4.** Has the COVID-19 crisis in your child's area led to any family members in the child's household(s) applying for or accessing any of the following supports? (Select all that apply)

- ☐ Residential Mortgage or Lease/ Rent Payment Deferral
- ☐ Commercial Mortgage or Lease/ Rent Payment Deferral
- ☐ Personal Income Support (e.g Canadian Emergency Response Benefit (CERB), Canadian Emergency Student Benefit (CESB), Employment Insurance (EI))
- ☐ Business Income Support (e.g Canadian Emergency Wage Subsidy (CEWS))
- ☐ Food Bank
- ☐ Other, Specify________________________________
- ☐ None of the above
- ☐ Don't Know

**5.** Has the child been exposed to someone likely to have COVID-19? (Select all that apply)

- ☐ Yes, someone with positive test
- ☐ Yes, someone with medical diagnosis, but no test
- ☐ Yes, someone with possible symptoms, but no diagnosis by doctor
- ☐ No, not to my knowledge

**6.** Has the child been suspected of having COVID-19?

- ○ Yes, positive test
- ○ Yes, medical diagnosis, but no test
- ○ Yes, has had some possible symptoms, but no diagnosis by health care professional
- ○ No symptoms or signs
- ○ Don't know

**7.** Has the child ever been tested for the virus that causes COVID-19? (Select all that apply)

(Do not include the antibody/ serology test (blood test for previous infection) that is part of this research study )

- ☐ No, I never tried to get the child tested (Answer Question 7.3)
- ☐ No, I tried to get the child tested but was not able to (Answer Question 7.1)
- ☐ Yes, and the child is waiting for the results (Answer Question 7.2)
- ☐ Yes, and the test showed that the child did not have it ("negative" test) (Answer Question 7.2)
- ☐ Yes, and the test showed that the child did have it ("positive" test) (Answer Question 7.2)

**7.1** Which type of test for the virus that causes COVID-19 did you try to get for the child but were not able to?

(Antibody/ Serology testing and saliva/ mouth rinse testing may not be currently available in all areas, but it may be available in the future.)

- ☐ Viral Test (swab test for current infection)
- ☐ Antibody/ Serology Test (blood test for previous infection)
- ☐ Saliva or Mouth Rinse Test
- ☐ Other Specify ______________________
- ☐ Don't Know

**7.2** Please specify why the child got tested or tried to get tested

- ☐ A contact or possible contact of the child tested positive for COVID-19
- ☐ The child had COVID-19 symptoms
- ☐ The child was admitted to hospital (unrelated to COVID-19)
- ☐ The child was admitted to hospital (COVID-19 related)
- ☐ The child was tested in another facility or instiution context (contact with someone in long term care, prison etc)
- ☐ The child was participating in another facility research study
- ☐ Travel-related testing
- ☐ Other, specify ______________________
- ☐ Don't Know

**7.3** Please specify why you didn't try to get the child tested

- ☐ Symptoms were due to another condition, specify__________
- ☐ Symptoms were mild
- ☐ A healthcare provider did not recommend testing
- ☐ Other, specify______________________

| | |
|---|---|
| **7.4** In the past two weeks how many times has your child been tested for COVID-19? | ◯ 0 (Go to Question 8)<br>◯ 1<br>◯ 2<br>◯ 3<br>◯ 4 |
| Details of First COVID-19 Test | |
| **7.5** What type of test did your child receive?<br><br>(Do not include the antibody/ serology test (blood test for previous infection) that is part of this reseach study ) | ◯ Viral Test (swab test for current infection)<br>◯ Antibody/ Serology Test (blood test for previous infection)<br>◯ Saliva or Mouth Rinse Test<br>◯ Other Specify________________<br>◯ Don't Know |
| **7.6** Date of test<br>(DD-MM-YYYY) | ________________ |
| **7.7** First test result | ◯ Negative ◯ Positive<br>◯ Don't know |
| Did your child complete a second COVID test? | ◯ Yes<br>◯ No (Go to Question 8) |
| Details of Second COVID-19 Test | |
| **7.8** What type of test did your child receive?<br><br>(Do not include the antibody/ serology test (blood test for previous infection) that is part of this reseach study ) | ◯ Viral Test (swab test for current infection)<br>◯ Antibody/ Serology Test (blood test for previous infection)<br>◯ Saliva or Mouth Rinse Test<br>◯ Other Specify________________<br>◯ Don't Know |
| **7.9** Date of test<br>(DD-MM-YYYY) | ________________ |
| **7.10** Second test result | ◯ Negative ◯ Positive<br>◯ Don't know |
| Did your child complete a third COVID test? | ◯ Yes<br>◯ No (Go to Question 8) |

Details of Third COVID-19 Test

**7.11** What type of test did your child receive?

(Do not include the antibody/ serology test (blood test for previous infection) that is part of this reseach study )

- ◯ Viral Test (swab test for current infection)
- ◯ Antibody/ Serology Test (blood test for previous infection)
- ◯ Saliva or Mouth Rinse Test
- ◯ Other Specify ______________________________
- ◯ Don't Know

**7.12** Date of test
(DD-MM-YYYY)

______________________________

**7.13** Third test result

◯ Negative ◯ Positive
◯ Don't know

Did your child complete a fourth COVID test?

◯ Yes
◯ No (Go to Question 8)

Details of Fourth COVID-19 Test

**7.14** What type of test did your child receive?

(Do not include the antibody/ serology test (blood test for previous infection) that is part of this reseach study )

- ◯ Viral Test (swab test for current infection)
- ◯ Antibody/ Serology Test (blood test for previous infection)
- ◯ Saliva or Mouth Rinse Test
- ◯ Other Specify ______________________________
- ◯ Don't Know

**7.15** Date of test
(DD-MM-YYYY)

______________________________

**7.16** Fourth test result

◯ Negative ◯ Positive
◯ Don't know

**8.** Since March 15th 2020, has the child had any colds?

◯ Yes ◯ No ◯ Don't Know

| Question | Response |
|---|---|
| **9.** Since March 15th 2020, has the child had any of the following symptoms? (Select all that apply) | ☐ New or worsening cough<br>☐ Shortness of breath<br>☐ Difficulty breathing<br>☐ Fever<br>☐ Chills<br>☐ Muscle or body aches<br>☐ Headache<br>☐ Sore throat<br>☐ New loss of sense of smell or taste<br>☐ Runny nose or nasal congestion*<br>☐ Nausea or vomiting<br>☐ Diarrhea<br>☐ Fatigue<br>☐ Skin Rash<br>☐ Conjunctivitis (pink eye)<br>☐ Other, Specify ______________________<br>☐ None of the above (Go to Question 10)<br><br>(*Runny nose or nasal congestion in the absence of underlying reason for these symptoms such as seasonal allergies, post nasal drip, etc) |
| **9.1** Do you remember the date when the child had their first symptom? | ◯ Yes<br>◯ No (Go to Question 9.3)<br>◯ Roughly |
| **9.2** What was the date of the child's first symptom? (DD-MM-YYYY) | ______________________ |
| **9.3** In the TWO WEEKS before the child had symptoms, did the child: (Select all that apply) | ☐ Have contact with someone who tested positive for COVID-19 (Go to Question 9.21)<br>☐ Have contact with someone who likely had COVID-19 (e.g., was not tested but had symptoms; was told by a healthcare provider that he/she likely had it) (Go to Question 9.21)<br>☐ Travel outside the province<br>☐ None of the above (Go to Question 9.21) |
| **9.4** How many places did your child travel? (Travel outside the province) | ◯ 0 (Go to Question 9.21)<br>◯ 1<br>◯ 2<br>◯ 3<br>◯ 4 |
| Place 1 | |
| **9.5** Location (e.g. Seattle, Washington OR Toronto, Ontario) | ______________________ |
| **9.6** Start date of travel (DD-MM-YYYY) | ______________________ |
| **9.7** End date of travel (DD-MM-YYYY) | ______________________ |
| **9.8** How? (Check all that apply) | ☐ Car ☐ Plane ☐ Train<br>☐ Ship ☐ Other ______________________ |

| | |
|---|---|
| Did your child travel to a second place? | ◯ Yes<br>◯ No (Go to Question 9.21) |
| Place 2 | |
| **9.9** Location<br>(e.g. Portland, Oregon OR Vancouver, BC) | ______________ |
| **9.10** Start date of travel<br>(DD-MM-YYYY) | ______________ |
| **9.11** End date of travel<br>(DD-MM-YYYY) | ______________ |
| **9.12** How? (Check all that apply) | ☐ Car ☐ Plane ☐ Train<br>☐ Ship ☐ Other ______________ |
| Did your child travel to a third place? | ◯ Yes<br>◯ No (Go to Question 9.21) |
| Place 3 | |
| **9.13** Location<br>(e.g. Paris, France OR Banff, Alberta) | ______________ |
| **9.14** Start date of travel<br>(DD-MM-YYYY) | ______________ |
| **9.15** End date of travel<br>(DD-MM-YYYY) | ______________ |
| **9.16** How? (Check all that apply) | ☐ Car ☐ Plane ☐ Train<br>☐ Ship ☐ Other ______________ |
| Did your child travel to a fourth place? | ◯ Yes<br>◯ No (Go to Question 9.21) |
| Place 4 | |
| **9.17** Location<br>(e.g. Portland, Oregon OR Vancouver, BC) | ______________ |
| **9.18** Start date of travel<br>(DD-MM-YYYY) | ______________ |
| **9.19** End date of travel<br>(DD-MM-YYYY) | ______________ |

| | |
|---|---|
| **9.20** How? (Check all that apply) | ☐ Car ☐ Plane ☐ Train<br>☐ Ship ☐ Other ______________ |
| **9.21** Which of the following occurred as a result of the child's symptoms? (Select all that apply) | ☐ You/ the child spoke to a healthcare provider over the phone, by email, or online<br>☐ The child saw a healthcare provider in person, such as in a clinic or doctor's office<br>☐ The child saw healthcare provider in person, such as in urgent care, or in an Emergency Department (ED)<br>☐ The child was kept overnight in a hospital because a healthcare provider thought he/she had COVID-19<br>☐ The child was admitted to a hospital's ICU (Intensive Care Unit)<br>☐ The child self-isolated or quarantined at home<br>☐ None of the above |
| **10.** Has anyone in the child's family been diagnosed with COVID-19? (Select all that apply) | ☐ Yes, member of household<br>☐ Yes, non-household member<br>☐ No<br>☐ Don't know |
| **11.** Have any of the following happened to the child's family members because of COVID-19? (Select all that apply) | ☐ Fallen ill physically<br>☐ Hospitalized<br>☐ Self-isolated or quarantine with symptoms<br>☐ Self-isolated or quarantine without symptoms (e.g., due to possible exposure)<br>☐ Lost job or been laid off from job<br>☐ Reduced ability to earn money<br>☐ Passed away<br>☐ None of the above<br>(Include any family members, not just those living in the same household as the child) |
| **12.** Has the child been vaccinated against COVID-19? | ○ Yes<br>○ No (Go to Question 13)<br>(Answer 'Yes' if the child has received at least one dose of the COVID-19 vaccine. Note: Certain types of vaccines require more than one dose to protect against COVID-19. You or the child would have been informed at the time of vaccination if the child needed a second dose.) |
| **12.1** How many doses of the COVID-19 vaccine has the child received so far? | ○ One dose ○ Two doses<br>○ More than two doses |
| **12.2** When did the child receive their first dose of the COVID-19 vaccine? | ______________ |

If your child received more than one dose, go to Question 12.3. If not, continue to Question 12.4.

| | |
|---|---|
| **12.3** When did the child receive their second dose of the COVID-19 vaccine? | ______________ |
| **12.4** Which vaccine did the child receive? | ○ Pfizer and BioNTech mRNA vaccine<br>○ Moderna mRNA vaccine<br>○ AstraZeneca Oxford vaccine<br>○ Other, specify ______________<br>○ Don't Know |

#### How often has the child done any of the following?

**13.** How often has the child worn a mask in public places between **January-March 2020**?

- Never
- Rarely
- Occasionally
- Often
- Always
- Don't Know

**14.** How often has the child worn a mask in public places between **April-August 2020**?

- Never
- Rarely
- Occasionally
- Often
- Always
- Don't Know

**15.** How often has the child worn a mask in public places since **September 2020**?

- Never
- Rarely
- Occasionally
- Often
- Always
- Don't Know

**16.** How often has the child practiced physical in public places between **January-March 2020?**

- Never
- Rarely
- Occasionally
- Often
- Always
- Don't Know

**17.** How often has the child practiced physical distancing in public places between **April-August 2020**?

- Never
- Rarely
- Occasionally
- Often
- Always
- Don't Know

**18.** How often has the child practiced physical distancing in public places since **September 2020**?

- Never
- Rarely
- Occasionally
- Often
- Always
- Don't Know

**19.** How often has the child avoided crowded places/gatherings between **January-March 2020?**

- Never
- Rarely
- Occasionally
- Often
- Always
- Don't Know

**20.** How often has the child avoided crowded places/gatherings between **April-August 2020**?

- Never
- Rarely
- Occasionally
- Often
- Always
- Don't Know

**21.** How often has the child avoided crowded places/gatherings since **September 2020**?

- Never
- Rarely
- Occasionally
- Often
- Always
- Don't Know

**22.** How often has the child avoided common greetings (e.g handshakes, hugs etc) between **January-March 2020**?

- Never
- Rarely
- Occasionally
- Often
- Always
- Don't Know

**23.** How often has the child avoided common greetings (e.g handshakes, hugs etc) between **April-August 2020**?

- Never
- Rarely
- Occasionally
- Often
- Always
- Don't Know

**24.** How often has the child avoided common greetings (e.g handshakes, hugs etc) since **September 2020**?

- Never
- Rarely
- Occasionally
- Often
- Always
- Don't Know

**25.** How often has the child limited contact with people at higher risk (e.g. elderly relative) between **January-March 2020**?

- Never
- Rarely
- Occasionally
- Often
- Always
- Don't Know

**26.** How often has the child limited contact with people at higher risk (e.g. elderly relative) between **April-August 2020**?

- Never
- Rarely
- Occasionally
- Often
- Always
- Don't Know

**27.** How often has the child limited contact with people at higher risk (e.g. elderly relative) since **September 2020**?

- Never
- Rarely
- Occasionally
- Often
- Always
- Don't Know

**28.** How often has the child self-Isolated because someone thought the child was infected with the virus that causes COVID-19 between **January-March 2020**?

- Never
- Rarely
- Occasionally
- Often
- Always
- Don't Know

**29.** How often has the child self-Isolated because someone thought the child was infected with the virus that causes COVID-19 between **April-August 2020**?

- ○ Never
- ○ Rarely
- ○ Occasionally
- ○ Often
- ○ Always
- ○ Don't Know

**30.** How often has the child self-Isolated because someone thought the child was infected with the virus that causes COVID-19 since **September 2020**?

- ○ Never
- ○ Rarely
- ○ Occasionally
- ○ Often
- ○ Always
- ○ Don't Know

**31.** How often has the child quarantined because the child may have been exposed to the virus that causes COVID-19, but did not show symptoms between **January-March 2020**?

- ○ Never
- ○ Rarely
- ○ Occasionally
- ○ Often
- ○ Always
- ○ Don't Know

**32.** How often has the child quarantined because the child may have been exposed to the virus that causes COVID-19, but did not show symptoms between **April-August 2020**?

- ○ Never
- ○ Rarely
- ○ Occasionally
- ○ Often
- ○ Always
- ○ Don't Know

**33.** How often has the child quarantined because the child may have been exposed to the virus that causes COVID-19, but did not show symptoms since **September 2020**?

- ○ Never
- ○ Rarely
- ○ Occasionally
- ○ Often
- ○ Always
- ○ Don't Know

**34.** Has the child participated in any organized sports or recreational activities, in person, since **March 15th 2020**?

- ○ Yes
- ○ No (Go to Question 35)

**34.1** If yes, how many children were in the activity?

○ 0-9 ○ 10-19 ○ 20-29 ○ 30+

**35.** How many times has the child been in a gathering* of >10 or more people (excluding school) since **March 15th 2020**?

____________________

(A gathering in this context is defined as any assembly or meeting of more than 10 persons, in open air or in a building or premises. Gatherings include a range of interpersonal gatherings/events of varying purposes, from gatherings/events of family and friends (e.g., weddings, funerals, baptisms, birthday parties and cultural ceremonies) to community gathering spaces (e.g., places of worship, places of traditional gatherings, libraries, community and recreation centres, drop-in centres, food banks and communal kitchens, museums, theatres, cinemas, and tourist attractions) to larger planned or structured gatherings/events, including mass gatherings (e.g., large meetings or conferences, national sporting events, large cultural/religious events, festivals).

#### During the PAST TWO WEEKS...

| | Not at all worried | Slightly worried | Moderately worried | Very worried | Extremely worried |
|---|---|---|---|---|---|
| **36.** How worried has the child been about being infected by the virus that causes COVID-19? | ○ | ○ | ○ | ○ | ○ |
| **37.** How worried has the child been about friends or family being infected by the virus that causes COVID-19? | ○ | ○ | ○ | ○ | ○ |
| **38.** How worried has the child been about his/her physical health being influenced by COVID-19? | ○ | ○ | ○ | ○ | ○ |
| **39.** How worried has the child been about his/her mental/ emotional health being influenced by COVID-19? | ○ | ○ | ○ | ○ | ○ |
| **40.** How worried was the child generally? | ○ | ○ | ○ | ○ | ○ |

**41.** How much is the child asking questions, reading, or talking about COVID-19?

- ○ Never
- ○ A few times a month
- ○ Weekly
- ○ A few times a week
- ○ Daily

**42.** Has the COVID-19 crisis in your child's area led to any POSITIVE changes in the child's life?

- ○ Yes
- ○ No (Go to Question 43)

**42.1** Please specify these POSITIVE changes in the child's life

- ☐ More time with family
- ☐ More time with friends
- ☐ More exercising
- ☐ Healthier eating
- ☐ New hobbies
- ☐ Other, specify ______________________

#### LIFE CHANGES DUE TO THE COVID-19 CRISIS

| | |
|---|---|
| **43.** Was the child enrolled in school for the previous academic year*?<br>(Previous academic year = Sept 2019 to June 2020) | ◯ Not in school (Go to Question 44)<br>◯ Home school (Go to Question 44)<br>◯ Elementary school<br>◯ Junior High or Middle School<br>◯ Secondary or High School |
| **43.1** Was the child's school building closed due to COVID-19? | ◯ Yes<br>◯ No<br>◯ N/A (Go to Question 44) |
| **43.2** If no, were they attending classes in person? | ◯ Yes (Go to Question 44)<br>◯ No (Go to Question 44)<br>◯ N/A (Go to Question 44) |
| **43.3** If yes, when did the school building closure begin? (DD-MM-YYYY) | ______________________ |
| **43.4** If yes, were classes resumed online? | ◯ Yes<br>◯ No (Go to Question 43.6)<br>◯ N/A (Go to Question 43.6) |
| **43.5** Did the child participate in classes online? | ◯ Yes ◯ Partially<br>◯ No<br>◯ N/A |
| **43.6** If yes, did the child have easy access to the internet and a computer? | ◯ Yes<br>◯ No<br>◯ N/A |
| **43.7** If yes, were there assignments for the child to complete? | ◯ Yes<br>◯ No (Go to Question 43.9)<br>◯ N/A (Go to Question 43.9) |
| **43.8** Did the child complete the assignments? | ◯ Yes ◯ Partially<br>◯ No<br>◯ N/A |
| **43.9** Did the child physically return to school before the summer break? | ◯ Yes<br>◯ No<br>◯ N/A |
| **43.10** If yes, when did the child return to school before the summer break? (DD-MM-YYYY) | ______________________ |
| **44.** Is the child enrolled in school for the current academic year*?<br>(Current academic year = Sept 2020 to June 2021) | ◯ Not in school (Go to Question 45)<br>◯ Home school (Go to Question 45)<br>◯ Elementary school<br>◯ Junior High or Middle School<br>◯ Secondary or High School |
| **44.1** Has the the child's school building closed due to COVID-19 since the start of the current academic year? | ◯ Yes<br>◯ No<br>◯ N/A (Go to Question 44.11) |
| **44.2** If no, are they attending classes in person? | ◯ Yes (Go to Question 44.11)<br>◯ No (Go to Question 44.11)<br>◯ N/A (Go to Question 44.11) |
| **44.3** If yes, when did the school building closure begin? | ______________________ |
| **44.4** If yes, were classes resumed online? | ◯ Yes<br>◯ No (Go to Question 44.6)<br>◯ N/A (Go to Question 44.6) |
| **44.5** Did the child participate in classes online? | ◯ Yes ◯ Partially<br>◯ No<br>◯ N/A |

| | |
|---|---|
| **44.6** If yes, did the child have easy access to the internet and a computer? | ◯ Yes<br>◯ No<br>◯ N/A |
| **44.7** If yes, were there assignments for the child to complete? | ◯ Yes<br>◯ No (Go to Question 44.9)<br>◯ N/A (Go to Question 44.9) |
| **44.8** Did the child complete the assignments? | ◯ Yes ◯ Partially<br>◯ No<br>◯ N/A |
| **44.9** Did the child physically return to school? | ◯ Yes<br>◯ No (Go to Question 44.11)<br>◯ N/A (Go to Question 44.11) |
| **44.10** When did the child physically return to school? (DD-MM-YYYY) | __________________________ |
| **44.11** How many days of school has the child missed/ been absent for during this current academic year? | __________________________ |
| **45.** During the PAST TWO WEEKS, how many people, from outside the child's household, has the child had an in-person conversation with? | __________________________<br>(In-person means that the person is in the same place as the child (NOT on the phone or video chat)) |
| **46.** During the PAST TWO WEEKS, how many times did the child go outside of their home (e.g., going to stores, parks, etc)? | ◯ Not at all<br>◯ 1-2 days per week or less<br>◯ 3-4 days per week<br>◯ 5-6 days per week<br>◯ Daily |
| **47.** During the PAST TWO WEEKS, how stressful have the restrictions on leaving home been for the child? | ◯ Not stressful<br>◯ A little bit stressful<br>◯ Stressful<br>◯ Very stressful<br>◯ Extremely stressful<br>◯ There are no restrictions on leaving home |
| **48.** During the PAST TWO WEEKS, how difficult has it been for the child to follow the recommendations for keeping away from close contact with people who they don't live with? | ◯ Not difficult<br>◯ A little bit difficult<br>◯ Difficult<br>◯ Very difficult<br>◯ Extremely difficult<br>◯ There are no recommendations for keeping away from close contact with people who they don't live with |
| **49.** During the PAST TWO WEEKS, has the quality of the relationships between the child and members of his/her family changed? | ◯ A lot better<br>◯ A little better<br>◯ About the same (Go to Question 50)<br>◯ A little worse<br>◯ A lot worse<br>◯ Don't know (Go to Question 50) |
| **49.1** During the PAST TWO WEEKS, how stressful have these changes in relationships between the child and members of his/her family been for the child? | ◯ Not stressful<br>◯ A little bit stressful<br>◯ Stressful<br>◯ Very stressful<br>◯ Extremely stressful<br>◯ Don't know |

| | |
|---|---|
| **50.** During the PAST TWO WEEKS, has the quality of the child's relationships with his/her friends changed? | ◯ A lot better<br>◯ A little better<br>◯ About the same (Go to Question 51)<br>◯ A little worse<br>◯ A lot worse<br>◯ Don't know (Go to Question 51) |
| **50.1** During the PAST TWO WEEKS, how stressful have these changes in relationships between the child and his/her friends been for the child? | ◯ Not stressful<br>◯ A little bit stressful<br>◯ Stressful<br>◯ Very stressful<br>◯ Extremely stressful<br>◯ Don't Know |
| **51.** During the PAST TWO WEEKS, how difficult has the cancellation of important events in the child's life (such as sporting events, school trips or vacations, etc.) been difficult for the child? | ◯ Not difficult<br>◯ A little bit difficult<br>◯ Difficult<br>◯ Very difficult<br>◯ Extremely difficult<br>◯ There hasn't been any cancellation of important events |
| **52.**How hopeful is the child that the COVID-19 crisis in the child's area will end soon? | ◯ Extremely hopeful<br>◯ Very hopeful<br>◯ Hopeful<br>◯ A little bit hopeful<br>◯ Not at all hopeful<br>◯ There is no COVID-19 crisis in my child's area |

#### 53. Compared to before the COVID-19 outbreak, how much is the child now:

| | Less | Same amount | More |
|---|---|---|---|
| Eating | ○ | ○ | ○ |
| Sleeping | ○ | ○ | ○ |
| Physical activity | ○ | ○ | ○ |
| Spending time outside | ○ | ○ | ○ |
| Spending time with friends in-person | ○ | ○ | ○ |
| Spending time with friends remotely (e.g., online, social media, texting) | ○ | ○ | ○ |
| Spending time watching TV, playing video/computer games, or using social media for educational purposes, including school work | ○ | ○ | ○ |
| Spending time watching TV, playing video/computer games, or using social media for non-educational purposes | ○ | ○ | ○ |

#### DAILY BEHAVIOURS (PAST TWO WEEKS)

**54.** During the PAST TWO WEEKS on average, what time did the child go to bed on WEEKDAYS?

- ○ Before 8 pm
- ○ 8 pm - 10 pm
- ○ 10 pm - 12 am (Midnight)
- ○ After 12 am (Midnight)

**55.** During the PAST TWO WEEKS on average, what time did the child go to bed on WEEKENDS?

- ○ Before 8 pm
- ○ 8 pm - 10 pm
- ○ 10 pm - 12 am (Midnight)
- ○ After 12 am (Midnight)

**56.** During the PAST TWO WEEKS on average, how many hours per night did the child sleep on WEEKDAYS?

- ○ Less than 6 hours
- ○ 6-8 hours
- ○ 8-10 hours
- ○ Greater than 10 hours

**57.** During the PAST TWO WEEKS on average, how many hours per night did the child sleep on WEEKENDS?

- ○ Less than 6 hours
- ○ 6-8 hours
- ○ 8-10 hours
- ○ Greater than 10 hours

**58.** During the PAST TWO WEEKS how many days per week did the child exercise (e.g., increased heart rate, breathing) for at least 30 minutes?

- ○ Not at all
- ○ 1-2 days per week or less
- ○ 3-4 days per week
- ○ 5-6 days per week
- ○ Daily

**59.** During the PAST TWO WEEKS how many days per week did the child spend time outdoors?

- ○ Not at all
- ○ 1-2 days per week or less
- ○ 3-4 days per week
- ○ 5-6 days per week
- ○ Daily

**Study ID:**

#### EMOTIONS/WORRIES (PAST TWO WEEKS)

**60.** During the PAST TWO WEEKS how happy versus sad was the child?

- ◯ Very happy/cheerful
- ◯ Moderately happy/cheerful
- ◯ Neutral
- ◯ Moderately sad/depressed/unhappy
- ◯ Very sad/depressed/unhappy

**61.** During the PAST TWO WEEKS how relaxed versus anxious was the child?

- ◯ Very relaxed/calm
- ◯ Moderately relaxed/calm
- ◯ Neutral
- ◯ Moderately nervous/anxious
- ◯ Very nervous/anxious

**62.** During the PAST TWO WEEKS how fidgety or restless was the child?

- ◯ Not fidgety/restless at all
- ◯ Slightly fidgety/restless
- ◯ Moderately fidgety/restless
- ◯ Very fidgety/restless
- ◯ Extremely fidgety/restless

**63.** During the PAST TWO WEEKS how tired was the child?

- ◯ Not tired at all
- ◯ Slightly tired
- ◯ Moderately tired
- ◯ Very tired
- ◯ Extremely tired

**64.** During the PAST TWO WEEKS how well was the child able to concentrate or focus?

- ◯ Very focused/attentive
- ◯ Moderately focused/attentive
- ◯ Neutral
- ◯ Moderately unfocused/distracted
- ◯ Very unfocused/distracted

**65.** During the PAST TWO WEEKS how irritable or easily angered was the child?

- ◯ Not irritable or easily angered at all
- ◯ Slightly irritable or easily angered
- ◯ Moderately irritable or easily angered
- ◯ Very irritable or easily angered
- ◯ Extremely irritable or easily angered

**66.** During the PAST TWO WEEKS how lonely was the child?

- ◯ Not lonely at all
- ◯ Slightly lonely
- ◯ Moderately lonely
- ◯ Very lonely
- ◯ Extremely lonely

#### MEDIA USE (PAST TWO WEEKS)

**67.** During the PAST TWO WEEKS, how much time per day did the child spend watching TV or digital media (e.g., Netflix, YouTube, web surfing) for educational purposes, including school work?

- ○ No TV or digital media
- ○ Under 1 hour
- ○ 1-3 hours
- ○ 4-6 hours
- ○ More than 6 hours

**68.** During the PAST TWO WEEKS, how much time per day did the child spend watching TV or digital media (e.g., Netflix, YouTube, web surfing) for non-educational purposes?

- ○ No TV or digital media
- ○ Under 1 hour
- ○ 1-3 hours
- ○ 4-6 hours
- ○ More than 6 hours

**69.** During the PAST TWO WEEKS, how much time per day did the child spend using social media (e.g., Facebook, Instagram, Snapchat, Twitter, TikTok)?

- ○ No social media
- ○ Under 1 hour
- ○ 1-3 hours
- ○ 4-6 hours
- ○ More than 6 hours

**70.** During the PAST TWO WEEKS, how much time per day did the child spend playing video games?

(Include time for educational purposes, including school work)

- ○ No video games
- ○ Under 1 hour
- ○ 1-3 hours
- ○ 4-6 hours
- ○ More than 6 hours

**71. Below are statements about families. You are to decide which of these statements are true of your family and which are false. If you think the statement is True or mostly True of your family, mark True. If you think the statement is False or mostly False of your family, mark False.**

**You may feel that some of the statements are true for some family members and false for others. Mark True if the statement is true for most members. Mark False if the statement is false for most members. If the members are evenly divided, decide what is the stronger overall impression and answer accordingly.**

**Remember, we would like to know what your family seems like to you. So do not try to figure out how other members see your family, but do give us your general impression of your family for each statement.**

**Selected Items from Family Environment Scale (FES) Instrument copyright 1974, 2002 by Rudolf H Moos. All rights reserved in all media. Published by Mind Garden, Inc., www.mindgarden.com**

| | True | False |
|---|---|---|
| Family members often criticise each other | ○ | ○ |
| Family members sometimes get so angry they throw things | ○ | ○ |
| Family members often try to one-up or out-do each other | ○ | ○ |
| We fight a lot in our family | ○ | ○ |
| Family members sometimes hit each other | ○ | ○ |
| In our family, we believe you don't ever get anywhere by raising your voice | ○ | ○ |
| If there's a disagreement in our family, we try hard to smooth things over and keep the peace | ○ | ○ |
| Family members hardly ever lose their tempers | ○ | ○ |
| Family members rarely become openly angry | ○ | ○ |

#### DEMOGRAPHIC INFORMATION

If you responded to CHILD Cohort Study questions in the past, you may have already been asked some of the questions contained in this survey. We are asking these questions again because the COVID-19 pandemic may have changed where families are living and who is in their household. We also hope to study how COVID-19 may impact people of different backgrounds and understand how COVID-19 infection can impact a child's medical condition(s) and health. We thank you for your patience in answering questions you may have previously answered as a CHILD Cohort Study participant.

**72.** Thinking about what you know of the child's family history, which of the following best describes the geographic regions from where the child's ancestors (i.e.great-great-grandparents) came from? You may select as many choices as needed

- ☐ England, Ireland, Scotland or Wales
- ☐ Australia - not of Aboriginal or Torres Strait Islander descent
- ☐ Australia - of Aboriginal or Torres Strait Islander descent
- ☐ New Zealand - not of Maori descent
- ☐ New Zealand - of Maori descent
- ☐ Northern Europe including Sweden, Norway, Finland and surrounding countries
- ☐ Western Europe including France, Germany, the Netherlands and surrounding countries
- ☐ Eastern Europe, including Russia, Poland, Hungary and surrounding countries
- ☐ Southern Europe including Italy, Greece, Spain, Portugal and surrounding countries
- ☐ Middle East including Lebanon, Turkey and surrounding countries
- ☐ Eastern Asia including China, Japan, South Korea, North Korea, Taiwan and Hong Kong
- ☐ South-East Asia including Thailand, Malaysia, Indonesia, Singapore and surrounding countries
- ☐ South Asia including India, Pakistan, Sri Lanka and surrounding countries
- ☐ Polynesia, Micronesia or Melanesia including Tonga, Fiji, Papua New Guinea and surrounding countries
- ☐ Africa
- ☐ North America - not of First Nations, Native American, Inuit or Metis descent
- ☐ North America - of First Nations, Native American, Inuit or Metis descent (Answer Question 72.1)
- ☐ Central or South America
- ☐ Don't know
- ☐ Other

**72.1** Which of the following groups does the child belong to?

- ☐ First Nations
- ☐ Inuit
- ☐ Metis
- ☐ Non-status First Nations
- ☐ Other Indigenous
- ☐ Prefer not to answer
- ☐ Don't Know

**73.** Does the child regularly stay at more than one home?

○ Yes ○ No
(Do not include occasional sleepovers at friends or relatives homes)

**74.** Please provide the postal code of the child's primary home

________________________

**75.** How many adults currently live at the child's primary home?

- ○ 0
- ○ 1
- ○ 2
- ○ 3
- ○ 4
- ○ 5
- ○ 6
- ○ 7
- ○ 8
- ○ 9
- ○ 10

**76.** How many children currently live at the child's primary home (excluding the child)?

- ○ 0
- ○ 1
- ○ 2
- ○ 3
- ○ 4
- ○ 5
- ○ 6
- ○ 7
- ○ 8
- ○ 9
- ○ 10

**77.** How many bedrooms are in the child's primary home?

- ○ 0
- ○ 1
- ○ 2
- ○ 3
- ○ 4
- ○ 5
- ○ 6
- ○ 7
- ○ 8
- ○ 9
- ○ 10

**78.** How many toilets are in the child's primary home?

- ○ 0
- ○ 1
- ○ 2
- ○ 3
- ○ 4
- ○ 5
- ○ 6
- ○ 7
- ○ 8
- ○ 9
- ○ 10

**79.** Choose the type of dwelling that best describes the child's primary home

- ○ Manufactured home/ mobile home or trailer
- ○ Single family detached house
- ○ Single family house attached to 1 or more houses (e.g townhouse/ semi-detached)
- ○ High rise apartment/ condo/ co-op (4 or more floors)
- ○ Low rise apartment/ condo/ co-op (1-3 floors)
- ○ Multi-family home (more than one family in a converted single home)
- ○ Other type

**80.** Does the child regularly stay at a secondary home?

○ Yes ○ No (Go to Question 87)
(Do not include occasional sleepovers at friends or relatives homes)

**81.** Please provide the postal code of the child's secondary home

________________________________

**82.** How many adults currently live at the child's secondary home?

- ○ 0
- ○ 1
- ○ 2
- ○ 3
- ○ 4
- ○ 5
- ○ 6
- ○ 7
- ○ 8
- ○ 9
- ○ 10

**83.** How many children currently live at the child's secondary home (excluding the child)?

- ○ 0
- ○ 1
- ○ 2
- ○ 3
- ○ 4
- ○ 5
- ○ 6
- ○ 7
- ○ 8
- ○ 9
- ○ 10

**84.** How many bedrooms are in the child's secondary home?

- ○ 0
- ○ 1
- ○ 2
- ○ 3
- ○ 4
- ○ 5
- ○ 6
- ○ 7
- ○ 8
- ○ 9
- ○ 10

**85.** How many toilets are in the child's secondary home?

- ○ 0
- ○ 1
- ○ 2
- ○ 3
- ○ 4
- ○ 5
- ○ 6
- ○ 7
- ○ 8
- ○ 9
- ○ 10

**86.** Choose the type of dwelling that best describes the child's secondary home

- ○ Manufactured home/ mobile home or trailer
- ○ Single family detached house
- ○ Single family house attached to 1 or more houses (e.g townhouse/ semi-detached)
- ○ High rise apartment/ condo/ co-op (4 or more floors)
- ○ Low rise apartment/ condo/ co-op (1-3 floors)
- ○ Multi-family home (more than one family in a converted single home)
- ○ Other type

| | |
|---|---|
| **87.** Did the child get the flu shot in the past year? | ○ Yes ○ No ○ Don't Know |
| **88.** Are any adults living in the home(s) an ESSENTIAL WORKER? | ☐ Yes, I am<br>☐ Yes, another adult is<br>☐ No (Go to Question 89)<br>☐ Don't Know (Go to Question 89)<br>(e.g., healthcare, delivery worker, store worker, security, building maintenance) |
| **88.1** If yes, do they come home each day? | ☐ Yes, I do<br>☐ Yes, the other adult does<br>☐ No, separated due to COVID-19 (Go to Question 89)<br>☐ No separated due to other reasons (Go to Question 89) |
| **88.2** If yes, are they a FIRST RESPONDER, HEALTHCARE PROVIDER or OTHER WORKER in a facility treating COVID-19? | ☐ Yes, I am<br>☐ Yes, another adult is<br>☐ No<br>☐ Don't Know |

**89.Which of the following medical conditions has the child experienced or received treatment for?(Mark all that apply)**

| | Currently | In the past, but not currently |
|---|---|---|
| Allergies | ☐ | ☐ |
| Anemia (low blood count) | ☐ | ☐ |
| Anxiety | ☐ | ☐ |
| Arthritis | ☐ | ☐ |
| Asthma/ wheeze | ☐ | ☐ |
| Attention Deficient Disorder (ADD, ADHD) | ☐ | ☐ |
| Autism | ☐ | ☐ |
| Behaviour Problems | ☐ | ☐ |
| Bipolar Disorder | ☐ | ☐ |
| Blood Pressure, High | ☐ | ☐ |
| Bronchitis | ☐ | ☐ |
| Cancer, Specify ______________________________ | ☐ | ☐ |
| Cholesterol, High | ☐ | ☐ |
| Chronic Heartburn/ Reflux | ☐ | ☐ |
| Croup | ☐ | ☐ |
| Depression | ☐ | ☐ |
| Diabetes (insulin dependent) | ☐ | ☐ |
| Diabetes (non-insulin dependent) | ☐ | ☐ |
| Epilepsy | ☐ | ☐ |
| Heart Disease | ☐ | ☐ |
| Immune Disorder | ☐ | ☐ |
| IInflammatory Bowel Disease (Crohn's, Ulcerative Colitis) | ☐ | ☐ |
| Irritable Bowel Syndrome (disorder of the intestines with pain and bloating) | ☐ | ☐ |
| Kidney Disease (including kidney stones) | ☐ | ☐ |
| Learning Disorder | ☐ | ☐ |
| Migraine Headaches | ☐ | ☐ |
| Osteoporosis | ☐ | ☐ |
| Pneumonia | ☐ | ☐ |
| Serious acne or skin problems | ☐ | ☐ |
| Severe Obesity (BMI greater-than or equal to 40) | ☐ | ☐ |
| Stroke | ☐ | ☐ |

| | | |
|---|---|---|
| Substance Abuse (excluding alcohol) | ☐ | ☐ |
| Urinary Infections, recurrent | ☐ | ☐ |
| Other, Specify ______________________ | ☐ | ☐ |
| No Medical Conditions | ☐ | ☐ |

**90.** Please share any comments, suggestions or clarifications that you have related to this survey

______________________________

**91.** What is your relationship to the child?

- ○ Biological Mother
- ○ Biological Father
- ○ Parent
- ○ Step Parent
- ○ Foster Parent
- ○ Grandparent
- ○ Great-Grandparent
- ○ Aunt or Uncle
- ○ Sibling
- ○ Step Sibling
- ○ Cousin
- ○ Other, Caregiver

**92.** Today's Date
(DD-MM-YYYY)

______________________

### Adult Self Report

**Study ID:**

We are interested in finding out about how you are feeling and what you are thinking about during the Coronavirus (COVID-19) pandemic.

COVID-19 is a new type of illness affecting many people in Canada. The virus that causes COVID-19 is related to the common cold virus. Some people affected by the virus may have symptoms that can be mild or severe, similar to a cold or flu. Other people with COVID-19 may not have any symptoms at all but they can spread the virus to others.

It is important for scientists to learn more about COVID-19. You can help us by answering this survey. Do the best you can to answer each question. This survey should take about 10 minutes to complete.

If you have questions or want help filling the survey out, please contact your site's Coordinator.

#### COVID-19 HEALTH/EXPOSURE STATUS

**1.** How would you rate your overall physical health before the COVID-19 crisis in your area?

- Excellent
- Very Good
- Good
- Fair
- Poor

**2.** How would you rate your overall mental/ emotional health before the COVID-19 crisis in your area?

- Excellent
- Very Good
- Good
- Fair
- Poor

**3**. In what ways has the COVID-19 outbreak affected your overall healthcare? (Select all that apply)

- ☐ I did not go to healthcare appointments because I was concerned about entering the healthcare providers office
- ☐ My healthcare provider cancelled appointments
- ☐ My healthcare provider changed to phone or online visits
- ☐ My healthcare provider told me to self-isolate or quarantine
- ☐ None of these apply

**4.** Has the COVID-19 crisis in your area led to any family members in your household(s) applying for any of the following government supports? (Select all that apply)

- ☐ Residential Mortgage or Lease/ Rent Payment Deferral
- ☐ Commercial Mortgage or Lease/ Rent Payment Deferral
- ☐ Personal Income Support (e.g Canadian Emergency Response Benefit (CERB), Canadian Emergency Student Benefit (CESB), Employment Insurance (EI))
- ☐ Business Income Support (e.g Canadian Emergency Wage Subsidy (CEWS))
- ☐ Food Bank
- ☐ Other, Specify ______________________
- ☐ None of the above
- ☐ Don't Know

**Study ID:**

| | |
|---|---|
| **5.** Have you been exposed to someone likely to have COVID-19? (Select all that apply) | ☐ Yes, someone with positive test<br>☐ Yes, someone with medical diagnosis, but no test<br>☐ Yes, someone with possible symptoms, but no diagnosis by doctor<br>☐ No, not to my knowledge |
| **6.** Have you been suspected of having COVID-19? | ☐ Yes, has positive test<br>☐ Yes, medical diagnosis, but no test<br>☐ Yes, has had some possible symptoms, but no diagnosis by health care professional<br>☐ No symptoms or signs |
| **7.** Have you ever been tested for the virus that causes COVID-19? (Select all that apply)<br><br>(Do not include the antibody/ serology test (blood test for previous infection) that is part of this research study ) | ☐ No, I never tried to get tested (Answer Question 7.3)<br>☐ No, I tried to get tested but was not able to (Answer Question 7.1)<br>☐ Yes, and I am waiting for the results (Answer Question 7.2)<br>☐ Yes, and the test showed that I did not have it ("negative" test) (Answer Question 7.2)<br>☐ Yes, and the test showed that I did have it ("positive" test) (Answer Question 7.2) |
| **7.1** Which type of test for the virus that causes COVID-19 did you try to get but were not able to?<br><br>(Antibody/ Serology testing and saliva/ mouth rinse testing may not be currently available in all areas, but it may be available in the future.) | ☐ Viral Test (swab test for current infection)<br>☐ Antibody/ Serology Test (blood test for previous infection)<br>☐ Saliva or Mouth Rinse Test<br>☐ Other Specify ______________________<br>☐ Don't Know |
| **7.2** Please specify why you got tested or tried to get tested | ☐ A contact or possible contact of mine tested positive for COVID-19<br>☐ I had COVID-19 symptoms<br>☐ I was admitted to hospital (unrelated to COVID-19)<br>☐ I was admitted to hospital (COVID-19 related)<br>☐ I was tested in another facility or instiution context (contact with someone in long term care, prison etc)<br>☐ I was participating in another facility research study<br>☐ Travel-related testing<br>☐ Routine workplace testing (health care workplace)<br>☐ Routine workplace testing (non-health care workplace)<br>☐ Other, specify ______________________<br>☐ Don't Know |
| **7.3** Please specify why you didn't try to get tested | ☐ Symptoms were due to another condition, specify __________<br>☐ Symptoms were mild<br>☐ A healthcare provider did not recommend testing<br>☐ Other, specify ______________________ |

| | |
|---|---|
| **7.4** In the past two weeks how many times have you been tested for COVID-19? | ○ 0 (Go to Question 8)<br>○ 1<br>○ 2<br>○ 3<br>○ 4 |
| Details of First COVID-19 Test | |
| **7.5** What type of test did you receive?<br><br>(Do not include the antibody/ serology test (blood test for previous infection) that is part of this reseach study ) | ○ Viral Test (swab test for current infection)<br>○ Antibody/ Serology Test (blood test for previous infection)<br>○ Saliva or Mouth Rinse Test<br>○ Other Specify ______________________<br>○ Don't Know |
| **7.6** Date of test<br>(DD-MM-YYYY) | ______________________ |
| **7.7** First test result | ○ Negative ○ Positive<br>○ Don't know |
| Did you complete a second COVID test? | ○ Yes<br>○ No (Go to Question 8) |
| Details of Second COVID-19 Test | |
| **7.8** What type of test did you receive?<br><br>(Do not include the antibody/ serology test (blood test for previous infection) that is part of this reseach study ) | ○ Viral Test (swab test for current infection)<br>○ Antibody/ Serology Test (blood test for previous infection)<br>○ Saliva or Mouth Rinse Test<br>○ Other Specify ______________________<br>○ Don't Know |
| **7.9** Date of test<br>(DD-MM-YYYY) | ______________________ |
| **7.10** Second test result | ○ Negative ○ Positive<br>○ Don't know |
| Did you complete a third COVID test? | ○ Yes<br>○ No (Go to Question 8) |

Details of Third COVID-19 Test

| | |
|---|---|
| **7.11** What type of test did you receive?<br><br>(Do not include the antibody/ serology test (blood test for previous infection) that is part of this reseach study ) | ○ Viral Test (swab test for current infection)<br>○ Antibody/ Serology Test (blood test for previous infection)<br>○ Saliva or Mouth Rinse Test<br>○ Other Specify ____________<br>○ Don't Know |
| **7.12** Date of test<br>(DD-MM-YYYY) | ____________ |
| **7.13** Third test result | ○ Negative ○ Positive<br>○ Don't know |
| Did you complete a fourth COVID test? | ○ Yes<br>○ No (Go to Question 8) |

Details of Fourth COVID-19 Test

| | |
|---|---|
| **7.14** What type of test did you receive?<br><br>(Do not include the antibody/ serology test (blood test for previous infection) that is part of this reseach study ) | ○ Viral Test (swab test for current infection)<br>○ Antibody/ Serology Test (blood test for previous infection)<br>○ Saliva or Mouth Rinse Test<br>○ Other Specify ____________<br>○ Don't Know |
| **7.15** Date of test<br>(DD-MM-YYYY) | |
| **7.16** Fourth test result | ○ Negative ○ Positive<br>○ Don't know |
| **8.** Since March 15th 2020, have you had any colds? | ○ Yes ○ No ○ Don't Know |

| | |
|---|---|
| **9.** Since March 15th 2020, have you had any of the following symptoms? (Select all that apply) | ☐ New or worsening cough<br>☐ Shortness of breath<br>☐ Difficulty breathing<br>☐ Fever<br>☐ Chills<br>☐ Muscle or body aches<br>☐ Headache<br>☐ Sore throat<br>☐ New loss of sense of smell or taste<br>☐ Runny nose or nasal congestion*<br>☐ Nausea or vomiting<br>☐ Diarrhea<br>☐ Fatigue<br>☐ Skin Rash<br>☐ Conjunctivitis (pink eye)<br>☐ Other, Specify ______________________<br>☐ None of the above (Go to Question 10)<br><br>(*Runny nose or nasal congestion in the absence of underlying reason for these symptoms such as seasonal allergies, post nasal drip, etc) |
| **9.1** Do you remember the date when you had your first symptom? | ◯ Yes<br>◯ No (Go to Question 9.3)<br>◯ Roughly |
| **9.2** What was the date of your first symptom?<br>(DD-MM-YYYY) | |
| **9.3** In the TWO WEEKS before you had symptoms, did you:(Select all that apply) | ☐ Have contact with someone who tested positive for COVID-19 (Go to Question 9.21)<br>☐ Have contact with someone who likely had COVID-19 (e.g., was not tested but had symptoms; was told by a healthcare provider that he/she likely had it) (Go to Question 9.21)<br>☐ Travel outside the province<br>☐ None of the above (Go to Question 9.21) |
| **9.4** How many places did you travel?<br>(Travel outside the province) | ◯ 0 (Go to Question 9.21)<br>◯ 1<br>◯ 2<br>◯ 3<br>◯ 4 |
| Place 1 | |
| **9.5** Location<br>(e.g. Seattle, Washington OR Toronto, Ontario) | ______________________ |
| **9.6** Start date of travel<br>(DD-MM-YYYY) | ______________________ |
| **9.7** End date of travel<br>(DD-MM-YYYY) | ______________________ |
| **9.8** How? (Check all that apply) | ☐ Car ☐ Plane ☐ Train<br>☐ Ship ☐ Other ______________________ |

| | |
|---|---|
| Did you travel to a second place? | ◯ Yes<br>◯ No (Go to Question 9.21) |
| Place 2 | |
| **9.9** Location<br>(e.g. Portland, Oregon OR Vancouver, BC) | ______________________ |
| **9.10** Start date of travel<br>(DD-MM-YYYY) | ______________________ |
| **9.11** End date of travel<br>(DD-MM-YYYY) | ______________________ |
| **9.12** How? (Check all that apply) | ☐ Car ☐ Plane ☐ Train<br>☐ Ship ☐ Other ______________________ |
| Did you travel to a third place? | ◯ Yes<br>◯ No (Go to Question 9.21) |
| Place 3 | |
| **9.13** Location<br>(e.g. Paris, France OR Banff, Alberta) | ______________________ |
| **9.14** Start date of travel<br>(DD-MM-YYYY) | ______________________ |
| **9.15** End date of travel<br>(DD-MM-YYYY) | ______________________ |
| **9.16** How? (Check all that apply) | ☐ Car ☐ Plane ☐ Train<br>☐ Ship ☐ Other ______________________ |
| Did you travel to a fourth place? | ◯ Yes<br>◯ No (Go to Question 9.21) |
| Place 4 | |
| **9.17** Location<br>(e.g. Portland, Oregon OR Vancouver, BC) | ______________________ |
| **9.18** Start date of travel<br>(DD-MM-YYYY) | ______________________ |
| **9.19** End date of travel<br>(DD-MM-YYYY) | ______________________ |

| | |
|---|---|
| **9.20** How? (Check all that apply) | ☐ Car ☐ Plane ☐ Train<br>☐ Ship ☐ Other ____________________ |
| **9.21** Which of the following occurred as a result of your symptoms? (Select all that apply) | ☐ You spoke to a healthcare provider over the phone, by email, or online<br>☐ You saw a healthcare provider in person, such as in a clinic or doctor's office<br>☐ You saw a healthcare provider in person, such as in urgent care, or in an Emergency Department (ED)<br>☐ You were kept overnight in a hospital because a healthcare provider thought you had COVID-19<br>☐ You were admitted to a hospital's ICU (Intensive Care Unit)<br>☐ You self-isolated or quarantined at home<br>☐ None of the above |
| **10.** Has anyone in your family been diagnosed with COVID-19? (Select all that apply) | ☐ Yes, member of household<br>☐ Yes, non-household member<br>☐ No<br>☐ Don't know |
| **11.** Have any of the following happened to your family members because of COVID-19? (Select all that apply) | ☐ Fallen ill physically<br>☐ Hospitalized<br>☐ Self-isolated or quarantined with symptoms<br>☐ Self-isolated or quarantined without symptoms (e.g., due to possible exposure)<br>☐ Lost job or been laid off from job<br>☐ Reduced ability to earn money<br>☐ Passed away<br>☐ None of the above<br>(Include any family members, not just those living in the same household as you) |
| **12.** Have you been vaccinated against COVID-19? | ○ Yes<br>○ No (Go to Question 13)<br>Answer 'Yes' if you have received at least one dose of the COVID-19 vaccine. Note: Certain types of vaccines require more than one dose to protect against COVID-19. You would have been informed at the time of vaccination if you needed a second dose.) |
| **12.1** How many doses of the COVID-19 vaccine have you received so far? | ○ One dose<br>○ Two doses<br>○ More than two doses |
| **12.2** When did you receive your first dose of the COVID-19 vaccine? | ____________________ |

If you received more than one dose, go to Question 12.3. If not, continue to Question 12.4.

| | |
|---|---|
| **12.3** When did you receive your second dose of the COVID-19 vaccine? | ____________________ |
| **12.4** Which vaccine did you receive? | ○ Pfizer and BioNTech mRNA vaccine<br>○ Moderna mRNA vaccine<br>○ AstraZeneca Oxford vaccine<br>○ Other, specify____________________<br>○ Don't Know |

#### How often have you done any of the following?

**13.** How often have you worn a mask in public places between **January-March 2020**?

- ○ Never
- ○ Rarely
- ○ Occasionally
- ○ Often
- ○ Always
- ○ Don't Know

**14.** How often have you worn a mask in public places between **April-August 2020**?

- ○ Never
- ○ Rarely
- ○ Occasionally
- ○ Often
- ○ Always
- ○ Don't Know

**15.** How often have you worn a mask in public places since **September 2020**?

- ○ Never
- ○ Rarely
- ○ Occasionally
- ○ Often
- ○ Always
- ○ Don't Know

**16.** How often have you practiced physical distancing in public places between **January-March 2020**?

- ○ Never
- ○ Rarely
- ○ Occasionally
- ○ Often
- ○ Always
- ○ Don't Know

**17.** How often have you practiced physical distancing in public places between **April-August 2020**?

- ○ Never
- ○ Rarely
- ○ Occasionally
- ○ Often
- ○ Always
- ○ Don't Know

**18.** How often have you practiced physical distancing in public places since **September 2020**?

- ○ Never
- ○ Rarely
- ○ Occasionally
- ○ Often
- ○ Always
- ○ Don't Know

**19.** How often have you avoided crowded places/gatherings between **January-March 2020**?

- ○ Never
- ○ Rarely
- ○ Occasionally
- ○ Often
- ○ Always
- ○ Don't Know

**20.** How often have you avoided crowded places/gatherings between **April-August 2020**?

- ○ Never
- ○ Rarely
- ○ Occasionally
- ○ Often
- ○ Always
- ○ Don't Know

**21.** How often have you avoided crowded places/gatherings since **September 2020**?

- Never
- Rarely
- Occasionally
- Often
- Always
- Don't Know

**22.** How often have you avoided common greetings (e.g handshakes, hugs etc) between **January-March 2020**?

- Never
- Rarely
- Occasionally
- Often
- Always
- Don't Know

**23.** How often have you avoided common greetings (e.g handshakes, hugs etc) between **April-August 2020**?

- Never
- Rarely
- Occasionally
- Often
- Always
- Don't Know

**24.** How often have you avoided common greetings (e.g handshakes, hugs etc) since **September 2020**?

- Never
- Rarely
- Occasionally
- Often
- Always
- Don't Know

**25.** How often have you limited contact with people at higher risk (e.g. elderly relative) between **January-March 2020**?

- Never
- Rarely
- Occasionally
- Often
- Always
- Don't Know

**26.** How often have you limited contact with people at higher risk (e.g. elderly relative) between **April-August 2020**?

- Never
- Rarely
- Occasionally
- Often
- Always
- Don't Know

**27.** How often have you limited contact with people at higher risk (e.g. elderly relative) since **September 2020**?

- Never
- Rarely
- Occasionally
- Often
- Always
- Don't Know

**28.** How often have you self-Isolated because someone thought you were infected with the virus that causes COVID-19 between **January-March 2020**?

- Never
- Rarely
- Occasionally
- Often
- Always
- Don't Know

| | |
|---|---|
| **29.** How often have you self-Isolated because someone thought you were infected with the virus that causes COVID-19 between **April-August 2020**? | ○ Never<br>○ Rarely<br>○ Occasionally<br>○ Often<br>○ Always<br>○ Don't Know |
| **30.** How often have you self-Isolated because someone thought you were infected with the virus that causes COVID-19 since **September 2020**? | ○ Never<br>○ Rarely<br>○ Occasionally<br>○ Often<br>○ Always<br>○ Don't Know |
| **31.** How often have you quarantined because you may have been exposed to the virus that causes did not show symptoms between **January-March 2020**? | ○ Never<br>○ Rarely<br>○ Occasionally<br>○ Often<br>○ Always<br>○ Don't Know |
| **32.** How often have you quarantined because you may have been exposed to the virus that causes but did not show symptoms between **April-August** | ○ Never<br>○ Rarely<br>○ Occasionally<br>○ Often<br>○ Always<br>○ Don't Know |
| **33.** How often have you quarantined because you may have been exposed to the virus that causes COVID-19, but did not show symptoms since **September 2020**? | ○ Never<br>○ Rarely<br>○ Occasionally<br>○ Often<br>○ Always<br>○ Don't Know |
| **34.** Have you participated in any organized sports or recreational activities, in person, since **March 15th 2020**? | ○ Yes<br>○ No (Go to Question 35) |
| **34.1** If yes, how many people were in the activity? | ○ 0-9 ○ 10-19 ○ 20-29<br>○ 30+ |
| **35.** How many times have you been in a gathering of >10 or more people (excluding school or work) since **March 15th 2020**? | ____________________<br>(A gathering in this context is defined as any assembly or meeting of more than 10 persons, in open air or in a building or premises. Gatherings include a range of interpersonal gatherings/events of varying purposes, from gatherings/events of family and friends (e.g., weddings, funerals, baptisms, birthday parties and cultural ceremonies) to community gathering spaces (e.g., places of worship, places of traditional gatherings, libraries, community and recreation centres, drop-in centres, food banks and communal kitchens, museums, theatres, cinemas, and tourist attractions) to larger planned or structured gatherings/events, including mass gatherings (e.g., large meetings or conferences, national sporting events, large cultural/religious events, festivals). |

**Study ID:**

#### During the PAST TWO WEEKS...

| | Not at all worried | Slightly worried | Moderately worried | Very worried | Extremely worried |
|---|---|---|---|---|---|
| **36.** How worried have you been about being infected by the virus that causes COVID-19? | ○ | ○ | ○ | ○ | ○ |
| **37.** How worried have you been about friends or family being infected by the virus that causes COVID-19? | ○ | ○ | ○ | ○ | ○ |
| **38.** How worried have you been about your physical health being influenced by COVID-19? | ○ | ○ | ○ | ○ | ○ |
| **39.** How worried have you been about your mental/emotional health being influenced by COVID-19? | ○ | ○ | ○ | ○ | ○ |
| **40.** How worried were you generally? | ○ | ○ | ○ | ○ | ○ |

**41.** How much are you reading or talking about COVID-19?

- ○ Never
- ○ A few times a month
- ○ Weekly
- ○ A few times a week
- ○ Daily

**42.** Has the COVID-19 crisis in your area led to any POSITIVE changes in your life?

- ○ Yes
- ○ No (Go to Question 43)

**42.1** Please specify these POSITIVE changes in your life (Select all that apply)

- ☐ More time with family
- ☐ More time with friends
- ☐ More exercising
- ☐ Healthier eating
- ☐ New hobbies
- ☐ Other, specify______________________________

#### LIFE CHANGES DUE TO THE COVID-19 CRISIS

| | |
|---|---|
| **43.** Were you enrolled in school for the previous academic year*?<br>(Previous academic year = Sept 2019 to June 2020 ) | ○ Not in school (Go to Question 44)<br>○ Home school (Go to Question 44)<br>○ Elementary school<br>○ Junior High or Middle School<br>○ Secondary or High School<br>○ College<br>○ |
| **43.1** Was your school building closed due to COVID-19? | ○ Yes<br>○ No<br>○ N/A (Go to Question 44) |
| **43.2** If no, were you attending classes in person? | ○ Yes (Go to Question 44)<br>○ No (Go to Question 44)<br>○ N/A (Go to Question 44) |
| **43.3** If yes, when did the school building closure begin?<br>(DD-MM-YYYY) | ______________________ |
| **43.4** If yes, were classes resumed online? | ○ Yes<br>○ No (Go to Question 43.6)<br>○ N/A (Go to Question 43.6) |
| **43.5** Did you participate in classes online? | ○ Yes ○ Partially<br>○ No<br>○ N/A |
| **43.6** If yes, did you have easy access to the internet and a computer? | ○ Yes<br>○ No<br>○ N/A |
| **43.7** If yes, were there assignments for you to complete? | ○ Yes<br>○ No (Go to Question 43.9)<br>○ N/A (Go to Question 43.9) |
| **43.8** Did you complete the assignments? | ○ Yes ○ Partially<br>○ No<br>○ N/A |
| **43.9** Did you physically return to school before the summer break? | ○ Yes<br>○ No<br>○ N/A |
| **43.10** If yes, when did the child return to school before the summer break?<br>(DD-MM-YYYY) | ______________________ |
| **44.** Are you enrolled in school for the current academic year*?<br>(Current academic year = Sept 2020 to June 2021) | ○ Not in school (Go to Question 45)<br>○ Home school (Go to Question 45)<br>○ Elementary school<br>○ Junior High or Middle School<br>○ Secondary or High School<br>○ College<br>○ University |
| **44.1** Has your school building closed due to COVID-19 since the start of the current academic year? | ○ Yes<br>○ No<br>○ N/A (Go to Question 44.11) |
| **44.2** If no, are you attending classes in person? | ○ Yes (Go to Question 44.11)<br>○ No (Go to Question 44.11)<br>○ N/A (Go to Question 44.11) |
| **44.3** If yes, when did the school building closure begin? | ______________________ |
| **44.4** If yes, were classes resumed online? | ○ Yes<br>○ No (Go to Question 44.6)<br>○ N/A (Go to Question 44.6) |
| **44.5** Did you participate in classes online? | ○ Yes ○ Partially<br>○ No<br>○ N/A |

| | |
|---|---|
| **44.6** If yes, did you have easy access to the internet and a computer? | ○ Yes<br>○ No<br>○ N/A |
| **44.7** If yes, were there assignments for you to complete? | ○ Yes<br>○ No (Go to Question 44.9)<br>○ N/A (Go to Question 44.9) |
| **44.8** Did you complete the assignments? | ○ Yes<br>○ N/A<br>○ No<br>○ Partially |
| **44.9** Did you physically return to school? | ○ Yes<br>○ No (Go to Question 44.11)<br>○ N/A (Go to Question 44.11) |
| **44.10** When did you return to school? | ________________ (DD-MM-YYYY) |
| **44.11** How many days of school have you missed/ been absent for during this current academic year? | ________________ |
| **45.** Are you currently working? | ☐ Working for pay<br>☐ On paid leave<br>☐ On unpaid leave<br>☐ Laid off or lost job<br>☐ Unemployed and looking for a job<br>☐ Retired<br>☐ Staying at home/ homemaker<br>☐ Other |
| **46.** In what ways has the COVID-19 outbreak affected your work? (Select all that apply) | ☐ I moved to working remotely or from home<br>☐ I lost my job permanently<br>☐ I lost my job temporarily, or was not told for how long<br>☐ I got a new job<br>☐ I reduced my work hours<br>☐ I increased my work hours<br>☐ My job put me at increased risk of getting COVID-19<br>☐ I laid off employees<br>☐ I did not have a paying job before the COVID-19 outbreak<br>☐ None of these apply |
| **47.** In what ways has the COVID-19 outbreak affected your spouse/partners work? (Select all that apply) | ☐ Not applicable I do not have a spouse/partner<br>☐ My spouse/partner moved to working remotely or from home<br>☐ My spouse/partner lost his/her job permanently<br>☐ My spouse/partner lost his/her job temporarily, or was not told for how long<br>☐ My spouse/partner got a new job<br>☐ My spouse/partner reduced his/her work hours<br>☐ My spouse/partner increased his/her work hours<br>☐ My spouse/partners job put him/her at increased risk of getting COVID-19<br>☐ My spouse/partner laid off employees<br>☐ My spouse/partner did not have a paying job before the COVID-19 outbreak<br>☐ None of these apply |
| **48.** During the PAST TWO WEEKS, how many people, from outside of your household, have you had an in-person conversation with? | ________________<br>(In-person means that the person is in the same place as you (NOT on the phone or video chat)) |

| | |
|---|---|
| **49.** During the PAST TWO WEEKS, how many times did you go out outside of your home (e.g., going to stores, parks, etc.)? | ○ Not at all<br>○ 1-2 days per week or less<br>○ 3-4 days per week<br>○ 5-6 days per week<br>○ Daily |
| **50.** During the PAST TWO WEEKS, how stressful have the restrictions on leaving home been for you? | ○ Not stressful<br>○ A little bit stressful<br>○ Stressful<br>○ Very stressful<br>○ Extremely stressful<br>○ There are no restrictions on leaving home |
| **51.** During the PAST TWO WEEKS, how difficult has it been for you to follow the recommendations for keeping away from close contact with people who you don't live with? | ○ Not difficult<br>○ A little bit difficult<br>○ Difficult<br>○ Very difficult<br>○ Extremely difficult<br>○ There are no recommendations for keeping away from close contact with people who I don't live with |
| **52.** During the PAST TWO WEEKS, has the quality of the relationships between you and members of your family changed? | ○ A lot better<br>○ A little better<br>○ About the same (Go to Question 53)<br>○ A little worse<br>○ A lot worse |
| **52.1** During the PAST TWO WEEKS, how stressful have these changes in relationships between you and members of your family been for you? | ○ Not stressful<br>○ A little bit stressful<br>○ Stressful<br>○ Very stressful<br>○ Extremely stressful |
| **53.** During the PAST TWO WEEKS, has the quality of your relationships with your friends changed? | ○ A lot better<br>○ A little better<br>○ About the same (Go to Question 54)<br>○ A little worse<br>○ A lot worse |
| **53.1** During the PAST TWO WEEKS, how stressful have these changes in relationships between you and your friends been for you? | ○ Not stressful<br>○ A little bit stressful<br>○ Stressful<br>○ Very stressful<br>○ Extremely stressful |
| **54.** During the PAST TWO WEEKS how difficult has the cancellation of important events in your life (such as weddings, vacations, etc.) been for you? | ○ Not difficult<br>○ A little bit difficult<br>○ Difficult<br>○ Very difficult<br>○ Extremely difficult<br>○ There hasn't been any cancellation of important events |
| **55.** During the PAST TWO WEEKS, to what degree have changes related to the COVID-19 crisis in your area created financial problems for your family? | ○ No financial problems<br>○ A few financial problems<br>○ Some financial problems<br>○ A lot of financial problems<br>○ Extreme financial problems |

**56.** During the PAST TWO WEEKS, to what degree are you concerned about the stability of your living situation?

- Not at all concerned
- A little concerned
- Concerned
- Very concerned
- Extremely concerned

(Stability of your living situation refers to forced moves that are the result of poor housing quality, unstable neighbourhood conditions and high costs of housing in relation to income. )

**57.** During the PAST TWO WEEKS, did you worry your food would run out because of a lack of money?

- Yes
- No

**58.** How hopeful are you that the COVID-19 crisis in your area will end soon?

- Extremely hopeful
- Very hopeful
- Hopeful
- A little bit hopeful
- Not at all hopeful
- There is no COVID-19 crisis in my area

**59. Compared to before the COVID-19 outbreak, how much are you now:**

| | Less | Same amount | More |
|---|---|---|---|
| Eating | ○ | ○ | ○ |
| Sleeping | ○ | ○ | ○ |
| Physical activity | ○ | ○ | ○ |
| Spending time outside | ○ | ○ | ○ |
| Spending time with friends in-person | ○ | ○ | ○ |
| Spending time with friends remotely (e.g., online, social media, texting) | ○ | ○ | ○ |
| Spending time watching TV, playing video/computer games, or using social media for educational or work purposes, including school work | ○ | ○ | ○ |
| Spending time watching TV, playing video/computer games, or using social media for non-educational or non-work purposes | ○ | ○ | ○ |

#### DAILY BEHAVIORS (PAST TWO WEEKS)

**60.** During the PAST TWO WEEKS on average, what time did you go to bed on WEEKDAYS?

- ○ Before 8 pm
- ○ 8 pm - 10 pm
- ○ 10 pm - 12 am (Midnight)
- ○ After 12 am (Midnight)

**61.** During the PAST TWO WEEKS on average, what time did you go to bed on WEEKENDS?

- ○ Before 8 pm
- ○ 8 pm - 10 pm
- ○ 10 pm - 12 am (Midnight)
- ○ After 12 am (Midnight)

**62.** During the PAST TWO WEEKS on average, how many hours per night did you sleep on WEEKDAYS?

- ○ Less than 6 hours
- ○ 6-8 hours
- ○ 8-10 hours
- ○ Greater than 10 hours

**63.** During the PAST TWO WEEKS on average, how many hours per night did you sleep on WEEKENDS?

- ○ Less than 6 hours
- ○ 6-8 hours
- ○ 8-10 hours
- ○ Greater than 10 hours

**64.** During the PAST TWO WEEKS how many days per week did you exercise (e.g., increased heart rate, breathing) for at least 30 minutes?

- ○ Not at all
- ○ 1-2 days per week or less
- ○ 3-4 days per week
- ○ 5-6 days per week
- ○ Daily

**65.** During the PAST TWO WEEKS how many days per week did you spend time outdoors?

- ○ Not at all
- ○ 1-2 days per week or less
- ○ 3-4 days per week
- ○ 5-6 days per week
- ○ Daily

**66.** How has the COVID-19 outbreak affected your regular childcare? (Select all that apply)

- ☐ I had difficulty arranging for childcare
- ☐ I had to pay more for childcare
- ☐ My spouse/partner or I had to change our work schedule to care for our children ourselves
- ☐ My spouse/partner or I had to reduce our paid work hours to care for our children ourselves
- ☐ My regular childcare has not been affected by the COVID-19 outbreak
- ☐ I do not have a child in childcare

**Study ID:**

#### EMOTIONS/WORRIES (PAST TWO WEEKS)

**67.** During the PAST TWO WEEKS how happy versus sad were you?

- Very happy/cheerful
- Moderately happy/cheerful
- Neutral
- Moderately sad/depressed/unhappy
- Very sad/depressed/unhappy

**68.** During the PAST TWO WEEKS how relaxed versus anxious were you?

- Very relaxed/calm
- Moderately relaxed/calm
- Neutral
- Moderately nervous/anxious
- Very nervous/anxious

**69.** During the PAST TWO WEEKS how fidgety or restless were you?

- Not fidgety/restless at all
- Slightly fidgety/restless
- Moderately fidgety/restless
- Very fidgety/restless
- Extremely fidgety/restless

**70.** During the PAST TWO WEEKS how tired were you?

- Not tired at all
- Slightly tired
- Moderately tired
- Very tired
- Extremely tired

**71.** During the PAST TWO WEEKS how well were you able to concentrate or focus?

- Very focused/attentive
- Moderately focused/attentive
- Neutral
- Moderately unfocused/distracted
- Very unfocused/distracted

**72.** During the PAST TWO WEEKS how irritable or easily angered were you?

- Not irritable or easily angered at all
- Slightly irritable or easily angered
- Moderately irritable or easily angered
- Very irritable or easily angered
- Extremely irritable or easily angered

**73.** During the PAST TWO WEEKS how lonely were you?

- Not lonely at all
- Slightly lonely
- Moderately lonely
- Very lonely
- Extremely lonely

**74.** What have been your greatest sources of stress from the COVID-19 outbreak? (Select all that apply)

- ☐ Health concerns
- ☐ Financial concerns
- ☐ Impact on work
- ☐ Impact on your child
- ☐ Impact on your community
- ☐ Impact on family members
- ☐ Impact on the economy
- ☐ Access to food
- ☐ Access to baby supplies (e.g., formula, diapers, wipes)
- ☐ Access to personal care products or household supplies
- ☐ Access to medical care, including mental health care
- ☐ Social distancing or being quarantined
- ☐ Other
- ☐ I am not stressed about the COVID-19 outbreak

**75.** What have you done to cope with your stress related to the COVID-19 outbreak? (Select all that apply)

- ☐ Meditation and/or mindfulness practices
- ☐ Talking with friends and family (e.g., by phone, text, or video)
- ☐ Engaging in more family activities (e.g., games, sports)
- ☐ Increased television watching or other screen time activities (e.g., video games, social media)
- ☐ Eating more often, including snacking
- ☐ Increasing time reading books, or doing activities like puzzles and crosswords
- ☐ Drinking alcohol
- ☐ Using tobacco (e.g., smoking, vaping)
- ☐ Using marijuana (e.g., vaping, smoking, eating) or cannabidiol (CBD)
- ☐ Talking to my healthcare providers more frequently, including mental healthcare provider (e.g., therapist, psychologist, counselor)
- ☐ Volunteer work
- ☐ Exercise
- ☐ Other
- ☐ I have not done any of these things to cope with the stress related to the COVID-19 outbreak
- ☐ I am not stressed about the COVID-19 outbreak

#### MEDIA USE (PAST TWO WEEKS)

**76.** During the PAST TWO WEEKS, how much time per day did you spend watching TV or digital media (e.g., Netflix, YouTube, web surfing) for educational or work purposes, including school work?

- No TV or digital media
- Under 1 hour
- 1-3 hours
- 4-6 hours
- More than 6 hours

**77.** During the PAST TWO WEEKS, how much time per day did you spend watching TV or digital media (e.g., Netflix, YouTube, web surfing) for non-educational or non- work purposes?

- No TV or digital media
- Under 1 hour
- 1-3 hours
- 4-6 hours
- More than 6 hours

**78.** During the PAST TWO WEEKS, how much time per day did you spend using social media (e.g.,Facebook, Instagram, Snapchat, Twitter, TikTok)?

- No social media
- Under 1 hour
- 1-3 hours
- 4-6 hours
- More than 6 hours

**79.** During the PAST TWO WEEKS, how much time per day did you spend playing video games?88.

- No video games
- Under 1 hour
- 1-3 hours
- 4-6 hours
- More than 6 hours

#### SUBSTANCE USE DURING THE PAST TWO WEEKS

| | Not at all | Rarely | Once a month | Several times a month | Once a week | Several times a week | Once a day | More than once a day |
|---|---|---|---|---|---|---|---|---|
| **80.** How frequently did you use alcohol? | ○ | ○ | ○ | ○ | ○ | ○ | ○ | ○ |
| **81.** How frequently did you use vaping products? | ○ | ○ | ○ | ○ | ○ | ○ | ○ | ○ |
| **82.** How frequently did you use cigarettes or other tobacco products? | ○ | ○ | ○ | ○ | ○ | ○ | ○ | ○ |
| **83.** How frequently did you use marijuana/cannabis (e.g., joint, blunt, pipe, bong)? | ○ | ○ | ○ | ○ | ○ | ○ | ○ | ○ |
| **84.** How frequently did you use opiates, heroin, cocaine, crack, amphetamine, methamphetamine, hallucinogens, or ecstasy? | ○ | ○ | ○ | ○ | ○ | ○ | ○ | ○ |

#### DEMOGRAPHIC INFORMATION

If you responded to CHILD Cohort Study questions in the past, you may have already been asked some of the questions contained in this survey. We are asking these questions again because the COVID-19 pandemic may have changed where families are living and who is in their household. We also hope to study how COVID-19 may impact people of different backgrounds and understand how COVID-19 infection can impact your medical condition(s) and health. We thank you for your patience in answering questions you may have previously answered as a CHILD Cohort Study participant.

**85.** Thinking about what you know of your family history, which of the following best describes the geographic regions from where your ancestors (i.e.great-great-grandparents) came from? You may select as many choices as needed

- ☐ England, Ireland, Scotland or Wales
- ☐ Australia - not of Aboriginal or Torres Strait Islander descent
- ☐ Australia - of Aboriginal or Torres Strait Islander descent
- ☐ New Zealand - not of Maori descent
- ☐ New Zealand - of Maori descent
- ☐ Northern Europe including Sweden, Norway, Finland and surrounding countries
- ☐ Western Europe including France, Germany, the Netherlands and surrounding countries
- ☐ Eastern Europe, including Russia, Poland, Hungary and surrounding countries
- ☐ Southern Europe including Italy, Greece, Spain, Portugal and surrounding countries
- ☐ Middle East including Lebanon, Turkey and surrounding countries
- ☐ Eastern Asia including China, Japan, South Korea, North Korea, Taiwan and Hong Kong
- ☐ South-East Asia including Thailand, Malaysia, Indonesia, Singapore and surrounding countries
- ☐ South Asia including India, Pakistan, Sri Lanka and surrounding countries
- ☐ Polynesia, Micronesia or Melanesia including Tonga, Fiji, Papua New Guinea and surrounding countries
- ☐ Africa
- ☐ North America - not of First Nations, Native American, Inuit or Metis descent
- ☐ North America - of First Nations, Native American, Inuit or Metis descent (Answer Question 85.1)
- ☐ Central or South America
- ☐ Don't know
- ☐ Other

**85.1** Which of the following groups do you belong to?

- ☐ First Nations
- ☐ Inuit
- ☐ Metis
- ☐ Non-status First Nations
- ☐ Other Indigenous
- ☐ Prefer not to answer
- ☐ Don't Know

**86.** What is the highest level of education you have completed?

- ○ Less than high school graduation
- ○ High school graduation
- ○ Trade certificate, vocational school, or apprenticeship training
- ○ Non-university certificate or diploma from a community college, CEGEP
- ○ University Bachelor's degree
- ○ University graduate degree (such as a Masters or Doctorate)
- ○ Prefer not to answer

**87.** Do you regularly stay at more than one home?

○ Yes ○ No
(Do not include occasional sleepovers at friends or relatives homes)

**88.** Please provide the postal code of your primary home

____________________

**89.** How many adults currently live at your primary home (excluding yourself)?

○ 0
○ 1
○ 2
○ 3
○ 4
○ 5
○ 6
○ 7
○ 8
○ 9
○ 10

**90.** How many children currently live at your primary home?

○ 0
○ 1
○ 2
○ 3
○ 4
○ 5
○ 6
○ 7
○ 8
○ 9
○ 10

**91.** How many bedrooms are in your primary home?

○ 0
○ 1
○ 2
○ 3
○ 4
○ 5
○ 6
○ 7
○ 8
○ 9
○ 10

**92.** How many toilets are in your primary home?

○ 0
○ 1
○ 2
○ 3
○ 4
○ 5
○ 6
○ 7
○ 8
○ 9
○ 10

**93.** Choose the type of dwelling that best describes your primary home

- ○ Manufactured home/ mobile home or trailer
- ○ Single family detached house
- ○ Single family house attached to 1 or more houses (e.g townhouse/ semi-detached)
- ○ High rise apartment/ condo/ co-op (4 or more floors)
- ○ Low rise apartment/ condo/ co-op (1-3 floors)
- ○ Multi-family home (more than one family in a converted single home)
- ○ Other type

**94.** Do you regularly stay at a secondary home?

○ Yes ○ No (Go to Question 101)

(Do not include occasional sleepovers at friends or relatives homes)

**95.** Please provide the postal code of your secondary home

______________________________

**96.** How many adults currently live at your secondary home (excluding yourself)?

- ○ 0
- ○ 1
- ○ 2
- ○ 3
- ○ 4
- ○ 5
- ○ 6
- ○ 7
- ○ 8
- ○ 9
- ○ 10

**97.** How many children currently live at your secondary home?

- ○ 0
- ○ 1
- ○ 2
- ○ 3
- ○ 4
- ○ 5
- ○ 6
- ○ 7
- ○ 8
- ○ 9
- ○ 10

**98.** How many bedrooms are in your secondary home?

- ○ 0
- ○ 1
- ○ 2
- ○ 3
- ○ 4
- ○ 5
- ○ 6
- ○ 7
- ○ 8
- ○ 9
- ○ 10

| | |
|---|---|
| **99.** How many toilets are in your secondary home? | ○ 0<br>○ 1<br>○ 2<br>○ 3<br>○ 4<br>○ 5<br>○ 6<br>○ 7<br>○ 8<br>○ 9<br>○ 10 |
| **100.** Choose the type of dwelling that best describes your secondary home | ○ Manufactured home/ mobile home or trailer<br>○ Single family detached house<br>○ Single family house attached to 1 or more houses (e.g townhouse/ semi-detached)<br>○ High rise apartment/ condo/ co-op (4 or more floors)<br>○ Low rise apartment/ condo/ co-op (1-3 floors)<br>○ Multi-family home (more than one family in a converted single home)<br>○ Other type |
| **101.** Did you get the flu shot in the past year? | ○ Yes ○ No ○ Don't Know |
| **102.** Have you been working in any of the following occupations or worksites since **March 15th 2020**?(Select all that apply) | ☐ Hospital or health care facility worker<br>☐ First responder (paramedic, firefighter, police officer)<br>☐ Childcare worker<br>☐ Teacher/ other school staff<br>☐ Transit Driver<br>☐ Food Service Industry<br>☐ Grocery Store<br>☐ Pharmacy<br>☐ Hairdresser/ barber<br>☐ Aesthetician<br>☐ Flight attendant<br>☐ Factory Worker |
| **103.** Are any adults living in any of your home(s) an ESSENTIAL WORKER? | ☐ Yes, I am<br>☐ Yes, another adult is<br>☐ No (Go to Question 104)<br>☐ Don't Know (Go to Question 104)<br>(e.g., healthcare, delivery worker, store worker, security, building maintenance) |
| **103.1** If yes, do they come home each day? | ☐ Yes, I do<br>☐ Yes, the other adult does<br>☐ No, separated due to COVID-19 (Go to Question 104)<br>☐ No separated due to other reasons (Go to Question 104) |
| **103.2** If yes, are they a FIRST RESPONDER, HEALTHCARE PROVIDER or OTHER WORKER in a facility treating COVID-19? | ☐ Yes, I am<br>☐ Yes, another adult is<br>☐ No<br>☐ Don't Know |

**104. Which of the following medical conditions have you experienced or received treatment for?(Mark all that apply)**

| | Currently | In the past, but not currently |
|---|---|---|
| Allergies | ☐ | ☐ |
| Anemia (low blood count) | ☐ | ☐ |
| Anxiety | ☐ | ☐ |
| Arthritis | ☐ | ☐ |
| Asthma/ wheeze | ☐ | ☐ |
| Attention Deficient Disorder (ADD, ADHD) | ☐ | ☐ |
| Autism | ☐ | ☐ |
| Behaviour Problems | ☐ | ☐ |
| Bipolar Disorder | ☐ | ☐ |
| Blood Pressure, High | ☐ | ☐ |
| Bronchitis | ☐ | ☐ |
| Cancer, Specify ________________________________ | ☐ | ☐ |
| Cholesterol, High | ☐ | ☐ |
| Chronic Heartburn/ Reflux | ☐ | ☐ |
| Croup | ☐ | ☐ |
| Depression | ☐ | ☐ |
| Diabetes (insulin dependent) | ☐ | ☐ |
| Diabetes (non-insulin dependent) | ☐ | ☐ |
| Epilepsy | ☐ | ☐ |
| Heart Disease | ☐ | ☐ |
| Immune Disorder | ☐ | ☐ |
| IInflammatory Bowel Disease (Crohn's, Ulcerative Colitis) | ☐ | ☐ |
| Irritable Bowel Syndrome (disorder of the intestines with pain and bloating) | ☐ | ☐ |
| Kidney Disease (including kidney stones) | ☐ | ☐ |
| Learning Disorder | ☐ | ☐ |
| Migraine Headaches | ☐ | ☐ |
| Osteoporosis | ☐ | ☐ |
| Pneumonia | ☐ | ☐ |
| Severe Obesity (BMI greater-than or equal to 40) | ☐ | ☐ |
| Serious acne or skin problems | ☐ | ☐ |
| Stroke | ☐ | ☐ |

| | | |
|---|---|---|
| Substance Abuse (excluding alcohol) | ☐ | ☐ |
| Urinary Infections, recurrent | ☐ | ☐ |
| Other, Specify ______________________ | ☐ | ☐ |
| No Medical Conditions | ☐ | ☐ |

**105.** Please share any comments, suggestions or clarifications that you have related to this survey

______________________________

**106.** Did **you** complete this survey?

Note: **You** refers to an adult (eg. parent or relative of the child participant) in the study

- ○ Yes - by myself
- ○ Yes - partially or with help from someone else
- ○ No - someone else completed this survey on their behalf

**107.** Today's Date
(DD-MM-YYYY)

______________________________

### Child Self Report Follow-up 1



---

ADDENDUM TO PARTICIPANT INFORMATION/CONSENT FORM: Child Self Report

When you signed the consent document to be part of the study, we told you that we would tell you if there were any changes to the study. We thought the survey, asking questions about you, would take only 10 minutes to complete. We have found out from feedback from other people that it can take up to 30 minutes to complete. You do not have to complete the survey if you do not want to. It is ok to say no. By continuing to answer the questions, you say that it is still ok to try and complete the survey and understand it may take longer to complete.

---

We are interested in learning more about how you are feeling and what you are thinking about during the Coronavirus (COVID-19) pandemic since you completed your first survey on [cv19c_date].

COVID-19 is a new type of disease affecting many people in Canada. The virus that causes COVID-19 can cause runny or stuffy noses, coughing, fever, muscle aches and tiredness. Other people with COVID-19 may not have any symptoms at all but they can spread the virus to others.

It is important for scientists to learn more about COVID-19. You can help us by answering this survey. Do the best you can to answer each question. This survey should take between 20 to 30 minutes to complete.

If you have questions or want help filling the survey out, please ask a family member to help you.

---

In general, how would you rate your overall PHYSICAL health (how healthy your body is)?

- ◯ Excellent
- ◯ Very Good
- ◯ Good
- ◯ Fair
- ◯ Poor
- ◯ I don't know
- ◯ I don't want to answer

---

In general, how would you rate your overall MENTAL/EMOTIONAL health (the way you think and feel)?

- ◯ Excellent
- ◯ Very Good
- ◯ Good
- ◯ Fair
- ◯ Poor
- ◯ I don't know
- ◯ I don't want to answer

---

Have you had any of these health problems, since you filled in the first survey on [cv19c_date]?
Check all of the problems you have had.

- ☐ Cough
- ☐ Shortness of breath
- ☐ Difficulty breathing
- ☐ Fever
- ☐ Chills
- ☐ Muscle or body aches
- ☐ Headache
- ☐ Sore throat
- ☐ New loss of sense of smell or taste
- ☐ Runny nose
- ☐ Other, Specify
- ☐ None of the above
- ☐ I don't know
- ☐ I don't want to answer

What other health problems have you had?

__________________________________
(Enter "666" if you don't know. Enter "555" if you don't want to answer.)

How often are you asking questions, reading, or talking about COVID-19, since you completed the first survey on [cv19c_date]?

- ◯ Never
- ◯ A few times a month
- ◯ Weekly
- ◯ A few times a week
- ◯ Daily
- ◯ I don't know
- ◯ I don't want to answer

Has the COVID-19 crisis in your area led to any POSITIVE (GOOD) changes in your life, since you completed the first survey on [cv19c_date]?

- ◯ Yes
- ◯ No
- ◯ I don't know
- ◯ I don't want to answer

What are the POSITIVE (GOOD) changes in your life? (Select all that apply)

- ☐ More time with family
- ☐ More time with friends
- ☐ More exercising
- ☐ Healthier eating
- ☐ New hobbies
- ☐ Got a new pet
- ☐ Learned new technology
- ☐ Made new friends online
- ☐ Saved money
- ☐ Less commuting
- ☐ Better hand hygiene
- ☐ Less busy / more downtime
- ☐ Working from home
- ☐ Schooling from home
- ☐ More time outdoors
- ☐ Not getting sick as often
- ☐ Avoiding bullying
- ☐ Home improvements
- ☐ Improved mental health
- ☐ Other, specify
- ☐ I don't know
- ☐ I don't want to answer

What are the other POSITIVE (GOOD) changes in your life?

__________________________________
(Enter "666" if you don't know. Enter "555" if you don't want to answer.)

A vaccine is a type of medicine that doesn't make you better, but keeps you from getting sick in the first place. It does this by teaching your body to fight off germs like viruses and bacteria. The COVID-19 vaccine is given by a needle in the arm (shot) that will help your body be prepared to fight the COVID-19 virus in case you are exposed to it out in the world.

| Question | Response |
| --- | --- |
| Have you had at least one dose (shot) of the COVID-19 vaccine? | ◯ Yes<br>◯ No<br>◯ I don't know<br>◯ I don't want to answer<br>(Answer 'Yes' if you have received at least one dose of the COVID-19 vaccine. Note: Certain types of vaccines require more than one dose to protect against COVID-19. You would have been informed at the time of vaccination if you needed a second dose.) |
| Did you feel any side effects (feeling not well) from the COVID-19 vaccine? For example: sore arm, feeling tired, achy muscles, headache, fever and chills. | ◯ Yes<br>◯ No<br>◯ I don't know<br>◯ I don't want to answer |
| If yes, check off all the side effects you felt. | ☐ Head ache<br>☐ Tiredness<br>☐ Fever or chills<br>☐ Muscle or joint pain<br>☐ Soreness where the vaccine was injected<br>☐ Nausea or vomiting<br>☐ Other<br>☐ I don't know<br>☐ I don't want to answer |
| Please specify other side effects | ______________________________<br>(Enter "666" if you don't know. Enter "555" if you don't want to answer.) |
| How many doses (shots) of the COVID-19 vaccine have you had so far? | ◯ One dose<br>◯ Two doses<br>◯ More than two doses I<br>◯ don't know<br>◯ I don't want to answer |
| When did you receive your first dose of the COVID-19 vaccine? | ______________________________<br>(If you do not recall the exact date, please provide an estimate. Enter "01-01-1900" if you would prefer not to answer.) |
| Which vaccine did you receive? (first dose) | ◯ Pfizer and BioNTech mRNA vaccine<br>◯ Moderna mRNA vaccine AstraZeneca<br>◯ Oxford vaccine Janssen (Johnson &<br>◯ Johnson) vaccine Other, specify<br>◯ I don't know<br>◯ I don't want to answer<br>◯ |
| Specify the other vaccine | ______________________________<br>(Enter "666" if you don't know. Enter "555" if you don't want to answer.) |

| | |
|---|---|
| When did you receive your second dose of the COVID-19 vaccine? | __________________________________<br>(If you do not recall the exact date, please provide an estimate. Enter "01-01-1900" if you would prefer not to answer.) |
| Which vaccine did you receive? (second dose) | ◯ Pfizer and BioNTech mRNA vaccine<br>◯ Moderna mRNA vaccine<br>◯ AstraZeneca Oxford vaccine<br>◯ Janssen (Johnson & Johnson) vaccine<br>◯ Other, specify<br>◯ I don't know<br>◯ I don't want to answer |
| Specify the other vaccine | __________________________________<br>(Enter "666" if you don't know. Enter "555" if you don't want to answer.) |
| When you are able to get the COVID-19 vaccine, how likely is it that you will choose to get it? | ◯ Very Likely<br>◯ Somewhat Likely<br>◯ Somewhat Unlikely<br>◯ Very Unlikely<br>◯ I don't know<br>◯ I don't want to answer |
| What are some of the reasons why you don't want to get the COVID-19 vaccine? | ☐ Not confident in the safety of the vaccine<br>☐ Concern about risks and side effects<br>☐ Will wait until it seems safe to get the vaccine<br>☐ Do not consider it necessary to get the vaccine<br>☐ Do not believe in vaccination<br>☐ Have not yet decided<br>☐ Have a pre-existing medical condition Already<br>☐ had or think I have had COVID-19 Other,<br>☐ Specify<br>☐ I don't know<br>☐ I don't want to answer |
| What are the other reasons you don't want to get the COVID-19 vaccine? | __________________________________<br>(Enter "666" if you don't know. Enter "555" if you don't want to answer.) |

**A vaccine is a shot that you get at the doctor's office so you won't get sick. We are interested in finding out how you feel about vaccines.**

| | Strongly Agree | Agree | Disagree | Strongly Disagree | I don't know | I don't want to answer |
|---|---|---|---|---|---|---|
| Getting the COVID-19 vaccine is important so I can help protect the health of other people. | ◯ | ◯ | ◯ | ◯ | ◯ | ◯ |
| Getting vaccines is a good way to protect myself from getting sick | ◯ | ◯ | ◯ | ◯ | ◯ | ◯ |
| I usually do what my doctor or healthcare provider recommends about getting a vaccine | ◯ | ◯ | ◯ | ◯ | ◯ | ◯ |
| I worry about feeling sick or having side effects after getting a vaccine. | ◯ | ◯ | ◯ | ◯ | ◯ | ◯ |

#### Social Support

**We would like to measure your feelings about how supported you feel. Choose 1 of the 4 responses to show whether you agree or disagree with the statement.**

| | Strongly Agree | Agree | Disagree | Strongly Disagree | I don't know | I don't want to answer |
|---|---|---|---|---|---|---|
| I have family and friends who help me feel safe, secure and happy | ○ | ○ | ○ | ○ | ○ | ○ |
| There is someone I trust whom I would turn to for advice if I were having problems | ○ | ○ | ○ | ○ | ○ | ○ |
| There are people I can count on in an emergency | ○ | ○ | ○ | ○ | ○ | ○ |
| I feel part of a group of people who share my attitudes and beliefs | ○ | ○ | ○ | ○ | ○ | ○ |

**The following are a number of mental health services that you may have accessed. Please check the relevant boxes for each service indicating if you have accessed the service since March 2020 or if you would be interested in accessing the service.**

| | Have accessed this service since March 2020 | Would be interested in accessing this service | Not accessed, not interested | I don't know | I don't want to answer |
|---|---|---|---|---|---|
| Individual therapy (you alone) with a professional (in person or virtual) | ○ | ○ | ○ | ○ | ○ |
| Group therapy (you and others) with a professional (in person or virtual) | ○ | ○ | ○ | ○ | ○ |
| Family therapy (you and your family members) with a professional (in person or virtual) | ○ | ○ | ○ | ○ | ○ |
| Mental health mobile app (general) | ○ | ○ | ○ | ○ | ○ |
| Mental health mobile app to interact with a therapist | ○ | ○ | ○ | ○ | ○ |
| Phone helpline for mental health support (e.g. Crisis Services Canada) | ○ | ○ | ○ | ○ | ○ |
| Online mental health information you looked up yourself | ○ | ○ | ○ | ○ | ○ |

**Families have different routines.**

**We would like to know about your family's routines. Choose 1 of the 4 responses to show whether you agree or disagree with the statement.**

| | Strongly Agree | Agree | Disagree | Strongly Disagree | I don't know | I don't want to answer |
|---|---|---|---|---|---|---|
| Talking with your parents about your day each day | ○ | ○ | ○ | ○ | ○ | ○ |
| Playing with your parents each day | ○ | ○ | ○ | ○ | ○ | ○ |
| Eating dinner together almost every night | ○ | ○ | ○ | ○ | ○ | ○ |
| Letting your parents know where you are going when you are about to leave the house or where you've been when you get home | ○ | ○ | ○ | ○ | ○ | ○ |
| Regular chores | ○ | ○ | ○ | ○ | ○ | ○ |
| Reading books together almost every day | ○ | ○ | ○ | ○ | ○ | ○ |
| Going to bed at the same time every night | ○ | ○ | ○ | ○ | ○ | ○ |

#### LIFE CHANGES DUE TO THE COVID-19 CRISIS

During the PAST TWO WEEKS how many people (who do not live with you) have you talked to in-person in an indoor environment?

__________________________________
(In-person means that the person is in the same place as you (NOT on the phone or video chat) Enter "666" if you don't know. Enter "555" if you don't want to answer.)

During the PAST TWO WEEKS how many people (who do not live with you) have you talked to in-person in an outdoor environment?

__________________________________
(In-person means that the person is in the same place as you (NOT on the phone or video chat) Enter "666" if you don't know. Enter "555" if you don't want to answer.)

During the PAST TWO WEEKS, how many times did you go outside of your home (e.g., going to stores, parks, etc.)?

- ◯ Not at all
- ◯ 1-2 days per week
- ◯ 3-4 days per week
- ◯ 5-6 days per week
- ◯ Every day
- ◯ I don't know
- ◯ I don't want to answer

During the PAST TWO WEEKS, how stressful have the restrictions on leaving home been for you?

- ◯ Not stressful
- ◯ A little bit stressful
- ◯ Stressful
- ◯ Very stressful
- ◯ Extremely stressful
- ◯ There are no restrictions on leaving home
- ◯ I don't know
- ◯ I don't want to answer

During the PAST TWO WEEKS, how difficult has it been for you to follow the recommendations for keeping away from close contact with people who you don't live with?

- ◯ Not difficult
- ◯ A little bit difficult
- ◯ Difficult
- ◯ Very difficult
- ◯ Extremely difficult
- ◯ There are no recommendations for keeping away from close contact with people who I don't live with
- ◯ I don't know
- ◯ I don't want to answer

During the PAST TWO WEEKS, has the quality of the relationships between you and members of your family changed?

- ◯ A lot better
- ◯ A little better
- ◯ About the same
- ◯ A little worse
- ◯ A lot worse
- ◯ I don't know
- ◯ I don't want to answer

During the PAST TWO WEEKS, how stressful have these changes in relationships between you and members of your family been for you?

- ◯ Not stressful
- ◯ A little bit stressful
- ◯ Stressful
- ◯ Very stressful
- ◯ Extremely stressful
- ◯ I don't know
- ◯ I don't want to answer

| | |
|---|---|
| During the PAST TWO WEEKS, has the quality of your relationships with your friends changed? | ◯ A lot better<br>◯ A little better<br>◯ About the same<br>◯ A little worse<br>◯ A lot worse<br>◯ I don't know<br>◯ I don't want to answer |
| During the PAST TWO WEEKS, how stressful have these changes in your relationships with your friends been for you? | ◯ Not stressful<br>◯ A little bit stressful<br>◯ Stressful<br>◯ Very stressful<br>◯ Extremely stressful<br>◯ I don't know<br>◯ I don't want to answer |
| During the PAST TWO WEEKS, how difficult has the cancellation of important events in your life (such as sporting events, school trips or vacations, etc.) been for you? | ◯ Not difficult<br>◯ A little bit difficult<br>◯ Difficult<br>◯ Very difficult<br>◯ Extremely difficult<br>◯ There hasn't been any cancellation of important events<br>◯ I don't know<br>◯ I don't want to answer |
| How hopeful are you that the COVID-19 crisis in your area will end soon? | ◯ Extremely hopeful<br>◯ Very hopeful<br>◯ Hopeful<br>◯ A little bit hopeful<br>◯ Not at all hopeful<br>◯ Neither hopeful nor not hopeful that COVID-19 will improve/resolve<br>◯ There is no COVID-19 crisis in my area<br>◯ I don't know<br>◯ I don't want to answer |

#### DAILY BEHAVIOURS (PAST TWO WEEKS)

| | |
|---|---|
| During the PAST TWO WEEKS, have you experienced problems with sleep? (e.g., trouble falling asleep, staying asleep, waking up much earlier than intended, sleeping alone without a parent/caregiver) | ◯ Yes<br>◯ No<br>◯ I don't know<br>◯ I don't want to answer |
| During the PAST TWO WEEKS on average, what time did you go to bed on WEEKDAYS? | ◯ Before 8 pm<br>◯ 8 pm - 10 pm<br>◯ 10 pm - 12 am (Midnight)<br>◯ After 12 am (Midnight)<br>◯ I don't know<br>◯ I don't want to answer |
| During the PAST TWO WEEKS on average, what time did you go to bed on WEEKENDS? | ◯ Before 8 pm<br>◯ 8 pm - 10 pm<br>◯ 10 pm - 12 am (Midnight)<br>◯ After 12 am (Midnight)<br>◯ I don't know<br>◯ I don't want to answer |
| During the PAST TWO WEEKS on average, how many hours per night did you sleep on WEEKDAYS? | ◯ Less than 6 hours<br>◯ 6-8 hours<br>◯ 8-10 hours<br>◯ Greater than 10 hours<br>◯ I don't know<br>◯ I don't want to answer |
| During the PAST TWO WEEKS on average, how many hours per night did you sleep on WEEKENDS? | ◯ Less than 6 hours<br>◯ 6-8 hours<br>◯ 8-10 hours<br>◯ Greater than 10 hours<br>◯ I don't know<br>◯ I don't want to answer |
| During the PAST TWO WEEKS, how many days per week did you exercise (where your heart rate and breathing increased) for at least 30 minutes? | ◯ None<br>◯ 1-2 days<br>◯ 3-4 days<br>◯ 5-6 days<br>◯ Daily<br>◯ I don't know<br>◯ I don't want to answer |
| During the PAST TWO WEEKS, how many days per week did you spend time outdoors? | ◯ None<br>◯ 1-2 days<br>◯ 3-4 days<br>◯ 5-6 days<br>◯ Daily<br>◯ I don't know<br>◯ I don't want to answer |

#### EMOTIONS/WORRIES (PAST TWO WEEKS)

| | Not at all worried | Slightly worried | Moderately worried | Very worried | Extremely worried | I don't know | I don't want to answer |
|---|---|---|---|---|---|---|---|
| During the PAST TWO WEEKS how worried have you been about being infected by the virus that causes COVID-19? | ◯ | ◯ | ◯ | ◯ | ◯ | ◯ | ◯ |
| During the PAST TWO WEEKS how worried have you been about friends or family being infected by the virus that causes COVID-19? | ◯ | ◯ | ◯ | ◯ | ◯ | ◯ | ◯ |
| During the PAST TWO WEEKS how worried have you been about your physical health (how healthy your body is) being influenced by COVID-19? | ◯ | ◯ | ◯ | ◯ | ◯ | ◯ | ◯ |
| During the PAST TWO WEEKS how worried have you been about your mental/emotional health (how you think and feel) being influenced by COVID-19? | ◯ | ◯ | ◯ | ◯ | ◯ | ◯ | ◯ |
| During the PAST TWO WEEKS how worried were you generally? | ◯ | ◯ | ◯ | ◯ | ◯ | ◯ | ◯ |

During the PAST TWO WEEKS, how happy versus sad were you?

◯ Very happy/cheerful
◯ Moderately happy/cheerful
◯ Neutral
◯ Moderately sad/depressed/unhappy
◯ Very sad/depressed/unhappy
◯ I don't know
◯ I don't want to answer

During the PAST TWO WEEKS, how relaxed versus anxious were you?

◯ Very relaxed/calm
◯ Moderately relaxed/calm
◯ Neutral
◯ Moderately nervous/anxious
◯ Very nervous/anxious
◯ I don't know
◯ I don't want to answer

During the PAST TWO WEEKS, how fidgety or restless were you?

◯ Not fidgety/restless at all
◯ Slightly fidgety/restless
◯ Moderately fidgety/restless
◯ Very fidgety/restless
◯ Extremely fidgety/restless
◯ I don't know
◯ I don't want to answer

| | |
|---|---|
| During the PAST TWO WEEKS, how tired were you? | ◯ Not tired at all<br>◯ Slightly tired<br>◯ Moderately tired<br>◯ Very tired<br>◯ Extremely tired<br>◯ I don't know<br>◯ I don't want to answer |
| During the PAST TWO WEEKS, how well were you able to concentrate or focus? | ◯ Very focused/attentive<br>◯ Moderately focused/attentive<br>◯ Neutral<br>◯ Moderately unfocused/distracted<br>◯ Very unfocused/distracted<br>◯ I don't know<br>◯ I don't want to answer |
| During the PAST TWO WEEKS, how irritable or easily angered were you? | ◯ Not irritable or easily angered at all<br>◯ Slightly irritable or easily angered<br>◯ Moderately irritable or easily angered<br>◯ Very irritable or easily angered<br>◯ Extremely irritable or easily angered<br>◯ I don't know<br>◯ I don't want to answer |
| During the PAST TWO WEEKS, how lonely were you? | ◯ Not lonely at all<br>◯ Slightly lonely<br>◯ Moderately lonely<br>◯ Very lonely Extremely<br>◯ lonely<br>◯ I don't know<br>◯ I don't want to answer |

#### MEDIA USE (PAST TWO WEEKS)

| | |
|---|---|
| During the PAST TWO WEEKS, how much time per day did you spend watching TV or digital media (e.g., Netflix, YouTube, web surfing)? | ◯ No TV or digital media<br>◯ Under 1 hour<br>◯ 1-3 hours<br>◯ 4-6 hours<br>◯ More than 6 hours<br>◯ I don't know<br>◯ I don't want to answer<br>(Including time for educational purposes and/or homework) |
| During the PAST TWO WEEKS, how much time per day did you spend using social media (e.g., Facebook, Instagram, Snapchat, Twitter, TikTok)? | ◯ No social media<br>◯ Under 1 hour<br>◯ 1-3 hours<br>◯ 4-6 hours<br>◯ More than 6 hours<br>◯ I don't know<br>◯ I don't want to answer<br>(Including time for educational purposes and/or homework) |
| During the PAST TWO WEEKS, how much time per day did you spend playing video games? | ◯ No video games<br>◯ Under 1 hour<br>◯ 1-3 hours<br>◯ 4-6 hours<br>◯ More than 6 hours<br>◯ I don't know<br>◯ I don't want to answer |
| Please share any comments, suggestions or clarifications that you have related to this survey | ________________________________ |
| Did [setup_arm_3][cv19binfo_firstname] complete this survey? | ◯ Yes - by themselves<br>◯ Yes - partially or with help from someone else<br>◯ No - someone else completed this survey on their behalf<br>◯ I don't know<br>◯ I don't want to answer |
| Today's Date | __________________________ |

### Parental Follow-up Survey 1



ADDENDUM TO PARTICIPANT INFORMATION/CONSENT FORM: Parent Self Report

Before beginning this research study, you signed an Informed Consent Form describing the study and your rights as a study participant. At that time, we explained that we would tell you about any new information that might affect your health, well-being or willingness to stay in the study. The Follow up survey was stated to take 10 minutes to complete. Based on feedback from participants we would like to inform you that the survey may take up to 30 minutes to complete.

By continuing to complete the survey questionnaire you consent to be in this part of the study and that you have read and understood the information provided above.

---

We are interested in learning more about how the child is feeling and what they are thinking about during the Coronavirus (COVID-19) pandemic since you completed the first survey on [cv19p_date].

COVID-19 is a new type of illness affecting many people in Canada. The virus that causes COVID-19 is related to the common cold virus. Some people affected by the virus may have symptoms that can be mild or severe, similar to a cold or flu. Other people with COVID-19 may not have any symptoms at all but they can spread the virus to others.

It is important for scientists to learn more about COVID-19. You can help us by answering this survey about the child. Do the best you can to answer each question. This survey should take between 20 to 30 minutes to complete.

---

In general, how would you rate the child's overall PHYSICAL health now?

- ◯ Excellent
- ◯ Very Good
- ◯ Good
- ◯ Fair
- ◯ Poor
- ◯ Prefer not to answer

---

In general, how would you rate the child's overall MENTAL/EMOTIONAL health now?

- ◯ Excellent
- ◯ Very Good
- ◯ Good
- ◯ Fair
- ◯ Poor
- ◯ Prefer not to answer

---

In what ways has the COVID-19 outbreak affected the child's overall healthcare, since you completed the first survey? (Select all that apply)

- ☐ The child did not go to healthcare appointments because I was concerned about entering the healthcare provider's office
- ☐ The child's healthcare provider cancelled appointments
- ☐ The child's healthcare provider changed to phone or online visits
- ☐ The child's healthcare provider told him/her to self-isolate or quarantine
- ☐ None of these apply
- ☐ Prefer not to answer

---

What kind of healthcare visits were affected since you completed the first survey? (Select all that apply)

- ☐ Routine visit/ regular checkup
- ☐ Vaccination visit
- ☐ Visit with a health care professional for a specific concern
- ☐ Visit with a specialist
- ☐ I did not miss any healthcare visits
- ☐ Prefer not to answer

| | |
|---|---|
| Has the COVID-19 crisis in the child's area led to any family members in the child's household(s) applying for or accessing any of the following supports, since you completed the first survey? (Select all that apply) | ☐ Residential Mortgage or Lease/ Rent Payment Deferral<br>☐ Commercial Mortgage or Lease/ Rent Payment Deferral<br>☐ Personal Income Support (e.g Canadian Emergency Response Benefit (CERB), Canadian Emergency Student Benefit (CESB), Employment Insurance (EI))<br>☐ Business Income Support (e.g Canadian Emergency Wage Subsidy (CEWS))<br>☐ Food Bank<br>☐ Other, Specify<br>☐ None of the above<br>☐ Don't Know<br>☐ Prefer not to answer |
| Specify other supports | __________________________________<br>(Enter "555" if you would prefer not to answer.) |
| Has the child been exposed to someone likely to have COVID-19, since you completed the first survey? (Select all that apply) | ☐ Yes, to someone with positive test<br>☐ Yes, to someone with medical diagnosis, but no test<br>☐ Yes, to someone with possible symptoms, but no diagnosis by doctor<br>☐ No, not to my knowledge<br>☐ Prefer not to answer |
| Has the child been suspected of having COVID-19, since you completed the first survey? | ◯ Yes, had positive test<br>◯ Yes, had medical diagnosis, but no test<br>◯ Yes, had some possible symptoms, but no diagnosis by health care professional<br>◯ No<br>◯ Don't know<br>◯ Prefer not to answer |
| Has the child been tested for the virus that causes COVID-19, since you completed the first survey? (Select all that apply)<br><br>Do not include the antibody/ serology test (blood test for previous infection) that is part of this research study. | ☐ No, I never tried to get the child tested<br>☐ No, I tried to get the child tested but was not able to<br>☐ Yes, and the child is waiting for the results<br>☐ Yes, and the test showed that the child did not have it ("negative" test)<br>☐ Yes, and the test showed that the child did have it ("positive" test)<br>☐ Prefer not to answer |
| Which type of test for the virus that causes COVID-19 did you try to get for the child but were not able to? | ☐ Viral Test (swab test for current infection)<br>☐ Antibody/ Serology Test (blood test for previous infection)<br>☐ Saliva or Mouth Rinse Test<br>☐ Rapid-Antigen Test<br>☐ Other Specify<br>☐ Don't Know<br>☐ Prefer not to answer<br>(Antibody/ Serology testing and saliva/ mouth rinse testing may not be currently available in all areas, but it may be available in the future.) |
| Specify the other type of test for the virus that causes COVID-19 that you tried to get for the child but were not able to. | __________________________________<br>(Enter "555" if you would prefer not to answer.) |

| | |
|---|---|
| Please specify why the child got tested or tried to get tested | ☐ A contact or possible contact of the child tested positive for COVID-19<br>☐ The child had COVID-19 symptoms<br>☐ The child was admitted to hospital (unrelated to COVID-19)<br>☐ The child was admitted to hospital (COVID-19 related)<br>☐ The child was tested in another facility or instiution context (contact with someone in long term care, prison etc)<br>☐ The child was participating in another facility research study<br>☐ Travel-related testing<br>☐ Other, specify<br>☐ Don't Know<br>☐ Prefer not to answer |
| Please specify the other reason(s) why the child got tested or tried to get tested | __________________________<br>(Enter "555" if you would prefer not to answer.) |
| Please specify why you didn't try to get the child tested | ☐ Symptoms were due to another condition, specify<br>☐ Symptoms were mild<br>☐ A healthcare provider did not recommend testing<br>☐ Other, specify<br>☐ Prefer not to answer |
| Please specify the condition that you believe the symptoms are from | __________________________<br>( Enter "555" if you would prefer not to answer.) |
| Please specify the other reason(s) you didn't try to get the child tested | __________________________<br>( Enter "555" if you would prefer not to answer.) |
| IN THE PAST TWO WEEKS how many times has the child been tested for COVID-19? | ◯ 0<br>◯ 1<br>◯ 2<br>◯ 3<br>◯ 4<br>◯ Prefer not to answer |
| Details of First COVID-19 Test | |
| What type of test did the child receive? | ◯ Viral Test (swab test for current infection)<br>◯ Antibody/ Serology Test (blood test for previous infection)<br>◯ Saliva or Mouth Rinse Test<br>◯ Rapid-Antigen Test<br>◯ Other Specify<br>◯ Don't Know<br>◯ Prefer not to answer<br>(Do not include the antibody/ serology test (blood test for previous infection) that is part of this reseach study ) |
| Specify other type of test the child received. | __________________________<br>( Enter "555" if you would prefer not to answer.) |

| | |
|---|---|
| Date of test | ________________________________<br>(If you do not recall the exact date, please provide an estimate. Enter "01-01-1900" if you would prefer not to answer.) |
| First test result | ◯ Negative<br>◯ Positive<br>◯ Don't know<br>◯ Prefer not to answer |
| Details of Second COVID-19 Test | |
| What type of test did the child receive? | ◯ Viral Test (swab test for current infection)<br>◯ Antibody/ Serology Test (blood test for previous infection)<br>◯ Saliva or Mouth Rinse Test<br>◯ Rapid-Antigen Test<br>◯ Other Specify<br>◯ Don't Know<br>◯ Prefer not to answer<br>(Do not include the antibody/ serology test (blood test for previous infection) that is part of this reseach study ) |
| Specify other type of test the child received. | ________________________________<br>(Enter "555" if you would prefer not to answer.) |
| Date of test | ________________________________<br>(If you do not recall the exact date, please provide an estimate. Enter "01-01-1900" if you would prefer not to answer.) |
| Second test result | ◯ Negative<br>◯ Positive<br>◯ Don't know<br>◯ Prefer not to answer |
| Details of Third COVID-19 Test | |
| What type of test did the child receive? | ◯ Viral Test (swab test for current infection)<br>◯ Antibody/ Serology Test (blood test for previous infection)<br>◯ Saliva or Mouth Rinse Test<br>◯ Rapid-Antigen Test<br>◯ Other Specify<br>◯ Don't Know<br>◯ Prefer not to answer<br>(Do not include the antibody/ serology test (blood test for previous infection) that is part of this reseach study ) |
| Specify other type of test the child received. | ________________________________<br>(Enter "555" if you would prefer not to answer.) |

| | |
|---|---|
| Date of test | __________________________________<br>(If you do not recall the exact date, please provide an estimate. Enter "01-01-1900" if you would prefer not to answer.) |
| Third test result | ◯ Negative<br>◯ Positive<br>◯ Don't know<br>◯ Prefer not to answer |
| Details of Fourth COVID-19 Test | |
| What type of test did the child receive? | ◯ Viral Test (swab test for current infection)<br>◯ Antibody/ Serology Test (blood test for previous infection)<br>◯ Saliva or Mouth Rinse Test<br>◯ Rapid-Antigen Test<br>◯ Other Specify<br>◯ Don't Know<br>◯ Prefer not to answer<br>(Do not include the antibody/ serology test (blood test for previous infection) that is part of this reseach study ) |
| Specify other type of test the child received. | __________________________________<br>(Enter "555" if you would prefer not to answer.) |
| Date of test | __________________________________<br>(If you do not recall the exact date, please provide an estimate. Enter "01-01-1900" if you would prefer not to answer.) |
| Fourth test result | ◯ Negative<br>◯ Positive<br>◯ Don't know<br>◯ Prefer not to answer |
| Has the child had any colds, since you completed the first survey? | ◯ Yes<br>◯ No<br>◯ Don't Know<br>◯ Prefer not to answer |

| | |
|---|---|
| Has the child had any of the following symptoms, since you completed the first survey? (Select all that apply) | ☐ New or worsening cough<br>☐ Shortness of breath<br>☐ Difficulty breathing<br>☐ Fever<br>☐ Chills<br>☐ Muscle or body aches<br>☐ Headache<br>☐ Sore throat<br>☐ New loss of sense of smell or taste<br>☐ Runny nose or nasal congestion*<br>☐ Nausea or vomiting<br>☐ Diarrhea<br>☐ Fatigue<br>☐ Skin Rash<br>☐ Conjunctivitis (pink eye)<br>☐ Other, Specify<br>☐ None of the above<br>☐ Prefer not to answer<br>(*Runny nose or nasal congestion in the absence of underlying reason for these symptoms such as seasonal allergies, post nasal drip, etc) |
| Specify the other symptoms that the child had since you completed the first survey? | __________________________________<br>( Enter "555" if you would prefer not to answer.) |
| Do you remember the date when the child had their first symptom? | ◯ Yes<br>◯ No<br>◯ Roughly<br>◯ Prefer not to answer |
| What was the date of the child's first symptom? | __________________________________<br>(If you do not recall the exact date, please provide an estimate. Enter "01-01-1900" if you would prefer not to answer.) |
| In the TWO WEEKS before the child had symptoms, did the child: (Select all that apply) | ☐ Have contact with someone who tested positive for COVID-19<br>☐ Have contact with someone who likely had COVID-19 (e.g., was not tested but had symptoms; was told by a healthcare provider that he/she likely had it)<br>☐ Travel outside the province<br>☐ None of the above<br>☐ Prefer not to answer |
| How many places did the child travel? | ◯ 0<br>◯ 1<br>◯ 2<br>◯ 3<br>◯ 4<br>◯ Prefer not to answer<br>(Travel outside the province) |
| Place 1 | |
| Location<br>(e.g. Seattle, Washington OR Toronto, Ontario) | __________________________________<br>(Enter "555" if you would prefer not to answer.) |

| | |
|---|---|
| Start date of travel | ______________________<br>(If you do not recall the exact date, please provide an estimate. Enter "01-01-1900" if you would prefer not to answer.) |
| End date of travel | ______________________<br>(If you do not recall the exact date, please provide an estimate. Enter "01-01-1900" if you would prefer not to answer.) |
| How? (Check all that apply) | ☐ Car<br>☐ Plane<br>☐ Train<br>☐ Ship<br>☐ Other<br>☐ Prefer not to answer |
| Please specify other mode of transportation | ______________________<br>(Enter "555" if you would prefer not to answer.) |
| Place 2 | |
| Location<br>(e.g. Portland, Oregon OR Vancouver, BC) | ______________________<br>(Enter "555" if you would prefer not to answer.) |
| Start date of travel | ______________________<br>(If you do not recall the exact date, please provide an estimate. Enter "01-01-1900" if you would prefer not to answer.) |
| End date of travel | ______________________<br>(If you do not recall the exact date, please provide an estimate. Enter "01-01-1900" if you would prefer not to answer.) |
| How? (Check all that apply) | ☐ Car<br>☐ Plane<br>☐ Train<br>☐ Ship<br>☐ Other<br>☐ Prefer not to answer |
| Please specify other mode of transportation | ______________________<br>(Enter "555" if you would prefer not to answer.) |
| Place 3 | |
| Location<br>(e.g. Paris, France OR Banff, Alberta) | ______________________<br>(Enter "555" if you would prefer not to answer.) |

Start date of travel

__________________________________
(If you do not recall the exact date, please provide an estimate. Enter "01-01-1900" if you would prefer not to answer.)

End date of travel

__________________________________
(If you do not recall the exact date, please provide an estimate. Enter "01-01-1900" if you would prefer not to answer.)

How? (Check all that apply)

- ☐ Car
- ☐ Plane
- ☐ Train
- ☐ Ship
- ☐ Other
- ☐ Prefer not to answer

Please specify other mode of transportation

__________________________________
(Enter "555" if you would prefer not to answer.)

Place 4

Location
(e.g. Portland, Oregon OR Vancouver, BC)

__________________________________
(Enter "555" if you would prefer not to answer.)

Start date of travel

__________________________________
(If you do not recall the exact date, please provide an estimate. Enter "01-01-1900" if you would prefer not to answer.)

End date of travel

__________________________________
(If you do not recall the exact date, please provide an estimate. Enter "01-01-1900" if you would prefer not to answer.)

How? (Check all that apply)

- ☐ Car
- ☐ Plane
- ☐ Train
- ☐ Ship
- ☐ Other
- ☐ Prefer not to answer

Please specify other mode of transportation

__________________________________
(Enter "555" if you would prefer not to answer.)

| Question | Options |
|---|---|
| Which of the following occurred as a result of the child's symptoms? (Select all that apply) | ☐ You/ the child spoke to a healthcare provider over the phone, by email, or online<br>☐ The child saw a healthcare provider in person, such as in a clinic or doctor's office<br>☐ The child saw a healthcare provider in person, such as in urgent care, or in an Emergency Department (ED)<br>☐ The child was kept overnight in a hospital because a healthcare provider thought he/she had COVID-19<br>☐ The child was admitted to a hospital's ICU (Intensive Care Unit)<br>☐ The child self-isolated or quarantined at home<br>☐ None of the above<br>☐ Prefer not to answer |
| Has anyone in the child's family been diagnosed with COVID-19, since you completed the first survey? (Select all that apply) | ☐ Yes, member of household<br>☐ Yes, non-household<br>☐ member No<br>☐ Don't know<br>☐ Prefer not to answer |
| Have any of the following happened to the child's family members because of COVID-19, since you completed the first survey? (Select all that apply) | ☐ Fallen physically ill<br>☐ Hospitalized<br>☐ Self-isolated or quarantined with symptoms<br>☐ Self-isolated or quarantined without symptoms (e.g., due to possible exposure)<br>☐ Lost job or been laid off from job<br>☐ Reduced ability to earn money<br>☐ Passed away<br>☐ None of the above<br>☐ Prefer not to answer<br>(Include any family members, not just those living in the same household as the child) |
| Did the child get the flu shot, since you completed the first survey? | ◯ Yes<br>◯ No<br>◯ Don't Know<br>◯ Prefer not to answer |
| Has the child received at least one dose of a COVID-19 vaccine? | ◯ Yes<br>◯ No<br>◯ Prefer not to answer<br>(Answer 'Yes' if the child has received at least one dose of the COVID-19 vaccine. Note: Certain types of vaccines require more than one dose to protect against COVID-19. You or the child would have been informed at the time of vaccination if the child needed a second dose.) |
| Did the child feel any side effects from the COVID-19 vaccine? | ◯ Yes<br>◯ No<br>◯ Prefer not to answer |

| | |
|---|---|
| If yes, check all that apply. | ☐ Head ache<br>☐ Tiredness<br>☐ Fever or chills<br>☐ Muscle or joint pain<br>☐ Soreness where the vaccine was injected<br>☐ Nausea or vomiting<br>☐ Other<br>☐ Prefer not to answer |
| Please specify other side effects | __________________________________<br>(Enter "555" if you would prefer not to answer.) |
| How many doses of the COVID-19 vaccine has the child received so far? | ◯ One dose<br>◯ Two doses<br>◯ More than two doses<br>◯ Prefer not to answer |
| When did the child receive their first dose of the COVID-19 vaccine? | __________________________________<br>(If you do not recall the exact date, please provide an estimate. Enter "01-01-1900" if you would prefer not to answer.) |
| Which vaccine did the child receive? (first dose) | ◯ Pfizer and BioNTech mRNA vaccine<br>◯ Moderna mRNA vaccine<br>◯ AstraZeneca Oxford vaccine Janssen<br>◯ (Johnson & Johnson) vaccine Other,<br>◯ specify<br>◯ Don't Know<br>◯ Prefer not to answer |
| Specify the other vaccine | __________________________________<br>(Enter "555" if you would prefer not to answer.) |
| When did the child receive their second dose of the COVID-19 vaccine? | __________________________________<br>(If you do not recall the exact date, please provide an estimate. Enter "01-01-1900" if you would prefer not to answer.) |
| Which vaccine did the child receive? (second dose) | ◯ Pfizer and BioNTech mRNA vaccine<br>◯ Moderna mRNA vaccine<br>◯ AstraZeneca Oxford vaccine Janssen<br>◯ (Johnson & Johnson) vaccine Other,<br>◯ specify<br>◯ Don't Know<br>◯ Prefer not to answer |
| Specify the other vaccine | __________________________________<br>(Enter "555" if you would prefer not to answer.) |

When a COVID-19 vaccine becomes available, how likely is it that you will choose for THE CHILD to get it?

- ◯ Very Likely
- ◯ Somewhat Likely
- ◯ Somewhat Unlikely
- ◯ Very Unlikely
- ◯ I Don't Know
- ◯ Prefer not to answer

What are some of the reasons why you don't want THE CHILD to get the COVID-19 vaccine?

- ☐ Not confident in the safety of the vaccine
- ☐ Concern about risks and side effects
- ☐ Will wait until it seems safe to get the vaccine
- ☐ Do not consider it necessary to get the vaccine
- ☐ Do not believe in vaccination
- ☐ Have not yet decided
- ☐ Have a pre-existing medical condition Already
- ☐ had or think I have had COVID-19 Other,
- ☐ Specify
- ☐ Prefer not to answer

(Check all that apply.)

What were the other reasons you didn't want THE CHILD to get the COVID-19 vaccine?

__________________________________
( Enter "555" if you would prefer not to answer.)

| | Strongly Agree | Agree | Disagree | Strongly Disagree | Prefer not to answer |
|---|---|---|---|---|---|
| Having the child vaccinated is important for the health of others in my community. | ◯ | ◯ | ◯ | ◯ | ◯ |
| Getting vaccines is a good way to protect the child/children from disease. | ◯ | ◯ | ◯ | ◯ | ◯ |
| Generally I do what my doctor or health care provider recommends about vaccines for the child/children. | ◯ | ◯ | ◯ | ◯ | ◯ |

| | Never | Rarely | Occasionally | Often | Always | Don't Know | Prefer not to answer |
|---|---|---|---|---|---|---|---|
| How often has the child worn a mask in public places since you completed the first survey? | ◯ | ◯ | ◯ | ◯ | ◯ | ◯ | ◯ |
| How often has the child practiced physical distancing in public places since you completed the first survey? | ◯ | ◯ | ◯ | ◯ | ◯ | ◯ | ◯ |
| How often has the child avoided crowded places/ gatherings since you completed the first survey? | ◯ | ◯ | ◯ | ◯ | ◯ | ◯ | ◯ |

| | | | | | | | |
|---|---|---|---|---|---|---|---|
| How often has the child avoided common greetings (e.g handshakes, hugs etc) since you completed the first survey? | ◯ | ◯ | ◯ | ◯ | ◯ | ◯ | ◯ |
| How often has the child limited contact with people at higher risk (e.g. elderly relative) since you completed the first survey? | ◯ | ◯ | ◯ | ◯ | ◯ | ◯ | ◯ |
| How often has the child self-Isolated because someone thought the child was infected with the virus that causes COVID-19 since you completed the first survey? | ◯ | ◯ | ◯ | ◯ | ◯ | ◯ | ◯ |
| How often has the child quarantined because the child may have been exposed to the virus that causes COVID-19, but did not show symptoms since you completed the first survey? | ◯ | ◯ | ◯ | ◯ | ◯ | ◯ | ◯ |

---

Has the child participated in any organized sports or recreational activities, in person, since you completed the first survey?

◯ Yes
◯ No
◯ Prefer not to answer

---

If yes, how many children were in the activity?

◯ 0-9
◯ 10-19
◯ 20-29
◯ 30+
◯ Prefer not to answer

---

How many times has the child been in a gathering* of >10 or more people (excluding school), since you completed the first survey?

__________________________________

(A gathering in this context is defined as any assembly or meeting of more than 10 persons, in open air or in a building or premises. Gatherings include a range of interpersonal gatherings/events of varying purposes, from gatherings/events of family and friends (e.g., weddings, funerals, baptisms, birthday parties and cultural ceremonies) to community gathering spaces (e.g., places of worship, places of traditional gatherings, libraries, community and recreation centres, drop-in centres, food banks and communal kitchens, museums, theatres, cinemas, and tourist attractions) to larger planned or structured gatherings/events, including mass gatherings (e.g., large meetings or conferences, national sporting events, large cultural/religious events, festivals). Enter "555" if you would prefer not to answer.)

**The following are a number of mental health services that the child may have accessed. Please check the relevant boxes for each service indicating if the child accessed the service since March 2020 or if the child would be interested in accessing the service.**

| | Have accessed this service since March 2020 | Would be interested in accessing this service | Not accessed, not interested | Prefer not to answer |
|---|---|---|---|---|
| Individual therapy (you alone) with a professional (in person or virtual) | ◯ | ◯ | ◯ | ◯ |
| Group therapy (you and others) with a professional (in person or virtual) | ◯ | ◯ | ◯ | ◯ |
| Family therapy (you and your family members) with a professional (in person or virtual) | ◯ | ◯ | ◯ | ◯ |
| Mental health mobile app (general) | ◯ | ◯ | ◯ | ◯ |
| Mental health mobile app to interact with a therapist | ◯ | ◯ | ◯ | ◯ |
| Phone helpline for mental health support (e.g. Crisis Services Canada) | ◯ | ◯ | ◯ | ◯ |
| Online mental health information you looked up yourself | ◯ | ◯ | ◯ | ◯ |

**Worrying (PAST TWO WEEKS)**
**You completed your first survey on [cv19p_date].**

| | Not at all worried | Slightly worried | Moderately worried | Very worried | Extremely worried | Prefer not to answer |
|---|---|---|---|---|---|---|
| During the PAST TWO WEEKS how worried has the child been about being infected by the virus that causes COVID-19? | ○ | ○ | ○ | ○ | ○ | ○ |
| During the PAST TWO WEEKS how worried has the child been about friends or family being infected by the virus that causes COVID-19? | ○ | ○ | ○ | ○ | ○ | ○ |
| During the PAST TWO WEEKS how worried has the child been about his/her physical health being influenced by COVID-19? | ○ | ○ | ○ | ○ | ○ | ○ |
| During the PAST TWO WEEKS how worried has the child been about his/her mental/emotional health being influenced by COVID-19? | ○ | ○ | ○ | ○ | ○ | ○ |
| During the PAST TWO WEEKS how worried was the child generally? | ○ | ○ | ○ | ○ | ○ | ○ |

How much is the child asking questions, reading, or talking about COVID-19, since you completed the first survey?

- ○ Never
- ○ A few times a month
- ○ Weekly
- ○ A few times a week
- ○ Daily
- ○ Prefer not to answer

Has the COVID-19 crisis in the child's area led to any POSITIVE changes in the child's life, since you completed the first survey?

- ○ Yes
- ○ No
- ○ Prefer not to answer

Please specify these POSITIVE changes in the child's life

- ☐ More time with family
- ☐ More time with friends
- ☐ More exercising
- ☐ Healthier eating
- ☐ New hobbies
- ☐ Other, specify
- ☐ Prefer not to answer

Please specify the other POSITIVE changes in the child's life

__________________________________
(Enter "555" if you would prefer not to answer.)

| Question | Response |
|---|---|
| Is the child enrolled in school for the current academic year*? | ◯ Not in school<br>◯ Home school<br>◯ Elementary school<br>◯ Junior High or Middle School<br>◯ Secondary or High School<br>◯ Prefer not to answer<br>(* Current academic year = Sept 2020 to June 2021) |
| Has the the child's school building closed since you completed the first survey? | ◯ Yes<br>◯ No<br>◯ N/A<br>◯ Prefer not to answer |
| If no, are they attending classes in person? | ◯ Yes<br>◯ No<br>◯ N/A<br>◯ Prefer not to answer |
| If yes, how many times did the school close? | __________________________<br>(Enter "666" if you don't know the answer.Enter "555" if you would prefer not to answer.) |
| If yes, when did the first school building closure begin? | __________________________<br>(If you do not recall the exact date, please provide an estimate. Enter "01-01-1900" if you would prefer not to answer.) |
| How many weeks in total was the school building closed since September 2020 due to COVID-19? | __________________________<br>((do not include Christmas and spring break) Enter "555" if you would prefer not to answer.) |
| If yes, were classes resumed online? | ◯ Yes<br>◯ No<br>◯ N/A<br>◯ Prefer not to answer |
| Did the child participate in classes online since September 2020? | ◯ Yes<br>◯ No<br>◯ Partially<br>◯ N/A<br>◯ Prefer not to answer |
| If yes, did the child have easy access to the internet and a computer? | ◯ Yes<br>◯ No<br>◯ N/A<br>◯ Prefer not to answer |
| If yes, were there assignments for the child to complete? | ◯ Yes<br>◯ No<br>◯ N/A<br>◯ Prefer not to answer |

| | |
|---|---|
| Did the child complete the assignments? | ◯ Yes<br>◯ No<br>◯ Partially<br>◯ N/A<br>◯ Prefer not to answer |
| Did the child physically return to school since the first closure? | ◯ Yes<br>◯ No<br>◯ N/A<br>◯ Prefer not to answer |
| When did the child physically return to school? | ______________________________<br>(If you do not recall the exact date, please provide an estimate. Enter "01-01-1900" if you would prefer not to answer.) |
| How many days of in-school or remote/online learning has the child missed/ been absent for, since you completed the first survey? | ______________________________<br>(Enter "555" if you would prefer not to answer.) |
| During the PAST TWO WEEKS, how many people, from outside the child's household, has the child had an in-person conversation with in an indoor environment? | ______________________________<br>(In-person means that the person is in the same place as the child (NOT on the phone or video chat) Enter "555" if you would prefer not to answer.) |
| During the PAST TWO WEEKS, how many people, from outside the child's household, has the child had an in-person conversation with in an outdoor environment? | ______________________________<br>(In-person means that the person is in the same place as the child (NOT on the phone or video chat) Enter "555" if you would prefer not to answer.) |
| During the PAST TWO WEEKS, how many times did the child go outside of their home (e.g., going to stores, parks, etc)? | ◯ Not at all<br>◯ 1-2 days per week or less<br>◯ 3-4 days per week<br>◯ 5-6 days per week<br>◯ Daily<br>◯ Prefer not to answer |
| During the PAST TWO WEEKS, how stressful have the restrictions on leaving home been for the child? | ◯ Not stressful<br>◯ A little bit stressful<br>◯ Stressful<br>◯ Very stressful<br>◯ Extremely stressful<br>◯ There are no restrictions on leaving home<br>◯ Prefer not to answer |
| During the PAST TWO WEEKS, how difficult has it been for the child to follow the recommendations for keeping away from close contact with people who they don't live with? | ◯ Not difficult<br>◯ A little bit difficult<br>◯ Difficult<br>◯ Very difficult<br>◯ Extremely difficult<br>◯ There are no recommendations for keeping away from close contact with people who they don't live with<br>◯ Prefer not to answer |

| | |
|---|---|
| During the PAST TWO WEEKS, has the quality of the relationships between the child and members of his/her family changed? | ◯ A lot better<br>◯ A little better<br>◯ About the same<br>◯ A little worse<br>◯ A lot worse<br>◯ Don't know<br>◯ Prefer not to answer |
| During the PAST TWO WEEKS, how stressful have these changes in relationships between the child and members of his/her family been for the child? | ◯ Not stressful<br>◯ A little bit stressful<br>◯ Stressful<br>◯ Very stressful<br>◯ Extremely stressful<br>◯ Don't know<br>◯ Prefer not to answer |
| During the PAST TWO WEEKS, has the quality of the child's relationships with his/her friends changed? | ◯ A lot better<br>◯ A little better<br>◯ About the same<br>◯ A little worse<br>◯ A lot worse<br>◯ Don't know<br>◯ Prefer not to answer |
| During the PAST TWO WEEKS, how stressful have these changes in relationships between the child and his/her friends been for the child? | ◯ Not stressful<br>◯ A little bit stressful<br>◯ Stressful<br>◯ Very stressful<br>◯ Extremely stressful<br>◯ Don't Know<br>◯ Prefer not to answer |
| During the PAST TWO WEEKS, how difficult has the cancellation of important events in the child's life (such as sporting events, school trips or vacations, etc.) been difficult for the child? | ◯ Not difficult<br>◯ A little bit difficult<br>◯ Difficult<br>◯ Very difficult<br>◯ Extremely difficult<br>◯ There hasn't been any cancellation of important events<br>◯ Prefer not to answer |
| How hopeful is the child that the COVID-19 crisis in the child's area will end soon? | ◯ Extremely hopeful<br>◯ Very hopeful<br>◯ Hopeful<br>◯ A little bit hopeful<br>◯ Not at all hopeful<br>◯ The child is neither hopeful nor not hopeful that COVID-19 will improve/resolve<br>◯ There is no COVID-19 crisis in the child's area<br>◯ Prefer not to answer |

**Compared to before the COVID-19 outbreak, how much is the child now:**

| | Less | Same amount | More | Prefer not to answer |
|---|---|---|---|---|
| Eating | ○ | ○ | ○ | ○ |
| Sleeping | ○ | ○ | ○ | ○ |
| Physically active | ○ | ○ | ○ | ○ |
| Spending time outside | ○ | ○ | ○ | ○ |
| Spending time with friends in-person | ○ | ○ | ○ | ○ |
| Spending time with friends remotely (e.g., online, social media, texting) | ○ | ○ | ○ | ○ |
| Spending time watching TV, playing video/computer games, or using social media for educational purposes, including school work | ○ | ○ | ○ | ○ |
| Spending time watching TV, playing video/computer games, or using social media for NON-educational purposes | ○ | ○ | ○ | ○ |

| DAILY BEHAVIOURS (PAST TWO WEEKS) | |
|---|---|
| During the PAST TWO WEEKS, has the child experienced problems with sleep? (e.g., trouble falling asleep, staying asleep, waking up much earlier than intended, sleeping alone without a parent/caregiver) | ◯ Yes<br>◯ No<br>◯ Prefer not to answer |
| During the PAST TWO WEEKS on average, what time did the child go to bed on WEEKDAYS? | ◯ Before 8 pm<br>◯ 8 pm - 10 pm<br>◯ 10 pm - 12 am (Midnight)<br>◯ After 12 am (Midnight)<br>◯ Prefer not to answer |
| During the PAST TWO WEEKS on average, what time did the child go to bed on WEEKENDS? | ◯ Before 8 pm<br>◯ 8 pm - 10 pm<br>◯ 10 pm - 12 am (Midnight)<br>◯ After 12 am (Midnight)<br>◯ Prefer not to answer |
| During the PAST TWO WEEKS on average, how many hours per night did the child sleep on WEEKDAYS? | ◯ Less than 6 hours<br>◯ 6-8 hours<br>◯ 8-10 hours<br>◯ Greater than 10 hours<br>◯ Prefer not to answer |
| During the PAST TWO WEEKS on average, how many hours per night did the child sleep on WEEKENDS? | ◯ Less than 6 hours<br>◯ 6-8 hours<br>◯ 8-10 hours<br>◯ Greater than 10 hours<br>◯ Prefer not to answer |
| During the PAST TWO WEEKS how many days per week did the child exercise (e.g., increased heart rate, breathing) for at least 30 minutes? | ◯ Not at all<br>◯ 1-2 days per week or less<br>◯ 3-4 days per week<br>◯ 5-6 days per week<br>◯ Daily<br>◯ Prefer not to answer |
| During the PAST TWO WEEKS how many days per week did the child spend time outdoors? | ◯ Not at all<br>◯ 1-2 days per week or less<br>◯ 3-4 days per week<br>◯ 5-6 days per week<br>◯ Daily<br>◯ Prefer not to answer |

#### EMOTIONS/WORRIES (PAST TWO WEEKS)

| | |
|---|---|
| During the PAST TWO WEEKS how happy versus sad was the child? | ◯ Very happy/cheerful<br>◯ Moderately happy/cheerful<br>◯ Neutral<br>◯ Moderately sad/depressed/unhappy<br>◯ Very sad/depressed/unhappy<br>◯ Prefer not to answer |
| During the PAST TWO WEEKS how relaxed versus anxious was the child? | ◯ Very relaxed/calm<br>◯ Moderately relaxed/calm<br>◯ Neutral<br>◯ Moderately nervous/anxious<br>◯ Very nervous/anxious<br>◯ Prefer not to answer |
| During the PAST TWO WEEKS how fidgety or restless was the child? | ◯ Not fidgety/restless at all<br>◯ Slightly fidgety/restless<br>◯ Moderately fidgety/restless<br>◯ Very fidgety/restless<br>◯ Extremely fidgety/restless<br>◯ Prefer not to answer |
| During the PAST TWO WEEKS how tired was the child? | ◯ Not tired at all<br>◯ Slightly tired<br>◯ Moderately tired<br>◯ Very tired<br>◯ Extremely tired<br>◯ Prefer not to answer |
| During the PAST TWO WEEKS how well was the child able to concentrate or focus? | ◯ Very focused/attentive<br>◯ Moderately focused/attentive<br>◯ Neutral<br>◯ Moderately unfocused/distracted<br>◯ Very unfocused/distracted<br>◯ Prefer not to answer |
| During the PAST TWO WEEKS how irritable or easily angered was the child? | ◯ Not irritable or easily angered at all<br>◯ Slightly irritable or easily angered<br>◯ Moderately irritable or easily angered<br>◯ Very irritable or easily angered<br>◯ Extremely irritable or easily angered<br>◯ Prefer not to answer |
| During the PAST TWO WEEKS how lonely was the child? | ◯ Not lonely at all<br>◯ Slightly lonely<br>◯ Moderately lonely<br>◯ Very lonely<br>◯ Extremely lonely<br>◯ Prefer not to answer |

#### MEDIA USE (PAST TWO WEEKS)

| | |
|---|---|
| During the PAST TWO WEEKS, how much time per day did the child spend watching TV or digital media (e.g., Netflix, YouTube, web surfing) for educational purposes, including school work? | ◯ No TV or digital media<br>◯ Under 1 hour<br>◯ 1-3 hours<br>◯ 4-6 hours<br>◯ More than 6 hours<br>◯ Prefer not to answer |
| During the PAST TWO WEEKS, how much time per day did the child spend watching TV or digital media (e.g., Netflix, YouTube, web surfing) for NON-educational purposes? | ◯ No TV or digital media<br>◯ Under 1 hour<br>◯ 1-3 hours<br>◯ 4-6 hours<br>◯ More than 6 hours<br>◯ Prefer not to answer |
| During the PAST TWO WEEKS, how much time per day did the child spend using social media (e.g., Facebook, Instagram, Snapchat, Twitter, TikTok)? | ◯ No social media<br>◯ Under 1 hour<br>◯ 1-3 hours<br>◯ 4-6 hours<br>◯ More than 6 hours<br>◯ Prefer not to answer |
| During the PAST TWO WEEKS, how much time per day did the child spend playing video games? | ◯ No video games<br>◯ Under 1 hour<br>◯ 1-3 hours<br>◯ 4-6 hours<br>◯ More than 6 hours<br>◯ Prefer not to answer<br>(Including time for educational purposes and/or homework) |

**Below are statements about families. You are to decide which of these statements are true of your family and which are false. If you think the statement is True or mostly True of your family, mark True. If you think the statement is False or mostly False of your family, mark False.**

**You may feel that some of the statements are true for some family members and false for others. Mark True if the statement is true for most members. Mark False if the statement is false for most members. If the members are evenly divided, decide what is the stronger overall impression and answer accordingly.**

**Remember, we would like to know what your family seems like to you. So do not try to figure out how other members see your family, but do give us your general impression of your family for each statement.**

**Selected Items from Family Environment Scale (FES) Instrument copyright 1974, 2002 by Rudolf H Moos. All rights reserved in all media. Published by Mind Garden, Inc., www.mindgarden.com**

| | True | False | Prefer not to answer |
|---|---|---|---|
| Family members often criticise each other | ○ | ○ | ○ |
| Family members sometimes get so angry they throw things | ○ | ○ | ○ |
| Family members often try to one-up or out-do each other | ○ | ○ | ○ |
| We fight a lot in our family | ○ | ○ | ○ |
| Family members sometimes hit each other | ○ | ○ | ○ |
| In our family, we believe you don't ever get anywhere by raising your voice | ○ | ○ | ○ |
| If there's a disagreement in our family, we try hard to smooth things over and keep the peace | ○ | ○ | ○ |
| Family members hardly ever lose their tempers | ○ | ○ | ○ |
| Family members rarely become openly angry | ○ | ○ | ○ |

**DEMOGRAPHIC INFORMATION**

**You completed your first survey on [cv19p_date].**

Has the child's living situation changed since you completed the first survey?

- ☐ No, the child's living situation has not changed
- ☐ Yes, the child has have moved (since the first survey)
- ☐ Yes, the child regularly stays in more than one home (since the first survey)
- ☐ Yes, the number of people living in the child's home has changed (since the first survey)
- ☐ Prefer not to answer

Please provide the postal code of the child's primary home

__________________________________
(Enter "555" if you would prefer not to answer.)

How many adults currently live at the child's primary home?

- ◯ 0
- ◯ 1
- ◯ 2
- ◯ 3
- ◯ 4
- ◯ 5
- ◯ 6
- ◯ 7
- ◯ 8
- ◯ 9
- ◯ 10
- ◯ Prefer not to answer

How many children currently live at the child's primary home (excluding the child)?

- ◯ 0
- ◯ 1
- ◯ 2
- ◯ 3
- ◯ 4
- ◯ 5
- ◯ 6
- ◯ 7
- ◯ 8
- ◯ 9
- ◯ 10
- ◯ Prefer not to answer

How many bedrooms are in the child's primary home?

- ◯ 0
- ◯ 1
- ◯ 2
- ◯ 3
- ◯ 4
- ◯ 5
- ◯ 6
- ◯ 7
- ◯ 8
- ◯ 9
- ◯ 10
- ◯ Prefer not to answer

| | |
|---|---|
| How many toilets are in the child's primary home? | ◯ 0<br>◯ 1<br>◯ 2<br>◯ 3<br>◯ 4<br>◯ 5<br>◯ 6<br>◯ 7<br>◯ 8<br>◯ 9<br>◯ 10<br>◯ Prefer not to answer |
| Choose the type of dwelling that best describes the child's primary home | ◯ Manufactured home/ mobile home or trailer<br>◯ Single family detached house<br>◯ Single family house attached to 1 or more houses (e.g townhouse/ semi-detached)<br>◯ High rise apartment/ condo/ co-op (4 or more floors)<br>◯ Low rise apartment/ condo/ co-op (1-3 floors)<br>◯ Multi-family home (more than one family in a converted single home)<br>◯ Other type<br>◯ Prefer not to answer |
| Please provide the postal code of the child's secondary home | ______________________________<br>(Enter "555" if you would prefer not to answer.) |
| How many adults currently live at the child's secondary home? | ◯ 0<br>◯ 1<br>◯ 2<br>◯ 3<br>◯ 4<br>◯ 5<br>◯ 6<br>◯ 7<br>◯ 8<br>◯ 9<br>◯ 10<br>◯ Prefer not to answer |
| How many children currently live at the child's secondary home (excluding the child)? | ◯ 0<br>◯ 1<br>◯ 2<br>◯ 3<br>◯ 4<br>◯ 5<br>◯ 6<br>◯ 7<br>◯ 8<br>◯ 9<br>◯ 10<br>◯ Prefer not to answer |

| | |
|---|---|
| How many bedrooms are in the child's secondary home? | ◯ 0<br>◯ 1<br>◯ 2<br>◯ 3<br>◯ 4<br>◯ 5<br>◯ 6<br>◯ 7<br>◯ 8<br>◯ 9<br>◯ 10<br>◯ Prefer not to answer |
| How many toilets are in the child's secondary home? | ◯ 0<br>◯ 1<br>◯ 2<br>◯ 3<br>◯ 4<br>◯ 5<br>◯ 6<br>◯ 7<br>◯ 8<br>◯ 9<br>◯ 10<br>◯ Prefer not to answer |
| Choose the type of dwelling that best describes the child's secondary home | ◯ Manufactured home/ mobile home or trailer<br>◯ Single family detached house<br>◯ Single family house attached to 1 or more houses (e.g townhouse/ semi-detached)<br>◯ High rise apartment/ condo/ co-op (4 or more floors)<br>◯ Low rise apartment/ condo/ co-op (1-3 floors)<br>◯ Multi-family home (more than one family in a converted single home)<br>◯ Other type<br>◯ Prefer not to answer |
| Are any adults living in the home(s) an ESSENTIAL WORKER? | ☐ Yes, I am<br>☐ Yes, another adult is<br>☐ No<br>☐ Don't Know<br>☐ Prefer not to answer<br>((e.g., healthcare, delivery worker, store worker, security, building maintenance)) |
| If yes, do they come home each day? | ☐ Yes, I do<br>☐ Yes, the other adult does<br>☐ No, separated due to COVID-19<br>☐ No separated due to other reasons<br>☐ Prefer not to answer |
| If yes, are they a FIRST RESPONDER, HEALTHCARE PROVIDER or OTHER WORKER in a facility treating COVID-19? | ☐ Yes, I am<br>☐ Yes, another adult is<br>☐ No<br>☐ Don't Know<br>☐ Prefer not to answer |
| Please share any comments, suggestions or clarifications that you have related to this survey | __________________________________ |

What is your relationship to the child?

◯ Biological Mother
◯ Biological Father
◯ Parent
◯ Step Parent
◯ Foster Parent
◯ Grandparent
◯ Great-Grandparent
◯ Aunt or Uncle
◯ Sibling
◯ Step Sibling
◯ Cousin
◯ Other, Caregiver
◯ Prefer not to answer

Today's Date

__________________________________

### Adult Self Report Follow-up Survey 1

---

ADDENDUM TO PARTICIPANT INFORMATION/CONSENT FORM: Adult Self Report

Before beginning this research study, you signed an Informed Consent Form describing the study and your rights as a study participant. At that time, we explained that we would tell you about any new information that might affect your health, well-being or willingness to stay in the study. The Follow up survey was stated to take 10 minutes to complete. Based on feedback from participants we would like to inform you that the survey may take up to 30 minutes to complete.

By continuing to complete the survey questionnaire you consent to be in this part of the study and that you have read and understood the information provided above.

---

We are interested in learning more about how you are feeling and what you are thinking about during the Coronavirus (COVID-19) pandemic since you completed your first survey on [cv19a_date].

COVID-19 is a new type of illness affecting many people in Canada. The virus that causes COVID-19 is related to the common cold virus. Some people affected by the virus may have symptoms that can be mild or severe, similar to a cold or flu. Other people with COVID-19 may not have any symptoms at all but they can spread the virus to others.

It is important for scientists to learn more about COVID-19. You can help us understand more about how COVID-19 affects families by answering this survey. Do the best you can to answer each question. This survey should take between 20 to 30 minutes to complete.

If you have questions or want help filling the survey out, please contact your site's Coordinator.

---

In general, how would you rate your overall PHYSICAL health now?

◯ Excellent
◯ Very Good
◯ Good
◯ Fair
◯ Poor
◯ Prefer not to answer

---

In general, how would you rate your overall MENTAL/EMOTIONAL health now?

◯ Excellent
◯ Very Good
◯ Good
◯ Fair
◯ Poor
◯ Prefer not to answer

**COVID-19 HEALTH/EXPOSURE STATUS**

**You completed your first survey on [cv19a_date].**

In what ways has the COVID-19 outbreak affected your overall healthcare, since you completed the first survey? (Select all that apply)

- ☐ I did not go to healthcare appointments because I was concerned about entering the healthcare providers office
- ☐ My healthcare provider cancelled appointments My
- ☐ healthcare provider changed to phone or online visits
- ☐ My healthcare provider told me to self-isolate or quarantine
- ☐ None of these apply
- ☐ Prefer not to answer

What kind of healthcare visits were affected since you completed the first survey? (Select all that apply)

- ☐ Routine visit/ regular checkup
- ☐ Vaccination visit
- ☐ Visit with a health care professional for a specific concern
- ☐ Visit with a specialist
- ☐ I did not miss any healthcare visits
- ☐ Prefer not to answer

Has the COVID-19 crisis in your area led to any family members in your household(s) applying for any of the following government supports, since you completed the first survey? (Select all that apply)

- ☐ Residential Mortgage or Lease/ Rent Payment Deferral
- ☐ Commercial Mortgage or Lease/ Rent Payment Deferral
- ☐ Personal Income Support (e.g Canadian Emergency Response Benefit (CERB), Canadian Emergency Student Benefit (CESB), Employment Insurance (EI))
- ☐ Business Income Support (e.g Canadian Emergency Wage Subsidy (CEWS))
- ☐ Food Bank
- ☐ Other, Specify
- ☐ None of the above
- ☐ Don't know
- ☐ Prefer not to answer

Specify other supports

______________________________
(Enter "555" if you would prefer not to answer.)

Have you been exposed to someone likely to have COVID-19, since you completed the first survey? (Select all that apply)

- ☐ Yes, to someone with positive test
- ☐ Yes, to someone with medical diagnosis, but no test
- ☐ Yes, to someone with possible symptoms, but no diagnosis by doctor
- ☐ No, not to my knowledge
- ☐ Prefer not to answer

Have you been suspected of having COVID-19, since you completed the first survey?

- ☐ Yes, had positive test
- ☐ Yes, had medical diagnosis, but no test
- ☐ Yes, had some possible symptoms, but no diagnosis by health care professional
- ☐ No
- ☐ Prefer not to answer

| | |
|---|---|
| Have you been tested for the virus that causes COVID-19, since you completed the first survey? (Select all that apply)<br><br>Do not include the antibody/ serology test (blood test for previous infection) that is part of this reseach study. | ☐ No, I never tried to get tested<br>☐ No, I tried to get tested but was not able to<br>☐ Yes, and I am waiting for the results<br>☐ Yes, and the test showed that I did not have it ("negative" test)<br>☐ Yes, and the test showed that I did have it ("positive" test)<br>☐ Prefer not to answer |
| Which type of test for the virus that causes COVID-19 did you try to get but were not able to? | ☐ Viral Test (swab test for current infection)<br>☐ Antibody/ Serology Test (blood test for previous infection)<br>☐ Saliva or Mouth Rinse Test<br>☐ Rapid-Antigen Test<br>☐ Other Specify<br>☐ Don't Know<br>☐ Prefer not to answer<br>(Antibody/ Serology testing and saliva/ mouth rinse testing may not be currently available in all areas, but it may be available in the future.) |
| Specify the other type of test for the virus that causes COVID-19 that you tried to get but were not able to. | ______________________________<br>(Enter "555" if you would prefer not to answer.) |
| Please specify why you got tested or tried to get tested | ☐ A contact or possible contact of mine tested positive for COVID-19<br>☐ I had COVID-19 symptoms<br>☐ I was admitted to hospital (unrelated to COVID-19)<br>☐ I was admitted to hospital (COVID-19 related)<br>☐ I was tested in another facility or institution context (contact with someone in long term care, prison etc)<br>☐ I was participating in another facility research study<br>☐ Travel-related testing<br>☐ Routine workplace testing (health care workplace)<br>☐ Routine workplace testing (non-health care workplace)<br>☐ Other, specify<br>☐ Don't Know<br>☐ Prefer not to answer |
| Please specify the other reason(s) why you got tested or tried to get tested | ______________________________<br>(Enter "555" if you would prefer not to answer.) |
| Please specify why you didn't try to get tested | ☐ Symptoms were due to another condition, specify<br>☐ Symptoms were mild<br>☐ A healthcare provider did not recommend testing<br>☐ Other, specify<br>☐ Prefer not to answer |
| Please specify the condition that you believe the symptoms are from | ______________________________<br>(Enter "555" if you would prefer not to answer.) |
| Please specify the other reason(s) you didn't try to get tested | ______________________________<br>(Enter "555" if you would prefer not to answer.) |

| | |
|---|---|
| IN THE PAST TWO WEEKS how many times have you been tested for COVID-19? | ◯ 0<br>◯ 1<br>◯ 2<br>◯ 3<br>◯ 4<br>◯ Prefer not to answer |
| Details of First COVID-19 Test | |
| What type of test did you receive? | ◯ Viral Test (swab test for current infection)<br>◯ Antibody/ Serology Test (blood test for previous infection)<br>◯ Saliva or Mouth Rinse Test<br>◯ Rapid-Antigen Test<br>◯ Other Specify<br>◯ Don't Know<br>◯ Prefer not to answer<br>(Do not include the antibody/ serology test (blood test for previous infection) that is part of this reseach study ) |
| Specify other type of test you received. | ______________________________<br>(Enter "555" if you would prefer not to answer.) |
| Date of test | ______________________________<br>(If you do not recall the exact date, please provide an estimate. Enter "01-01-1900" if you would prefer not to answer.) |
| First test result | ◯ Negative<br>◯ Positive<br>◯ Don't know<br>◯ Prefer not to answer |
| Details of Second COVID-19 Test | |
| What type of test did you receive? | ◯ Viral Test (swab test for current infection)<br>◯ Antibody/ Serology Test (blood test for previous infection)<br>◯ Saliva or Mouth Rinse Test<br>◯ Rapid-Antigen Test<br>◯ Other Specify<br>◯ Don't Know<br>◯ Prefer not to answer<br>(Do not include the antibody/ serology test (blood test for previous infection) that is part of this reseach study ) |
| Specify other type of test you received. | ______________________________<br>(Enter "555" if you would prefer not to answer.) |
| Date of test | ______________________________<br>(If you do not recall the exact date, please provide an estimate. Enter "01-01-1900" if you would prefer not to answer.) |

| | |
|---|---|
| Second test result | ◯ Negative<br>◯ Positive<br>◯ Don't know<br>◯ Prefer not to answer |
| Details of Third COVID-19 Test | |
| What type of test did you receive? | ◯ Viral Test (swab test for current infection)<br>◯ Antibody/ Serology Test (blood test for previous infection)<br>◯ Saliva or Mouth Rinse Test<br>◯ Rapid-Antigen Test<br>◯ Other Specify<br>◯ Don't Know<br>◯ Prefer not to answer<br>(Do not include the antibody/ serology test (blood test for previous infection) that is part of this reseach study ) |
| Specify other type of test you received. | __________________________________<br>(Enter "555" if you would prefer not to answer.) |
| Date of test | __________________________________<br>(If you do not recall the exact date, please provide an estimate. Enter "01-01-1900" if you would prefer not to answer.) |
| Third test result | ◯ Negative<br>◯ Positive<br>◯ Don't know<br>◯ Prefer not to answer |
| Details of Fourth COVID-19 Test | |
| What type of test did you receive? | ◯ Viral Test (swab test for current infection)<br>◯ Antibody/ Serology Test (blood test for previous infection)<br>◯ Saliva or Mouth Rinse Test<br>◯ Rapid-Antigen Test<br>◯ Other Specify<br>◯ Don't Know<br>◯ Prefer not to answer<br>(Do not include the antibody/ serology test (blood test for previous infection) that is part of this reseach study ) |
| Specify other type of test you received. | __________________________________<br>(Enter "555" if you would prefer not to answer.) |
| Date of test | __________________________________<br>(If you do not recall the exact date, please provide an estimate. Enter "01-01-1900" if you would prefer not to answer.) |

| | |
|---|---|
| Fourth test result | ◯ Negative<br>◯ Positive<br>◯ Don't know<br>◯ Prefer not to answer |
| Have you had any colds, since you completed the first survey? | ◯ Yes<br>◯ No<br>◯ Don't Know<br>◯ Prefer not to answer |
| Have you had any of the following symptoms, since you completed the first survey? (Select all that apply) | ☐ New or worsening cough<br>☐ Shortness of breath<br>☐ Difficulty breathing<br>☐ Fever<br>☐ Chills<br>☐ Muscle or body aches<br>☐ Headache<br>☐ Sore throat<br>☐ New loss of sense of smell or taste<br>☐ Runny nose or nasal congestion*<br>☐ Nausea or vomiting<br>☐ Diarrhea<br>☐ Fatigue<br>☐ Skin Rash<br>☐ Conjunctivitis (pink eye)<br>☐ Other, Specify<br>☐ None of the above<br>☐ Prefer not to answer<br>(*Runny nose or nasal congestion in the absence of underlying reason for these symptoms such as seasonal allergies, post nasal drip, etc) |
| Specify the other symptoms that you had | __________________________________<br>(Enter "555" if you would prefer not to answer.) |
| Do you remember the date when you had your first symptom? | ◯ Yes<br>◯ No<br>◯ Roughly<br>◯ Prefer not to answer |
| What was the date of your first symptom? | __________________________________<br>(If you do not recall the exact date, please provide an estimate. Enter "01-01-1900" if you would prefer not to answer.) |
| In the TWO WEEKS before you had symptoms, did you:(Select all that apply) | ☐ Have contact with someone who tested positive for COVID-19<br>☐ Have contact with someone who likely had COVID-19 (e.g., was not tested but had symptoms; was told by a healthcare provider that he/she likely had it)<br>☐ Travel outside the province<br>☐ None of the above<br>☐ Prefer not to answer |

| | |
|---|---|
| How many places did you travel? | ◯ 0<br>◯ 1<br>◯ 2<br>◯ 3<br>◯ 4<br>◯ Prefer not to answer<br>(Travel outside the province) |
| Place 1 | |
| Location<br>(e.g. Seattle, Washington OR Toronto, Ontario) | ______________________________<br>(Enter "555" if you would prefer not to answer.) |
| Start date of travel | ______________________________<br>(If you do not recall the exact date, please provide an estimate. Enter "01-01-1900" if you would prefer not to answer.) |
| End date of travel | ______________________________<br>(If you do not recall the exact date, please provide an estimate. Enter "01-01-1900" if you would prefer not to answer.) |
| How? (Check all that apply) | ☐ Car<br>☐ Plane<br>☐ Train<br>☐ Ship<br>☐ Other<br>☐ Prefer not to answer |
| Please specify other mode of transportation | ______________________________<br>(Enter "555" if you would prefer not to answer.) |
| Place 2 | |
| Location<br>(e.g. Portland, Oregon OR Vancouver, BC) | ______________________________<br>(Enter "555" if you would prefer not to answer.) |
| Start date of travel | ______________________________<br>(If you do not recall the exact date, please provide an estimate. Enter "01-01-1900" if you would prefer not to answer.) |
| End date of travel | ______________________________<br>(If you do not recall the exact date, please provide an estimate. Enter "01-01-1900" if you would prefer not to answer.) |

| | |
|---|---|
| How? (Check all that apply) | ☐ Car<br>☐ Plane<br>☐ Train<br>☐ Ship<br>☐ Other<br>☐ Prefer not to answer |
| Please specify other mode of transportation | ______________________<br>(Enter "555" if you would prefer not to answer.) |
| Place 3 | |
| Location<br>(e.g. Paris, France OR Banff, Alberta) | ______________________<br>(Enter "555" if you would prefer not to answer.) |
| Start date of travel | ______________________<br>(If you do not recall the exact date, please provide an estimate. Enter "01-01-1900" if you would prefer not to answer.) |
| End date of travel | ______________________<br>(If you do not recall the exact date, please provide an estimate. Enter "01-01-1900" if you would prefer not to answer.) |
| How? (Check all that apply) | ☐ Car<br>☐ Plane<br>☐ Train<br>☐ Ship<br>☐ Other<br>☐ Prefer not to answer |
| Please specify other mode of transportation | ______________________<br>(Enter "555" if you would prefer not to answer.) |
| Place 4 | |
| Location<br>(e.g. Portland, Oregon OR Vancouver, BC) | ______________________<br>(Enter "555" if you would prefer not to answer.) |
| Start date of travel | ______________________<br>(If you do not recall the exact date, please provide an estimate. Enter "01-01-1900" if you would prefer not to answer.) |
| End date of travel | ______________________<br>(If you do not recall the exact date, please provide an estimate. Enter "01-01-1900" if you would prefer not to answer.) |

| | |
|---|---|
| How? (Check all that apply) | ☐ Car<br>☐ Plane<br>☐ Train<br>☐ Ship<br>☐ Other<br>☐ Prefer not to answer |
| Please specify other mode of transportation | ________________________<br>(Enter "555" if you would prefer not to answer.) |
| Which of the following occurred as a result of your symptoms? (Select all that apply) | ☐ You spoke to a healthcare provider over the phone, by email, or online<br>☐ You saw a healthcare provider in person, such as in a clinic or doctor's office<br>☐ You saw a healthcare provider in person, such as in urgent care, or in an Emergency Department (ED)<br>☐ You were kept overnight in a hospital because a healthcare provider thought you had COVID-19 You<br>☐ were admitted to a hospital's ICU (Intensive Care Unit)<br>☐ You self-isolated or quarantined at home<br>☐ None of the above<br>☐ Prefer not to answer |
| Has anyone in your family been diagnosed with COVID-19, since you completed the first survey? (Select all that apply) | ☐ Yes, member of household<br>☐ Yes, non-household<br>☐ member No<br>☐ Don't know<br>☐ Prefer not to answer |
| Have any of the following happened to your family members because of COVID-19, since you completed the first survey? (Select all that apply) | ☐ Fallen physically ill<br>☐ Hospitalized<br>☐ Self-isolated or quarantined with symptoms<br>☐ Self-isolated or quarantined without symptoms (e.g., due to possible exposure)<br>☐ Lost job or been laid off from job<br>☐ Reduced ability to earn money<br>☐ Passed away<br>☐ None of the above<br>☐ Prefer not to answer<br>(Include any family members, not just those living in the same household as you) |
| Did you get the flu shot, since you completed the first survey? | ◯ Yes<br>◯ No<br>◯ Don't Know<br>◯ Prefer not to answer |
| Have you received at least one dose of a COVID-19 vaccine? | ◯ Yes<br>◯ No<br>◯ Prefer not to answer<br>(Answer 'Yes' if you have received at least one dose of the COVID-19 vaccine. Note: Certain types of vaccines require more than one dose to protect against COVID-19. You would have been informed at the time of vaccination if you needed a second dose.) |

| | |
|---|---|
| Did you feel any side effects from the COVID-19 vaccine? | ◯ Yes<br>◯ No<br>◯ Prefer not to answer |
| If yes, check all that apply. | ☐ Head ache<br>☐ Tiredness<br>☐ Fever or chills<br>☐ Muscle or joint pain<br>☐ Soreness where the vaccine was injected<br>☐ Nausea or vomiting<br>☐ Other<br>☐ Prefer not to answer |
| Please specify other side effects | __________________________________<br>(Enter "555" if you would prefer not to answer.) |
| How many doses of the COVID-19 vaccine have you received so far? | ◯ One dose<br>◯ Two doses<br>◯ More than two doses<br>◯ Prefer not to answer |
| When did you receive your first dose of the COVID-19 vaccine? | __________________________________<br>(If you do not recall the exact date, please provide an estimate. Enter "01-01-1900" if you would prefer not to answer.) |
| Which vaccine did you receive? (first dose) | ◯ Pfizer and BioNTech mRNA vaccine<br>◯ Moderna mRNA vaccine<br>◯ AstraZeneca Oxford vaccine Janssen<br>◯ (Johnson & Johnson) vaccine Other,<br>◯ specify<br>◯ Don't Know<br>◯ Prefer not to answer |
| Specify the other vaccine | __________________________________<br>(Enter "555" if you would prefer not to answer.) |
| When did you receive your second dose of the COVID-19 vaccine? | __________________________________<br>(If you do not recall the exact date, please provide an estimate. Enter "01-01-1900" if you would prefer not to answer.) |
| Which vaccine did you receive? (second dose) | ◯ Pfizer and BioNTech mRNA vaccine<br>◯ Moderna mRNA vaccine<br>◯ AstraZeneca Oxford vaccine Janssen<br>◯ (Johnson & Johnson) vaccine Other,<br>◯ specify<br>◯ Don't Know<br>◯ Prefer not to answer |
| Specify the other vaccine | __________________________________<br>(Enter "555" if you would prefer not to answer.) |

| | |
|---|---|
| When a COVID-19 vaccine becomes available, how likely is it that you will choose to get it? | ◯ Very Likely<br>◯ Somewhat Likely<br>◯ Somewhat Unlikely<br>◯ Very Unlikely<br>◯ I Don't Know<br>◯ Prefer not to answer |
| What are some of the reasons why you don't want to get the COVID-19 vaccine? | ☐ Not confident in the safety of the vaccine<br>☐ Concern about risks and side effects<br>☐ Will wait until it seems safe to get the vaccine<br>☐ Do not consider it necessary to get the vaccine<br>☐ Do not believe in vaccination<br>☐ Have not yet decided<br>☐ Have a pre-existing medical condition Already<br>☐ had or think I have had COVID-19 Other,<br>☐ Specify<br>☐ Prefer not to answer<br>(Check all that apply.) |
| What were the other reasons you didn't want to get the COVID-19 vaccine? | __________________________________<br>(Enter "555" if you would prefer not to answer.) |

| | Strongly Agree | Agree | Disagree | Strongly Disagree | Prefer not to answer |
|---|---|---|---|---|---|
| Having myself vaccinated is important for the health of others in my community. | ◯ | ◯ | ◯ | ◯ | ◯ |
| Getting vaccines is a good way to protect myself from disease. | ◯ | ◯ | ◯ | ◯ | ◯ |
| Generally I do what my doctor or health care provider recommends about vaccines for myself | ◯ | ◯ | ◯ | ◯ | ◯ |
| I am concerned about serious adverse effects of vaccines. | ◯ | ◯ | ◯ | ◯ | ◯ |

| | Never | Rarely | Occasionally | Often | Always | Don't Know | Prefer not to answer |
|---|---|---|---|---|---|---|---|
| How often have you worn a mask in public places since you completed your first survey? | ◯ | ◯ | ◯ | ◯ | ◯ | ◯ | ◯ |
| How often have you practiced physical distancing in public places since you completed your first survey? | ◯ | ◯ | ◯ | ◯ | ◯ | ◯ | ◯ |
| How often have you avoided crowded places/ gatherings since you completed your first survey? | ◯ | ◯ | ◯ | ◯ | ◯ | ◯ | ◯ |

| | | | | | | | |
|---|---|---|---|---|---|---|---|
| How often have you avoided common greetings (e.g handshakes, hugs etc) since you completed your first survey? | ◯ | ◯ | ◯ | ◯ | ◯ | ◯ | ◯ |
| How often have you limited contact with people at higher risk (e.g. elderly relative) since you completed your first survey? | ◯ | ◯ | ◯ | ◯ | ◯ | ◯ | ◯ |
| How often have you self-Isolated because someone thought you were infected with the virus that causes COVID-19 since you completed your first survey? | ◯ | ◯ | ◯ | ◯ | ◯ | ◯ | ◯ |
| How often have you quarantined because you may have been exposed to the virus that causes COVID-19, but did not show symptoms since you completed your first survey? | ◯ | ◯ | ◯ | ◯ | ◯ | ◯ | ◯ |

---

Have you participated in any organized sports or recreational activities, in person, since you completed the first survey?

◯ Yes
◯ No
◯ Prefer not to answer

---

If yes, how many people were in the activity?

◯ 0-9
◯ 10-19
◯ 20-29
◯ 30+
◯ Prefer not to answer

---

How many times have you been in a gathering of >10 or more people (excluding school or work), since you completed the first survey?

__________________________________
(A gathering in this context is defined as any assembly or meeting of more than 10 persons, in open air or in a building or premises. Gatherings include a range of interpersonal gatherings/events of varying purposes, from gatherings/events of family and friends (e.g., weddings, funerals, baptisms, birthday parties and cultural ceremonies) to community gathering spaces (e.g., places of worship, places of traditional gatherings, libraries, community and recreation centres, drop-in centres, food banks and communal kitchens, museums, theatres, cinemas, and tourist attractions) to larger planned or structured gatherings/events, including mass gatherings (e.g., large meetings or conferences, national sporting events, large cultural/religious events, festivals). Enter "555" if you would prefer not to answer.)

**The following are a number of mental health services that you may have accessed. Please check the relevant boxes for each service indicating if you have accessed the service since March 2020 or if you would be interested in accessing the service.**

| | Have accessed this service since March 2020 | Would be interested in accessing this service | Not accessed, not interested | Prefer not to answer |
|---|---|---|---|---|
| Individual therapy (you alone) with a professional (in person or virtual) | ○ | ○ | ○ | ○ |
| Group therapy (you and others) with a professional (in person or virtual) | ○ | ○ | ○ | ○ |
| Couples therapy (you and your partner) with a professional (in person or virtual) | ○ | ○ | ○ | ○ |
| Family therapy (you and your family members) with a professional (in person or virtual) | ○ | ○ | ○ | ○ |
| Mental health mobile app (general) | ○ | ○ | ○ | ○ |
| Mental health mobile app to interact with a therapist | ○ | ○ | ○ | ○ |
| Phone helpline for mental health support (e.g. Crisis Services Canada) | ○ | ○ | ○ | ○ |
| Online mental health information you looked up yourself | ○ | ○ | ○ | ○ |

**Social Support**

**We would like to measure your feelings about how supported you feel. Choose 1 of the 4 responses to show whether you agree or disagree with the statement.**

| | Strongly Agree | Agree | Disagree | Strongly Disagree | Prefer not to answer |
|---|---|---|---|---|---|
| I have family and friends who help me feel safe, secure and happy | ○ | ○ | ○ | ○ | ○ |
| There is someone I trust whom I would turn to for advice if I were having problems | ○ | ○ | ○ | ○ | ○ |
| There are people I can count on in an emergency | ○ | ○ | ○ | ○ | ○ |
| I feel part of a group of people who share my attitudes and beliefs | ○ | ○ | ○ | ○ | ○ |

**Worrying (PAST TWO WEEKS)**

**You completed your first survey on [cv19a_date].**

| | Not at all worried | Slightly worried | Moderately worried | Very worried | Extremely worried | Prefer not to answer |
|---|---|---|---|---|---|---|
| During the PAST TWO WEEKS how worried have you been about being infected by the virus that causes COVID-19? | ◯ | ◯ | ◯ | ◯ | ◯ | ◯ |
| During the PAST TWO WEEKS how worried have you been about friends or family being infected by the virus that causes COVID-19? | ◯ | ◯ | ◯ | ◯ | ◯ | ◯ |
| During the PAST TWO WEEKS how worried have you been about your physical health being influenced by COVID-19? | ◯ | ◯ | ◯ | ◯ | ◯ | ◯ |
| During the PAST TWO WEEKS how worried have you been about your mental/emotional health being influenced by COVID-19? | ◯ | ◯ | ◯ | ◯ | ◯ | ◯ |
| During the PAST TWO WEEKS how worried were you generally? | ◯ | ◯ | ◯ | ◯ | ◯ | ◯ |

How much are you reading or talking about COVID-19, since you completed the first survey?

◯ Never
◯ A few times a month
◯ Weekly
◯ A few times a week
◯ Daily
◯ Prefer not to answer

Has the COVID-19 crisis in your area led to any POSITIVE changes in your life, since you completed the first survey?

◯ Yes
◯ No
◯ Prefer not to answer

Please specify these POSITIVE changes in your life (Select all that apply)

- ☐ More time with family
- ☐ More time with friends
- ☐ More exercising
- ☐ Healthier eating
- ☐ New hobbies
- ☐ Got a new pet
- ☐ Learned new technology
- ☐ Made new friends online
- ☐ Saved money
- ☐ Less commuting
- ☐ Better hand hygiene
- ☐ Less busy / more downtime
- ☐ Working from home
- ☐ Schooling from home
- ☐ More time outdoors
- ☐ Not getting sick as often
- ☐ Avoiding bullying
- ☐ Home improvements
- ☐ Improved mental health
- ☐ Other, specify
- ☐ Prefer not to answer

---

Please specify these other POSITIVE changes in your life

________________________________________
(Enter "555" if you would prefer not to answer.)

**Families have different routines.**

**We would like to know about your family's routines. Choose 1 of the 4 responses to show whether you agree or disagree with the statement.**

| | Strongly Agree | Agree | Disagree | Strongly Disagree | Prefer not to answer |
|---|---|---|---|---|---|
| Parents have some times each day for just talking with children | ◯ | ◯ | ◯ | ◯ | ◯ |
| Parents and children play together each day | ◯ | ◯ | ◯ | ◯ | ◯ |
| The whole family eats dinner together almost every night | ◯ | ◯ | ◯ | ◯ | ◯ |
| Family checks in or out with each other when someone leaves the home | ◯ | ◯ | ◯ | ◯ | ◯ |
| Children do regular household chores | ◯ | ◯ | ◯ | ◯ | ◯ |
| Parents read with children together almost every day | ◯ | ◯ | ◯ | ◯ | ◯ |
| Children go to bed at the same time each night during the week | ◯ | ◯ | ◯ | ◯ | ◯ |

**LIFE CHANGES DUE TO THE COVID-19 CRISIS**
**You completed your first survey on [cv19a_date].**

| | |
|---|---|
| Are you enrolled in school for the current academic year*? | ◯ Not in school<br>◯ Home school<br>◯ Elementary school<br>◯ Junior High or Middle<br>◯ School Secondary or High<br>◯ School College<br>◯ University<br>◯ Prefer not to answer<br>(Current academic year = Sept 2020 to June 2021) |
| Has your school building closed since you completed your first survey? | ◯ Yes<br>◯ No<br>◯ N/A<br>◯ Prefer not to answer |
| If no, are you attending classes in person? | ◯ Yes<br>◯ No<br>◯ N/A<br>◯ Prefer not to answer |
| If yes, how many times did the school close? | ________________________<br>(Enter "666" if you don't know the answer.Enter "555" if you would prefer not to answer.) |
| If yes, when did the first school building closure begin? | ________________________<br>(If you do not recall the exact date, please provide an estimate. Enter "01-01-1900" if you would prefer not to answer.) |
| How many weeks in total was the school building closed since September 2020 due to COVID-19? | ________________________<br>((do not include Christmas and spring break) Enter "555" if you would prefer not to answer.) |
| If yes, were classes resumed online? | ◯ Yes<br>◯ No<br>◯ N/A<br>◯ Prefer not to answer |
| Did you participate in classes online since September 2020? | ◯ Yes<br>◯ No<br>◯ Partially<br>◯ N/A<br>◯ Prefer not to answer |
| If yes, did you have easy access to the internet and a computer? | ◯ Yes<br>◯ No<br>◯ N/A<br>◯ Prefer not to answer |
| If yes, were there assignments for you to complete? | ◯ Yes<br>◯ No<br>◯ N/A<br>◯ Prefer not to answer |

| | |
|---|---|
| Did you complete the assignments? | ◯ Yes<br>◯ No<br>◯ Partially<br>◯ N/A<br>◯ Prefer not to answer |
| Did you physically return to school since the first closure? | ◯ Yes<br>◯ No<br>◯ N/A<br>◯ Prefer not to answer |
| When did you return to school? | ____________________<br>(If you do not recall the exact date, please provide an estimate. Enter "01-01-1900" if you would prefer not to answer.) |
| How many days of in-school or remote/online learning have you missed/ been absent for since you completed your first survey? | ____________________<br>(Enter "555" if you would prefer not to answer.) |
| Are you currently working? | ☐ Working for pay<br>☐ On paid leave<br>☐ On unpaid leave<br>☐ Laid off or lost job<br>☐ Unemployed and looking for a<br>☐ job Retired<br>☐ Staying at home/ homemaker<br>☐ Other<br>☐ Prefer not to answer |
| In what ways has the COVID-19 outbreak affected your work, since you completed the first survey? (Select all that apply) | ☐ I moved to working remotely or from home<br>☐ I lost my job permanently<br>☐ I lost my job temporarily, or was not told for how long<br>☐ I got a new job<br>☐ I reduced my work hours<br>☐ I increased my work hours<br>☐ My job put me at increased risk of getting COVID-19<br>☐ I laid off employees<br>☐ I did not have a paying job before the COVID-19 outbreak<br>☐ None of these apply<br>☐ Prefer not to answer |

| | |
|---|---|
| In what ways has the COVID-19 outbreak affected your spouse/partners work, since you completed the first survey? (Select all that apply) | ☐ Not applicable I do not have a spouse/partner My<br>☐ spouse/partner moved to working remotely or from home<br>☐ My spouse/partner lost his/her job permanently My<br>☐ spouse/partner lost his/her job temporarily, or was not told for how long<br>☐ My spouse/partner got a new job<br>☐ My spouse/partner reduced his/her work hours My<br>☐ spouse/partner increased his/her work hours My<br>☐ spouse/partners job put him/her at increased risk of getting COVID-19<br>☐ My spouse/partner laid off employees<br>☐ My spouse/partner did not have a paying job before the COVID-19 outbreak<br>☐ None of these apply<br>☐ Prefer not to answer |
| During the PAST TWO WEEKS, how many people (from outside of your household) have you had an in-person conversation with in an indoor environment? | __________________________________<br>(In-person means that the person is in the same place as you (NOT on the phone or video chat) Enter "555" if you would prefer not to answer.) |
| During the PAST TWO WEEKS, how many people (from outside of your household) have you had an in-person conversation with in an outdoor environment? | __________________________________<br>(In-person means that the person is in the same place as you (NOT on the phone or video chat) Enter "555" if you would prefer not to answer.) |
| During the PAST TWO WEEKS, how many times did you go out outside of your home (e.g., going to stores, parks, etc.)? | ◯ Not at all<br>◯ 1-2 days per week or less<br>◯ 3-4 days per week<br>◯ 5-6 days per week<br>◯ Daily<br>◯ Prefer not to answer |
| During the PAST TWO WEEKS, how stressful have the restrictions on leaving home been for you? | ◯ Not stressful<br>◯ A little bit stressful<br>◯ Stressful<br>◯ Very stressful<br>◯ Extremely stressful<br>◯ There are no restrictions on leaving home<br>◯ Prefer not to answer |
| During the PAST TWO WEEKS, how difficult has it been for you to follow the recommendations for keeping away from close contact with people who you don't live with? | ◯ Not difficult<br>◯ A little bit difficult<br>◯ Difficult<br>◯ Very difficult<br>◯ Extremely difficult<br>◯ There are no recommendations for keeping away from close contact with people who I don't live with<br>◯ Prefer not to answer |
| During the PAST TWO WEEKS, has the quality of the relationships between you and members of your family changed? | ◯ A lot better<br>◯ A little better About<br>◯ the same<br>◯ A little worse<br>◯ A lot worse<br>◯ Prefer not to answer |

| | |
|---|---|
| During the PAST TWO WEEKS, how stressful have these changes in relationships between you and members of your family been for you? | ◯ Not stressful<br>◯ A little bit stressful<br>◯ Stressful<br>◯ Very stressful<br>◯ Extremely stressful<br>◯ Prefer not to answer |
| During the PAST TWO WEEKS, has the quality of your relationships with your friends changed? | ◯ A lot better<br>◯ A little better About<br>◯ the same<br>◯ A little worse<br>◯ A lot worse<br>◯ Prefer not to answer |
| During the PAST TWO WEEKS, how stressful have these changes in relationships between you and your friends been for you? | ◯ Not stressful<br>◯ A little bit stressful<br>◯ Stressful<br>◯ Very stressful<br>◯ Extremely stressful<br>◯ Prefer not to answer |
| During the PAST TWO WEEKS how difficult has the cancellation of important events in your life (such as weddings, vacations, etc.) been for you? | ◯ Not difficult<br>◯ A little bit difficult<br>◯ Difficult<br>◯ Very difficult<br>◯ Extremely difficult<br>◯ There hasn't been any cancellation of important events<br>◯ Prefer not to answer |
| During the PAST TWO WEEKS, to what degree have changes related to the COVID-19 crisis in your area created financial problems for your family? | ◯ No financial problems<br>◯ A few financial problems<br>◯ Some financial problems A<br>◯ lot of financial problems<br>◯ Extreme financial<br>◯ problems Prefer not to answer |
| During the PAST TWO WEEKS, to what degree are you concerned about the stability of your living situation? | ◯ Not at all concerned<br>◯ A little concerned<br>◯ Concerned<br>◯ Very concerned<br>◯ Extremely concerned<br>◯ Prefer not to answer<br>(Stability of your living situation refers to forced moves that are the result of poor housing quality, unstable neighbourhood conditions and high costs of housing in relation to income. ) |
| During the PAST TWO WEEKS, did you worry your food would run out because of a lack of money? | ◯ Yes<br>◯ No<br>◯ Prefer not to answer |
| How hopeful are you that the COVID-19 crisis in your area will end soon? | ◯ Extremely hopeful<br>◯ Very hopeful<br>◯ Hopeful<br>◯ A little bit hopeful<br>◯ Not at all hopeful<br>◯ There is no COVID-19 crisis in my area<br>◯ Prefer not to answer |

**Compared to before the COVID-19 outbreak, how much are you now:**

| | Less | Same amount | More | Prefer not to answer |
|---|---|---|---|---|
| Eating | ○ | ○ | ○ | ○ |
| Sleeping | ○ | ○ | ○ | ○ |
| Physically active | ○ | ○ | ○ | ○ |
| Spending time outside | ○ | ○ | ○ | ○ |
| Spending time with friends in-person | ○ | ○ | ○ | ○ |
| Spending time with friends remotely (e.g., online, social media, texting) | ○ | ○ | ○ | ○ |
| Spending time watching TV, playing video/computer games, or using social media for educational or work purposes, including school work | ○ | ○ | ○ | ○ |
| Spending time watching TV, playing video/computer games, or using social media for NON-educational or NON-work purposes | ○ | ○ | ○ | ○ |

| **DAILY BEHAVIORS (Past TWO WEEKS)**<br><br>**You completed your first survey on [cv19a_date].** | |
|---|---|
| During the PAST TWO WEEKS on average, what time did you go to bed on WEEKDAYS? | ◯ Before 8 pm<br>◯ 8 pm - 10 pm<br>◯ 10 pm - 12 am (Midnight)<br>◯ After 12 am (Midnight)<br>◯ Prefer not to answer |
| During the PAST TWO WEEKS on average, what time did you go to bed on WEEKENDS? | ◯ Before 8 pm<br>◯ 8 pm - 10 pm<br>◯ 10 pm - 12 am (Midnight)<br>◯ After 12 am (Midnight)<br>◯ Prefer not to answer |
| During the PAST TWO WEEKS on average, how many hours per night did you sleep on WEEKDAYS? | ◯ Less than 6 hours<br>◯ 6-8 hours<br>◯ 8-10 hours<br>◯ Greater than 10 hours<br>◯ Prefer not to answer |
| During the PAST TWO WEEKS on average, how many hours per night did you sleep on WEEKENDS? | ◯ Less than 6 hours<br>◯ 6-8 hours<br>◯ 8-10 hours<br>◯ Greater than 10 hours<br>◯ Prefer not to answer |
| During the PAST TWO WEEKS how many days per week did you exercise (e.g., increased heart rate, breathing) for at least 30 minutes? | ◯ Not at all<br>◯ 1-2 days per week or less<br>◯ 3-4 days per week<br>◯ 5-6 days per week<br>◯ Daily<br>◯ Prefer not to answer |
| During the PAST TWO WEEKS how many days per week did you spend time outdoors? | ◯ Not at all<br>◯ 1-2 days per week or less<br>◯ 3-4 days per week<br>◯ 5-6 days per week<br>◯ Daily<br>◯ Prefer not to answer |
| How has the COVID-19 outbreak affected your regular childcare, since you completed the first survey?<br>(Select all that apply) | ☐ I had difficulty arranging for childcare<br>☐ I had to pay more for childcare<br>☐ My spouse/partner or I had to change our work schedule to care for our children ourselves<br>☐ My spouse/partner or I had to reduce our paid work hours to care for our children ourselves<br>☐ My regular childcare has not been affected by the COVID-19 outbreak<br>☐ I do not have a child in childcare<br>☐ Prefer not to answer |

**EMOTIONS/WORRIES (PAST TWO WEEKS)**

**You completed your first survey on [cv19a_date].**

| | |
|---|---|
| During the PAST TWO WEEKS how happy versus sad were you? | ◯ Very happy/cheerful<br>◯ Moderately happy/cheerful<br>◯ Neutral<br>◯ Moderately sad/depressed/unhappy<br>◯ Very sad/depressed/unhappy<br>◯ Prefer not to answer |
| During the PAST TWO WEEKS how relaxed versus anxious were you? | ◯ Very relaxed/calm<br>◯ Moderately relaxed/calm<br>◯ Neutral<br>◯ Moderately nervous/anxious<br>◯ Very nervous/anxious<br>◯ Prefer not to answer |
| During the PAST TWO WEEKS how fidgety or restless were you? | ◯ Not fidgety/restless at all<br>◯ Slightly fidgety/restless<br>◯ Moderately fidgety/restless<br>◯ Very fidgety/restless<br>◯ Extremely fidgety/restless<br>◯ Prefer not to answer |
| During the PAST TWO WEEKS how tired were you? | ◯ Not tired at all<br>◯ Slightly tired<br>◯ Moderately tired<br>◯ Very tired<br>◯ Extremely tired<br>◯ Prefer not to answer |
| During the PAST TWO WEEKS how well were you able to concentrate or focus? | ◯ Very focused/attentive<br>◯ Moderately focused/attentive<br>◯ Neutral<br>◯ Moderately unfocused/distracted<br>◯ Very unfocused/distracted<br>◯ Prefer not to answer |
| During the PAST TWO WEEKS how irritable or easily angered were you? | ◯ Not irritable or easily angered at all<br>◯ Slightly irritable or easily angered<br>◯ Moderately irritable or easily angered<br>◯ Very irritable or easily angered<br>◯ Extremely irritable or easily angered<br>◯ Prefer not to answer |
| During the PAST TWO WEEKS how lonely were you? | ◯ Not lonely at all<br>◯ Slightly lonely<br>◯ Moderately lonely<br>◯ Very lonely<br>◯ Extremely lonely<br>◯ Prefer not to answer |

| | |
|---|---|
| What have been your greatest sources of stress from the COVID-19 outbreak, since you completed the first survey? (Select all that apply) | ☐ Health concerns<br>☐ Financial concerns<br>☐ Impact on work<br>☐ Impact on your child<br>☐ Impact on your community<br>☐ Impact on family members<br>☐ Impact on the economy<br>☐ Access to food<br>☐ Access to baby supplies (e.g., formula, diapers, wipes)<br>☐ Access to personal care products or household supplies<br>☐ Access to medical care, including mental health care<br>☐ Social distancing or being quarantined<br>☐ Other<br>☐ I am not stressed about the COVID-19 outbreak<br>☐ Prefer not to answer |
| What have you done to cope with your stress related to the COVID-19 outbreak, since you completed the first survey? (Select all that apply) | ☐ Meditation and/or mindfulness practices<br>☐ Talking with friends and family (e.g., by phone, text, or video)<br>☐ Engaging in more family activities (e.g., games, sports)<br>☐ Increased television watching or other screen time activities (e.g., video games, social media)<br>☐ Eating more often, including snacking<br>☐ Increasing time reading books, or doing activities like puzzles and crosswords<br>☐ Drinking alcohol<br>☐ Using tobacco (e.g., smoking, vaping)<br>☐ Using marijuana (e.g., vaping, smoking, eating) or cannabidiol (CBD)<br>☐ Talking to my healthcare providers more frequently, including mental healthcare provider (e.g., therapist, psychologist, counselor)<br>☐ Volunteer work<br>☐ Exercise<br>☐ Other<br>☐ I have not done any of these things to cope with the stress related to the COVID-19 outbreak<br>☐ I am not stressed about the COVID-19 outbreak<br>☐ Prefer not to answer |

#### MEDIA USE (PAST TWO WEEKS)

During the PAST TWO WEEKS, how much time per day did you spend watching TV or digital media (e.g., Netflix, YouTube, web surfing) for educational or work purposes, including school work?

◯ No TV or digital media
◯ Under 1 hour
◯ 1-3 hours
◯ 4-6 hours
◯ More than 6 hours
◯ Prefer not to answer

During the PAST TWO WEEKS, how much time per day did you spend watching TV or digital media (e.g., Netflix, YouTube, web surfing) for NON-educational or NON-work purposes?

◯ No TV or digital media
◯ Under 1 hour
◯ 1-3 hours
◯ 4-6 hours
◯ More than 6 hours
◯ Prefer not to answer

During the PAST TWO WEEKS, how much time per day did you spend using social media (e.g.,Facebook, Instagram, Snapchat, Twitter, TikTok)?

◯ No social media
◯ Under 1 hour
◯ 1-3 hours
◯ 4-6 hours
◯ More than 6 hours
◯ Prefer not to answer

During the PAST TWO WEEKS, how much time per day did you spend playing video games?

◯ No video games
◯ Under 1 hour
◯ 1-3 hours
◯ 4-6 hours
◯ More than 6 hours
◯ Prefer not to answer

#### SUBSTANCE USE (Past TWO WEEKS)

| | Not at all | Rarely | Once a month | Several times a month | Once a week | Several times a week | Once a day | More than once a day | Prefer not to answer |
|---|---|---|---|---|---|---|---|---|---|
| During the PAST TWO WEEKS how frequently did you use alcohol? | ○ | ○ | ○ | ○ | ○ | ○ | ○ | ○ | ○ |
| During the PAST TWO WEEKS how frequently did you use vaping products? | ○ | ○ | ○ | ○ | ○ | ○ | ○ | ○ | ○ |
| During the PAST TWO WEEKS how frequently did you use cigarettes or other tobacco products? | ○ | ○ | ○ | ○ | ○ | ○ | ○ | ○ | ○ |
| During the PAST TWO WEEKS how frequently did you inhale marijuana/cannabis (e.g., joint, blunt, pipe, bong)? | ○ | ○ | ○ | ○ | ○ | ○ | ○ | ○ | ○ |
| During the PAST TWO WEEKS how frequently did you ingest/eat or apply marijuana/cannabis (e.g. edibles, pills, topical creams, patches, tinctures) | ○ | ○ | ○ | ○ | ○ | ○ | ○ | ○ | ○ |
| During the PAST TWO WEEKS how frequently did you use opiates, heroin, cocaine, crack, amphetamine, methamphetamine, hallucinogens, or ecstasy? | ○ | ○ | ○ | ○ | ○ | ○ | ○ | ○ | ○ |

**DEMOGRAPHIC INFORMATION**

**You completed your first survey on [cv19a_date].**

Has your living situation changed since you completed your first survey?

- ☐ No, my living situation has not changed
- ☐ Yes, I have moved (since the first survey)
- ☐ Yes, I regularly stay in more than one home (since the first survey)
- ☐ Yes, the number of people living in my home has changed (since the first survey)
- ☐ Prefer not to answer

Please provide the postal code of your primary home

__________________________________
(Enter "555" if you would prefer not to answer.)

How many adults currently live at your primary home (excluding yourself)?

- ◯ 0
- ◯ 1
- ◯ 2
- ◯ 3
- ◯ 4
- ◯ 5
- ◯ 6
- ◯ 7
- ◯ 8
- ◯ 9
- ◯ 10
- ◯ Prefer not to answer

How many children currently live at your primary home?

- ◯ 0
- ◯ 1
- ◯ 2
- ◯ 3
- ◯ 4
- ◯ 5
- ◯ 6
- ◯ 7
- ◯ 8
- ◯ 9
- ◯ 10
- ◯ Prefer not to answer

How many bedrooms are in your primary home?

- ◯ 0
- ◯ 1
- ◯ 2
- ◯ 3
- ◯ 4
- ◯ 5
- ◯ 6
- ◯ 7
- ◯ 8
- ◯ 9
- ◯ 10
- ◯ Prefer not to answer

| | |
|---|---|
| How many toilets are in your primary home? | ◯ 0<br>◯ 1<br>◯ 2<br>◯ 3<br>◯ 4<br>◯ 5<br>◯ 6<br>◯ 7<br>◯ 8<br>◯ 9<br>◯ 10<br>◯ Prefer not to answer |
| Choose the type of dwelling that best describes your primary home | ◯ Manufactured home/ mobile home or trailer<br>◯ Single family detached house<br>◯ Single family house attached to 1 or more houses (e.g townhouse/ semi-detached)<br>◯ High rise apartment/ condo/ co-op (4 or more floors)<br>◯ Low rise apartment/ condo/ co-op (1-3 floors)<br>◯ Multi-family home (more than one family in a converted single home)<br>◯ Other type<br>◯ Prefer not to answer |
| Please provide the postal code of your secondary home | ______________________________<br>(Enter "555" if you would prefer not to answer.) |
| How many adults currently live at your secondary home (excluding yourself)? | ◯ 0<br>◯ 1<br>◯ 2<br>◯ 3<br>◯ 4<br>◯ 5<br>◯ 6<br>◯ 7<br>◯ 8<br>◯ 9<br>◯ 10<br>◯ Prefer not to answer |
| How many children currently live at your secondary home? | ◯ 0<br>◯ 1<br>◯ 2<br>◯ 3<br>◯ 4<br>◯ 5<br>◯ 6<br>◯ 7<br>◯ 8<br>◯ 9<br>◯ 10<br>◯ Prefer not to answer |

| Question | Response options |
|---|---|
| How many bedrooms are in your secondary home? | ◯ 0<br>◯ 1<br>◯ 2<br>◯ 3<br>◯ 4<br>◯ 5<br>◯ 6<br>◯ 7<br>◯ 8<br>◯ 9<br>◯ 10<br>◯ Prefer not to answer |
| How many toilets are in your secondary home? | ◯ 0<br>◯ 1<br>◯ 2<br>◯ 3<br>◯ 4<br>◯ 5<br>◯ 6<br>◯ 7<br>◯ 8<br>◯ 9<br>◯ 10<br>◯ Prefer not to answer |
| Choose the type of dwelling that best describes your secondary home | ◯ Manufactured home/ mobile home or trailer<br>◯ Single family detached house<br>◯ Single family house attached to 1 or more houses (e.g townhouse/ semi-detached)<br>◯ High rise apartment/ condo/ co-op (4 or more floors)<br>◯ Low rise apartment/ condo/ co-op (1-3 floors)<br>◯ Multi-family home (more than one family in a converted single home)<br>◯ Other type<br>◯ Prefer not to answer |
| Have you been working in any of the following occupations or worksites since you completed your first survey? (Select all that apply) | ☐ Hospital or health care facility worker<br>☐ First responder (paramedic, firefighter, police officer)<br>☐ Childcare worker<br>☐ Teacher/ other school staff<br>☐ Transit Driver<br>☐ Food Service Industry<br>☐ Grocery Store<br>☐ Pharmacy<br>☐ Hairdresser/ barber<br>☐ Aesthetician<br>☐ Flight attendant<br>☐ Factory Worker<br>☐ None of the above<br>☐ Prefer not to answer |
| Are any adults living in any of your home(s) an ESSENTIAL WORKER? | ☐ Yes, I am<br>☐ Yes, another adult is<br>☐ No<br>☐ Don't Know<br>☐ Prefer not to answer<br>((e.g., healthcare, delivery worker, store worker, security, building maintenance)) |

| | |
|---|---|
| If yes, do they come home each day? | ☐ Yes, I do<br>☐ Yes, the other adult does<br>☐ No, separated due to COVID-19<br>☐ No separated due to other reasons<br>☐ Prefer not to answer |
| If yes, are they a FIRST RESPONDER, HEALTHCARE PROVIDER or OTHER WORKER in a facility treating COVID-19? | ☐ Yes, I am<br>☐ Yes, another adult is<br>☐ No<br>☐ Don't Know<br>☐ Prefer not to answer |
| Please share any comments, suggestions or clarifications that you have related to this survey | ________________________________ |
| Did [setup_arm_2][cv19binfo_firstname] complete this survey? | ◯ Yes - by themselves<br>◯ Yes - partially or with help from someone else<br>◯ No - someone else completed this survey on their behalf |
| Today's Date | __________________________ |

### Child Self Report Follow-up Survey 2

We are interested in learning more about how you are feeling and what you are thinking about during the Coronavirus (COVID-19) pandemic since you completed your second survey on [cv19c1_date] (Day-Month-Year).

COVID-19 is a new type of disease affecting many people in Canada. The virus that causes COVID-19 can cause runny or stuffy noses, coughing, fever, muscle aches and tiredness. Other people with COVID-19 may not have any symptoms at all but they can spread the virus to others.

It is important for scientists to learn more about COVID-19. You can help us by answering this survey. Do the best you can to answer each question. This survey should take between 20 to 30 minutes to complete.

If you have questions or want help filling the survey out, please ask a family member to help you.

---

In general, how would you rate your overall PHYSICAL health (how healthy your body is)?

- ◯ Excellent
- ◯ Very Good
- ◯ Good
- ◯ Fair
- ◯ Poor
- ◯ I don't know
- ◯ I don't want to answer

---

In general, how would you rate your overall MENTAL/EMOTIONAL health (the way you think and feel)?

- ◯ Excellent
- ◯ Very Good
- ◯ Good
- ◯ Fair
- ◯ Poor
- ◯ I don't know
- ◯ I don't want to answer

---

How tall are you (in centimeters)?

__________________________________
(Enter "666" if you don't know. Enter "555" if you don't want to answer.)

---

How much do you weigh (in kilograms)?

__________________________________
(Enter "666" if you don't know. Enter "555" if you don't want to answer.)

---

Have you had any of these health problems, since you filled in the second survey on [cv19c1_date]?
Check all of the problems you have had.

- ☐ Cough
- ☐ Shortness of breath
- ☐ Difficulty breathing
- ☐ Fever
- ☐ Chills
- ☐ Muscle or body aches
- ☐ Headache
- ☐ Sore throat
- ☐ New loss of sense of smell or taste
- ☐ Runny nose
- ☐ Other, Specify
- ☐ None of the above
- ☐ I don't know
- ☐ I don't want to answer

---

What other health problems have you had?

__________________________________
(Enter "666" if you don't know. Enter "555" if you don't want to answer.)

---

How often are you asking questions, reading, or talking about COVID-19, since you completed the second survey on [cv19c1_date]?

- ◯ Never
- ◯ A few times a month
- ◯ Weekly
- ◯ A few times a week
- ◯ Daily
- ◯ I don't know
- ◯ I don't want to answer

---

Has the COVID-19 pandemic in your area led to any POSITIVE (GOOD) changes in your life, since you completed the second survey on [cv19c1_date]?

- ☐ Yes
- ☐ No
- ☐ I don't know
- ☐ I don't want to answer

---

What are the POSITIVE (GOOD) changes in your life? (Select all that apply)

- ☐ More time with family
- ☐ More time with friends
- ☐ More exercising
- ☐ Healthier eating
- ☐ New hobbies
- ☐ Got a new pet
- ☐ Learned new technology
- ☐ Made new friends online
- ☐ Saved money
- ☐ Less commuting
- ☐ Better hand hygiene
- ☐ Less busy / more downtime
- ☐ Working from home
- ☐ Schooling from home
- ☐ More time outdoors
- ☐ Not getting sick as often
- ☐ Avoiding bullying
- ☐ Home improvements
- ☐ Improved mental health
- ☐ Other, specify
- ☐ I don't know
- ☐ I don't want to answer

---

What are the other POSITIVE (GOOD) changes in your life?

__________________________________
(Enter "666" if you don't know. Enter "555" if you don't want to answer.)

| | |
|---|---|
| Do you have your own cell phone (one that you do not have to share with anyone else)? | ◯ Yes<br>◯ No |

A vaccine is a type of medicine that doesn't make you better, but keeps you from getting sick in the first place. It does this by teaching your body to fight off germs like viruses and bacteria. The COVID-19 vaccine is given by a needle in the arm (shot) that will help your body be prepared to fight the COVID-19 virus in case you are exposed to it out in the world.

| | |
|---|---|
| Have you had at least one dose (shot) of the COVID-19 vaccine? | ◯ Yes<br>◯ No<br>◯ I don't know<br>◯ I don't want to answer<br>(Answer 'Yes' if you have received at least one dose of the COVID-19 vaccine. Note: Certain types of vaccines require more than one dose to protect against COVID-19. You would have been informed at the time of vaccination if you needed a second dose.) |
| Did you feel any side effects (feeling not well) from the COVID-19 vaccine? For example: sore arm, feeling tired, achy muscles, headache, fever and chills. | ◯ Yes<br>◯ No<br>◯ I don't know<br>◯ I don't want to answer |
| If yes, check off all the side effects you felt. | ☐ Head ache<br>☐ Tiredness<br>☐ Fever or chills<br>☐ Muscle or joint pain<br>☐ Soreness where the vaccine was injected<br>☐ Nausea or vomiting<br>☐ Other<br>☐ I don't know<br>☐ I don't want to answer |
| Please specify other side effects | __________________________________<br>(Enter "666" if you don't know. Enter "555" if you don't want to answer.) |
| How many doses (shots) of the COVID-19 vaccine have you had so far? | ◯ One dose<br>◯ Two doses<br>◯ Three doses<br>◯ Four doses<br>◯ More than four doses<br>◯ I don't know<br>◯ I don't want to answer |
| When did you receive your first dose of the COVID-19 vaccine? | __________________________________<br>(If you do not recall the exact date, please provide an estimate. Enter "01-01-1900" if you would prefer not to answer.) |

| | |
|---|---|
| Which vaccine did you receive? (first dose) | ◯ Pfizer and BioNTech mRNA vaccine<br>◯ Moderna mRNA vaccine<br>◯ AstraZeneca Oxford vaccine<br>◯ Janssen (Johnson & Johnson) vaccine<br>◯ Other, specify<br>◯ I don't know<br>◯ I don't want to answer |
| Specify the other vaccine | __________________________________<br>(Enter "666" if you don't know. Enter "555" if you don't want to answer.) |
| When did you receive your second dose of the COVID-19 vaccine? | __________________________________<br>(If you do not recall the exact date, please provide an estimate. Enter "01-01-1900" if you would prefer not to answer.) |
| Which vaccine did you receive? (second dose) | ◯ Pfizer and BioNTech mRNA vaccine<br>◯ Moderna mRNA vaccine<br>◯ AstraZeneca Oxford vaccine<br>◯ Janssen (Johnson & Johnson) vaccine<br>◯ Other, specify<br>◯ I don't know<br>◯ I don't want to answer |
| Specify the other vaccine | __________________________________<br>(Enter "666" if you don't know. Enter "555" if you don't want to answer.) |
| When did you receive your third dose of the COVID-19 vaccine? | __________________________________<br>(If you do not recall the exact date, please provide an estimate. Enter "01-01-1900" if you would prefer not to answer.) |
| Which vaccine did you receive? (third dose) | ◯ Pfizer and BioNTech mRNA vaccine<br>◯ Moderna mRNA vaccine<br>◯ AstraZeneca Oxford vaccine<br>◯ Janssen (Johnson & Johnson) vaccine<br>◯ Other, specify<br>◯ I don't know<br>◯ I don't want to answer |
| Specify the other vaccine | __________________________________<br>(Enter "666" if you don't know. Enter "555" if you don't want to answer.) |
| When did you receive your fourth dose of the COVID-19 vaccine? | __________________________________<br>(If you do not recall the exact date, please provide an estimate. Enter "01-01-1900" if you would prefer not to answer.) |

| | |
|---|---|
| Which vaccine did you receive? (fourth dose) | ◯ Pfizer and BioNTech mRNA vaccine<br>◯ Moderna mRNA vaccine<br>◯ AstraZeneca Oxford vaccine<br>◯ Janssen (Johnson & Johnson) vaccine<br>◯ Other, specify<br>◯ I don't know<br>◯ I don't want to answer |
| Specify the other vaccine | ______________________________<br>(Enter "666" if you don't know. Enter "555" if you don't want to answer.) |
| Are you old enough to receive a COVID-19 vaccine? | ◯ Yes and I plan on getting a COVID-19 shot<br>◯ Yes I'm old enough, but I DO NOT plan on getting a COVID-19 shot<br>◯ No, I am not old enough yet<br>◯ I don't know<br>◯ I don't want to answer |
| What are some of the reasons why you don't want to get the COVID-19 vaccine? | ☐ Not confident in the safety of the vaccine<br>☐ Concern about risks and side effects<br>☐ Will wait until it seems safe to get the vaccine<br>☐ Do not consider it necessary to get the vaccine<br>☐ Do not believe in vaccination<br>☐ Have not yet decided<br>☐ Have a pre-existing medical condition<br>☐ Already had or think I have had COVID-19<br>☐ Other, Specify<br>☐ I don't know<br>☐ I don't want to answer |
| What are the other reasons you don't want to get the COVID-19 vaccine? | ______________________________<br>(Enter "666" if you don't know. Enter "555" if you don't want to answer.) |
| When you are able to get the COVID-19 vaccine, how likely is it that you will choose to get it? | ◯ Very Likely<br>◯ Somewhat Likely<br>◯ Somewhat Unlikely<br>◯ Very Unlikely<br>◯ I don't know<br>◯ I don't want to answer |

**A vaccine is a shot that you get at the doctor's office so you won't get sick. We are interested in finding out how you feel about vaccines.**

| | Strongly Agree | Agree | Disagree | Strongly Disagree | I don't know | I don't want to answer |
|---|---|---|---|---|---|---|
| Getting the COVID-19 vaccine is important so I can help protect the health of other people. | ◯ | ◯ | ◯ | ◯ | ◯ | ◯ |

| | | | | | | |
|---|---|---|---|---|---|---|
| Getting vaccines is a good way to protect myself from getting sick. | ○ | ○ | ○ | ○ | ○ | ○ |
| I usually do what my doctor or healthcare provider recommends about getting a vaccine. | ○ | ○ | ○ | ○ | ○ | ○ |
| I worry about feeling sick or having side effects after getting a vaccine. | ○ | ○ | ○ | ○ | ○ | ○ |

**Social Support**

**We would like to measure your feelings about how supported you feel. Choose 1 of the 4 responses to show whether you agree or disagree with the statement.**

| | Strongly Agree | Agree | Disagree | Strongly Disagree | I don't know | I don't want to answer |
|---|---|---|---|---|---|---|
| I have family and friends who help me feel safe, secure and happy | ○ | ○ | ○ | ○ | ○ | ○ |
| There is someone I trust whom I would turn to for advice if I were having problems | ○ | ○ | ○ | ○ | ○ | ○ |
| There are people I can count on in an emergency | ○ | ○ | ○ | ○ | ○ | ○ |
| I feel part of a group of people who share my attitudes and beliefs | ○ | ○ | ○ | ○ | ○ | ○ |

**The following are a number of mental health services that you may have accessed. Please check the relevant boxes for each service indicating if you have accessed the service since you completed the second survey on [cv19c1_date]  or if you would be interested in accessing the service.**

| | Have accessed this service since completing the second survey on [cv19c1_date] | Would be interested in accessing this service | Not accessed, not interested | I don't know | I don't want to answer |
|---|---|---|---|---|---|
| Individual therapy (you alone) with a professional (in person or virtual) | ○ | ○ | ○ | ○ | ○ |
| Group therapy (you and others) with a professional (in person or virtual) | ○ | ○ | ○ | ○ | ○ |

| | | | | | |
|---|---|---|---|---|---|
| Family therapy (you and your family members) with a professional (in person or virtual) | ○ | ○ | ○ | ○ | ○ |
| Mental health mobile app (general) | ○ | ○ | ○ | ○ | ○ |
| Mental health mobile app to interact with a therapist | ○ | ○ | ○ | ○ | ○ |
| Phone helpline for mental health support (e.g. Crisis Services Canada) | ○ | ○ | ○ | ○ | ○ |
| Online mental health information you looked up yourself | ○ | ○ | ○ | ○ | ○ |

**Families have different routines.**

**We would like to know about your family's routines. Choose 1 of the 4 responses that best fits the statement with your family's routine.**

| | Always | Often | Rarely | Never | I don't know | I don't want to answer |
|---|---|---|---|---|---|---|
| Talking with your parents about your day each day | ○ | ○ | ○ | ○ | ○ | ○ |
| Playing with your parents each day | ○ | ○ | ○ | ○ | ○ | ○ |
| Eating dinner together almost every night | ○ | ○ | ○ | ○ | ○ | ○ |
| Letting your parents know where you are going when you are about to leave the house or where you've been when you get home | ○ | ○ | ○ | ○ | ○ | ○ |
| Regular chores | ○ | ○ | ○ | ○ | ○ | ○ |
| Reading books together almost every day | ○ | ○ | ○ | ○ | ○ | ○ |
| Going to bed at the same time every night | ○ | ○ | ○ | ○ | ○ | ○ |

#### LIFE CHANGES DUE TO THE COVID-19 PANDEMIC

Based on what you know at this time, how safe do you think it is for students in your community 12 years of age and OLDER to attend school in-person?

- ○ Very safe
- ○ Somewhat safe
- ○ Not very safe
- ○ Not safe at all
- ○ I don't Know
- ○ I don't want to answer

---

Based on what you know at this time, how safe do you think it is for students in your community YOUNGER than 12 years of age to attend school in-person?

- ◯ Very safe
- ◯ Somewhat safe
- ◯ Not very safe
- ◯ Not safe at all
- ◯ I don't Know
- ◯ I don't want to answer

---

Since you completed the second survey on [cv19c1_date], has the quality of the relationships between you and members of your family changed?

- ◯ A lot better
- ◯ A little better
- ◯ About the same
- ◯ A little worse
- ◯ A lot worse
- ◯ I don't know
- ◯ I don't want to answer

---

Since you completed the second survey on [cv19c1_date], how stressful have these changes in relationships between you and members of your family been for you?

- ◯ Not stressful
- ◯ A little bit stressful
- ◯ Stressful
- ◯ Very stressful
- ◯ Extremely stressful
- ◯ I don't know
- ◯ I don't want to answer

---

Since you completed the second survey on [cv19c1_date], has the quality of your relationships with your friends changed?

- ◯ A lot better
- ◯ A little better
- ◯ About the same
- ◯ A little worse
- ◯ A lot worse
- ◯ I don't know
- ◯ I don't want to answer

---

Since you completed the second survey on [cv19c1_date], how stressful have these changes in your relationships with your friends been for you?

- ◯ Not stressful
- ◯ A little bit stressful
- ◯ Stressful
- ◯ Very stressful
- ◯ Extremely stressful
- ◯ I don't know
- ◯ I don't want to answer

---

Since you completed the second survey on [cv19c1_date], how difficult has the cancellation of important events in your life (such as sporting events, school trips or vacations, etc.) been for you?

- ◯ Not difficult
- ◯ A little bit difficult
- ◯ Difficult
- ◯ Very difficult
- ◯ Extremely difficult
- ◯ There hasn't been any cancellation of important events
- ◯ I don't know
- ◯ I don't want to answer

---

During the PAST TWO WEEKS, how many people (who do not live with you) have you talked to in-person in an indoor environment?

__________________________________
(In-person means that the person is in the same place as you (NOT on the phone or video chat) Enter "666" if you don't know. Enter "555" if you don't want to answer.)

| | |
|---|---|
| During the PAST TWO WEEKS, how many people (who do not live with you) have you talked to in-person in an outdoor environment? | ________________________________ (In-person means that the person is in the same place as you (NOT on the phone or video chat) Enter "666" if you don't know. Enter "555" if you don't want to answer.) |
| During the PAST TWO WEEKS, how many times did you go outside of your home (e.g., going to stores, parks, etc.)? | ◯ Not at all<br>◯ 1-2 days per week<br>◯ 3-4 days per week<br>◯ 5-6 days per week<br>◯ Every day<br>◯ I don't know<br>◯ I don't want to answer |
| During the PAST TWO WEEKS, how stressful have the restrictions on leaving home been for you? | ◯ Not stressful<br>◯ A little bit stressful<br>◯ Stressful<br>◯ Very stressful<br>◯ Extremely stressful<br>◯ There are no restrictions on leaving home<br>◯ I don't know<br>◯ I don't want to answer |
| During the PAST TWO WEEKS, how difficult has it been for you to follow the recommendations for keeping away from close contact with people who you don't live with? | ◯ Not difficult<br>◯ A little bit difficult<br>◯ Difficult<br>◯ Very difficult<br>◯ Extremely difficult<br>◯ There are no recommendations for keeping away from close contact with people who I don't live with<br>◯ I don't know<br>◯ I don't want to answer |
| How hopeful are you that the COVID-19 pandemic in your area will end soon? | ◯ Extremely hopeful<br>◯ Very hopeful<br>◯ Hopeful<br>◯ A little bit hopeful<br>◯ Neither hopeful nor not hopeful<br>◯ Not at all hopeful<br>◯ There is no COVID-19 pandemic in my area<br>◯ I don't know<br>◯ I don't want to answer |

#### DAILY BEHAVIOURS (PAST TWO WEEKS)

| | |
|---|---|
| During the PAST TWO WEEKS, have you experienced problems with sleep? (e.g., trouble falling asleep, staying asleep, waking up much earlier than intended, sleeping alone without a parent/caregiver) | ◯ Yes<br>◯ No<br>◯ I don't know<br>◯ I don't want to answer |
| During the PAST TWO WEEKS, what time did you go to bed on WEEKDAYS (on average)? | ◯ Before 8 pm<br>◯ 8 pm - 10 pm<br>◯ 10 pm - 12 am (Midnight)<br>◯ After 12 am (Midnight)<br>◯ I don't know<br>◯ I don't want to answer |

| | |
|---|---|
| During the PAST TWO WEEKS, what time did you go to bed on WEEKENDS (on average)? | ◯ Before 8 pm<br>◯ 8 pm - 10 pm<br>◯ 10 pm - 12 am (Midnight)<br>◯ After 12 am (Midnight)<br>◯ I don't know<br>◯ I don't want to answer |
| During the PAST TWO WEEKS, how many hours per night did you sleep on WEEKDAYS (on average)? | ◯ Less than 6 hours<br>◯ 6-8 hours<br>◯ 8-10 hours<br>◯ Greater than 10 hours<br>◯ I don't know<br>◯ I don't want to answer |
| During the PAST TWO WEEKS, how many hours per night did you sleep on WEEKENDS (on average)? | ◯ Less than 6 hours<br>◯ 6-8 hours<br>◯ 8-10 hours<br>◯ Greater than 10 hours<br>◯ I don't know<br>◯ I don't want to answer |
| During the PAST TWO WEEKS, how many days per week did you exercise (where your heart rate and breathing increased) for at least 30 minutes over the entire day? | ◯ None<br>◯ 1-2 days<br>◯ 3-4 days<br>◯ 5-6 days<br>◯ Daily<br>◯ I don't know<br>◯ I don't want to answer |
| During the PAST TWO WEEKS, how many days per week did you spend time outdoors? | ◯ None<br>◯ 1-2 days<br>◯ 3-4 days<br>◯ 5-6 days<br>◯ Daily<br>◯ I don't know<br>◯ I don't want to answer |

#### EMOTIONS/WORRIES (PAST TWO WEEKS)

| | Not at all worried | Slightly worried | Moderately worried | Very worried | Extremely worried | I don't know | I don't want to answer |
|---|---|---|---|---|---|---|---|
| During the PAST TWO WEEKS, how worried have you been about being infected by the virus that causes COVID-19? | ◯ | ◯ | ◯ | ◯ | ◯ | ◯ | ◯ |
| During the PAST TWO WEEKS, how worried have you been about friends or family being infected by the virus that causes COVID-19? | ◯ | ◯ | ◯ | ◯ | ◯ | ◯ | ◯ |

| | | | | | | | |
|---|---|---|---|---|---|---|---|
| During the PAST TWO WEEKS, how worried have you been about your physical health (how healthy your body is) being influenced by COVID-19? | ◯ | ◯ | ◯ | ◯ | ◯ | ◯ | ◯ |
| During the PAST TWO WEEKS, how worried have you been about your mental/emotional health (how you think and feel) being influenced by COVID-19? | ◯ | ◯ | ◯ | ◯ | ◯ | ◯ | ◯ |
| During the PAST TWO WEEKS, how worried were you generally? | ◯ | ◯ | ◯ | ◯ | ◯ | ◯ | ◯ |

---

During the PAST TWO WEEKS, how happy versus sad were you?

◯ Very happy/cheerful
◯ Moderately happy/cheerful
◯ Neutral
◯ Moderately sad/depressed/unhappy
◯ Very sad/depressed/unhappy
◯ I don't know
◯ I don't want to answer

---

During the PAST TWO WEEKS, how relaxed versus anxious were you?

◯ Very relaxed/calm
◯ Moderately relaxed/calm
◯ Neutral
◯ Moderately nervous/anxious
◯ Very nervous/anxious
◯ I don't know
◯ I don't want to answer

---

During the PAST TWO WEEKS, how fidgety or restless were you?

◯ Not fidgety/restless at all
◯ Slightly fidgety/restless
◯ Moderately fidgety/restless
◯ Very fidgety/restless
◯ Extremely fidgety/restless
◯ I don't know
◯ I don't want to answer

---

During the PAST TWO WEEKS, how tired were you?

◯ Not  tired at all
◯ Slightly tired
◯ Moderately tired
◯ Very tired
◯ Extremely tired
◯ I don't know
◯ I don't want to answer

---

During the PAST TWO WEEKS, how well were you able to concentrate or focus?

◯ Very focused/attentive
◯ Moderately focused/attentive
◯ Neutral
◯ Moderately unfocused/distracted
◯ Very unfocused/distracted
◯ I don't know
◯ I don't want to answer

| | |
|---|---|
| During the PAST TWO WEEKS, how irritable or easily angered were you? | ◯ Not irritable or easily angered at all<br>◯ Slightly irritable or easily angered<br>◯ Moderately irritable or easily angered<br>◯ Very irritable or easily angered<br>◯ Extremely irritable or easily angered<br>◯ I don't know<br>◯ I don't want to answer |
| During the PAST TWO WEEKS, how lonely were you? | ◯ Not lonely at all<br>◯ Slightly lonely<br>◯ Moderately lonely<br>◯ Very lonely<br>◯ Extremely lonely<br>◯ I don't know<br>◯ I don't want to answer |

**MEDIA USE (PAST TWO WEEKS)**

| | |
|---|---|
| During the PAST TWO WEEKS, how much time PER WEEKDAY did you spend watching TV or digital media (e.g., Netflix, YouTube, web surfing)? | ◯ No TV or digital media<br>◯ Under 1 hour<br>◯ 1-3 hours<br>◯ 4-6 hours<br>◯ More than 6 hours<br>◯ I don't know<br>◯ I don't want to answer<br>(Including time for educational purposes and/or homework) |
| During the PAST TWO WEEKS, how much time PER WEEKEND DAY did you spend watching TV or digital media (e.g., Netflix, YouTube, web surfing)? | ◯ No TV or digital media<br>◯ Under 1 hour<br>◯ 1-3 hours<br>◯ 4-6 hours<br>◯ More than 6 hours<br>◯ I don't know<br>◯ I don't want to answer<br>(Including time for educational purposes and/or homework) |
| During the PAST TWO WEEKS, how much time PER WEEKDAY did you spend using social media (e.g., Facebook, Instagram, Snapchat, Twitter, TikTok)? | ◯ No social media<br>◯ Under 1 hour<br>◯ 1-3 hours<br>◯ 4-6 hours<br>◯ More than 6 hours<br>◯ I don't know<br>◯ I don't want to answer<br>(Including time for educational purposes and/or homework) |
| During the PAST TWO WEEKS, how much time PER WEEKEND DAY did you spend using social media (e.g., Facebook, Instagram, Snapchat, Twitter, TikTok)? | ◯ No social media<br>◯ Under 1 hour<br>◯ 1-3 hours<br>◯ 4-6 hours<br>◯ More than 6 hours<br>◯ I don't know<br>◯ I don't want to answer<br>(Including time for educational purposes and/or homework) |

| | |
|---|---|
| During the PAST TWO WEEKS, how much time PER WEEKDAY did you spend playing video games? | ◯ No video games<br>◯ Under 1 hour<br>◯ 1-3 hours<br>◯ 4-6 hours<br>◯ More than 6 hours<br>◯ I don't know<br>◯ I don't want to answer |
| During the PAST TWO WEEKS, how much time PER WEEKEND DAY did you spend playing video games? | ◯ No video games<br>◯ Under 1 hour<br>◯ 1-3 hours<br>◯ 4-6 hours<br>◯ More than 6 hours<br>◯ I don't know<br>◯ I don't want to answer |
| Please share any comments, suggestions or clarifications that you have related to this survey | ______________________________ |
| Did [setup_arm_3][cv19binfo_firstname] complete this survey? | ◯ Yes - by themselves<br>◯ Yes - partially or with help from someone else<br>◯ No - someone else completed this survey on their behalf<br>◯ I don't know<br>◯ I don't want to answer |
| Today's Date | ________________________ |

### Parental Follow-up Survey 2

We are interested in learning more about how the child is feeling and what they are thinking about during the Coronavirus (COVID-19) pandemic since you completed your second survey on [cv19p1_date] (Day-Month-Year).

COVID-19 is a new type of illness affecting many people in Canada. The virus that causes COVID-19 is related to the common cold virus. Some people affected by the virus may have symptoms that can be mild or severe, similar to a cold or flu. Other people with COVID-19 may not have any symptoms at all but they can spread the virus to others.

It is important for scientists to learn more about COVID-19. You can help us by answering this survey about the child. Do the best you can to answer each question. This survey should take between 20 to 30 minutes to complete.

In general, how would you rate the child's overall PHYSICAL health now?

- ◯ Excellent
- ◯ Very Good
- ◯ Good
- ◯ Fair
- ◯ Poor
- ◯ Prefer not to answer

In general, how would you rate the child's overall MENTAL/EMOTIONAL health now?

- ◯ Excellent
- ◯ Very Good
- ◯ Good
- ◯ Fair
- ◯ Poor
- ◯ Prefer not to answer

Child's height in centimeters?

__________________________________
(Enter "555" if you would prefer not to answer.)

Child's weight in kilograms?

__________________________________
(Enter "555" if you would prefer not to answer.)

In what ways has the COVID-19 outbreak affected the child's overall healthcare, since you completed the second survey on [cv19p1_date]? (Select all that apply)

- ☐ The child did not go to healthcare appointments because I was concerned about entering the healthcare provider's office
- ☐ The child's healthcare provider cancelled appointments
- ☐ The child's healthcare provider changed to phone or online visits
- ☐ The child's healthcare provider told him/her to self-isolate or quarantine
- ☐ None of these apply
- ☐ Prefer not to answer

| | |
|---|---|
| What kind of healthcare visits were affected since you completed the second survey on [cv19p1_date]? (Select all that apply) | ☐ Routine visit/ regular checkup<br>☐ Vaccination visit<br>☐ Visit with a health care professional for a specific concern<br>☐ Visit with a specialist<br>☐ I did not miss any healthcare visits<br>☐ Prefer not to answer |
| Has the COVID-19 pandemic in the child's area led to any family members in the child's household(s) applying for or accessing any of the following supports, since you completed the second survey on [cv19p1_date]? (Select all that apply) | ☐ Residential Mortgage or Lease/ Rent Payment Deferral<br>☐ Commercial Mortgage or Lease/ Rent Payment Deferral<br>☐ Personal Income Support (e.g Canadian Emergency Response Benefit (CERB), Canadian Emergency Student Benefit (CESB), Employment Insurance (EI))<br>☐ Business Income Support (e.g Canadian Emergency Wage Subsidy (CEWS))<br>☐ Food Bank<br>☐ Other, Specify<br>☐ None of the above<br>☐ Don't Know<br>☐ Prefer not to answer |
| Specify other supports | __________________________________<br>(Enter "555" if you would prefer not to answer.) |
| Has the child been exposed to someone likely to have COVID-19, since you completed the second survey on [cv19p1_date]? (Select all that apply) | ☐ Yes, to someone with positive test<br>☐ Yes, to someone with medical diagnosis, but no test<br>☐ Yes, to someone with possible symptoms, but no diagnosis by doctor<br>☐ No, not to my knowledge<br>☐ Prefer not to answer |
| Has the child been suspected of having COVID-19, since you completed the second survey on [cv19p1_date]? | ◯ Yes, had positive test<br>◯ Yes, had medical diagnosis, but no test<br>◯ Yes, had some possible symptoms, but no diagnosis by health care professional<br>◯ No<br>◯ Don't know<br>◯ Prefer not to answer |
| Has the child been tested for the virus that causes COVID-19, since you completed the second survey on [cv19p1_date]? (Select all that apply)<br><br>Do not include the antibody/ serology test (blood test for previous infection) that is part of this research study. | ☐ No, I never tried to get the child tested<br>☐ No, I tried to get the child tested but was not able to<br>☐ Yes, and the child is waiting for the results<br>☐ Yes, and the test showed that the child did not have it ("negative" test)<br>☐ Yes, and the test showed that the child did have it ("positive" test)<br>☐ Prefer not to answer |

| | |
|---|---|
| Which type of test for the virus that causes COVID-19 did you try to get for the child but were not able to? | ☐ Viral Test (swab test for current infection)<br>☐ Antibody/ Serology Test (blood test for previous infection)<br>☐ Saliva or Mouth Rinse Test<br>☐ Rapid-Antigen Test<br>☐ Other Specify<br>☐ Don't Know<br>☐ Prefer not to answer<br>(Antibody/ Serology testing and saliva/ mouth rinse testing may not be currently available in all areas, but it may be available in the future.) |
| Specify the other type of test for the virus that causes COVID-19 that you tried to get for the child but were not able to. | ______________________________<br>(Enter "555" if you would prefer not to answer.) |
| Please specify why the child got tested or tried to get tested | ☐ A contact or possible contact of the child tested positive for COVID-19<br>☐ The child had COVID-19 symptoms<br>☐ The child was admitted to hospital (unrelated to COVID-19)<br>☐ The child was admitted to hospital (COVID-19 related)<br>☐ The child was tested in another facility or instiution context (contact with someone in long term care, prison etc)<br>☐ The child was participating in another facility research study<br>☐ Travel-related testing<br>☐ Other, specify<br>☐ Don't Know<br>☐ Prefer not to answer |
| Please specify the other reason(s) why the child got tested or tried to get tested | ______________________________<br>(Enter "555" if you would prefer not to answer.) |
| Please specify why you didn't try to get the child tested | ☐ Symptoms were due to another condition, specify<br>☐ Symptoms were mild<br>☐ A healthcare provider did not recommend testing<br>☐ Other, specify<br>☐ Prefer not to answer |
| Please specify the condition that you believe the symptoms are from | ______________________________<br>( Enter "555" if you would prefer not to answer.) |
| Please specify the other reason(s) you didn't try to get the child tested | ______________________________<br>( Enter "555" if you would prefer not to answer.) |
| IN THE PAST TWO WEEKS, how many times has the child been tested for COVID-19? | ◯ 0<br>◯ 1<br>◯ 2<br>◯ 3<br>◯ 4<br>◯ Prefer not to answer |
| Details of First COVID-19 Test | |

| | |
|---|---|
| What type of test did the child receive? | ◯ Viral Test (swab test for current infection)<br>◯ Antibody/ Serology Test (blood test for previous infection)<br>◯ Saliva or Mouth Rinse Test<br>◯ Rapid-Antigen Test<br>◯ Other Specify<br>◯ Don't Know<br>◯ Prefer not to answer<br>(Do not include the antibody/ serology test (blood test for previous infection) that is part of this reseach study ) |
| Specify other type of test the child received. | ______________________________<br>( Enter "555" if you would prefer not to answer.) |
| Date of test | ______________________________<br>(If you do not recall the exact date, please provide an estimate. Enter "01-01-1900" if you would prefer not to answer.) |
| First test result | ◯ Negative<br>◯ Positive<br>◯ Don't know<br>◯ Prefer not to answer |
| Details of Second COVID-19 Test | |
| What type of test did the child receive? | ◯ Viral Test (swab test for current infection)<br>◯ Antibody/ Serology Test (blood test for previous infection)<br>◯ Saliva or Mouth Rinse Test<br>◯ Rapid-Antigen Test<br>◯ Other Specify<br>◯ Don't Know<br>◯ Prefer not to answer<br>(Do not include the antibody/ serology test (blood test for previous infection) that is part of this reseach study ) |
| Specify other type of test the child received. | ______________________________<br>(Enter "555" if you would prefer not to answer.) |
| Date of test | ______________________________<br>(If you do not recall the exact date, please provide an estimate. Enter "01-01-1900" if you would prefer not to answer.) |
| Second test result | ◯ Negative<br>◯ Positive<br>◯ Don't know<br>◯ Prefer not to answer |
| Details of Third COVID-19 Test | |

| | |
|---|---|
| What type of test did the child receive? | ◯ Viral Test (swab test for current infection)<br>◯ Antibody/ Serology Test (blood test for previous infection)<br>◯ Saliva or Mouth Rinse Test<br>◯ Rapid-Antigen Test<br>◯ Other Specify<br>◯ Don't Know<br>◯ Prefer not to answer<br>(Do not include the antibody/ serology test (blood test for previous infection) that is part of this reseach study ) |
| Specify other type of test the child received. | __________________________________<br>(Enter "555" if you would prefer not to answer.) |
| Date of test | __________________________________<br>(If you do not recall the exact date, please provide an estimate. Enter "01-01-1900" if you would prefer not to answer.) |
| Third test result | ◯ Negative<br>◯ Positive<br>◯ Don't know<br>◯ Prefer not to answer |
| Details of Fourth COVID-19 Test | |
| What type of test did the child receive? | ◯ Viral Test (swab test for current infection)<br>◯ Antibody/ Serology Test (blood test for previous infection)<br>◯ Saliva or Mouth Rinse Test<br>◯ Rapid-Antigen Test<br>◯ Other Specify<br>◯ Don't Know<br>◯ Prefer not to answer<br>(Do not include the antibody/ serology test (blood test for previous infection) that is part of this reseach study ) |
| Specify other type of test the child received. | __________________________________<br>(Enter "555" if you would prefer not to answer.) |
| Date of test | __________________________________<br>(If you do not recall the exact date, please provide an estimate. Enter "01-01-1900" if you would prefer not to answer.) |
| Fourth test result | ◯ Negative<br>◯ Positive<br>◯ Don't know<br>◯ Prefer not to answer |

| | |
|---|---|
| Has the child had any colds, since you completed the second survey on [cv19p1_date]? | ◯ Yes<br>◯ No<br>◯ Don't Know<br>◯ Prefer not to answer |
| Has the child had any of the following symptoms, since you completed the second survey on [cv19p1_date]? (Select all that apply) | ☐ New or worsening cough<br>☐ Shortness of breath<br>☐ Difficulty breathing<br>☐ Fever<br>☐ Chills<br>☐ Muscle or body aches<br>☐ Headache<br>☐ Sore throat<br>☐ New loss of sense of smell or taste<br>☐ Runny nose or nasal congestion*<br>☐ Nausea or vomiting<br>☐ Diarrhea<br>☐ Fatigue<br>☐ Skin Rash<br>☐ Conjunctivitis (pink eye)<br>☐ Other, Specify<br>☐ None of the above<br>☐ Prefer not to answer<br>(*Runny nose or nasal congestion in the absence of underlying reason for these symptoms such as seasonal allergies, post nasal drip, etc) |
| Specify the other symptoms that the child had since you completed the second survey on [cv19p1_date]? | ____________________________________<br>( Enter "555" if you would prefer not to answer.) |
| Do you remember the date when the child had their first symptom? | ◯ Yes<br>◯ No<br>◯ Roughly<br>◯ Prefer not to answer |
| What was the date of the child's first symptom? | ____________________________________<br>(If you do not recall the exact date, please provide an estimate. Enter "01-01-1900" if you would prefer not to answer.) |
| In the TWO WEEKS before the child had symptoms, did the child: (Select all that apply) | ☐ Have contact with someone who tested positive for COVID-19<br>☐ Have contact with someone who likely had COVID-19 (e.g., was not tested but had symptoms; was told by a healthcare provider that he/she likely had it)<br>☐ Travel outside the province<br>☐ None of the above<br>☐ Prefer not to answer |
| How many places did the child travel? | ◯ 0<br>◯ 1<br>◯ 2<br>◯ 3<br>◯ 4<br>◯ Prefer not to answer<br>(Travel outside the province) |
| Place 1 | |

Location
(e.g. Seattle, Washington OR Toronto, Ontario)

__________________________________
(Enter "555" if you would prefer not to answer.)

Start date of travel

__________________________________
(If you do not recall the exact date, please provide an estimate. Enter "01-01-1900" if you would prefer not to answer.)

End date of travel

__________________________________
(If you do not recall the exact date, please provide an estimate. Enter "01-01-1900" if you would prefer not to answer.)

How? (Check all that apply)

- ☐ Car
- ☐ Plane
- ☐ Train
- ☐ Ship
- ☐ Other
- ☐ Prefer not to answer

Please specify other mode of transportation

__________________________________
(Enter "555" if you would prefer not to answer.)

Place 2

Location
(e.g. Portland, Oregon OR Vancouver, BC)

__________________________________
(Enter "555" if you would prefer not to answer.)

Start date of travel

__________________________________
(If you do not recall the exact date, please provide an estimate. Enter "01-01-1900" if you would prefer not to answer.)

End date of travel

__________________________________
(If you do not recall the exact date, please provide an estimate. Enter "01-01-1900" if you would prefer not to answer.)

How? (Check all that apply)

- ☐ Car
- ☐ Plane
- ☐ Train
- ☐ Ship
- ☐ Other
- ☐ Prefer not to answer

Please specify other mode of transportation

__________________________________
(Enter "555" if you would prefer not to answer.)

Place 3

| | |
|---|---|
| Location<br>(e.g. Paris, France OR Banff, Alberta) | ______________________________<br>(Enter "555" if you would prefer not to answer.) |
| Start date of travel | ______________________________<br>(If you do not recall the exact date, please provide an estimate. Enter "01-01-1900" if you would prefer not to answer.) |
| End date of travel | ______________________________<br>(If you do not recall the exact date, please provide an estimate. Enter "01-01-1900" if you would prefer not to answer.) |
| How? (Check all that apply) | ☐ Car<br>☐ Plane<br>☐ Train<br>☐ Ship<br>☐ Other<br>☐ Prefer not to answer |
| Please specify other mode of transportation | ______________________________<br>(Enter "555" if you would prefer not to answer.) |
| Place 4 | |
| Location<br>(e.g. Portland, Oregon OR Vancouver, BC) | ______________________________<br>(Enter "555" if you would prefer not to answer.) |
| Start date of travel | ______________________________<br>(If you do not recall the exact date, please provide an estimate. Enter "01-01-1900" if you would prefer not to answer.) |
| End date of travel | ______________________________<br>(If you do not recall the exact date, please provide an estimate. Enter "01-01-1900" if you would prefer not to answer.) |
| How? (Check all that apply) | ☐ Car<br>☐ Plane<br>☐ Train<br>☐ Ship<br>☐ Other<br>☐ Prefer not to answer |
| Please specify other mode of transportation | ______________________________<br>(Enter "555" if you would prefer not to answer.) |

| | |
|---|---|
| Which of the following occurred as a result of the child's symptoms? (Select all that apply) | ☐ You/ the child spoke to a healthcare provider over the phone, by email, or online<br>☐ The child saw a healthcare provider in person, such as in a clinic or doctor's office<br>☐ The child saw a healthcare provider in person, such as in urgent care, or in an Emergency Department (ED)<br>☐ The child was kept overnight in a hospital because a healthcare provider thought he/she had COVID-19<br>☐ The child was admitted to a hospital's ICU (Intensive Care Unit)<br>☐ The child self-isolated or quarantined at home<br>☐ None of the above<br>☐ Prefer not to answer |
| Has anyone in the child's family been diagnosed with COVID-19, since you completed the second survey on [cv19p1_date]? (Select all that apply) | ☐ Yes, member of household<br>☐ Yes, non-household member<br>☐ No<br>☐ Don't know<br>☐ Prefer not to answer |
| Have any of the following happened to the child's family members because of COVID-19, since you completed the second survey on [cv19p1_date]? (Select all that apply) | ☐ Fallen physically ill<br>☐ Hospitalized<br>☐ Self-isolated or quarantined with symptoms<br>☐ Self-isolated or quarantined without symptoms (e.g., due to possible exposure)<br>☐ Lost job or been laid off from job<br>☐ Reduced ability to earn money<br>☐ Passed away<br>☐ None of the above<br>☐ Prefer not to answer<br>(Include any family members, not just those living in the same household as the child) |
| Did the child get the flu shot, since you completed the second survey on [cv19p1_date]? | ◯ Yes<br>◯ No<br>◯ Don't Know<br>◯ Prefer not to answer |
| Has the child received at least one dose of a COVID-19 vaccine? | ◯ Yes<br>◯ No<br>◯ Prefer not to answer<br>(Answer 'Yes' if the child has received at least one dose of the COVID-19 vaccine. Note: Certain types of vaccines require more than one dose to protect against COVID-19. You or the child would have been informed at the time of vaccination if the child needed a second dose.) |
| Did the child feel any side effects from the COVID-19 vaccine? | ◯ Yes<br>◯ No<br>◯ Prefer not to answer |

| | |
|---|---|
| If yes, check all that apply. | ☐ Head ache<br>☐ Tiredness<br>☐ Fever or chills<br>☐ Muscle or joint pain<br>☐ Soreness where the vaccine was injected<br>☐ Nausea or vomiting<br>☐ Other<br>☐ Prefer not to answer |
| Please specify other side effects | __________________________________<br>(Enter "555" if you would prefer not to answer.) |
| How many doses of the COVID-19 vaccine has the child received so far? | ◯ One dose<br>◯ Two doses<br>◯ Three doses<br>◯ Four doses<br>◯ More than four doses<br>◯ Prefer not to answer |
| When did the child receive their first dose of the COVID-19 vaccine? | __________________________________<br>(If you do not recall the exact date, please provide an estimate. Enter "01-01-1900" if you would prefer not to answer.) |
| Which vaccine did the child receive? (first dose) | ◯ Pfizer and BioNTech mRNA vaccine<br>◯ Moderna mRNA vaccine<br>◯ AstraZeneca Oxford vaccine<br>◯ Janssen (Johnson & Johnson) vaccine<br>◯ Other, specify<br>◯ Don't Know<br>◯ Prefer not to answer |
| Specify the other vaccine | __________________________________<br>(Enter "555" if you would prefer not to answer.) |
| When did the child receive their second dose of the COVID-19 vaccine? | __________________________________<br>(If you do not recall the exact date, please provide an estimate. Enter "01-01-1900" if you would prefer not to answer.) |
| Which vaccine did the child receive? (second dose) | ◯ Pfizer and BioNTech mRNA vaccine<br>◯ Moderna mRNA vaccine<br>◯ AstraZeneca Oxford vaccine<br>◯ Janssen (Johnson & Johnson) vaccine<br>◯ Other, specify<br>◯ Don't Know<br>◯ Prefer not to answer |
| Specify the other vaccine | __________________________________<br>(Enter "555" if you would prefer not to answer.) |

| | |
|---|---|
| When did the child receive their third dose of the COVID-19 vaccine? | ______________________________ (If you do not recall the exact date, please provide an estimate. Enter "01-01-1900" if you would prefer not to answer.) |
| Which vaccine did the child receive? (third dose) | ◯ Pfizer and BioNTech mRNA vaccine<br>◯ Moderna mRNA vaccine<br>◯ AstraZeneca Oxford vaccine<br>◯ Janssen (Johnson & Johnson) vaccine<br>◯ Other, specify<br>◯ Don't Know<br>◯ Prefer not to answer |
| Specify the other vaccine | ______________________________ (Enter "555" if you would prefer not to answer.) |
| When did the child receive their fourth dose of the COVID-19 vaccine? | ______________________________ (If you do not recall the exact date, please provide an estimate. Enter "01-01-1900" if you would prefer not to answer.) |
| Which vaccine did the child receive? (fourth dose) | ◯ Pfizer and BioNTech mRNA vaccine<br>◯ Moderna mRNA vaccine<br>◯ AstraZeneca Oxford vaccine<br>◯ Janssen (Johnson & Johnson) vaccine<br>◯ Other, specify<br>◯ Don't Know<br>◯ Prefer not to answer |
| Specify the other vaccine | ______________________________ (Enter "555" if you would prefer not to answer.) |
| Is the child eligible to receive a COVID-19 vaccine? | ◯ Yes and we plan to schedule a COVID-19 vaccination for the child<br>◯ Yes, but we DO NOT plan to schedule a COVID-19 vaccination for the child<br>◯ No, the child is not eligible for a COVID-19 vaccine yet<br>◯ Don't know<br>◯ Prefer not to answer |
| What are some of the reasons why you don't want THE CHILD to get the COVID-19 vaccine? | ☐ Not confident in the safety of the vaccine<br>☐ Concern about risks and side effects<br>☐ Will wait until it seems safe to get the vaccine<br>☐ Do not consider it necessary to get the vaccine<br>☐ Do not believe in vaccination<br>☐ Have not yet decided<br>☐ Have a pre-existing medical condition<br>☐ Already had or think I have had COVID-19<br>☐ Other, Specify<br>☐ Prefer not to answer<br>(Check all that apply.) |
| What were the other reasons you didn't want THE CHILD to get the COVID-19 vaccine? | ______________________________ ( Enter "555" if you would prefer not to answer.) |

When a COVID-19 vaccine becomes available, how likely is it that you will choose for THE CHILD to get it?

- ◯ Very Likely
- ◯ Somewhat Likely
- ◯ Somewhat Unlikely
- ◯ Very Unlikely
- ◯ Don't Know
- ◯ Prefer not to answer

| | Strongly Agree | Agree | Disagree | Strongly Disagree | Prefer not to answer |
|---|---|---|---|---|---|
| Having the child vaccinated is important for the health of others in my community. | ◯ | ◯ | ◯ | ◯ | ◯ |
| Getting vaccines is a good way to protect the child/children from disease. | ◯ | ◯ | ◯ | ◯ | ◯ |
| Generally I do what my doctor or health care provider recommends about vaccines for the child/children. | ◯ | ◯ | ◯ | ◯ | ◯ |

| | Never | Rarely | Occasionall y | Often | Always | Don't Know | Prefer not to answer |
|---|---|---|---|---|---|---|---|
| How often has the child worn a mask in public places since you completed the second survey on [cv19p1_date]? | ◯ | ◯ | ◯ | ◯ | ◯ | ◯ | ◯ |
| How often has the child practiced physical distancing in public places since you completed the second survey on [cv19p1_date]? | ◯ | ◯ | ◯ | ◯ | ◯ | ◯ | ◯ |
| How often has the child avoided crowded places/ gatherings since you completed the second survey on [cv19p1_date]? | ◯ | ◯ | ◯ | ◯ | ◯ | ◯ | ◯ |
| How often has the child avoided common greetings (e.g handshakes, hugs etc) since you completed the second survey on [cv19p1_date]? | ◯ | ◯ | ◯ | ◯ | ◯ | ◯ | ◯ |
| How often has the child limited contact with people at higher risk (e.g. elderly relative) since you completed the second survey on [cv19p1_date]? | ◯ | ◯ | ◯ | ◯ | ◯ | ◯ | ◯ |

| | | | | | | | |
|---|---|---|---|---|---|---|---|
| How often has the child self-Isolated because someone thought the child was infected with the virus that causes COVID-19 since you completed the second survey on [cv19p1_date]? | ◯ | ◯ | ◯ | ◯ | ◯ | ◯ | ◯ |
| How often has the child quarantined because the child may have been exposed to the virus that causes COVID-19, but did not show symptoms since you completed the second survey on [cv19p1_date]? | ◯ | ◯ | ◯ | ◯ | ◯ | ◯ | ◯ |

---

Has the child participated in any organized sports or recreational activities, in person, since you completed the second survey on [cv19p1_date]?

◯ Yes
◯ No
◯ Prefer not to answer

---

If yes, how many children were in the activity?

◯ 0-9
◯ 10-19
◯ 20-29
◯ 30+
◯ Prefer not to answer

---

How many times has the child been in a gathering* of >10 or more people (excluding school), since you completed the second survey on [cv19p1_date]?

__________________________________
(A gathering in this context is defined as any assembly or meeting of more than 10 persons, in open air or in a building or premises. These can include gatherings with or without masks or social distancing. Gatherings include a range of interpersonal gatherings/events of varying purposes, from gatherings/events of family and friends (e.g., sports, weddings, funerals, baptisms, birthday parties and cultural ceremonies) to community gathering spaces (e.g., places of worship, places of traditional gatherings, libraries, community and recreation centres, camps, drop-in centres, food banks and communal kitchens, museums, theatres, cinemas, and tourist attractions) to larger planned or structured gatherings/events, including mass gatherings (e.g., large meetings or conferences, national sporting events, large cultural/religious events, festivals). Enter "555" if you would prefer not to answer.)

**The following are a number of mental health services that the child may have accessed. Please check the relevant boxes for each service indicating if the child accessed the service since completing the second survey on [cv19p1_date] or if the child would be interested in accessing the service.**

| | Have accessed this service since completing the second survey on [cv19p1_date] | Would be interested in accessing this service | Not accessed, not interested | Prefer not to answer |
|---|---|---|---|---|
| Individual therapy (you alone) with a professional (in person or virtual) | ○ | ○ | ○ | ○ |
| Group therapy (you and others) with a professional (in person or virtual) | ○ | ○ | ○ | ○ |
| Family therapy (you and your family members) with a professional (in person or virtual) | ○ | ○ | ○ | ○ |
| Mental health mobile app (general) | ○ | ○ | ○ | ○ |
| Mental health mobile app to interact with a therapist | ○ | ○ | ○ | ○ |
| Phone helpline for mental health support (e.g. Crisis Services Canada) | ○ | ○ | ○ | ○ |
| Online mental health information you looked up yourself | ○ | ○ | ○ | ○ |

40% Complete

**Worrying (PAST TWO WEEKS)**

| | Not at all worried | Slightly worried | Moderately worried | Very worried | Extremely worried | Prefer not to answer |
|---|---|---|---|---|---|---|
| During the PAST TWO WEEKS, how worried has the child been about being infected by the virus that causes COVID-19? | ○ | ○ | ○ | ○ | ○ | ○ |
| During the PAST TWO WEEKS, how worried has the child been about friends or family being infected by the virus that causes COVID-19? | ○ | ○ | ○ | ○ | ○ | ○ |

| | | | | | | |
|---|---|---|---|---|---|---|
| During the PAST TWO WEEKS, how worried has the child been about his/her physical health being influenced by COVID-19? | ◯ | ◯ | ◯ | ◯ | ◯ | ◯ |
| During the PAST TWO WEEKS, how worried has the child been about his/her mental/emotional health being influenced by COVID-19? | ◯ | ◯ | ◯ | ◯ | ◯ | ◯ |
| During the PAST TWO WEEKS, how worried was the child generally? | ◯ | ◯ | ◯ | ◯ | ◯ | ◯ |

---

How much is the child asking questions, reading, or talking about COVID-19, since you completed the second survey on [cv19p1_date]?

◯ Never
◯ A few times a month
◯ Weekly
◯ A few times a week
◯ Daily
◯ Prefer not to answer

---

Has the COVID-19 pandemic in the child's area led to any POSITIVE changes in the child's life, since you completed the second survey on [cv19p1_date]?

☐ Yes
☐ No
☐ I don't know
☐ Prefer not to answer

---

Please specify these POSITIVE changes in the child's life

☐ More time with family
☐ More time with friends
☐ More exercising
☐ Healthier eating
☐ New hobbies
☐ Other, specify
☐ Prefer not to answer

---

Please specify the other POSITIVE changes in the child's life

__________________________________
(Enter "555" if you would prefer not to answer.)

---

Does the child have a personal cell phone (one that the child doesn't not have to share with anyone else)?

◯ Yes
◯ No

---

Based on what you know at this time, how safe do you think it is for students in your community 12 years of age and OLDER to attend school in-person?

◯ Very safe
◯ Somewhat safe
◯ Not very safe
◯ Not safe at all
◯ Don't Know
◯ Prefer not to answer

---

Based on what you know at this time, how safe do you think it is for students in your community YOUNGER than 12 years of age to attend school in-person?

◯ Very safe
◯ Somewhat safe
◯ Not very safe
◯ Not safe at all
◯ Don't Know
◯ Prefer not to answer

| | |
|---|---|
| Is the child enrolled in school for the current academic year*? | ◯ Not in school<br>◯ Preschool or daycare<br>◯ Home school<br>◯ Elementary school<br>◯ Junior High or Middle School<br>◯ Secondary or High School<br>◯ Prefer not to answer<br>(* Current academic year = Sept 2021 to June 2022) |
| Has the the child's school building closed since you completed the second survey on [cv19p1_date]? | ◯ Yes<br>◯ No<br>◯ N/A<br>◯ Prefer not to answer |
| If no, are they attending classes in person? | ◯ Yes<br>◯ No<br>◯ N/A<br>◯ Prefer not to answer |
| If yes, how many times did the school close? | __________________________________<br>(Enter "666" if you don't know the answer.Enter "555" if you would prefer not to answer.) |
| If yes, when did the first school building closure begin? | __________________________________<br>(If you do not recall the exact date, please provide an estimate. Enter "01-01-1900" if you would prefer not to answer.) |
| How many weeks in total was the school building closed since September 2021 due to COVID-19? | __________________________________<br>((do not include Christmas and spring break) Enter "555" if you would prefer not to answer.) |
| If yes, were classes resumed online? | ◯ Yes<br>◯ No<br>◯ N/A<br>◯ Prefer not to answer |
| Did the child participate in classes online since September 2021? | ◯ Yes<br>◯ No<br>◯ Partially<br>◯ N/A<br>◯ Prefer not to answer |
| If yes, did the child have easy access to the internet and a computer? | ◯ Yes<br>◯ No<br>◯ N/A<br>◯ Prefer not to answer |
| If yes, were there assignments for the child to complete? | ◯ Yes<br>◯ No<br>◯ N/A<br>◯ Prefer not to answer |

| | |
|---|---|
| Did the child complete the assignments? | ◯ Yes<br>◯ No<br>◯ Partially<br>◯ N/A<br>◯ Prefer not to answer |
| Did the child physically return to school since the first closure? | ◯ Yes<br>◯ No<br>◯ N/A<br>◯ Prefer not to answer |
| When did the child physically return to school? | __________________________<br>(If you do not recall the exact date, please provide an estimate. Enter "01-01-1900" if you would prefer not to answer.) |
| How many days of in-school or remote/online learning has the child missed/ been absent for, since you completed the second survey on [cv19p1_date]? | __________________________<br>(Enter "555" if you would prefer not to answer.) |
| Since you completed the second survey on [cv19p1_date], has the quality of the relationships between the child and members of his/her family changed? | ◯ A lot better<br>◯ A little better<br>◯ About the same<br>◯ A little worse<br>◯ A lot worse<br>◯ Don't know<br>◯ Prefer not to answer |
| Since you completed the second survey on [cv19p1_date], how stressful have these changes in relationships between the child and members of his/her family been for the child? | ◯ Not stressful<br>◯ A little bit stressful<br>◯ Stressful<br>◯ Very stressful<br>◯ Extremely stressful<br>◯ Don't know<br>◯ Prefer not to answer |
| Since you completed the second survey on [cv19p1_date], has the quality of the child's relationships with his/her friends changed? | ◯ A lot better<br>◯ A little better<br>◯ About the same<br>◯ A little worse<br>◯ A lot worse<br>◯ Don't know<br>◯ Prefer not to answer |
| Since you completed the second survey on [cv19p1_date], how stressful have these changes in relationships between the child and his/her friends been for the child? | ◯ Not stressful<br>◯ A little bit stressful<br>◯ Stressful<br>◯ Very stressful<br>◯ Extremely stressful<br>◯ Don't Know<br>◯ Prefer not to answer |

| | |
|---|---|
| Since you completed the second survey on [cv19p1_date], how difficult has the cancellation of important events in the child's life (such as sporting events, school trips or vacations, etc.) been difficult for the child? | ◯ Not difficult<br>◯ A little bit difficult<br>◯ Difficult<br>◯ Very difficult<br>◯ Extremely difficult<br>◯ There hasn't been any cancellation of important events<br>◯ Prefer not to answer |
| During the PAST TWO WEEKS, how many people, from outside the child's household, has the child had an in-person conversation with in an indoor environment? This can include things like work, school or other activities with or without masks and/or social distancing. If you aren't sure, please estimate a number. | ___________________________________<br>(In-person means that the person is in the same place as the child (NOT on the phone or video chat) Enter "555" if you would prefer not to answer.) |
| During the PAST TWO WEEKS, how many people, from outside the child's household, has the child had an in-person conversation with in an outdoor environment? This can include things like work, school or other activities with or without masks and/or social distancing. If you aren't sure, please estimate a number. | ___________________________________<br>(In-person means that the person is in the same place as the child (NOT on the phone or video chat) Enter "555" if you would prefer not to answer.) |
| During the PAST TWO WEEKS, how many times did the child go outside of their home (e.g., going to stores, parks, etc)? | ◯ Not at all<br>◯ 1-2 days per week or less<br>◯ 3-4 days per week<br>◯ 5-6 days per week<br>◯ Daily<br>◯ Prefer not to answer |
| During the PAST TWO WEEKS, how stressful have the restrictions on leaving home been for the child? | ◯ Not stressful<br>◯ A little bit stressful<br>◯ Stressful<br>◯ Very stressful<br>◯ Extremely stressful<br>◯ There are no restrictions on leaving home<br>◯ Prefer not to answer |
| During the PAST TWO WEEKS, how difficult has it been for the child to follow the recommendations for keeping away from close contact with people who they don't live with? | ◯ Not difficult<br>◯ A little bit difficult<br>◯ Difficult<br>◯ Very difficult<br>◯ Extremely difficult<br>◯ There are no recommendations for keeping away from close contact with people who they don't live with<br>◯ Prefer not to answer |
| How hopeful is the child that the COVID-19 pandemic in the child's area will end soon? | ◯ Extremely hopeful<br>◯ Very hopeful<br>◯ Hopeful<br>◯ A little bit hopeful<br>◯ Neither hopeful nor not hopeful<br>◯ Not at all hopeful<br>◯ There is no COVID-19 pandemic in the child's area<br>◯ Prefer not to answer |

60% Complete

**Compared to before the COVID-19 outbreak, how much is the child now:**

| | Less | Same amount | More | Prefer not to answer |
|---|---|---|---|---|
| Eating | ◯ | ◯ | ◯ | ◯ |
| Sleeping | ◯ | ◯ | ◯ | ◯ |
| Physically active | ◯ | ◯ | ◯ | ◯ |
| Spending time outside | ◯ | ◯ | ◯ | ◯ |
| Spending time with friends in-person | ◯ | ◯ | ◯ | ◯ |
| Spending time with friends remotely (e.g., online, social media, texting) | ◯ | ◯ | ◯ | ◯ |
| Spending time watching TV, playing video/computer games, or using social media for educational purposes, including school work | ◯ | ◯ | ◯ | ◯ |
| Spending time watching TV, playing video/computer games, or using social media for NON-educational purposes | ◯ | ◯ | ◯ | ◯ |

**DAILY BEHAVIOURS (PAST TWO WEEKS)**

| | |
|---|---|
| During the PAST TWO WEEKS, has the child experienced problems with sleep? (e.g., trouble falling asleep, staying asleep, waking up much earlier than intended, sleeping alone without a parent/caregiver) | ◯ Yes<br>◯ No<br>◯ Prefer not to answer |
| During the PAST TWO WEEKS, what time did the child go to bed on WEEKDAYS (on average)? | ◯ Before 8 pm<br>◯ 8 pm - 10 pm<br>◯ 10 pm - 12 am (Midnight)<br>◯ After 12 am (Midnight)<br>◯ Prefer not to answer |
| During the PAST TWO WEEKS, what time did the child go to bed on WEEKENDS (on average)? | ◯ Before 8 pm<br>◯ 8 pm - 10 pm<br>◯ 10 pm - 12 am (Midnight)<br>◯ After 12 am (Midnight)<br>◯ Prefer not to answer |
| During the PAST TWO WEEKS, how many hours per night did the child sleep on WEEKDAYS (on average)? | ◯ Less than 6 hours<br>◯ 6-8 hours<br>◯ 8-10 hours<br>◯ Greater than 10 hours<br>◯ Prefer not to answer |
| During the PAST TWO WEEKS, how many hours per night did the child sleep on WEEKENDS (on average)? | ◯ Less than 6 hours<br>◯ 6-8 hours<br>◯ 8-10 hours<br>◯ Greater than 10 hours<br>◯ Prefer not to answer |

| | |
|---|---|
| During the PAST TWO WEEKS, how many days per week did the child exercise (e.g., increased heart rate, breathing) for at least 30 minutes over the entire day? | ◯ Not at all<br>◯ 1-2 days per week or less<br>◯ 3-4 days per week<br>◯ 5-6 days per week<br>◯ Daily<br>◯ Prefer not to answer |
| During the PAST TWO WEEKS, how many days per week did the child spend time outdoors? | ◯ Not at all<br>◯ 1-2 days per week or less<br>◯ 3-4 days per week<br>◯ 5-6 days per week<br>◯ Daily<br>◯ Prefer not to answer |

#### EMOTIONS/WORRIES (PAST TWO WEEKS)

| | |
|---|---|
| During the PAST TWO WEEKS, how happy versus sad was the child? | ◯ Very happy/cheerful<br>◯ Moderately happy/cheerful<br>◯ Neutral<br>◯ Moderately sad/depressed/unhappy<br>◯ Very sad/depressed/unhappy<br>◯ Prefer not to answer |
| During the PAST TWO WEEKS, how relaxed versus anxious was the child? | ◯ Very relaxed/calm<br>◯ Moderately relaxed/calm<br>◯ Neutral<br>◯ Moderately nervous/anxious<br>◯ Very nervous/anxious<br>◯ Prefer not to answer |
| During the PAST TWO WEEKS, how fidgety or restless was the child? | ◯ Not fidgety/restless at all<br>◯ Slightly fidgety/restless<br>◯ Moderately fidgety/restless<br>◯ Very fidgety/restless<br>◯ Extremely fidgety/restless<br>◯ Prefer not to answer |
| During the PAST TWO WEEKS, how tired was the child? | ◯ Not tired at all<br>◯ Slightly tired<br>◯ Moderately tired<br>◯ Very tired<br>◯ Extremely tired<br>◯ Prefer not to answer |
| During the PAST TWO WEEKS, how well was the child able to concentrate or focus? | ◯ Very focused/attentive<br>◯ Moderately focused/attentive<br>◯ Neutral<br>◯ Moderately unfocused/distracted<br>◯ Very unfocused/distracted<br>◯ Prefer not to answer |
| During the PAST TWO WEEKS, how irritable or easily angered was the child? | ◯ Not irritable or easily angered at all<br>◯ Slightly irritable or easily angered<br>◯ Moderately irritable or easily angered<br>◯ Very irritable or easily angered<br>◯ Extremely irritable or easily angered<br>◯ Prefer not to answer |

| | |
|---|---|
| During the PAST TWO WEEKS, how lonely was the child? | ◯ Not lonely at all<br>◯ Slightly lonely<br>◯ Moderately lonely<br>◯ Very lonely<br>◯ Extremely lonely<br>◯ Prefer not to answer |

#### MEDIA USE (PAST TWO WEEKS)

| | |
|---|---|
| During the PAST TWO WEEKS, how much time PER WEEKDAY did the child spend watching TV or digital media (e.g., Netflix, YouTube, web surfing) for NON-educational purposes? | ◯ No TV or digital media<br>◯ Under 1 hour<br>◯ 1-3 hours<br>◯ 4-6 hours<br>◯ More than 6 hours<br>◯ Prefer not to answer |
| During the PAST TWO WEEKS, how much time PER WEEKEND DAY did the child spend watching TV or digital media (e.g., Netflix, YouTube, web surfing) for NON-educational purposes? | ◯ No TV or digital media<br>◯ Under 1 hour<br>◯ 1-3 hours<br>◯ 4-6 hours<br>◯ More than 6 hours<br>◯ Prefer not to answer |
| During the PAST TWO WEEKS, how much time PER WEEKDAY did the child spend using social media (e.g., Facebook, Instagram, Snapchat, Twitter, TikTok)? | ◯ No social media<br>◯ Under 1 hour<br>◯ 1-3 hours<br>◯ 4-6 hours<br>◯ More than 6 hours<br>◯ Prefer not to answer |
| During the PAST TWO WEEKS, how much time PER WEEKEND DAY did the child spend using social media (e.g., Facebook, Instagram, Snapchat, Twitter, TikTok)? | ◯ No social media<br>◯ Under 1 hour<br>◯ 1-3 hours<br>◯ 4-6 hours<br>◯ More than 6 hours<br>◯ Prefer not to answer |
| During the PAST TWO WEEKS, how much time PER WEEKDAY did the child spend playing video games for NON-EDUCATIONAL purposes? | ◯ No video games<br>◯ Under 1 hour<br>◯ 1-3 hours<br>◯ 4-6 hours<br>◯ More than 6 hours<br>◯ Prefer not to answer<br>(Including time for educational purposes and/or homework) |
| During the PAST TWO WEEKS, how much time PER WEEKEND DAY did the child spend playing video games for NON-EDUCATIONAL purposes? | ◯ No video games<br>◯ Under 1 hour<br>◯ 1-3 hours<br>◯ 4-6 hours<br>◯ More than 6 hours<br>◯ Prefer not to answer<br>(Including time for educational purposes and/or homework) |

**Below are statements about families. You are to decide which of these statements are true of your family and which are false. If you think the statement is True or mostly True of your family, mark True. If you think the statement is False or mostly False of your family, mark False.**

**You may feel that some of the statements are true for some family members and false for others. Mark True if the statement is true for most members. Mark False if the statement is false for most members. If the members are evenly divided, decide what is the stronger overall impression and answer accordingly.**

**Remember, we would like to know what your family seems like to you. So do not try to figure out how other members see your family, but do give us your general impression of your family for each statement.**

**Selected Items from Family Environment Scale (FES) Instrument copyright 1974, 2002 by Rudolf H Moos. All rights reserved in all media. Published by Mind Garden, Inc., www.mindgarden.com**

| | True | False | Prefer not to answer |
|---|---|---|---|
| Family members often criticise each other | ○ | ○ | ○ |
| Family members sometimes get so angry they throw things | ○ | ○ | ○ |
| Family members often try to one-up or out-do each other | ○ | ○ | ○ |
| We fight a lot in our family | ○ | ○ | ○ |
| Family members sometimes hit each other | ○ | ○ | ○ |
| In our family, we believe you don't ever get anywhere by raising your voice | ○ | ○ | ○ |
| If there's a disagreement in our family, we try hard to smooth things over and keep the peace | ○ | ○ | ○ |
| Family members hardly ever lose their tempers | ○ | ○ | ○ |
| Family members rarely become openly angry | ○ | ○ | ○ |

#### DEMOGRAPHIC INFORMATION

| Question | Response |
|---|---|
| Has the child's living situation changed since you completed the second survey on [cv19p1_date]? | ☐ No, the child's living situation has not changed<br>☐ Yes, the child has have moved (since the second survey)<br>☐ Yes, the child regularly stays in more than one home (since the second survey)<br>☐ Yes, the number of people living in the child's home has changed (since the second survey)<br>☐ Prefer not to answer |
| Please provide the postal code of the child's primary home | ________________________<br>(Enter "555" if you would prefer not to answer.) |
| How many adults currently live at the child's primary home? | ◯ 0<br>◯ 1<br>◯ 2<br>◯ 3<br>◯ 4<br>◯ 5<br>◯ 6<br>◯ 7<br>◯ 8<br>◯ 9<br>◯ 10<br>◯ Prefer not to answer |
| How many children currently live at the child's primary home (excluding the child)? | ◯ 0<br>◯ 1<br>◯ 2<br>◯ 3<br>◯ 4<br>◯ 5<br>◯ 6<br>◯ 7<br>◯ 8<br>◯ 9<br>◯ 10<br>◯ Prefer not to answer |
| How many bedrooms are in the child's primary home? | ◯ 0<br>◯ 1<br>◯ 2<br>◯ 3<br>◯ 4<br>◯ 5<br>◯ 6<br>◯ 7<br>◯ 8<br>◯ 9<br>◯ 10<br>◯ Prefer not to answer |

| | |
|---|---|
| How many toilets are in the child's primary home? | ◯ 0<br>◯ 1<br>◯ 2<br>◯ 3<br>◯ 4<br>◯ 5<br>◯ 6<br>◯ 7<br>◯ 8<br>◯ 9<br>◯ 10<br>◯ Prefer not to answer |
| Choose the type of dwelling that best describes the child's primary home | ◯ Manufactured home/ mobile home or trailer<br>◯ Single family detached house<br>◯ Single family house attached to 1 or more houses (e.g townhouse/ semi-detached)<br>◯ High rise apartment/ condo/ co-op (4 or more floors)<br>◯ Low rise apartment/ condo/ co-op (1-3 floors)<br>◯ Multi-family home (more than one family in a converted single home)<br>◯ Other type<br>◯ Prefer not to answer |
| Please provide the postal code of the child's secondary home | ______________________________<br>(Enter "555" if you would prefer not to answer.) |
| How many adults currently live at the child's secondary home? | ◯ 0<br>◯ 1<br>◯ 2<br>◯ 3<br>◯ 4<br>◯ 5<br>◯ 6<br>◯ 7<br>◯ 8<br>◯ 9<br>◯ 10<br>◯ Prefer not to answer |
| How many children currently live at the child's secondary home (excluding the child)? | ◯ 0<br>◯ 1<br>◯ 2<br>◯ 3<br>◯ 4<br>◯ 5<br>◯ 6<br>◯ 7<br>◯ 8<br>◯ 9<br>◯ 10<br>◯ Prefer not to answer |

| | |
|---|---|
| How many bedrooms are in the child's secondary home? | ◯ 0<br>◯ 1<br>◯ 2<br>◯ 3<br>◯ 4<br>◯ 5<br>◯ 6<br>◯ 7<br>◯ 8<br>◯ 9<br>◯ 10<br>◯ Prefer not to answer |
| How many toilets are in the child's secondary home? | ◯ 0<br>◯ 1<br>◯ 2<br>◯ 3<br>◯ 4<br>◯ 5<br>◯ 6<br>◯ 7<br>◯ 8<br>◯ 9<br>◯ 10<br>◯ Prefer not to answer |
| Choose the type of dwelling that best describes the child's secondary home | ◯ Manufactured home/ mobile home or trailer<br>◯ Single family detached house<br>◯ Single family house attached to 1 or more houses (e.g townhouse/ semi-detached)<br>◯ High rise apartment/ condo/ co-op (4 or more floors)<br>◯ Low rise apartment/ condo/ co-op (1-3 floors)<br>◯ Multi-family home (more than one family in a converted single home)<br>◯ Other type<br>◯ Prefer not to answer |
| Are any adults living in the home(s) considered to be an ESSENTIAL WORKER? | ☐ Yes, I am<br>☐ Yes, another adult is<br>☐ No<br>☐ Don't Know<br>☐ Prefer not to answer<br>((e.g., healthcare, delivery worker, store worker, security, building maintenance)) |
| If yes, do they come home each day? | ☐ Yes, I do<br>☐ Yes, the other adult does<br>☐ No, separated due to COVID-19<br>☐ No separated due to other reasons<br>☐ Prefer not to answer |
| If yes, are they a FIRST RESPONDER, HEALTHCARE PROVIDER or OTHER WORKER in a facility treating COVID-19? | ☐ Yes, I am<br>☐ Yes, another adult is<br>☐ No<br>☐ Don't Know<br>☐ Prefer not to answer |
| Please share any comments, suggestions or clarifications that you have related to this survey | ________________________________ |

What is your relationship to the child?

◯ Biological Mother
◯ Biological Father
◯ Parent
◯ Step Parent
◯ Foster Parent
◯ Grandparent
◯ Great-Grandparent
◯ Aunt or Uncle
◯ Sibling
◯ Step Sibling
◯ Cousin
◯ Other, Caregiver
◯ Prefer not to answer

Today's Date

__________________________________

### Adult Self Report Follow-up Survey 2



We are interested in learning more about how you are feeling and what you are thinking about during the Coronavirus (COVID-19) pandemic since you completed your second survey on [cv19a1_date] (Day-Month-Year).

COVID-19 is a new type of illness affecting many people in Canada. The virus that causes COVID-19 is related to the common cold virus. Some people affected by the virus may have symptoms that can be mild or severe, similar to a cold or flu. Other people with COVID-19 may not have any symptoms at all but they can spread the virus to others.

It is important for scientists to learn more about COVID-19. You can help us understand more about how COVID-19 affects families by answering this survey. Do the best you can to answer each question. This survey should take between 20 to 30 minutes to complete.

If you have questions or want help filling the survey out, please contact your site's Coordinator.

---

In general, how would you rate your overall PHYSICAL health now?

◯ Excellent
◯ Very Good
◯ Good
◯ Fair
◯ Poor
◯ Prefer not to answer

---

In general, how would you rate your overall MENTAL/EMOTIONAL health now?

◯ Excellent
◯ Very Good
◯ Good
◯ Fair
◯ Poor
◯ Prefer not to answer

---

What is your current height in centimeters?

__________________________________
(Enter "555" if you would prefer not to answer.)

---

What is your current weight in kilograms?

__________________________________
(Enter "555" if you would prefer not to answer.)

#### COVID-19 HEALTH/EXPOSURE STATUS

| | |
|---|---|
| In what ways has the COVID-19 outbreak affected your overall healthcare, since you completed the second survey on [cv19a1_date]? (Select all that apply) | ☐ I did not go to healthcare appointments because I was concerned about entering the healthcare providers office<br>☐ My healthcare provider cancelled appointments<br>☐ My healthcare provider changed to phone or online visits<br>☐ My healthcare provider told me to self-isolate or quarantine<br>☐ None of these apply<br>☐ Prefer not to answer |
| What kind of healthcare visits were affected since you completed the second survey on [cv19a1_date]? (Select all that apply) | ☐ Routine visit/ regular checkup<br>☐ Vaccination visit<br>☐ Visit with a health care professional for a specific concern<br>☐ Visit with a specialist<br>☐ I did not miss any healthcare visits<br>☐ Prefer not to answer |
| Has the COVID-19 pandemic in your area led to any family members in your household(s) applying for any of the following government supports, since you completed the second survey on [cv19a1_date]? (Select all that apply) | ☐ Residential Mortgage or Lease/ Rent Payment Deferral<br>☐ Commercial Mortgage or Lease/ Rent Payment Deferral<br>☐ Personal Income Support (e.g Canadian Emergency Response Benefit (CERB), Canadian Emergency Student Benefit (CESB), Employment Insurance (EI))<br>☐ Business Income Support (e.g Canadian Emergency Wage Subsidy (CEWS))<br>☐ Food Bank<br>☐ Other, Specify<br>☐ None of the above<br>☐ Don't know<br>☐ Prefer not to answer |
| Specify other supports | ______________________________<br>(Enter "555" if you would prefer not to answer.) |
| Have you been exposed to someone likely to have COVID-19, since you completed the second survey on [cv19a1_date]? (Select all that apply) | ☐ Yes, to someone with positive test<br>☐ Yes, to someone with medical diagnosis, but no test<br>☐ Yes, to someone with possible symptoms, but no diagnosis by doctor<br>☐ No, not to my knowledge<br>☐ Prefer not to answer |
| Have you been suspected of having COVID-19, since you completed the second survey on [cv19a1_date]? | ☐ Yes, had positive test<br>☐ Yes, had medical diagnosis, but no test<br>☐ Yes, had some possible symptoms, but no diagnosis by health care professional<br>☐ No<br>☐ Prefer not to answer |
| Have you been tested for the virus that causes COVID-19, since you completed the second survey on [cv19a1_date]? (Select all that apply)<br><br>Do not include the antibody/ serology test (blood test for previous infection) that is part of this reseach study. | ☐ No, I never tried to get tested<br>☐ No, I tried to get tested but was not able to<br>☐ Yes, and I am waiting for the results<br>☐ Yes, and the test showed that I did not have it ("negative" test)<br>☐ Yes, and the test showed that I did have it ("positive" test)<br>☐ Prefer not to answer |

| | |
|---|---|
| Which type of test for the virus that causes COVID-19 did you try to get but were not able to? | ☐ Viral Test (swab test for current infection)<br>☐ Antibody/ Serology Test (blood test for previous infection)<br>☐ Saliva or Mouth Rinse Test<br>☐ Rapid-Antigen Test<br>☐ Other Specify<br>☐ Don't Know<br>☐ Prefer not to answer<br>(Antibody/ Serology testing and saliva/ mouth rinse testing may not be currently available in all areas, but it may be available in the future.) |
| Specify the other type of test for the virus that causes COVID-19 that you tried to get but were not able to. | ______________________<br>(Enter "555" if you would prefer not to answer.) |
| Please specify why you got tested or tried to get tested | ☐ A contact or possible contact of mine tested positive for COVID-19<br>☐ I had COVID-19 symptoms<br>☐ I was admitted to hospital (unrelated to COVID-19)<br>☐ I was admitted to hospital (COVID-19 related)<br>☐ I was tested in another facility or institution context (contact with someone in long term care, prison etc)<br>☐ I was participating in another facility research study<br>☐ Travel-related testing<br>☐ Routine workplace testing (health care workplace)<br>☐ Routine workplace testing (non-health care workplace)<br>☐ Other, specify<br>☐ Don't Know<br>☐ Prefer not to answer |
| Please specify the other reason(s) why you got tested or tried to get tested | ______________________<br>(Enter "555" if you would prefer not to answer.) |
| Please specify why you didn't try to get tested | ☐ Symptoms were due to another condition, specify<br>☐ Symptoms were mild<br>☐ A healthcare provider did not recommend testing<br>☐ Other, specify<br>☐ Prefer not to answer |
| Please specify the condition that you believe the symptoms are from | ______________________<br>(Enter "555" if you would prefer not to answer.) |
| Please specify the other reason(s) you didn't try to get tested | ______________________<br>(Enter "555" if you would prefer not to answer.) |
| IN THE PAST TWO WEEKS, how many times have you been tested for COVID-19? | ◯ 0<br>◯ 1<br>◯ 2<br>◯ 3<br>◯ 4<br>◯ Prefer not to answer |
| Details of First COVID-19 Test | |

| | |
|---|---|
| What type of test did you receive? | ◯ Viral Test (swab test for current infection)<br>◯ Antibody/ Serology Test (blood test for previous infection)<br>◯ Saliva or Mouth Rinse Test<br>◯ Rapid-Antigen Test<br>◯ Other Specify<br>◯ Don't Know<br>◯ Prefer not to answer<br>(Do not include the antibody/ serology test (blood test for previous infection) that is part of this reseach study ) |
| Specify other type of test you received. | __________________________________<br>(Enter "555" if you would prefer not to answer.) |
| Date of test | __________________________________<br>(If you do not recall the exact date, please provide an estimate. Enter "01-01-1900" if you would prefer not to answer.) |
| First test result | ◯ Negative<br>◯ Positive<br>◯ Don't know<br>◯ Prefer not to answer |
| Details of Second COVID-19 Test | |
| What type of test did you receive? | ◯ Viral Test (swab test for current infection)<br>◯ Antibody/ Serology Test (blood test for previous infection)<br>◯ Saliva or Mouth Rinse Test<br>◯ Rapid-Antigen Test<br>◯ Other Specify<br>◯ Don't Know<br>◯ Prefer not to answer<br>(Do not include the antibody/ serology test (blood test for previous infection) that is part of this reseach study ) |
| Specify other type of test you received. | __________________________________<br>(Enter "555" if you would prefer not to answer.) |
| Date of test | __________________________________<br>(If you do not recall the exact date, please provide an estimate. Enter "01-01-1900" if you would prefer not to answer.) |
| Second test result | ◯ Negative<br>◯ Positive<br>◯ Don't know<br>◯ Prefer not to answer |
| Details of Third COVID-19 Test | |

| | |
|---|---|
| What type of test did you receive? | ◯ Viral Test (swab test for current infection)<br>◯ Antibody/ Serology Test (blood test for previous infection)<br>◯ Saliva or Mouth Rinse Test<br>◯ Rapid-Antigen Test<br>◯ Other Specify<br>◯ Don't Know<br>◯ Prefer not to answer<br>(Do not include the antibody/ serology test (blood test for previous infection) that is part of this reseach study ) |
| Specify other type of test you received. | __________________________________<br>(Enter "555" if you would prefer not to answer.) |
| Date of test | __________________________________<br>(If you do not recall the exact date, please provide an estimate. Enter "01-01-1900" if you would prefer not to answer.) |
| Third test result | ◯ Negative<br>◯ Positive<br>◯ Don't know<br>◯ Prefer not to answer |
| Details of Fourth COVID-19 Test | |
| What type of test did you receive? | ◯ Viral Test (swab test for current infection)<br>◯ Antibody/ Serology Test (blood test for previous infection)<br>◯ Saliva or Mouth Rinse Test<br>◯ Rapid-Antigen Test<br>◯ Other Specify<br>◯ Don't Know<br>◯ Prefer not to answer<br>(Do not include the antibody/ serology test (blood test for previous infection) that is part of this reseach study ) |
| Specify other type of test you received. | __________________________________<br>(Enter "555" if you would prefer not to answer.) |
| Date of test | __________________________________<br>(If you do not recall the exact date, please provide an estimate. Enter "01-01-1900" if you would prefer not to answer.) |
| Fourth test result | ◯ Negative<br>◯ Positive<br>◯ Don't know<br>◯ Prefer not to answer |

| | |
|---|---|
| Have you had any colds, since you completed the second survey on [cv19a1_date]? | ◯ Yes<br>◯ No<br>◯ Don't Know<br>◯ Prefer not to answer |
| Have you had any of the following symptoms, since you completed the second survey on [cv19a1_date]? (Select all that apply) | ☐ New or worsening cough<br>☐ Shortness of breath<br>☐ Difficulty breathing<br>☐ Fever<br>☐ Chills<br>☐ Muscle or body aches<br>☐ Headache<br>☐ Sore throat<br>☐ New loss of sense of smell or taste<br>☐ Runny nose or nasal congestion*<br>☐ Nausea or vomiting<br>☐ Diarrhea<br>☐ Fatigue<br>☐ Skin Rash<br>☐ Conjunctivitis (pink eye)<br>☐ Other, Specify<br>☐ None of the above<br>☐ Prefer not to answer<br>(*Runny nose or nasal congestion in the absence of underlying reason for these symptoms such as seasonal allergies, post nasal drip, etc) |
| Specify the other symptoms that you had | ________________________________<br>(Enter "555" if you would prefer not to answer.) |
| Do you remember the date when you had your first symptom? | ◯ Yes<br>◯ No<br>◯ Roughly<br>◯ Prefer not to answer |
| What was the date of your first symptom? | ________________________________<br>(If you do not recall the exact date, please provide an estimate. Enter "01-01-1900" if you would prefer not to answer.) |
| In the TWO WEEKS, before you had symptoms, did you: (Select all that apply) | ☐ Have contact with someone who tested positive for COVID-19<br>☐ Have contact with someone who likely had COVID-19 (e.g., was not tested but had symptoms; was told by a healthcare provider that he/she likely had it)<br>☐ Travel outside the province<br>☐ None of the above<br>☐ Prefer not to answer |
| How many places did you travel? | ◯ 0<br>◯ 1<br>◯ 2<br>◯ 3<br>◯ 4<br>◯ Prefer not to answer<br>(Travel outside the province) |
| Place 1 | |

Location
(e.g. Seattle, Washington OR Toronto, Ontario)

__________________________________
(Enter "555" if you would prefer not to answer.)

Start date of travel

__________________________________
(If you do not recall the exact date, please provide an estimate. Enter "01-01-1900" if you would prefer not to answer.)

End date of travel

__________________________________
(If you do not recall the exact date, please provide an estimate. Enter "01-01-1900" if you would prefer not to answer.)

How? (Check all that apply)

- ☐ Car
- ☐ Plane
- ☐ Train
- ☐ Ship
- ☐ Other
- ☐ Prefer not to answer

Please specify other mode of transportation

__________________________________
(Enter "555" if you would prefer not to answer.)

Place 2

Location
(e.g. Portland, Oregon OR Vancouver, BC)

__________________________________
(Enter "555" if you would prefer not to answer.)

Start date of travel

__________________________________
(If you do not recall the exact date, please provide an estimate. Enter "01-01-1900" if you would prefer not to answer.)

End date of travel

__________________________________
(If you do not recall the exact date, please provide an estimate. Enter "01-01-1900" if you would prefer not to answer.)

How? (Check all that apply)

- ☐ Car
- ☐ Plane
- ☐ Train
- ☐ Ship
- ☐ Other
- ☐ Prefer not to answer

Please specify other mode of transportation

__________________________________
(Enter "555" if you would prefer not to answer.)

Place 3

| | |
|---|---|
| Location<br>(e.g. Paris, France OR Banff, Alberta) | ______________________________<br>(Enter "555" if you would prefer not to answer.) |
| Start date of travel | ______________________________<br>(If you do not recall the exact date, please provide an estimate. Enter "01-01-1900" if you would prefer not to answer.) |
| End date of travel | ______________________________<br>(If you do not recall the exact date, please provide an estimate. Enter "01-01-1900" if you would prefer not to answer.) |
| How? (Check all that apply) | ☐ Car<br>☐ Plane<br>☐ Train<br>☐ Ship<br>☐ Other<br>☐ Prefer not to answer |
| Please specify other mode of transportation | ______________________________<br>(Enter "555" if you would prefer not to answer.) |
| Place 4 | |
| Location<br>(e.g. Portland, Oregon OR Vancouver, BC) | ______________________________<br>(Enter "555" if you would prefer not to answer.) |
| Start date of travel | ______________________________<br>(If you do not recall the exact date, please provide an estimate. Enter "01-01-1900" if you would prefer not to answer.) |
| End date of travel | ______________________________<br>(If you do not recall the exact date, please provide an estimate. Enter "01-01-1900" if you would prefer not to answer.) |
| How? (Check all that apply) | ☐ Car<br>☐ Plane<br>☐ Train<br>☐ Ship<br>☐ Other<br>☐ Prefer not to answer |
| Please specify other mode of transportation | ______________________________<br>(Enter "555" if you would prefer not to answer.) |

| | |
|---|---|
| Which of the following occurred as a result of your symptoms? (Select all that apply) | ☐ You spoke to a healthcare provider over the phone, by email, or online<br>☐ You saw a healthcare provider in person, such as in a clinic or doctor's office<br>☐ You saw a healthcare provider in person, such as in urgent care, or in an Emergency Department (ED)<br>☐ You were kept overnight in a hospital because a healthcare provider thought you had COVID-19<br>☐ You were admitted to a hospital's ICU (Intensive Care Unit)<br>☐ You self-isolated or quarantined at home<br>☐ None of the above<br>☐ Prefer not to answer |
| Has anyone in your family been diagnosed with COVID-19, since you completed the second survey on [cv19a1_date]? (Select all that apply) | ☐ Yes, member of household<br>☐ Yes, non-household member<br>☐ No<br>☐ Don't know<br>☐ Prefer not to answer |
| Have any of the following happened to your family members because of COVID-19, since you completed the second survey on [cv19a1_date]? (Select all that apply) | ☐ Fallen physically ill<br>☐ Hospitalized<br>☐ Self-isolated or quarantined with symptoms<br>☐ Self-isolated or quarantined without symptoms (e.g., due to possible exposure)<br>☐ Lost job or been laid off from job<br>☐ Reduced ability to earn money<br>☐ Passed away<br>☐ None of the above<br>☐ Prefer not to answer<br>(Include any family members, not just those living in the same household as you) |
| Did you get the flu shot, since you completed the second survey on [cv19a1_date]? | ◯ Yes<br>◯ No<br>◯ Don't Know<br>◯ Prefer not to answer |
| Have you received at least one dose of a COVID-19 vaccine? | ◯ Yes<br>◯ No<br>◯ Prefer not to answer<br>(Answer 'Yes' if you have received at least one dose of the COVID-19 vaccine. Note: Certain types of vaccines require more than one dose to protect against COVID-19. You would have been informed at the time of vaccination if you needed a second dose. Some groups of people are also now eligible to receive a third booster shot.) |
| Did you feel any side effects from the COVID-19 vaccine? | ◯ Yes<br>◯ No<br>◯ Prefer not to answer |

| | |
|---|---|
| If yes, check all that apply. | ☐ Head ache<br>☐ Tiredness<br>☐ Fever or chills<br>☐ Muscle or joint pain<br>☐ Soreness where the vaccine was injected<br>☐ Nausea or vomiting<br>☐ Other<br>☐ Prefer not to answer |
| Please specify other side effects | ______________________<br>(Enter "555" if you would prefer not to answer.) |
| How many doses of the COVID-19 vaccine have you received so far? | ◯ One dose<br>◯ Two doses<br>◯ Three doses<br>◯ Four doses<br>◯ More than four doses<br>◯ Prefer not to answer |
| When did you receive your first dose of the COVID-19 vaccine? | ______________________<br>(If you do not recall the exact date, please provide an estimate. Enter "01-01-1900" if you would prefer not to answer.) |
| Which vaccine did you receive? (first dose) | ◯ Pfizer and BioNTech mRNA vaccine<br>◯ Moderna mRNA vaccine<br>◯ AstraZeneca Oxford vaccine<br>◯ Janssen (Johnson & Johnson) vaccine<br>◯ Other, specify<br>◯ Don't Know<br>◯ Prefer not to answer |
| Specify the other vaccine | ______________________<br>(Enter "555" if you would prefer not to answer.) |
| When did you receive your second dose of the COVID-19 vaccine? | ______________________<br>(If you do not recall the exact date, please provide an estimate. Enter "01-01-1900" if you would prefer not to answer.) |
| Which vaccine did you receive? (second dose) | ◯ Pfizer and BioNTech mRNA vaccine<br>◯ Moderna mRNA vaccine<br>◯ AstraZeneca Oxford vaccine<br>◯ Janssen (Johnson & Johnson) vaccine<br>◯ Other, specify<br>◯ Don't Know<br>◯ Prefer not to answer |
| Specify the other vaccine | ______________________<br>(Enter "555" if you would prefer not to answer.) |

---

When did you receive your third dose of the COVID-19 vaccine?

__________________________________
(If you do not recall the exact date, please provide an estimate. Enter "01-01-1900" if you would prefer not to answer.)

---

Which vaccine did you receive? (third dose)

- ◯ Pfizer and BioNTech mRNA vaccine
- ◯ Moderna mRNA vaccine
- ◯ AstraZeneca Oxford vaccine
- ◯ Janssen (Johnson & Johnson) vaccine
- ◯ Other, specify
- ◯ Don't Know
- ◯ Prefer not to answer

---

Specify the other vaccine

__________________________________
(Enter "555" if you would prefer not to answer.)

---

When did you receive your fourth dose of the COVID-19 vaccine?

__________________________________
(If you do not recall the exact date, please provide an estimate. Enter "01-01-1900" if you would prefer not to answer.)

---

Which vaccine did you receive? (fourth dose)

- ◯ Pfizer and BioNTech mRNA vaccine
- ◯ Moderna mRNA vaccine
- ◯ AstraZeneca Oxford vaccine
- ◯ Janssen (Johnson & Johnson) vaccine
- ◯ Other, specify
- ◯ Don't Know
- ◯ Prefer not to answer

---

Specify the other vaccine

__________________________________
(Enter "555" if you would prefer not to answer.)

---

When a COVID-19 vaccine becomes available, how likely is it that you will choose to get it?

- ◯ Very Likely
- ◯ Somewhat Likely
- ◯ Somewhat Unlikely
- ◯ Very Unlikely
- ◯ Don't Know
- ◯ Prefer not to answer

---

What are some of the reasons why you don't want to get the COVID-19 vaccine?

- ☐ Not confident in the safety of the vaccine
- ☐ Concern about risks and side effects
- ☐ Will wait until it seems safe to get the vaccine
- ☐ Do not consider it necessary to get the vaccine
- ☐ Do not believe in vaccination
- ☐ Have not yet decided
- ☐ Have a pre-existing medical condition
- ☐ Already had or think I have had COVID-19
- ☐ Other, Specify
- ☐ Prefer not to answer

(Check all that apply.)

---

What were the other reasons you didn't want to get the COVID-19 vaccine?

__________________________________
(Enter "555" if you would prefer not to answer.)

| | Strongly Agree | Agree | Disagree | Strongly Disagree | Prefer not to answer |
|---|---|---|---|---|---|
| Having myself vaccinated is important for the health of others in my community. | ◯ | ◯ | ◯ | ◯ | ◯ |
| Getting vaccines is a good way to protect myself from disease. | ◯ | ◯ | ◯ | ◯ | ◯ |
| Generally I do what my doctor or health care provider recommends about vaccines for myself | ◯ | ◯ | ◯ | ◯ | ◯ |
| I am concerned about serious adverse effects of vaccines. | ◯ | ◯ | ◯ | ◯ | ◯ |

| | Never | Rarely | Occasionally | Often | Always | Don't Know | Prefer not to answer |
|---|---|---|---|---|---|---|---|
| How often have you worn a mask in public places since you completed your second survey on [cv19a1_date]? | ◯ | ◯ | ◯ | ◯ | ◯ | ◯ | ◯ |
| How often have you practiced physical distancing in public places since you completed your second survey on [cv19a1_date]? | ◯ | ◯ | ◯ | ◯ | ◯ | ◯ | ◯ |
| How often have you avoided crowded places/ gatherings since you completed your second survey on [cv19a1_date]? | ◯ | ◯ | ◯ | ◯ | ◯ | ◯ | ◯ |
| How often have you avoided common greetings (e.g handshakes, hugs etc) since you completed your second survey on [cv19a1_date]? | ◯ | ◯ | ◯ | ◯ | ◯ | ◯ | ◯ |
| How often have you limited contact with people at higher risk (e.g. elderly relative) since you completed your second survey on [cv19a1_date]? | ◯ | ◯ | ◯ | ◯ | ◯ | ◯ | ◯ |
| How often have you self-Isolated because someone thought you were infected with the virus that causes COVID-19 since you completed your second survey on [cv19a1_date]? | ◯ | ◯ | ◯ | ◯ | ◯ | ◯ | ◯ |

| | | | | | | | |
|---|---|---|---|---|---|---|---|
| How often have you quarantined because you may have been exposed to the virus that causes COVID-19, but did not show symptoms since you completed your second survey on [cv19a1_date]? | ◯ | ◯ | ◯ | ◯ | ◯ | ◯ | ◯ |

Have you participated in any organized sports or recreational activities, in person, since you completed the second survey on [cv19a1_date]?

◯ Yes
◯ No
◯ Prefer not to answer

If yes, how many people were in the activity?

◯ 0-9
◯ 10-19
◯ 20-29
◯ 30+
◯ Prefer not to answer

How many times have you been in a gathering of >10 or more people (excluding school or work), since you completed the second survey on [cv19a1_date]?

__________________________________
(A gathering in this context is defined as any assembly or meeting of more than 10 persons, in open air or in a building or premises. These can include gatherings with or without masks or social distancing. Gatherings include a range of interpersonal gatherings/events of varying purposes, from gatherings/events of family and friends (e.g., sports, weddings, funerals, baptisms, birthday parties and cultural ceremonies) to community gathering spaces (e.g., places of worship, places of traditional gatherings, libraries, community and recreation centres, camps, drop-in centres, food banks and communal kitchens, museums, theatres, cinemas, and tourist attractions) to larger planned or structured gatherings/events, including mass gatherings (e.g., large meetings or conferences, national sporting events, large cultural/religious events, festivals). Enter "555" if you would prefer not to answer.)

**The following are a number of mental health services that you may have accessed. Please check the relevant boxes for each service indicating if you have accessed the service since you completed the second survey on [cv19a1_date] or if you would be interested in accessing the service.**

| | Have accessed this service since completing the second survey on [cv19a1_date] | Would be interested in accessing this service | Not accessed, not interested | Prefer not to answer |
|---|---|---|---|---|

| | | | | |
|---|---|---|---|---|
| Individual therapy (you alone) with a professional (in person or virtual) | ○ | ○ | ○ | ○ |
| Group therapy (you and others) with a professional (in person or virtual) | ○ | ○ | ○ | ○ |
| Couples therapy (you and your partner) with a professional (in person or virtual) | ○ | ○ | ○ | ○ |
| Family therapy (you and your family members) with a professional (in person or virtual) | ○ | ○ | ○ | ○ |
| Mental health mobile app (general) | ○ | ○ | ○ | ○ |
| Mental health mobile app to interact with a therapist | ○ | ○ | ○ | ○ |
| Phone helpline for mental health support (e.g. Crisis Services Canada) | ○ | ○ | ○ | ○ |
| Online mental health information you looked up yourself | ○ | ○ | ○ | ○ |

**Social Support**

**We would like to measure your feelings about how supported you feel. Choose 1 of the 4 responses to show whether you agree or disagree with the statement.**

| | Strongly Agree | Agree | Disagree | Strongly Disagree | Prefer not to answer |
|---|---|---|---|---|---|
| I have family and friends who help me feel safe, secure and happy | ○ | ○ | ○ | ○ | ○ |
| There is someone I trust whom I would turn to for advice if I were having problems | ○ | ○ | ○ | ○ | ○ |
| There are people I can count on in an emergency | ○ | ○ | ○ | ○ | ○ |
| I feel part of a group of people who share my attitudes and beliefs | ○ | ○ | ○ | ○ | ○ |

#### Worrying (PAST TWO WEEKS)

| | Not at all worried | Slightly worried | Moderately worried | Very worried | Extremely worried | Prefer not to answer |
|---|---|---|---|---|---|---|
| During the PAST TWO WEEKS, how worried have you been about being infected by the virus that causes COVID-19? | ◯ | ◯ | ◯ | ◯ | ◯ | ◯ |
| During the PAST TWO WEEKS, how worried have you been about friends or family being infected by the virus that causes COVID-19? | ◯ | ◯ | ◯ | ◯ | ◯ | ◯ |
| During the PAST TWO WEEKS, how worried have you been about your physical health being influenced by COVID-19? | ◯ | ◯ | ◯ | ◯ | ◯ | ◯ |
| During the PAST TWO WEEKS, how worried have you been about your mental/emotional health being influenced by COVID-19? | ◯ | ◯ | ◯ | ◯ | ◯ | ◯ |
| During the PAST TWO WEEKS, how worried were you generally? | ◯ | ◯ | ◯ | ◯ | ◯ | ◯ |

How much are you reading or talking about COVID-19, since you completed the second survey on [cv19a1_date]?

◯ Never
◯ A few times a month
◯ Weekly
◯ A few times a week
◯ Daily
◯ Prefer not to answer

Has the COVID-19 pandemic in your area led to any POSITIVE changes in your life, since you completed the second survey on [cv19a1_date]?

☐ Yes
☐ No
☐ I don't know
☐ Prefer not to answer

Please specify these POSITIVE changes in your life (Select all that apply)

- ☐ More time with family
- ☐ More time with friends
- ☐ More exercising
- ☐ Healthier eating
- ☐ New hobbies
- ☐ Got a new pet
- ☐ Learned new technology
- ☐ Made new friends online
- ☐ Saved money
- ☐ Less commuting
- ☐ Better hand hygiene
- ☐ Less busy / more downtime
- ☐ Working from home
- ☐ Schooling from home
- ☐ More time outdoors
- ☐ Not getting sick as often
- ☐ Avoiding bullying
- ☐ Home improvements
- ☐ Improved mental health
- ☐ Other, specify
- ☐ Prefer not to answer

Please specify these other POSITIVE changes in your life

______________________________
(Enter "555" if you would prefer not to answer.)

**Families have different routines.**

**We would like to know about your family's routines. Choose 1 of the 4 responses that best fits the statement with your family's routine.**

**For those with shared custody of your children, please only think of your household's routines.**

| | Always | Often | Rarely | Never | Prefer not to answer |
|---|---|---|---|---|---|
| Parents have some times each day for just talking with children | ○ | ○ | ○ | ○ | ○ |
| Parents and children play together each day | ○ | ○ | ○ | ○ | ○ |
| The whole family eats dinner together almost every night | ○ | ○ | ○ | ○ | ○ |
| Family checks in or out with each other when someone leaves the home | ○ | ○ | ○ | ○ | ○ |
| Children do regular household chores | ○ | ○ | ○ | ○ | ○ |
| Parents read with children together almost every day | ○ | ○ | ○ | ○ | ○ |

| | | | | | |
|---|---|---|---|---|---|
| Children go to bed at the same time each night during the week | ◯ | ◯ | ◯ | ◯ | ◯ |

#### LIFE CHANGES DUE TO THE COVID-19 PANDEMIC

| | |
|---|---|
| Based on what you know at this time, how safe do you think it is for students in your community 12 years of age and OLDER to attend school in-person? | ◯ Very safe<br>◯ Somewhat safe<br>◯ Not very safe<br>◯ Not safe at all<br>◯ Don't Know<br>◯ Prefer not to answer |
| Based on what you know at this time, how safe do you think it is for students in your community YOUNGER than 12 years of age to attend school in-person? | ◯ Very safe<br>◯ Somewhat safe<br>◯ Not very safe<br>◯ Not safe at all<br>◯ Don't Know<br>◯ Prefer not to answer |
| Are you enrolled in school for the current academic year*? | ◯ Not in school<br>◯ Home school<br>◯ Elementary school<br>◯ Junior High or Middle School<br>◯ Secondary or High School<br>◯ College<br>◯ University<br>◯ Prefer not to answer<br>(Current academic year = Sept 2021 to June 2022) |
| Has your school building closed since you completed your second survey on [cv19a1_date]? | ◯ Yes<br>◯ No<br>◯ N/A<br>◯ Prefer not to answer |
| If no, are you attending classes in person? | ◯ Yes<br>◯ No<br>◯ N/A<br>◯ Prefer not to answer |
| If yes, how many times did the school close? | ____________________________<br>(Enter "666" if you don't know the answer.Enter "555" if you would prefer not to answer.) |
| If yes, when did the first school building closure begin? | ____________________________<br>(If you do not recall the exact date, please provide an estimate. Enter "01-01-1900" if you would prefer not to answer.) |
| If yes, were classes resumed online? | ◯ Yes<br>◯ No<br>◯ N/A<br>◯ Prefer not to answer |

| Question | Response |
| --- | --- |
| How many weeks in total was the school building closed since September 2021 due to COVID-19? | __________________________________ ((do not include Christmas and spring break) Enter "555" if you would prefer not to answer.) |
| Did you participate in classes online since September 2021? | ◯ Yes<br>◯ No<br>◯ Partially<br>◯ N/A<br>◯ Prefer not to answer |
| If yes, did you have easy access to the internet and a computer? | ◯ Yes<br>◯ No<br>◯ N/A<br>◯ Prefer not to answer |
| If yes, were there assignments for you to complete? | ◯ Yes<br>◯ No<br>◯ N/A<br>◯ Prefer not to answer |
| Did you complete the assignments? | ◯ Yes<br>◯ No<br>◯ Partially<br>◯ N/A<br>◯ Prefer not to answer |
| Did you physically return to school since the first closure? | ◯ Yes<br>◯ No<br>◯ N/A<br>◯ Prefer not to answer |
| When did you return to school? | __________________________________ (If you do not recall the exact date, please provide an estimate. Enter "01-01-1900" if you would prefer not to answer.) |
| How many days of in-school or remote/online learning have you missed/ been absent for since you completed your second survey on [cv19a1_date]? | __________________________________ (Enter "555" if you would prefer not to answer.) |
| Are you currently working? | ☐ Working for pay<br>☐ On paid leave<br>☐ On unpaid leave<br>☐ Laid off or lost job<br>☐ Unemployed and looking for a job<br>☐ Retired<br>☐ Staying at home, homemaker or unpaid caregiver to someone inside or outside of the home<br>☐ Other<br>☐ Prefer not to answer |

| | |
|---|---|
| In what ways has the COVID-19 outbreak affected your work, since you completed the second survey on [cv19a1_date]? (Select all that apply) | ☐ I moved to working remotely or from home<br>☐ I lost my job permanently<br>☐ I lost my job temporarily, or was not told for how long<br>☐ I got a new job<br>☐ I reduced my work hours<br>☐ I increased my work hours<br>☐ My job put me at increased risk of getting COVID-19<br>☐ I laid off employees<br>☐ I did not have a paying job before the COVID-19 outbreak<br>☐ None of these apply<br>☐ Prefer not to answer |
| In what ways has the COVID-19 outbreak affected your spouse/partners work, since you completed the second survey on [cv19a1_date]? (Select all that apply) | ☐ Not applicable I do not have a spouse/partner<br>☐ My spouse/partner moved to working remotely or from home<br>☐ My spouse/partner lost his/her job permanently<br>☐ My spouse/partner lost his/her job temporarily, or was not told for how long<br>☐ My spouse/partner got a new job<br>☐ My spouse/partner reduced his/her work hours<br>☐ My spouse/partner increased his/her work hours<br>☐ My spouse/partners job put him/her at increased risk of getting COVID-19<br>☐ My spouse/partner laid off employees<br>☐ My spouse/partner did not have a paying job before the COVID-19 outbreak<br>☐ None of these apply<br>☐ Prefer not to answer |
| Since you completed the second survey on [cv19a1_date], has the quality of the relationships between you and members of your family changed? | ◯ A lot better<br>◯ A little better<br>◯ About the same<br>◯ A little worse<br>◯ A lot worse<br>◯ Prefer not to answer |
| Since you completed the second survey on [cv19a1_date], how stressful have these changes in relationships between you and members of your family been for you? | ◯ Not stressful<br>◯ A little bit stressful<br>◯ Stressful<br>◯ Very stressful<br>◯ Extremely stressful<br>◯ Prefer not to answer |
| Since you completed the second survey on [cv19a1_date], has the quality of your relationships with your friends changed? | ◯ A lot better<br>◯ A little better<br>◯ About the same<br>◯ A little worse<br>◯ A lot worse<br>◯ Prefer not to answer |
| Since you completed the second survey on [cv19a1_date], how stressful have these changes in relationships between you and your friends been for you? | ◯ Not stressful<br>◯ A little bit stressful<br>◯ Stressful<br>◯ Very stressful<br>◯ Extremely stressful<br>◯ Prefer not to answer |

| | |
|---|---|
| Since you completed the second survey on [cv19a1_date], how difficult has the cancellation of important events in your life (such as weddings, vacations, etc.) been for you? | ◯ Not difficult<br>◯ A little bit difficult<br>◯ Difficult<br>◯ Very difficult<br>◯ Extremely difficult<br>◯ There hasn't been any cancellation of important events<br>◯ Prefer not to answer |
| During the PAST TWO WEEKS, how many people (from outside of your household) have you had an in-person conversation with in an indoor environment?<br><br>This can include things like work, school or other activities with or without masks and/or social distancing. If you aren't sure, please estimate a number. | ______________________________<br>(In-person means that the person is in the same place as you (NOT on the phone or video chat) Enter "555" if you would prefer not to answer.) |
| During the PAST TWO WEEKS, how many people (from outside of your household) have you had an in-person conversation with in an outdoor environment?<br><br>This can include things like work, school or other activities with or without masks and/or social distancing. If you aren't sure, please estimate a number | ______________________________<br>(In-person means that the person is in the same place as you (NOT on the phone or video chat) Enter "555" if you would prefer not to answer.) |
| During the PAST TWO WEEKS, how many times did you go out outside of your home (e.g., going to stores, parks, etc.)? | ◯ Not at all<br>◯ 1-2 days per week or less<br>◯ 3-4 days per week<br>◯ 5-6 days per week<br>◯ Daily<br>◯ Prefer not to answer |
| During the PAST TWO WEEKS, how stressful have the restrictions on leaving home been for you? | ◯ Not stressful<br>◯ A little bit stressful<br>◯ Stressful<br>◯ Very stressful<br>◯ Extremely stressful<br>◯ There are no restrictions on leaving home<br>◯ Prefer not to answer |
| During the PAST TWO WEEKS, how difficult has it been for you to follow the recommendations for keeping away from close contact with people who you don't live with? | ◯ Not difficult<br>◯ A little bit difficult<br>◯ Difficult<br>◯ Very difficult<br>◯ Extremely difficult<br>◯ There are no recommendations for keeping away from close contact with people who I don't live with<br>◯ Prefer not to answer |
| During the PAST TWO WEEKS, to what degree have changes related to the COVID-19 pandemic in your area created financial problems for your family? | ◯ No financial problems<br>◯ A few financial problems<br>◯ Some financial problems<br>◯ A lot of financial problems<br>◯ Extreme financial problems<br>◯ Prefer not to answer |

During the PAST TWO WEEKS, to what degree are you concerned about the stability of your living situation?

◯ Not at all concerned
◯ A little concerned
◯ Concerned
◯ Very concerned
◯ Extremely concerned
◯ Prefer not to answer
(Stability of your living situation refers to forced moves that are the result of poor housing quality, unstable neighbourhood conditions and high costs of housing in relation to income. )

During the PAST TWO WEEKS, did you worry your food would run out because of a lack of money?

◯ Yes
◯ No
◯ Prefer not to answer

How hopeful are you that the COVID-19 pandemic in your area will end soon?

◯ Extremely hopeful
◯ Very hopeful
◯ Hopeful
◯ A little bit hopeful
◯ Not at all hopeful
◯ There is no COVID-19 pandemic in my area
◯ Prefer not to answer

**Compared to before the COVID-19 outbreak, how much are you now:**

| | Less | Same amount | More | Prefer not to answer |
|---|---|---|---|---|
| Eating | ◯ | ◯ | ◯ | ◯ |
| Sleeping | ◯ | ◯ | ◯ | ◯ |
| Physically active | ◯ | ◯ | ◯ | ◯ |
| Spending time outside | ◯ | ◯ | ◯ | ◯ |
| Spending time with friends in-person | ◯ | ◯ | ◯ | ◯ |
| Spending time with friends remotely (e.g., online, social media, texting) | ◯ | ◯ | ◯ | ◯ |
| Spending time watching TV, playing video/computer games, or using social media for educational or work purposes, including school work | ◯ | ◯ | ◯ | ◯ |
| Spending time watching TV, playing video/computer games, or using social media for NON-educational or NON-work purposes | ◯ | ◯ | ◯ | ◯ |

#### DAILY BEHAVIORS (Past TWO WEEKS)

During the PAST TWO WEEKS, what time did you go to bed on WEEKDAYS (on average)?

- ◯ Before 8 pm
- ◯ 8 pm - 10 pm
- ◯ 10 pm - 12 am (Midnight)
- ◯ After 12 am (Midnight)
- ◯ I slept during daytime hours (example: shift or on-call work)
- ◯ Prefer not to answer

During the PAST TWO WEEKS, what time did you go to bed on WEEKENDS (on average)?

- ◯ Before 8 pm
- ◯ 8 pm - 10 pm
- ◯ 10 pm - 12 am (Midnight)
- ◯ After 12 am (Midnight)
- ◯ I slept during daytime hours (example: shift or on-call work)
- ◯ Prefer not to answer

During the PAST TWO WEEKS, how many hours per night did you sleep on WEEKDAYS (on average)?

- ◯ Less than 6 hours
- ◯ 6-8 hours
- ◯ 8-10 hours
- ◯ Greater than 10 hours
- ◯ Prefer not to answer

During the PAST TWO WEEKS, how many hours per night did you sleep on WEEKENDS (on average)?

- ◯ Less than 6 hours
- ◯ 6-8 hours
- ◯ 8-10 hours
- ◯ Greater than 10 hours
- ◯ Prefer not to answer

During the PAST TWO WEEKS, how many days per week did you exercise (e.g., increased heart rate, breathing) for at least 30 minutes over the entire day?

- ◯ Not at all
- ◯ 1-2 days per week or less
- ◯ 3-4 days per week
- ◯ 5-6 days per week
- ◯ Daily
- ◯ Prefer not to answer

During the PAST TWO WEEKS, how many days per week did you spend time outdoors?

- ◯ Not at all
- ◯ 1-2 days per week or less
- ◯ 3-4 days per week
- ◯ 5-6 days per week
- ◯ Daily
- ◯ Prefer not to answer

How has the COVID-19 outbreak affected your regular childcare, since you completed the second survey on [cv19a1_date]? (Select all that apply)

- ☐ I had difficulty arranging for childcare
- ☐ I had to pay more for childcare
- ☐ My spouse/partner or I had to change our work schedule to care for our children ourselves
- ☐ My spouse/partner or I had to reduce our paid work hours to care for our children ourselves
- ☐ My regular childcare has not been affected by the COVID-19 outbreak
- ☐ I do not have a child in childcare
- ☐ Prefer not to answer

#### EMOTIONS/WORRIES (PAST TWO WEEKS)

| | |
|---|---|
| During the PAST TWO WEEKS, how happy versus sad were you? | ◯ Very happy/cheerful<br>◯ Moderately happy/cheerful<br>◯ Neutral<br>◯ Moderately sad/depressed/unhappy<br>◯ Very sad/depressed/unhappy<br>◯ Prefer not to answer |
| During the PAST TWO WEEKS, how relaxed versus anxious were you? | ◯ Very relaxed/calm<br>◯ Moderately relaxed/calm<br>◯ Neutral<br>◯ Moderately nervous/anxious<br>◯ Very nervous/anxious<br>◯ Prefer not to answer |
| During the PAST TWO WEEKS, how fidgety or restless were you? | ◯ Not fidgety/restless at all<br>◯ Slightly fidgety/restless<br>◯ Moderately fidgety/restless<br>◯ Very fidgety/restless<br>◯ Extremely fidgety/restless<br>◯ Prefer not to answer |
| During the PAST TWO WEEKS, how tired were you? | ◯ Not tired at all<br>◯ Slightly tired<br>◯ Moderately tired<br>◯ Very tired<br>◯ Extremely tired<br>◯ Prefer not to answer |
| During the PAST TWO WEEKS, how well were you able to concentrate or focus? | ◯ Very focused/attentive<br>◯ Moderately focused/attentive<br>◯ Neutral<br>◯ Moderately unfocused/distracted<br>◯ Very unfocused/distracted<br>◯ Prefer not to answer |
| During the PAST TWO WEEKS, how irritable or easily angered were you? | ◯ Not irritable or easily angered at all<br>◯ Slightly irritable or easily angered<br>◯ Moderately irritable or easily angered<br>◯ Very irritable or easily angered<br>◯ Extremely irritable or easily angered<br>◯ Prefer not to answer |
| During the PAST TWO WEEKS, how lonely were you? | ◯ Not lonely at all<br>◯ Slightly lonely<br>◯ Moderately lonely<br>◯ Very lonely<br>◯ Extremely lonely<br>◯ Prefer not to answer |

| | |
|---|---|
| What have been your greatest sources of stress from the COVID-19 outbreak, since you completed the second survey on [cv19a1_date]? (Select all that apply) | ☐ Health concerns<br>☐ Financial concerns<br>☐ Impact on work<br>☐ Impact on your child<br>☐ Impact on your community<br>☐ Impact on family members<br>☐ Impact on the economy<br>☐ Access to food<br>☐ Access to baby supplies (e.g., formula, diapers, wipes)<br>☐ Access to personal care products or household supplies<br>☐ Access to medical care, including mental health care<br>☐ Social distancing or being quarantined<br>☐ Other<br>☐ I am not stressed about the COVID-19 outbreak<br>☐ Prefer not to answer |
| What have you done to cope with your stress related to the COVID-19 outbreak, since you completed the second survey on [cv19a1_date]? (Select all that apply) | ☐ Meditation and/or mindfulness practices<br>☐ Talking with friends and family (e.g., by phone, text, or video)<br>☐ Engaging in more family activities (e.g., games, sports)<br>☐ Increased television watching or other screen time activities (e.g., video games, social media)<br>☐ Eating more often, including snacking<br>☐ Increasing time reading books, or doing activities like puzzles and crosswords<br>☐ Drinking alcohol<br>☐ Using tobacco (e.g., smoking, vaping)<br>☐ Using marijuana / cannabis (e.g., vaping, smoking, eating) or cannabidiol (CBD)<br>☐ Talking to my healthcare providers more frequently, including mental healthcare provider (e.g., therapist, psychologist, counselor)<br>☐ Volunteer work<br>☐ Exercise<br>☐ Other<br>☐ I have not done any of these things to cope with the stress related to the COVID-19 outbreak<br>☐ I am not stressed about the COVID-19 outbreak<br>☐ Prefer not to answer |

#### MEDIA USE (PAST TWO WEEKS)

| | |
|---|---|
| During the PAST TWO WEEKS, how much time PER WEEKDAY did you spend watching TV or digital media (e.g., Netflix, YouTube, web surfing) for educational or work purposes, including school work? | ◯ No TV or digital media<br>◯ Under 1 hour<br>◯ 1-3 hours<br>◯ 4-6 hours<br>◯ More than 6 hours<br>◯ Prefer not to answer |
| During the PAST TWO WEEKS, how much time PER WEEKEND DAY did you spend watching TV or digital media (e.g., Netflix, YouTube, web surfing) for educational or work purposes, including school work? | ◯ No TV or digital media<br>◯ Under 1 hour<br>◯ 1-3 hours<br>◯ 4-6 hours<br>◯ More than 6 hours<br>◯ Prefer not to answer |

| | |
|---|---|
| During the PAST TWO WEEKS, how much time PER WEEKDAY did you spend watching TV or digital media (e.g., Netflix, YouTube, web surfing) for NON-educational or NON-work purposes? | ◯ No TV or digital media<br>◯ Under 1 hour<br>◯ 1-3 hours<br>◯ 4-6 hours<br>◯ More than 6 hours<br>◯ Prefer not to answer |
| During the PAST TWO WEEKS, how much time PER WEEKEND DAY did you spend watching TV or digital media (e.g., Netflix, YouTube, web surfing) for NON-educational or NON-work purposes? | ◯ No TV or digital media<br>◯ Under 1 hour<br>◯ 1-3 hours<br>◯ 4-6 hours<br>◯ More than 6 hours<br>◯ Prefer not to answer |
| During the PAST TWO WEEKS, how much time PER WEEKDAY did you spend using social media (e.g.,Facebook, Instagram, Snapchat, Twitter, TikTok)? | ◯ No social media<br>◯ Under 1 hour<br>◯ 1-3 hours<br>◯ 4-6 hours<br>◯ More than 6 hours<br>◯ Prefer not to answer |
| During the PAST TWO WEEKS, how much time PER WEEKEND DAY did you spend using social media (e.g.,Facebook, Instagram, Snapchat, Twitter, TikTok)? | ◯ No social media<br>◯ Under 1 hour<br>◯ 1-3 hours<br>◯ 4-6 hours<br>◯ More than 6 hours<br>◯ Prefer not to answer |
| During the PAST TWO WEEKS, how much time PER WEEKDAY did you spend playing video games? | ◯ No video games<br>◯ Under 1 hour<br>◯ 1-3 hours<br>◯ 4-6 hours<br>◯ More than 6 hours<br>◯ Prefer not to answer |
| During the PAST TWO WEEKS, how much time PER WEEKEND DAY did you spend playing video games? | ◯ No video games<br>◯ Under 1 hour<br>◯ 1-3 hours<br>◯ 4-6 hours<br>◯ More than 6 hours<br>◯ Prefer not to answer |

#### SUBSTANCE USE (Past TWO WEEKS)

| | Not at all | Rarely | Once a month | Several times a month | Once a week | Several times a week | Once a day | More than once a day | Prefer not to answer |
|---|---|---|---|---|---|---|---|---|---|
| During the PAST TWO WEEKS, how frequently did you use alcohol? | ◯ | ◯ | ◯ | ◯ | ◯ | ◯ | ◯ | ◯ | ◯ |
| During the PAST TWO WEEKS, how frequently did you use vaping products? | ◯ | ◯ | ◯ | ◯ | ◯ | ◯ | ◯ | ◯ | ◯ |

| | | | | | | | | | |
|---|---|---|---|---|---|---|---|---|---|
| During the PAST TWO WEEKS, how frequently did you use cigarettes or other tobacco products? | ◯ | ◯ | ◯ | ◯ | ◯ | ◯ | ◯ | ◯ | ◯ |
| During the PAST TWO WEEKS, how frequently did you inhale marijuana/cannabis (e.g., joint, blunt, pipe, bong)? | ◯ | ◯ | ◯ | ◯ | ◯ | ◯ | ◯ | ◯ | ◯ |
| During the PAST TWO WEEKS, how frequently did you ingest/eat or apply marijuana/cannabis (e.g. edibles, pills, topical creams, patches, tinctures) | ◯ | ◯ | ◯ | ◯ | ◯ | ◯ | ◯ | ◯ | ◯ |
| During the PAST TWO WEEKS, how frequently did you use opiates, heroin, cocaine, crack, amphetamine, methamphetamine, hallucinogens, or ecstasy? | ◯ | ◯ | ◯ | ◯ | ◯ | ◯ | ◯ | ◯ | ◯ |

#### DEMOGRAPHIC INFORMATION

Has your living situation changed since you completed your second survey on [cv19a1_date]?

- ☐ No, my living situation has not changed
- ☐ Yes, I have moved (since the second survey)
- ☐ Yes, I regularly stay in more than one home (since the second survey)
- ☐ Yes, the number of people living in my home has changed (since the second survey)
- ☐ Prefer not to answer

Please provide the postal code of your primary home

__________________________________
(Enter "555" if you would prefer not to answer.)

How many adults currently live at your primary home (excluding yourself)?

- ◯ 0
- ◯ 1
- ◯ 2
- ◯ 3
- ◯ 4
- ◯ 5
- ◯ 6
- ◯ 7
- ◯ 8
- ◯ 9
- ◯ 10
- ◯ Prefer not to answer

How many children currently live at your primary home?

◯ 0
◯ 1
◯ 2
◯ 3
◯ 4
◯ 5
◯ 6
◯ 7
◯ 8
◯ 9
◯ 10
◯ Prefer not to answer

How many bedrooms are in your primary home?

◯ 0
◯ 1
◯ 2
◯ 3
◯ 4
◯ 5
◯ 6
◯ 7
◯ 8
◯ 9
◯ 10
◯ Prefer not to answer

How many toilets are in your primary home?

◯ 0
◯ 1
◯ 2
◯ 3
◯ 4
◯ 5
◯ 6
◯ 7
◯ 8
◯ 9
◯ 10
◯ Prefer not to answer

Choose the type of dwelling that best describes your primary home

◯ Manufactured home/ mobile home or trailer
◯ Single family detached house
◯ Single family house attached to 1 or more houses (e.g townhouse/ semi-detached)
◯ High rise apartment/ condo/ co-op (4 or more floors)
◯ Low rise apartment/ condo/ co-op (1-3 floors)
◯ Multi-family home (more than one family in a converted single home)
◯ Other type
◯ Prefer not to answer

Please provide the postal code of your secondary home

__________________________________
(Enter "555" if you would prefer not to answer.)

How many adults currently live at your secondary home (excluding yourself)?

◯ 0
◯ 1
◯ 2
◯ 3
◯ 4
◯ 5
◯ 6
◯ 7
◯ 8
◯ 9
◯ 10
◯ Prefer not to answer

How many children currently live at your secondary home?

◯ 0
◯ 1
◯ 2
◯ 3
◯ 4
◯ 5
◯ 6
◯ 7
◯ 8
◯ 9
◯ 10
◯ Prefer not to answer

How many bedrooms are in your secondary home?

◯ 0
◯ 1
◯ 2
◯ 3
◯ 4
◯ 5
◯ 6
◯ 7
◯ 8
◯ 9
◯ 10
◯ Prefer not to answer

How many toilets are in your secondary home?

◯ 0
◯ 1
◯ 2
◯ 3
◯ 4
◯ 5
◯ 6
◯ 7
◯ 8
◯ 9
◯ 10
◯ Prefer not to answer

| | |
|---|---|
| Choose the type of dwelling that best describes your secondary home | ◯ Manufactured home/ mobile home or trailer<br>◯ Single family detached house<br>◯ Single family house attached to 1 or more houses (e.g townhouse/ semi-detached)<br>◯ High rise apartment/ condo/ co-op (4 or more floors)<br>◯ Low rise apartment/ condo/ co-op (1-3 floors)<br>◯ Multi-family home (more than one family in a converted single home)<br>◯ Other type<br>◯ Prefer not to answer |
| Have you been working in any of the following occupations or worksites since you completed your second survey on [cv19a1_date]? (Select all that apply) | ☐ Hospital or health care facility worker<br>☐ First responder (paramedic, firefighter, police officer)<br>☐ Childcare worker<br>☐ Teacher/ other school staff (grade school, post-secondary, trade school, etc.)<br>☐ Transit Driver<br>☐ Food Service Industry<br>☐ Grocery Store<br>☐ Pharmacy<br>☐ Hairdresser/ barber<br>☐ Aesthetician<br>☐ Flight attendant<br>☐ Factory Worker<br>☐ Banker<br>☐ Truck Driver<br>☐ None of the above<br>☐ Prefer not to answer |
| Are any individuals living in any of your home(s) considered to be an ESSENTIAL WORKER? | ☐ Yes, I am<br>☐ Yes, another individual is<br>☐ No<br>☐ Don't Know<br>☐ Prefer not to answer<br>((e.g., healthcare, delivery worker, store worker, security, building maintenance)) |
| If yes, do they come home each day? | ☐ Yes, I do<br>☐ Yes, another individual does<br>☐ No, separated due to COVID-19<br>☐ No separated due to other reasons<br>☐ Prefer not to answer |
| If yes, are they a FIRST RESPONDER, HEALTHCARE PROVIDER or OTHER WORKER in a facility treating COVID-19? | ☐ Yes, I am<br>☐ Yes, another adult is<br>☐ No<br>☐ Don't Know<br>☐ Prefer not to answer |
| Please share any comments, suggestions or clarifications that you have related to this survey | __________________________________ |
| Did [setup_arm_2][cv19binfo_firstname] complete this survey? | ◯ Yes - by themselves<br>◯ Yes - partially or with help from someone else<br>◯ No - someone else completed this survey on their behalf |
| Today's Date | ____________________________ |

### Child Self Report Follow-up Survey 3

**Study ID: _ _ _ _ _ C _**

---

We are interested in learning more about how you are feeling and what you are thinking about during the Coronavirus (COVID-19) pandemic since you completed your third survey on ________________________.
(DD/MM/YYYY)

COVID-19 is a new type of disease affecting many people in Canada. The virus that causes COVID-19 can cause runny or stuffy noses, coughing, fever, muscle aches and tiredness. Other people with COVID-19 may not have any symptoms at all but they can spread the virus to others.

It is important for scientists to learn more about COVID-19. You can help us by answering this survey. Do the best you can to answer each question. This survey should take between 20 to 30 minutes to complete.

If you have questions or want help filling the survey out, please ask a family member to help you.

---

**1.**In general, how would you rate your overall PHYSICAL health (how healthy your body is)?

- ◯ Excellent
- ◯ Very Good
- ◯ Good
- ◯ Fair
- ◯ Poor
- ◯ I don't know
- ◯ I don't want to answer

---

**2.** In general, how would you rate your overall MENTAL/EMOTIONAL health (the way you think and feel)?

- ◯ Excellent
- ◯ Very Good
- ◯ Good
- ◯ Fair
- ◯ Poor
- ◯ I don't know
- ◯ I don't want to answer

---

**3.** How much do you weigh (in kilograms)?

______________________________
(Enter "666" if you don't know. Enter "555" if you don't want to answer.)

**4.** How often are you asking questions, reading, or talking about COVID-19, since you completed the third survey on ___________________________?
(DD/MM/YYYY)

- ○ Never
- ○ A few times a month
- ○ Weekly
- ○ A few times a week
- ○ Daily
- ○ I don't know
- ○ I don't want to answer

A vaccine is a type of medicine that doesn't make you better, but keeps you from getting sick in the first place. It does this by teaching your body to fight off germs like viruses and bacteria. The COVID-19 vaccine is given by a needle in the arm (shot) that will help your body be prepared to fight the COVID-19 virus in case you are exposed to it out in the world.

**5.** Have you had at least one dose (shot) of the COVID-19 vaccine?

- ○ Yes
- ○ No (Answer Question 5.12)
- ○ I don't know (Go to Question 6)
- ○ I don't want to answer (Go to Question 6)

(Answer 'Yes' if you have received at least one dose of the COVID-19 vaccine. Note: Certain types of vaccines require more than one dose to protect against COVID-19. You would have been informed at the time of vaccination if you needed a second dose.)

**5.1** Did you feel any side effects (feeling not well) from the COVID-19 vaccine? For example: sore arm, feeling tired, achy muscles, headache, fever and chills.

- ○ Yes
- ○ No (Go to Question 5.3)
- ○ I don't know (Go to Question 5.3)
- ○ I don't want to answer (Go to Question 5.3)

**5.2** If yes, check off all the side effects you felt.

- ☐ Head ache
- ☐ Tiredness
- ☐ Fever or chills
- ☐ Muscle or joint pain
- ☐ Soreness where the vaccine was injected
- ☐ Nausea or vomiting
- ☐ Other, specify ______________________________

(Enter "666" if you don't know. Enter "555" if you don't want to answer.)

- ☐ I don't know
- ☐ I don't want to answer

**5.3** How many doses (shots) of the COVID-19 vaccine have you had so far?

- ○ One dose
- ○ Two doses
- ○ Three doses
- ○ Four doses
- ○ More than four doses
- ○ I don't know (Go to Questio 6)
- ○ I don't want to answer (Go to Question 6)

**5.4** When did you receive your first dose of the COVID-19 vaccine?

______________________________ (DD/MM/YYYY)

(If you do not recall the exact date, please provide an estimate. Enter "01-01-1900" if you would prefer not to answer.)

**5.5** Which vaccine did you receive? (first dose)

- ○ Pfizer and BioNTech mRNA vaccine
- ○ Moderna mRNA vaccine
- ○ AstraZeneca Oxford vaccine
- ○ Janssen (Johnson & Johnson) vaccine
- ○ Other, specify ______________________

  (Enter "666" if you don't know. Enter "555" if you don't want to answer.)
- ○ I don't know
- ○ I don't want to answer

If you only received 1 dose of the COVID-19 vaccine, go to Question 6.
If you received more than 1 dose of the COVID-19 vaccine, continue to answer Question 6.6.

**5.6** When did you receive your second dose of the COVID-19 vaccine?

______________________ (DD/MM/YYYY)
(If you do not recall the exact date, please provide an estimate. Enter "01-01-1900" if you would prefer not to answer.)

**5.7** Which vaccine did you receive? (second dose)

- ○ Pfizer and BioNTech mRNA vaccine
- ○ Moderna mRNA vaccine
- ○ AstraZeneca Oxford vaccine
- ○ Janssen (Johnson & Johnson) vaccine
- ○ Other, specify ______________________

  (Enter "666" if you don't know. Enter "555" if you don't want to answer.)
- ○ I don't know
- ○ I don't want to answer

If you only received 2 doses of the COVID-19 vaccine, go to Question 6.
If you received more than 2 doses of the COVID-19 vaccine, continue to answer Question 5.8.

**5.8** When did you receive your third dose of the COVID-19 vaccine?

______________________ (DD/MM/YYYY)
(If you do not recall the exact date, please provide an estimate. Enter "01-01-1900" if you would prefer not to answer.)

**5.9** Which vaccine did you receive? (third dose)

- ○ Pfizer and BioNTech mRNA vaccine
- ○ Moderna mRNA vaccine
- ○ AstraZeneca Oxford vaccine
- ○ Janssen (Johnson & Johnson) vaccine
- ○ Other, specify ______________________

  (Enter "666" if you don't know. Enter "555" if you don't want to answer.)
- ○ I don't know
- ○ I don't want to answer

If you only received 3 doses of the COVID-19 vaccine, go to Question 6.
If you received more than 3 doses of the COVID-19 vaccine, continue to answer Question 5.10.

**5.10** When did you receive your fourth dose of the COVID-19 vaccine?

______________________ (DD/MM/YYYY)
(If you do not recall the exact date, please provide an estimate. Enter "01-01-1900" if you would prefer not to answer.)

**5.11** Which vaccine did you receive? (fourth dose)

- ◯ Pfizer and BioNTech mRNA vaccine
- ◯ Moderna mRNA vaccine
- ◯ AstraZeneca Oxford vaccine
- ◯ Janssen (Johnson & Johnson) vaccine
- ◯ Other, specify________________________________
  (Enter "666" if you don't know. Enter "555" if you don't want to answer.)
- ◯ I don't know
- ◯ I don't want to answer

(Go to Question 6)

**5.12** Are you old enough to receive a COVID-19 vaccine?

- ◯ Yes and I plan on getting a COVID-19 shot (Go to Question 6)
- ◯ Yes I'm old enough, but I DO NOT plan on getting a COVID-19 shot (Answer Question 5.13)
- ◯ No, I am not old enough yet (Answer Question 5.14)
- ◯ I don't know (Go to Question 6)
- ◯ I don't want to answer (Go to Question 6)

**5.13** What are some of the reasons why you don't want to get the COVID-19 vaccine?

- ☐ Not confident in the safety of the vaccine
- ☐ Concern about risks and side effects
- ☐ Will wait until it seems safe to get the vaccine
- ☐ Do not consider it necessary to get the vaccine
- ☐ Do not believe in vaccination
- ☐ Have not yet decided
- ☐ Have a pre-existing medical condition
- ☐ Already had or think I have had COVID-19
- ☐ Other, Specify________________________________
  (Enter "666" if you don't know. Enter "555" if you don't want to answer.)
- ☐ I don't know
- ☐ I don't want to answer

(Go to Question 6)

**5.14** When you are able to get the COVID-19 vaccine, how likely is it that you will choose to get it?

- ◯ Very Likely
- ◯ Somewhat Likely
- ◯ Somewhat Unlikely
- ◯ Very Unlikely
- ◯ I don't know
- ◯ I don't want to answer

**6.** Where do you normally look or go to for information on the COVID-19 vaccine? (check all that apply)

- ◯ My parents
- ◯ Family members other than my parents
- ◯ Family doctor/pediatrician
- ◯ Provincial government websites
- ◯ Government of Canada websites (example: PHAC, Health Canada)
- ◯ Other internet sites
- ◯ Social media/networking (examples: TikTok, Twitter, Facebook, Instagram)
- ◯ Religious leaders
- ◯ Friends
- ◯ Co-workers
- ◯ School/School Teachers
- ◯ Other, Specify________________________________
- ◯ None of these places
- ◯ Prefer not to answer
- ◯ I don't know

**Study ID: _ _ _ _ _ C _**

**7. The following are a number of mental health services that you may have accessed. Please check the relevant boxes for each service indicating if you have accessed the service since you completed the third survey on ____________________ or if you would be interested in accessing the service.**
(DD/MM/YYYY)

| | Have accessed this service since completing the third survey on _______________ (DD/MM/YYYY) | Would be interested in accessing this service | Not accessed, not interested | I don't know | I don't want to answer |
|---|---|---|---|---|---|
| Individual therapy (you alone) with a professional (in person or virtual) | ○ | ○ | ○ | ○ | ○ |
| Group therapy (you and others) with a professional (in person or virtual) | ○ | ○ | ○ | ○ | ○ |
| Family therapy (you and your family members) with a professional (in person or virtual) | ○ | ○ | ○ | ○ | ○ |
| Mental health mobile app (general) | ○ | ○ | ○ | ○ | ○ |
| Mental health mobile app to interact with a therapist | ○ | ○ | ○ | ○ | ○ |
| Phone helpline for mental health support (e.g. Crisis Services Canada) | ○ | ○ | ○ | ○ | ○ |
| Online mental health information you looked up yourself | ○ | ○ | ○ | ○ | ○ |

#### LIFE CHANGES DUE TO THE COVID-19 PANDEMIC

**8.** Based on what you know at this time, how safe do you think it is for students in your community to attend school in-person?

- ◯ Very safe
- ◯ Somewhat safe
- ◯ Not very safe
- ◯ Not safe at all
- ◯ I don't Know
- ◯ I don't want to answer

**9.** Since you completed the third survey on ______________________, (DD/MM/YYYY) how difficult has the cancellation of important events in your life (such as sporting events, school trips or vacations, etc.) been for you?

- ◯ Not difficult
- ◯ A little bit difficult
- ◯ Difficult
- ◯ Very difficult
- ◯ Extremely difficult
- ◯ There hasn't been any cancellation of important events
- ◯ I don't know
- ◯ I don't want to answer

**10.** During the PAST TWO WEEKS, how many people (who do not live with you) have you talked to in-person in an indoor environment?

- ◯ 0
- ◯ Less than 10
- ◯ 10 - 19
- ◯ 20 - 29
- ◯ 30 or more
- ◯ I don't know
- ◯ I don't want to answer

(In-person means that the person is in the same place as you (NOT on the phone or video chat))

**11.** During the PAST TWO WEEKS, how many people (who do not live with you) have you talked to in-person in an outdoor environment?

- ◯ 0
- ◯ Less than 10
- ◯ 10 - 19
- ◯ 20 - 29
- ◯ 30 or more
- ◯ I don't know
- ◯ I don't want to answer

(In-person means that the person is in the same place as you (NOT on the phone or video chat))

**12.** During the PAST TWO WEEKS, how many times did you go outside of your home (e.g., going to stores, parks, etc.)?

- ◯ Not at all
- ◯ 1-2 days per week
- ◯ 3-4 days per week
- ◯ 5-6 days per week
- ◯ Every day
- ◯ I don't know
- ◯ I don't want to answer

**13.** During the PAST TWO WEEKS, how stressful have the restrictions on leaving home been for you?

- ◯ Not stressful
- ◯ A little bit stressful
- ◯ Stressful
- ◯ Very stressful
- ◯ Extremely stressful
- ◯ There are no restrictions on leaving home
- ◯ I don't know
- ◯ I don't want to answer

**14.** During the PAST TWO WEEKS, how difficult has it been for you to follow the recommendations for keeping away from close contact with people who you don't live with?

- ◯ Not difficult
- ◯ A little bit difficult
- ◯ Difficult
- ◯ Very difficult
- ◯ Extremely difficult
- ◯ There are no recommendations for keeping away from close contact with people who I don't live with
- ◯ I don't know
- ◯ I don't want to answer

**15.** How hopeful are you that the COVID-19 pandemic in your area will end soon?

- ◯ Extremely hopeful
- ◯ Very hopeful
- ◯ Hopeful
- ◯ A little bit hopeful
- ◯ Neither hopeful nor not hopeful
- ◯ Not at all hopeful
- ◯ There is no COVID-19 pandemic in my area
- ◯ I don't know
- ◯ I don't want to answer

**Study ID: _ _ _ _ _ C _**

#### DAILY BEHAVIOURS (PAST TWO WEEKS)

**16.** During the PAST TWO WEEKS, have you experienced problems with sleep? (e.g., trouble falling asleep, staying asleep, waking up much earlier than intended, sleeping alone without a parent/caregiver)

- ◯ Yes
- ◯ No
- ◯ I don't know
- ◯ I don't want to answer

**17.** During the PAST TWO WEEKS, what time did you go to bed on WEEKDAYS (on average)?

- ◯ Before 8 pm
- ◯ 8 pm - 10 pm
- ◯ 10 pm - 12 am (Midnight)
- ◯ After 12 am (Midnight)
- ◯ I don't know
- ◯ I don't want to answer

**18.** During the PAST TWO WEEKS, what time did you go to bed on WEEKENDS (on average)?

- ◯ Before 8 pm
- ◯ 8 pm - 10 pm
- ◯ 10 pm - 12 am (Midnight)
- ◯ After 12 am (Midnight)
- ◯ I don't know
- ◯ I don't want to answer

**19.** During the PAST TWO WEEKS, how many hours per night did you sleep on WEEKDAYS (on average)?

- ◯ Less than 6 hours
- ◯ 6-8 hours
- ◯ 8-10 hours
- ◯ Greater than 10 hours
- ◯ I don't know
- ◯ I don't want to answer

**20.** During the PAST TWO WEEKS, how many hours per night did you sleep on WEEKENDS (on average)?

- ◯ Less than 6 hours
- ◯ 6-8 hours
- ◯ 8-10 hours
- ◯ Greater than 10 hours
- ◯ I don't know
- ◯ I don't want to answer

**21.** During the PAST TWO WEEKS, how many days per week did you exercise (where your heart rate and breathing increased) for at least 30 minutes over the entire day?

- ◯ None
- ◯ 1-2 days
- ◯ 3-4 days
- ◯ 5-6 days
- ◯ Daily
- ◯ I don't know
- ◯ I don't want to answer

**22.** During the PAST TWO WEEKS, how many days per week did you spend time outdoors?

- ◯ None
- ◯ 1-2 days
- ◯ 3-4 days
- ◯ 5-6 days
- ◯ Daily
- ◯ I don't know
- ◯ I don't want to answer

**Study ID: _ _ _ _ _ C _**

#### EMOTIONS/WORRIES (PAST TWO WEEKS)

| | Not at all worried | Slightly worried | Moderately worried | Very worried | Extremely worried | I don't know | I don't want to answer |
|---|---|---|---|---|---|---|---|
| **23.** During the PAST TWO WEEKS, how worried have you been about being infected by the virus that causes COVID-19? | ○ | ○ | ○ | ○ | ○ | ○ | ○ |
| **24.** During the PAST TWO WEEKS, how worried have you been about friends or family being infected by the virus that causes COVID-19? | ○ | ○ | ○ | ○ | ○ | ○ | ○ |
| **25.** During the PAST TWO WEEKS, how worried have you been about your physical health (how healthy your body is) being influenced by COVID-19? | ○ | ○ | ○ | ○ | ○ | ○ | ○ |
| **26.** During the PAST TWO WEEKS, how worried have you been about your mental/emotional health (how you think and feel) being influenced by COVID-19? | ○ | ○ | ○ | ○ | ○ | ○ | ○ |
| **27.** During the PAST TWO WEEKS, how worried were you generally? | ○ | ○ | ○ | ○ | ○ | ○ | ○ |

**28.** During the PAST TWO WEEKS, how happy versus sad were you?

- ○ Very happy/cheerful
- ○ Moderately happy/cheerful
- ○ Neutral
- ○ Moderately sad/depressed/unhappy
- ○ Very sad/depressed/unhappy
- ○ I don't know
- ○ I don't want to answer

**29.** During the PAST TWO WEEKS, how relaxed versus anxious were you?

- ○ Very relaxed/calm
- ○ Moderately relaxed/calm
- ○ Neutral
- ○ Moderately nervous/anxious
- ○ Very nervous/anxious
- ○ I don't know
- ○ I don't want to answer

**30.** During the PAST TWO WEEKS, how fidgety or restless were you?

- ○ Not fidgety/restless at all
- ○ Slightly fidgety/restless
- ○ Moderately fidgety/restless
- ○ Very fidgety/restless
- ○ Extremely fidgety/restless
- ○ I don't know
- ○ I don't want to answer

| | |
|---|---|
| **31.** During the PAST TWO WEEKS, how tired were you? | ◯ Not tired at all<br>◯ Slightly tired<br>◯ Moderately tired<br>◯ Very tired<br>◯ Extremely tired<br>◯ I don't know<br>◯ I don't want to answer |
| **32.** During the PAST TWO WEEKS, how well were you able to concentrate or focus? | ◯ Very focused/attentive<br>◯ Moderately focused/attentive<br>◯ Neutral<br>◯ Moderately unfocused/distracted<br>◯ Very unfocused/distracted<br>◯ I don't know<br>◯ I don't want to answer |
| **33.** During the PAST TWO WEEKS, how irritable or easily angered were you? | ◯ Not irritable or easily angered at all<br>◯ Slightly irritable or easily angered<br>◯ Moderately irritable or easily angered<br>◯ Very irritable or easily angered<br>◯ Extremely irritable or easily angered<br>◯ I don't know<br>◯ I don't want to answer |
| **34.** During the PAST TWO WEEKS, how lonely were you? | ◯ Not lonely at all<br>◯ Slightly lonely<br>◯ Moderately lonely<br>◯ Very lonely<br>◯ Extremely lonely<br>◯ I don't know<br>◯ I don't want to answer |
| **35.** Please share any comments, suggestions or clarifications that you have related to this survey | ________________________________ |
| **36.** Did **you** complete this survey on your own?<br>Note: **You** refers to the child participating in the study, who was assigned this survey | ◯ Yes - by myself<br>◯ Yes - partially or with help from someone else<br>◯ No - someone else completed this survey on my behalf<br>◯ I don't know<br>◯ I don't want to answer |
| **37.** Today's Date | _________________________ (DD/MM/YYYY) |

### Parental Report Follow-up Survey 3

Study ID: _ _ _ _ _ C _

We are interested in learning more about how the child is feeling and what they are thinking about during the Coronavirus (COVID-19) pandemic since you completed your third survey on ____________________.
(DD/MM/YYYY)

COVID-19 is a new type of illness affecting many people in Canada. The virus that causes COVID-19 is related to the common cold virus. Some people affected by the virus may have symptoms that can be mild or severe, similar to a cold or flu. Other people with COVID-19 may not have any symptoms at all but they can spread the virus to others.

It is important for scientists to learn more about COVID-19. You can help us by answering this survey about the child. Do the best you can to answer each question. This survey should take between 20 to 30 minutes to complete.

**1.** In general, how would you rate the child's overall PHYSICAL health now?

- Excellent
- Very Good
- Good
- Fair
- Poor
- Prefer not to answer

**2.** In general, how would you rate the child's overall MENTAL/EMOTIONAL health now?

- Excellent
- Very Good
- Good
- Fair
- Poor
- Prefer not to answer

**3.** Child's weight in kilograms?

______________________________
(Enter "555" if you would prefer not to answer.)

**Study ID:** _ _ _ _ _ C _

---

**4.** In what ways has the COVID-19 outbreak affected the child's overall healthcare, since you completed the third survey on______________________? (Select all that apply) (DD/MM/YYYY)

- ☐ The child did not go to healthcare appointments because I was concerned about entering the healthcare provider's office
- ☐ The child's healthcare provider cancelled appointments
- ☐ The child's healthcare provider changed to phone or online visits (Go to Question 5)
- ☐ The child's healthcare provider told him/her to self-isolate or quarantine (Go to Question 5)
- ☐ None of these apply (Go to Question 5)
- ☐ Prefer not to answer (Go to Question 5)

---

**4.1** What kind of healthcare visits were affected since you completed the third survey on ______________________? (Select all that apply) (DD/MM/YYYY)

- ☐ Routine visit/ regular checkup
- ☐ Vaccination visit
- ☐ Visit with a health care professional for a specific concern
- ☐ Visit with a specialist
- ☐ I did not miss any healthcare visits
- ☐ Prefer not to answer

---

**5.** Has the child been exposed to someone likely to have COVID-19, since you completed the third survey on ____________________________? (Select all that apply) (DD/MM/YYYY)

- ☐ Yes, to someone with positive test
- ☐ Yes, to someone with medical diagnosis, but no test
- ☐ Yes, to someone with possible symptoms, but no diagnosis by doctor
- ☐ No, not to my knowledge
- ☐ Prefer not to answer

---

**6.** Has the child been suspected of having COVID-19, since you completed the third survey on ______________________? (DD/MM/YYYY)

- ○ Yes, had positive test
- ○ Yes, had medical diagnosis, but no test
- ○ Yes, had some possible symptoms, but no diagnosis by health care professional
- ○ No
- ○ Don't know
- ○ Prefer not to answer

---

**7.** Has the child been tested for the virus that causes COVID-19, since you completed the third survey on ______________________? (DD/MM/YYYY) (Select all that apply)

Do not include the antibody/ serology test (blood test for previous infection) that is part of this research study.

- ☐ No, I never tried to get the child tested (Go to Question 7.3)
- ☐ No, I tried to get the child tested but was not able to (Go to Question 7.1)
- ☐ Yes, and the child is waiting for the results (Go to Question 7.2)
- ☐ Yes, and the test showed that the child did not have it ("negative" test) (Go to Question 7.2)
- ☐ Yes, and the test showed that the child did have it ("positive" test) (Go to Question 7.2)
- ☐ Prefer not to answer (Go to Question 8)

---

**7.1** Which type of test for the virus that causes COVID-19 did you try to get for the child but were not able to?

- ☐ Viral Test (swab test for current infection)
- ☐ Antibody/ Serology Test (blood test for previous infection)
- ☐ Saliva or Mouth Rinse Test
- ☐ Rapid-Antigen Test
- ☐ Other Specify ____________________________
  (Enter "555" if you would prefer not to answer.)
- ☐ Don't Know
- ☐ Prefer not to answer

(Antibody/ Serology testing and saliva/ mouth rinse testing may not be currently available in all areas, but it may be available in the future.)

| | |
|---|---|
| **7.2** Please specify why the child got tested or tried to get tested | ☐ A contact or possible contact of the child tested positive for COVID-19<br>☐ The child had COVID-19 symptoms<br>☐ The child was admitted to hospital (unrelated to COVID-19)<br>☐ The child was admitted to hospital (COVID-19 related)<br>☐ The child was tested in another facility or instiution context (contact with someone in long term care, prison etc)<br>☐ The child was participating in another facility research study<br>☐ Travel-related testing<br>☐ Other, specify ____________<br>(Enter "555" if you would prefer not to answer.)<br>☐ Don't Know<br>☐ Prefer not to answer<br>(Go to Question 7.4) |
| **7.3** Please specify why you didn't try to get the child tested | ☐ Symptoms were due to another condition, specify ____________<br>(Enter "555" if you would prefer not to answer.)<br>☐ Symptoms were mild<br>☐ A healthcare provider did not recommend testing<br>☐ Other, specify ____________<br>(Enter "555" if you would prefer not to answer.)<br>☐ Prefer not to answer<br>(Go to Question 8) |
| **7.4** IN THE PAST TWO WEEKS, how many times has the child been tested for COVID-19?<br><br>This can include routine testing (e.g. for work, school, events, etc.) | ○ 0 (Go to Question 8)<br>○ 1<br>○ 2<br>○ 3<br>○ 4<br>○ Prefer not to answer (Go to Question 8) |
| Details of First COVID-19 Test | |
| **7.5** First test result | ○ Negative<br>○ Positive<br>○ Don't know<br>○ Prefer not to answer |
| In the past two weeks has the child been tested for COVID-19 a second time? | ○ Yes<br>○ No (If you had a positive COVID test, go to Question 7.9. If not, go to Question 8) |
| Details of Second COVID-19 Test | |
| **7.6** Second test result | ○ Negative<br>○ Positive<br>○ Don't know<br>○ Prefer not to answer |
| In the past two weeks has the child been tested for COVID-19 a third time? | ○ Yes<br>○ No (If you had a positive COVID test, go to Question 7.9. If not, go to Question 8) |

**Study ID:** _ _ _ _ _ **C** _

Details of Third COVID-19 Test

**7.7** Third test result

- ◯ Negative
- ◯ Positive
- ◯ Don't know
- ◯ Prefer not to answer

In the past two weeks has the child been tested for COVID-19 a fourth time?

- ◯ Yes
- ◯ No (If you positive COVID test, go to Question 7.9. If not, go to Question 8)

Details of Fourth COVID-19 Test

**7.8** Fourth test result

- ◯ Negative
- ◯ Positive
- ◯ Don't know
- ◯ Prefer not to answer

**7.9** When people still show symptoms of COVID-19 for weeks or months after their initial recovery, it's called post or long COVID-19 condition. Post-COVID-19 can happen anywhere from 4 to 12 weeks after you first have COVID-19. Does the child have any post-COVID symptoms? (select all that apply)

- ☐ Tired/fatigue
- ☐ Headaches
- ☐ Weight loss
- ☐ Muscle Pain
- ☐ Trouble sleeping or staying asleep
- ☐ Stuffy or runny nose
- ☐ Difficulty thinking or concentrating
- ☐ Prefer not to answer

**8.** Has the child had any of the following symptoms, since you completed the third survey on ______________________? (Select all that apply)

(DD/MM/YYYY)

- ☐ New or worsening cough
- ☐ Shortness of breath
- ☐ Difficulty breathing
- ☐ Fever
- ☐ Chills
- ☐ Muscle or body aches
- ☐ Headache
- ☐ Sore throat
- ☐ New loss of sense of smell or taste
- ☐ Runny nose or nasal congestion*
- ☐ Nausea or vomiting
- ☐ Diarrhea
- ☐ Fatigue
- ☐ Skin Rash
- ☐ Conjunctivitis (pink eye)
- ☐ Other, Specify______________________________ ( Enter "555" if you would prefer not to answer.)
- ☐ None of the above (Go to Question 9)
- ☐ Prefer not to answer (Go to Question 9)

(*Runny nose or nasal congestion in the absence of underlying reason for these symptoms such as seasonal allergies, post nasal drip, etc)

**8.1** In the TWO WEEKS before the child had symptoms, did the child: (Select all that apply)

- ☐ Have contact with someone who tested positive for COVID-19 (Go to Question 8.7)
- ☐ Have contact with someone who likely had COVID-19 (e.g., was not tested but had symptoms; was told by a healthcare provider that he/she likely had it) (Go to Question 8.7)
- ☐ Travel outside the province
- ☐ None of the above (Go to Question 8.7)
- ☐ Prefer not to answer (Go to Question 8.7)

**Study ID: _ _ _ _ _ C _**

**8.2** How many places did the child travel?

- ○ 0 (Go to Question 8.7)
- ○ 1
- ○ 2
- ○ 3
- ○ 4
- ○ Prefer not to answer (Go to Question 8.7)

(Travel outside the province)

Place 1

**8.3** Location
(e.g. Seattle, Washington OR Toronto, Ontario)

________________________
(Enter "555" if you would prefer not to answer.)

Did the child travel to a second place?

- ○ Yes
- ○ No (Go to Question 8.7)

Place 2

**8.4** Location
(e.g. Portland, Oregon OR Vancouver, BC)

________________________
(Enter "555" if you would prefer not to answer.)

Did the child travel to a third place?

- ○ Yes
- ○ No (Go to Question 8.7)

Place 3

**8.5** Location
(e.g. Paris, France OR Banff, Alberta)

________________________
(Enter "555" if you would prefer not to answer.)

Did the child travel to a third place?

- ○ Yes
- ○ No (Go to Question 8.7)

Place 4

**8.6** Location
(e.g. Portland, Oregon OR Vancouver, BC)

________________________
(Enter "555" if you would prefer not to answer.)

**8.7** Which of the following occurred as a result of the child's symptoms? (Select all that apply)

- ☐ You/ the child spoke to a healthcare provider over the phone, by email, or online
- ☐ The child saw a healthcare provider in person, such as in a clinic or doctor's office
- ☐ The child saw a healthcare provider in person, such as in urgent care, or in an Emergency Department (ED)
- ☐ The child was kept overnight in a hospital because a healthcare provider thought he/she had COVID-19
- ☐ The child was admitted to a hospital's ICU (Intensive Care Unit)
- ☐ The child self-isolated or quarantined at home
- ☐ None of the above
- ☐ Prefer not to answer

**9.** Has anyone in the child's family been diagnosed with COVID-19, since you completed the third survey on ________________________? (Select all that apply)
(DD/MM/YYYY)

- ☐ Yes, member of household
- ☐ Yes, non-household member
- ☐ No
- ☐ Don't know
- ☐ Prefer not to answer

**10.** Have any of the following happened to family members living inside the child's household because of COVID-19, since you completed the third survey on ________________________? (Select all that apply)
(DD/MM/YYYY)

- ☐ Fallen physically ill
- ☐ Hospitalized
- ☐ Self-isolated or quarantined with symptoms
- ☐ Self-isolated or quarantined without symptoms (e.g., due to possible exposure)
- ☐ Lost job or been laid off from job
- ☐ Reduced ability to earn money
- ☐ Passed away
- ☐ None of the above
- ☐ Prefer not to answer

**11.** Have any of the following happened to family members living outside the child's household because of COVID-19, since you completed the third survey on ________________________? (Select all that apply)
(DD/MM/YYYY)

- ☐ Fallen physically ill
- ☐ Hospitalized
- ☐ Self-isolated or quarantined with symptoms
- ☐ Self-isolated or quarantined without symptoms (e.g., due to possible exposure)
- ☐ Lost job or been laid off from job
- ☐ Reduced ability to earn money
- ☐ Passed away
- ☐ None of the above
- ☐ Prefer not to answer

**12.** Did the child get the flu shot, since you completed the third survey on ____________________?
(DD/MM/YYYY)

- ○ Yes
- ○ No
- ○ Don't Know
- ○ Prefer not to answer

**13.** Has the child received at least one dose of a COVID-19 vaccine?

- ○ Yes
- ○ No (Go to Question 13.12)
- ○ Prefer not to answer (Go to Question 14)

(Answer 'Yes' if the child has received at least one dose of the COVID-19 vaccine. Note: Certain types of vaccines require more than one dose to protect against COVID-19. You or the child would have been informed at the time of vaccination if the child needed a second dose.)

**13.1** Did the child feel any side effects from the COVID-19 vaccine?

- ○ Yes
- ○ No
- ○ Prefer not to answer

**13.2** If yes, check all that apply.

- ☐ Head ache
- ☐ Tiredness
- ☐ Fever or chills
- ☐ Muscle or joint pain
- ☐ Soreness where the vaccine was injected
- ☐ Nausea or vomiting
- ☐ Other, specify ______________________________
  (Enter "555" if you would prefer not to answer.)
- ☐ Prefer not to answer

**13.3** How many doses of the COVID-19 vaccine has the child received so far?

- ○ One dose
- ○ Two doses
- ○ Three doses
- ○ Four doses
- ○ More than four doses
- ○ Prefer not to answer

**13.4** When did the child receive their first dose of the COVID-19 vaccine?

________________________

(If you do not recall the exact date, please provide an estimate. Enter "01-01-1900" if you would prefer not to answer.)

**13.5** Which vaccine did the child receive? (first dose)

- ○ Pfizer and BioNTech mRNA vaccine
- ○ Moderna mRNA vaccine
- ○ AstraZeneca Oxford vaccine
- ○ Janssen (Johnson & Johnson) vaccine
- ○ Other, specify________________________
  (Enter "555" if you would prefer not to answer.)
- ○ Don't Know
- ○ Prefer not to answer

If the child only received 1 dose of the COVID-19 vaccine, go to Question 14.
If the child received more than 1 dose of the COVID-19 vaccine, continue to answer Question 13.6.

**13.6** When did the child receive their second dose of the COVID-19 vaccine?

________________________

(If you do not recall the exact date, please provide an estimate. Enter "01-01-1900" if you would prefer not to answer.)

**13.7** Which vaccine did the child receive? (second dose)

- ○ Pfizer and BioNTech mRNA vaccine
- ○ Moderna mRNA vaccine
- ○ AstraZeneca Oxford vaccine
- ○ Janssen (Johnson & Johnson) vaccine
- ○ Other, specify________________________
  (Enter "555" if you would prefer not to answer.)
- ○ Don't Know
- ○ Prefer not to answer

If the child only received 2 doses of the COVID-19 vaccine, go to Question 14.
If the child received more than 2 doses of the COVID-19 vaccine, continue to answer Question 13.8.

**13.8** When did the child receive their third dose of the COVID-19 vaccine?

________________________

(If you do not recall the exact date, please provide an estimate. Enter "01-01-1900" if you would prefer not to answer.)

**13.9** Which vaccine did the child receive? (third dose)

- ○ Pfizer and BioNTech mRNA vaccine
- ○ Moderna mRNA vaccine
- ○ AstraZeneca Oxford vaccine
- ○ Janssen (Johnson & Johnson) vaccine
- ○ Other, specify________________________
  (Enter "555" if you would prefer not to answer.)
- ○ Don't Know
- ○ Prefer not to answer

If the child only received 3 doses of the COVID-19 vaccine, go to Question 14.
If the child received more than 3 doses of the COVID-19 vaccine, continue to answer Question 13.10.

**13.10** When did the child receive their fourth dose of the COVID-19 vaccine?

________________________

(If you do not recall the exact date, please provide an estimate. Enter "01-01-1900" if you would prefer not to answer.)

**13.11** Which vaccine did the child receive? (fourth dose)

- ◯ Pfizer and BioNTech mRNA vaccine
- ◯ Moderna mRNA vaccine
- ◯ AstraZeneca Oxford vaccine
- ◯ Janssen (Johnson & Johnson) vaccine
- ◯ Other, specify ______________________
  (Enter "555" if you would prefer not to answer.)
- ◯ Don't Know
- ◯ Prefer not to answer

---

**13.12** Is the child eligible to receive a COVID-19 vaccine?

- ◯ Yes and we plan to schedule a COVID-19 vaccination for the child (Go to Question 14)
- ◯ Yes, but we DO NOT plan to schedule a COVID-19 vaccination for the child
- ◯ No, the child is not eligible for a COVID-19 vaccine yet (Go to Question 13.14)
- ◯ Don't know (Go to Question 13.14)
- ◯ Prefer not to answer(Go to Question 14)

---

**13.13** What are some of the reasons why you don't want THE CHILD to get the COVID-19 vaccine?

- ☐ Not confident in the safety of the vaccine
- ☐ Concern about risks and side effects
- ☐ Will wait until it seems safe to get the vaccine
- ☐ Do not consider it necessary to get the vaccine
- ☐ Do not believe in vaccination
- ☐ Have not yet decided
- ☐ Have a pre-existing medical condition
- ☐ Already had or think I have had COVID-19
- ☐ Other, Specify ______________________
  ( Enter "555" if you would prefer not to answer.)
- ☐ Prefer not to answer

(Check all that apply.)

(Go to Question 14)

---

**13.14** When a COVID-19 vaccine becomes available, how likely is it that you will choose for THE CHILD to get it?

- ◯ Very Likely
- ◯ Somewhat Likely
- ◯ Somewhat Unlikely
- ◯ Very Unlikely
- ◯ Don't Know
- ◯ Prefer not to answer

---

| | Never | Rarely | Occasionally | Often | Always | Don't Know | Prefer not to answer |
|---|---|---|---|---|---|---|---|
| **14.** How often has the child worn a mask in public places since you completed the third survey on ______ _______ ? (DD/MM/YYYY) | ◯ | ◯ | ◯ | ◯ | ◯ | ◯ | ◯ |

**Study ID:** _ _ _ _ _ C _

| | | | | | | | |
|---|---|---|---|---|---|---|---|
| **15.** How often has the child practiced physical distancing in public places since you completed the third survey on ____________________ ? (DD/MM/YYYY) | ○ | ○ | ○ | ○ | ○ | ○ | ○ |
| **16.** How often has the child avoided crowded places/ gatherings since you completed the third survey on ____________________? (DD/MM/YYYY) | ○ | ○ | ○ | ○ | ○ | ○ | ○ |
| **17.** How often has the child avoided common greetings (e.g handshakes, hugs etc) since you completed the third survey on ____________________? (DD/MM/YYYY) | ○ | ○ | ○ | ○ | ○ | ○ | ○ |
| **18.** How often has the child limited contact with people at higher risk (e.g. elderly relative) since you completed the third survey on ______________? (DD/MM/YYYY) | ○ | ○ | ○ | ○ | ○ | ○ | ○ |
| **19.** How often has the child self-Isolated because someone thought the child was infected with the virus that causes COVID-19 since you completed the third survey on ____________________? (DD/MM/YYYY) | ○ | ○ | ○ | ○ | ○ | ○ | ○ |
| **20.** How often has the child quarantined because the child may have been exposed to the virus that causes COVID-19, but did not show symptoms since you completed the third survey on________________ ? (DD/MM/YYYY) | ○ | ○ | ○ | ○ | ○ | ○ | ○ |

**21.** Has the child participated in any organized sports or recreational activities, in person, since you completed the third survey on ____________________? (DD/MM/YYYY)

- ○ Yes
- ○ No (Go to Question 22)
- ○ Prefer not to answer (Go to Question 22)

**21.1** If yes, how many children were in the activity?

- ○ 0-9
- ○ 10-19
- ○ 20-29
- ○ 30+
- ○ Prefer not to answer

**22.** How many times has the child been in a gathering* of >10 or more people (excluding school), since you completed the third survey on ________________________?
(DD/MM/YYYY)

________________________________

(A gathering in this context is defined as any assembly or meeting of more than 10 persons, in open air or in a building or premises. These can include gatherings with or without masks or social distancing. Gatherings include a range of interpersonal gatherings/events of varying purposes, from gatherings/events of family and friends (e.g., sports, weddings, funerals, baptisms, birthday parties and cultural ceremonies) to community gathering spaces (e.g., places of worship, places of traditional gatherings, libraries, community and recreation centres, camps, drop-in centres, food banks and communal kitchens, museums, theatres, cinemas, and tourist attractions) to larger planned or structured gatherings/events, including mass gatherings (e.g., large meetings or conferences, national sporting events, large cultural/religious events, festivals). Enter "555" if you would prefer not to answer.)

**Study ID: _ _ _ _ _ C _**

**23. The following are a number of mental health services that the child may have accessed. Please check the relevant boxes for each service indicating if the child accessed the service since completing the third survey on ____________________________ or if the child would be interested in accessing the service.** (DD/MM/YYYY)

| | Have accessed this service since completing the third survey on ____________________ (DD/MM/YYYY) | Would be interested in accessing this service | Not accessed, not interested | Prefer not to answer |
|---|---|---|---|---|
| Individual therapy (you alone) with a professional (in person or virtual) | ○ | ○ | ○ | ○ |
| Group therapy (you and others) with a professional (in person or virtual) | ○ | ○ | ○ | ○ |
| Family therapy (you and your family members) with a professional (in person or virtual) | ○ | ○ | ○ | ○ |
| Mental health mobile app (general) | ○ | ○ | ○ | ○ |
| Mental health mobile app to interact with a therapist | ○ | ○ | ○ | ○ |
| Phone helpline for mental health support (e.g. Crisis Services Canada) | ○ | ○ | ○ | ○ |
| Online mental health information you looked up yourself | ○ | ○ | ○ | ○ |

#### Worrying (PAST TWO WEEKS)

| | Not at all worried | Slightly worried | Moderately worried | Very worried | Extremely worried | Prefer not to answer |
|---|---|---|---|---|---|---|
| **24.** During the PAST TWO WEEKS, how worried has the child been about being infected by the virus that causes COVID-19? | ○ | ○ | ○ | ○ | ○ | ○ |
| **25.** During the PAST TWO WEEKS, how worried has the child been about friends or family being infected by the virus that causes COVID-19? | ○ | ○ | ○ | ○ | ○ | ○ |
| **26.** During the PAST TWO WEEKS, how worried has the child been about his/her physical health being influenced by COVID-19? | ○ | ○ | ○ | ○ | ○ | ○ |
| **27.** During the PAST TWO WEEKS, how worried has the child been about his/her mental/emotional health being influenced by COVID-19? | ○ | ○ | ○ | ○ | ○ | ○ |
| **28.** During the PAST TWO WEEKS, how worried was the child generally? | ○ | ○ | ○ | ○ | ○ | ○ |

**29.** How much is the child asking questions, reading, or talking about COVID-19, since you completed the third survey on _________________________? (DD/MM/YYYY)

- ○ Never
- ○ A few times a month
- ○ Weekly
- ○ A few times a week
- ○ Daily
- ○ Prefer not to answer

**30.** Based on what you know at this time, how safe do you think it is for students in your community to attend school in-person?

- ○ Very safe
- ○ Somewhat safe
- ○ Not very safe
- ○ Not safe at all
- ○ Don't Know
- ○ Prefer not to answer

**31.** Since you completed the third survey on _________________________, (DD/MM/YYYY) how difficult has the cancellation of important events in the child's life (such as sporting events, school trips or vacations, etc.) been difficult for the child?

- ○ Not difficult
- ○ A little bit difficult
- ○ Difficult
- ○ Very difficult
- ○ Extremely difficult
- ○ There hasn't been any cancellation of important events
- ○ Prefer not to answer

**32.** During the PAST TWO WEEKS, how many people, from outside the child's household, has the child had an in-person conversation with in an indoor environment? This can include things like work, school or other activities with or without masks and/or social distancing. If you aren't sure, please estimate a number.

- 0
- Less than 10
- 10 - 19
- 20 - 29
- 30 or more
- Prefer not to answer

(In-person means that the person is in the same place as the child (NOT on the phone or video chat))

**33.** During the PAST TWO WEEKS, how many people, from outside the child's household, has the child had an in-person conversation with in an outdoor environment? This can include things like work, school or other activities with or without masks and/or social distancing. If you aren't sure, please estimate a number.

- 0
- Less than 10
- 10 - 19
- 20 - 29
- 30 or more
- Prefer not to answer

(In-person means that the person is in the same place as the child (NOT on the phone or video chat) )

**34.** During the PAST TWO WEEKS, how many times did the child go outside of their home (e.g., going to stores, parks, etc)?

- Not at all
- 1-2 days per week or less
- 3-4 days per week
- 5-6 days per week
- Daily
- Prefer not to answer

**35.** During the PAST TWO WEEKS, how stressful have the restrictions on leaving home been for the child?

- Not stressful
- A little bit stressful
- Stressful
- Very stressful
- Extremely stressful
- There are no restrictions on leaving home
- Prefer not to answer

**36.** During the PAST TWO WEEKS, how difficult has it been for the child to follow the recommendations for keeping away from close contact with people who they don't live with?

- Not difficult
- A little bit difficult
- Difficult
- Very difficult
- Extremely difficult
- There are no recommendations for keeping away from close contact with people who they don't live with
- Prefer not to answer

**37.** How hopeful is the child that the COVID-19 pandemic in the child's area will end soon?

- Extremely hopeful
- Very hopeful
- Hopeful
- A little bit hopeful
- Neither hopeful nor not hopeful
- Not at all hopeful
- There is no COVID-19 pandemic in the child's area
- Prefer not to answer

#### DAILY BEHAVIOURS (PAST TWO WEEKS)

**38.** During the PAST TWO WEEKS, has the child experienced problems with sleep? (e.g., trouble falling asleep, staying asleep, waking up much earlier than intended, sleeping alone without a parent/caregiver)

- ◯ Yes
- ◯ No
- ◯ Prefer not to answer

**39.** During the PAST TWO WEEKS, what time did the child go to bed on WEEKDAYS (on average)?

- ◯ Before 8 pm
- ◯ 8 pm - 10 pm
- ◯ 10 pm - 12 am (Midnight)
- ◯ After 12 am (Midnight)
- ◯ Prefer not to answer

**40.** During the PAST TWO WEEKS, what time did the child go to bed on WEEKENDS (on average)?

- ◯ Before 8 pm
- ◯ 8 pm - 10 pm
- ◯ 10 pm - 12 am (Midnight)
- ◯ After 12 am (Midnight)
- ◯ Prefer not to answer

**41.** During the PAST TWO WEEKS, how many hours per night did the child sleep on WEEKDAYS (on average)?

- ◯ Less than 6 hours
- ◯ 6-8 hours
- ◯ 8-10 hours
- ◯ Greater than 10 hours
- ◯ Prefer not to answer

**42.** During the PAST TWO WEEKS, how many hours per night did the child sleep on WEEKENDS (on average)?

- ◯ Less than 6 hours
- ◯ 6-8 hours
- ◯ 8-10 hours
- ◯ Greater than 10 hours
- ◯ Prefer not to answer

**43.** During the PAST TWO WEEKS, how many days per week did the child exercise (e.g., increased heart rate, breathing) for at least 30 minutes over the entire day?

- ◯ Not at all
- ◯ 1-2 days per week or less
- ◯ 3-4 days per week
- ◯ 5-6 days per week
- ◯ Daily
- ◯ Prefer not to answer

**44.** During the PAST TWO WEEKS, how many days per week did the child spend time outdoors?

- ◯ Not at all
- ◯ 1-2 days per week or less
- ◯ 3-4 days per week
- ◯ 5-6 days per week
- ◯ Daily
- ◯ Prefer not to answer

#### EMOTIONS/WORRIES (PAST TWO WEEKS)

**45.** During the PAST TWO WEEKS, how happy versus sad was the child?

- ○ Very happy/cheerful
- ○ Moderately happy/cheerful
- ○ Neutral
- ○ Moderately sad/depressed/unhappy
- ○ Very sad/depressed/unhappy
- ○ Prefer not to answer

**46.** During the PAST TWO WEEKS, how relaxed versus anxious was the child?

- ○ Very relaxed/calm
- ○ Moderately relaxed/calm
- ○ Neutral
- ○ Moderately nervous/anxious
- ○ Very nervous/anxious
- ○ Prefer not to answer

**47.** During the PAST TWO WEEKS, how fidgety or restless was the child?

- ○ Not fidgety/restless at all
- ○ Slightly fidgety/restless
- ○ Moderately fidgety/restless
- ○ Very fidgety/restless
- ○ Extremely fidgety/restless
- ○ Prefer not to answer

**48.** During the PAST TWO WEEKS, how tired was the child?

- ○ Not tired at all
- ○ Slightly tired
- ○ Moderately tired
- ○ Very tired
- ○ Extremely tired
- ○ Prefer not to answer

**49.** During the PAST TWO WEEKS, how well was the child able to concentrate or focus?

- ○ Very focused/attentive
- ○ Moderately focused/attentive
- ○ Neutral
- ○ Moderately unfocused/distracted
- ○ Very unfocused/distracted
- ○ Prefer not to answer

**50.** During the PAST TWO WEEKS, how irritable or easily angered was the child?

- ○ Not irritable or easily angered at all
- ○ Slightly irritable or easily angered
- ○ Moderately irritable or easily angered
- ○ Very irritable or easily angered
- ○ Extremely irritable or easily angered
- ○ Prefer not to answer

**51.** During the PAST TWO WEEKS, how lonely was the child?

- ○ Not lonely at all
- ○ Slightly lonely
- ○ Moderately lonely
- ○ Very lonely
- ○ Extremely lonely
- ○ Prefer not to answer

#### DEMOGRAPHIC INFORMATION

**52.** Has the child's living situation changed since you completed the third survey on ________________________? (DD/MM/YYYY)

- ☐ No, the child's living situation has not changed (Go to Question 53)
- ☐ Yes, the child has have moved (since the third survey)
- ☐ Yes, the child regularly stays in more than one home (since the third survey)
- ☐ Yes, the number of people living in the child's home has changed (since the third survey) (Go to Question 52.2)
- ☐ Prefer not to answer (Go to Question 53)

**52.1** Please provide the postal code of the child's primary home

______________________________

(Enter "555" if you would prefer not to answer.)

**52.2** How many adults currently live at the child's primary home?

- ◯ 0
- ◯ 1
- ◯ 2
- ◯ 3
- ◯ 4
- ◯ 5
- ◯ 6
- ◯ 7
- ◯ 8
- ◯ 9
- ◯ 10
- ◯ Prefer not to answer

**52.3** How many children currently live at the child's primary home (excluding the child)?

- ◯ 0
- ◯ 1
- ◯ 2
- ◯ 3
- ◯ 4
- ◯ 5
- ◯ 6
- ◯ 7
- ◯ 8
- ◯ 9
- ◯ 10
- ◯ Prefer not to answer

**52.4** How many bedrooms are in the child's primary home?

- ◯ 0
- ◯ 1
- ◯ 2
- ◯ 3
- ◯ 4
- ◯ 5
- ◯ 6
- ◯ 7
- ◯ 8
- ◯ 9
- ◯ 10
- ◯ Prefer not to answer

**52.5** How many toilets are in the child's primary home?

- ◯ 0
- ◯ 1
- ◯ 2
- ◯ 3
- ◯ 4
- ◯ 5
- ◯ 6
- ◯ 7
- ◯ 8
- ◯ 9
- ◯ 10
- ◯ Prefer not to answer

---

**52.6** Choose the type of dwelling that best describes the child's primary home

- ◯ Manufactured home/ mobile home or trailer
- ◯ Single family detached house
- ◯ Single family house attached to 1 or more houses (e.g townhouse/ semi-detached)
- ◯ High rise apartment/ condo/ co-op (4 or more floors)
- ◯ Low rise apartment/ condo/ co-op (1-3 floors)
- ◯ Multi-family home (more than one family in a converted single home)
- ◯ Other type
- ◯ Prefer not to answer

---

Does the child regularly stay in more than one home (since the third survey)?

- ◯ Yes
- ◯ No (Go to Question 53)

---

**52.7** Please provide the postal code of the child's secondary home

____________________

(Enter "555" if you would prefer not to answer.)

---

**52.8** How many adults currently live at the child's secondary home?

- ◯ 0
- ◯ 1
- ◯ 2
- ◯ 3
- ◯ 4
- ◯ 5
- ◯ 6
- ◯ 7
- ◯ 8
- ◯ 9
- ◯ 10
- ◯ Prefer not to answer

---

**52.9** How many children currently live at the child's secondary home (excluding the child)?

- ◯ 0
- ◯ 1
- ◯ 2
- ◯ 3
- ◯ 4
- ◯ 5
- ◯ 6
- ◯ 7
- ◯ 8
- ◯ 9
- ◯ 10
- ◯ Prefer not to answer

**52.10** How many bedrooms are in the child's secondary home?

- 0
- 1
- 2
- 3
- 4
- 5
- 6
- 7
- 8
- 9
- 10
- Prefer not to answer

**52.11** How many toilets are in the child's secondary home?

- 0
- 1
- 2
- 3
- 4
- 5
- 6
- 7
- 8
- 9
- 10
- Prefer not to answer

**52.12** Choose the type of dwelling that best describes the child's secondary home

- Manufactured home/ mobile home or trailer
- Single family detached house
- Single family house attached to 1 or more houses (e.g townhouse/ semi-detached)
- High rise apartment/ condo/ co-op (4 or more floors)
- Low rise apartment/ condo/ co-op (1-3 floors)
- Multi-family home (more than one family in a converted single home)
- Other type
- Prefer not to answer

**53.** Are any adults living in the home(s) considered to be an ESSENTIAL WORKER?

- Yes, I am
- Yes, another adult is
- No (Go to Question 54)
- Don't Know (Go to Question 54)
- Prefer not to answer (Go to Question 54)

((e.g., healthcare, delivery worker, store worker, security, building maintenance))

**53.1** If yes, do they come home each day?

- Yes, I do
- Yes, the other adult does
- No, separated due to COVID-19
- No separated due to other reasons
- Prefer not to answer

**53.2** If yes, are they a FIRST RESPONDER, HEALTHCARE PROVIDER or OTHER WORKER in a facility treating COVID-19?

- Yes, I am
- Yes, another adult is
- No
- Don't Know
- Prefer not to answer

**54.** Please share any comments, suggestions or clarifications that you have related to this survey

__________________________________________

**Study ID: _ _ _ _ _ C _**

**55.** What is your relationship to the child?

- ◯ Biological Mother
- ◯ Biological Father
- ◯ Parent
- ◯ Step Parent
- ◯ Foster Parent
- ◯ Grandparent
- ◯ Great-Grandparent
- ◯ Aunt or Uncle
- ◯ Sibling
- ◯ Step Sibling
- ◯ Cousin
- ◯ Other, Caregiver
- ◯ Prefer not to answer

**56.** Today's Date

______________________________ (DD/MM/YYYY)

### Adult Self Report Follow-up Survey 3

**Study ID: _ _ _ _ _ C _**

The following questions are being asked so we can better understand how COVID 19 has impacted the lives of families in the CHILD Study. If any of these questions are triggering for extreme concern about physical and/or mental health, resources can be found at the following link: http://www.bcchildrens.ca/our-services/mental-health-services/help-finding-resources. You can also contact our site at 604-875-2000 local 6390 to speak with a member of our team.

We are interested in learning more about how you are feeling and what you are thinking about during the Coronavirus (COVID-19) pandemic since you completed your third survey on ______________________ .
(DD/MM/YYYY)

COVID-19 is a new type of illness affecting many people in Canada. The virus that causes COVID-19 is related to the common cold virus. Some people affected by the virus may have symptoms that can be mild or severe, similar to a cold or flu. Other people with COVID-19 may not have any symptoms at all but they can spread the virus to others.

It is important for scientists to learn more about COVID-19. You can help us understand more about how COVID-19 affects families by answering this survey. Do the best you can to answer each question. This survey should take between 20 to 30 minutes to complete.

If you have questions or want help filling the survey out, please contact your site's Coordinator.

**1.** In general, how would you rate your overall PHYSICAL health now?

- ○ Excellent
- ○ Very Good
- ○ Good
- ○ Fair
- ○ Poor
- ○ Prefer not to answer

**2.** In general, how would you rate your overall MENTAL/EMOTIONAL health now?

- ○ Excellent
- ○ Very Good
- ○ Good
- ○ Fair
- ○ Poor
- ○ Prefer not to answer

**3.** What is your current weight in kilograms?

______________________________
(Enter "555" if you would prefer not to answer.)

#### COVID-19 HEALTH/EXPOSURE STATUS

**4.** In what ways has the COVID-19 outbreak affected your overall healthcare, since you completed the third survey on __________________? (Select all that apply)
(DD/MM/YYYY)

- ☐ I did not go to healthcare appointments because I was concerned about entering the healthcare providers office
- ☐ My healthcare provider cancelled appointments
- ☐ My healthcare provider changed to phone or online visits (Go to Question 5)
- ☐ My healthcare provider told me to self-isolate or quarantine (Go to Question 5)
- ☐ None of these apply (Go to Question 5)
- ☐ Prefer not to answer (Go to Question 5)

**4.1** What kind of healthcare visits were affected since you completed the third survey on __________________? (Select all that apply)
(DD/MM/YYYY)

- ☐ Routine visit/ regular checkup
- ☐ Vaccination visit
- ☐ Visit with a health care professional for a specific concern
- ☐ Visit with a specialist
- ☐ I did not miss any healthcare visits
- ☐ Prefer not to answer

**5.** Have you been exposed to someone likely to have COVID-19, since you completed the third survey on __________________? (Select all that apply)
(DD/MM/YYYY)

- ☐ Yes, to someone with positive test
- ☐ Yes, to someone with medical diagnosis, but no test
- ☐ Yes, to someone with possible symptoms, but no diagnosis by doctor
- ☐ No, not to my knowledge
- ☐ Prefer not to answer

**6.** Have you been suspected of having COVID-19, since you completed the third survey on __________________?
(DD/MM/YYYY)

- ☐ Yes, had positive test
- ☐ Yes, had medical diagnosis, but no test
- ☐ Yes, had some possible symptoms, but no diagnosis by health care professional
- ☐ No
- ☐ Prefer not to answer

**7.** Have you been tested for the virus that causes COVID-19, since you completed the third survey on __________________? (Select all that apply)
(DD/MM/YYYY)
Do not include the antibody/ serology test (blood test for previous infection) that is part of this reseach study.

This can include routine testing (e.g. for work, school, events, etc.)

- ☐ No, I never tried to get tested (Answer Question 7.3)
- ☐ No, I tried to get tested but was not able to (Answer Question 7.1)
- ☐ Yes, and I am waiting for the results (Answer Question 7.2)
- ☐ Yes, and the test showed that I did not have it ("negative" test) (Answer Question 7.2)
- ☐ Yes, and the test showed that I did have it ("positive" test) (Answer Question 7.2)
- ☐ Prefer not to answer (Answer Question 8)

**7.1** Which type of test for the virus that causes COVID-19 did you try to get but were not able to?

- ☐ Viral Test (swab test for current infection)
- ☐ Antibody/ Serology Test (blood test for previous infection)
- ☐ Saliva or Mouth Rinse Test
- ☐ Rapid-Antigen Test
- ☐ Other Specify______________________________
  (Enter "555" if you would prefer not to answer.)
- ☐ Don't Know
- ☐ Prefer not to answer

(Antibody/ Serology testing and saliva/ mouth rinse testing may not be currently available in all areas, but it may be available in the future.

**7.2** Please specify why you got tested or tried to get tested

- ☐ A contact or possible contact of mine tested positive for COVID-19
- ☐ I had COVID-19 symptoms
- ☐ I was admitted to hospital (unrelated to COVID-19)
- ☐ I was admitted to hospital (COVID-19 related)
- ☐ I was tested in another facility or institution context (contact with someone in long term care, prison etc)
- ☐ I was participating in another facility research study
- ☐ Travel-related testing
- ☐ Routine workplace testing (health care workplace)
- ☐ Routine workplace testing (non-health care workplace)
- ☐ Other, specify ______________________

  (Enter "555" if you would prefer not to answer.)
- ☐ Don't Know
- ☐ Prefer not to answer

(Go to Question 7.4)

**7.3** Please specify why you didn't try to get tested

- ☐ Symptoms were due to another condition, specify

  ______________________

  (Enter "555" if you would prefer not to answer.)
- ☐ Symptoms were mild
- ☐ A healthcare provider did not recommend testing
- ☐ Other, specify ______________________

  (Enter "555" if you would prefer not to answer.)
- ☐ Prefer not to answer

(Go to Question 7.9)

**7.4** IN THE PAST TWO WEEKS, how many times have you been tested for COVID-19?

- ○ 0 (Go to Question 8)
- ○ 1
- ○ 2
- ○ 3
- ○ 4
- ○ Prefer not to answer (Go to Question 8)

Details of First COVID-19 Test

**7.5** First test result

- ○ Negative
- ○ Positive
- ○ Don't know
- ○ Prefer not to answer

In the past two weeks have you been tested for COVID-19 a second time?

- ○ Yes
- ○ No (If you had a positive COVID test, go to Question 7.9. If not, go to Question 8)

Details of Second COVID-19 Test

**7.6** Second test result

- ○ Negative
- ○ Positive
- ○ Don't know
- ○ Prefer not to answer

In the past two weeks have you been tested for COVID-19 a third time?

- ○ Yes
- ○ No (If you had a positive COVID test, go to Question 7.9. If not, go to Question 8)

**Study ID:** _ _ _ _ _ **C** _

Details of Third COVID-19 Test

**7.7** Third test result

- ◯ Negative
- ◯ Positive
- ◯ Don't know
- ◯ Prefer not to answer

In the past two weeks have you been tested for COVID-19 a fourth time?

- ◯ Yes
- ◯ No (If you had a positive COVID test, go to Question 7.9. If not, go to Question 8)

Details of Fourth COVID-19 Test

**7.8** Fourth test result

- ◯ Negative
- ◯ Positive
- ◯ Don't know
- ◯ Prefer not to answer

**7.9** When people still show symptoms of COVID-19 for weeks or months after their initial recovery, it's called post or long COVID-19 condition. Post-COVID-19 can happen anywhere from 4 to 12 weeks after you first have COVID-19. Do you have post-COVID symptoms? (select all that apply)

- ☐ Tired/fatigue
- ☐ Memory problems
- ☐ Trouble sleeping or staying asleep
- ☐ Difficulty breathing
- ☐ Anxiety or depression
- ☐ Pain or discomfort anywhere in your body
- ☐ Difficulty thinking or concentrating
- ☐ Post-traumatic stress disorder (PTSD)
- ☐ Prefer not to answer

**8.** Have you had any of the following symptoms, since you completed the third survey on ____________________? (Select all that apply)
(DD/MM/YYYY)

- ☐ New or worsening cough
- ☐ Shortness of breath
- ☐ Difficulty breathing
- ☐ Fever
- ☐ Chills
- ☐ Muscle or body aches
- ☐ Headache
- ☐ Sore throat
- ☐ New loss of sense of smell or taste
- ☐ Runny nose or nasal congestion*
- ☐ Nausea or vomiting
- ☐ Diarrhea
- ☐ Fatigue
- ☐ Skin Rash
- ☐ Conjunctivitis (pink eye)
- ☐ Other, Specify ______________________________
  (Enter "555" if you would prefer not to answer.)

- ☐ None of the above (Go to Question 9)
- ☐ Prefer not to answer (Go to Question 9)

(*Runny nose or nasal congestion in the absence of underlying reason for these symptoms such as seasonal allergies, post nasal drip, etc)

**8.1** In the TWO WEEKS, before you had symptoms, did you: (Select all that apply)

- ☐ Have contact with someone who tested positive for COVID-19 (Go to Question 8.7)
- ☐ Have contact with someone who likely had COVID-19 (e.g., was not tested but had symptoms; was told by a healthcare provider that he/she likely had it) (Go to Question 8.7)
- ☐ Travel outside the province
- ☐ None of the above (Go to Question 8.7)
- ☐ Prefer not to answer (Go to Question 8.7)

**8.2** How many places did you travel?

- ○ 0 (Go to Question 8.7)
- ○ 1
- ○ 2
- ○ 3
- ○ 4
- ○ Prefer not to answer (Go to Question 8.7)

(Travel outside the province)

Place 1

**8.3** Location
(e.g. Seattle, Washington OR Toronto, Ontario)

______________________
(Enter "555" if you would prefer not to answer.)

Did the you travel to a second place?

- ○ Yes
- ○ No (Go to Question 8.7)

Place 2

**8.4** Location
(e.g. Portland, Oregon OR Vancouver, BC)

______________________
(Enter "555" if you would prefer not to answer.)

Did the you travel to a third place?

- ○ Yes
- ○ No (Go to Question 8.7)

Place 3

**8.5** Location
(e.g. Paris, France OR Banff, Alberta)

______________________
(Enter "555" if you would prefer not to answer.)

Did the you travel to a fourth place?

- ○ Yes
- ○ No (Go to Question 8.7)

Place 4

**8.6** Location
(e.g. Portland, Oregon OR Vancouver, BC)

______________________
(Enter "555" if you would prefer not to answer.)

**8.7** Which of the following occurred as a result of your symptoms? (Select all that apply)

- ☐ You spoke to a healthcare provider over the phone, by email, or online
- ☐ You saw a healthcare provider in person, such as in a clinic or doctor's office
- ☐ You saw a healthcare provider in person, such as in urgent care, or in an Emergency Department (ED)
- ☐ You were kept overnight in a hospital because a healthcare provider thought you had COVID-19
- ☐ You were admitted to a hospital's ICU (Intensive Care Unit)
- ☐ You self-isolated or quarantined at home
- ☐ None of the above
- ☐ Prefer not to answer

**9.** Has anyone in your family been diagnosed with COVID-19, since you completed the third survey on ___________________? (DD/MM/YYYY)
(Select all that apply)

- ☐ Yes, member of household
- ☐ Yes, non-household member
- ☐ No
- ☐ Don't know
- ☐ Prefer not to answer

**Study ID: _ _ _ _ _ C _**

| | |
|---|---|
| **10.** Have any of the following happened to family members living inside your household because of COVID-19, since you completed the third survey on ____________________? (DD/MM/YYYY) | ☐ Fallen physically ill<br>☐ Hospitalized<br>☐ Self-isolated or quarantined with symptoms<br>☐ Self-isolated or quarantined without symptoms (e.g., due to possible exposure)<br>☐ Lost job or been laid off from job<br>☐ Reduced ability to earn money<br>☐ Passed away<br>☐ None of the above<br>☐ Prefer not to answer |
| **11.** Have any of the following happened to family members living outside your household because of COVID-19, since you completed the third survey on ____________________? (DD/MM/YYYY) | ☐ Fallen physically ill<br>☐ Hospitalized<br>☐ Self-isolated or quarantined with symptoms<br>☐ Self-isolated or quarantined without symptoms (e.g., due to possible exposure)<br>☐ Lost job or been laid off from job<br>☐ Reduced ability to earn money<br>☐ Passed away<br>☐ None of the above<br>☐ Prefer not to answer |
| **12.** Did you get the flu shot, since you completed the third survey on ____________________? (DD/MM/YYYY) | ◯ Yes<br>◯ No<br>◯ Don't Know<br>◯ Prefer not to answer |
| **13.** Have you received at least one dose of a COVID-19 vaccine? | ◯ Yes<br>◯ No (Go to Question 13.12)<br>◯ Prefer not to answer (Go to Question 14)<br>(Answer 'Yes' if you have received at least one dose of the COVID-19 vaccine. Note: Certain types of vaccines require more than one dose to protect against COVID-19. You would have been informed at the time of vaccination if you needed a second dose. Some groups of people are also now eligible to receive a third booster shot.) |
| **13.1** Did you feel any side effects from the COVID-19 vaccine? | ◯ Yes<br>◯ No (Go to Question 13.3)<br>◯ Prefer not to answer(Go to Question 13.3) |
| **13.2** If yes, check all that apply. | ☐ Head ache<br>☐ Tiredness<br>☐ Fever or chills<br>☐ Muscle or joint pain<br>☐ Soreness where the vaccine was injected<br>☐ Nausea or vomiting<br>☐ Other, specify______________________________<br>(Enter "555" if you would prefer not to answer.)<br>☐ Prefer not to answer |

**13.3** How many doses of the COVID-19 vaccine have you received so far?

- ◯ One dose
- ◯ Two doses
- ◯ Three doses
- ◯ Four doses
- ◯ More than four doses
- ◯ Prefer not to answer (Go to Question 14)

**13.4** When did you receive your first dose of the COVID-19 vaccine?

______________________ (DD/MM/YYYY)
(If you do not recall the exact date, please provide an estimate. Enter "01-01-1900" if you would prefer not to answer.)

**13.5** Which vaccine did you receive? (first dose)

- ◯ Pfizer and BioNTech mRNA vaccine
- ◯ Moderna mRNA vaccine
- ◯ AstraZeneca Oxford vaccine
- ◯ Janssen (Johnson & Johnson) vaccine
- ◯ Other, specify______________________
  (Enter "555" if you would prefer not to answer.)
- ◯ Don't Know
- ◯ Prefer not to answer

If you only received 1 dose of the COVID-19 vaccine, go to Question 14.
If you received more than 1 dose of the COVID-19 vaccine, continue to answer Question 13.6.

**13.6** When did you receive your second dose of the COVID-19 vaccine?

______________________ (DD/MM/YYYY)
(If you do not recall the exact date, please provide an estimate. Enter "01-01-1900" if you would prefer not to answer.)

**13.7** Which vaccine did you receive? (second dose)

- ◯ Pfizer and BioNTech mRNA vaccine
- ◯ Moderna mRNA vaccine
- ◯ AstraZeneca Oxford vaccine
- ◯ Janssen (Johnson & Johnson) vaccine
- ◯ Other, specify______________________
  (Enter "555" if you would prefer not to answer.)
- ◯ Don't Know
- ◯ Prefer not to answer

If you only received 2 doses of the COVID-19 vaccine, go to Question 14.
If you received more than 2 doses of the COVID-19 vaccine, continue to answer Question 13.8.

**13.8** When did you receive your third dose of the COVID-19 vaccine?

______________________ (DD/MM/YYYY)
(If you do not recall the exact date, please provide an estimate. Enter "01-01-1900" if you would prefer not to answer.)

**13.9** Which vaccine did you receive? (third dose)

- ◯ Pfizer and BioNTech mRNA vaccine
- ◯ Moderna mRNA vaccine
- ◯ AstraZeneca Oxford vaccine
- ◯ Janssen (Johnson & Johnson) vaccine
- ◯ Other, specify______________________
  (Enter "555" if you would prefer not to answer.)
- ◯ Don't Know
- ◯ Prefer not to answer

If you only received 3 doses of the COVID-19 vaccine, go to Question 14.
If you received more than 3 doses of the COVID-19 vaccine, continue to answer Question 13.10.

**13.10** When did you receive your fourth dose of the COVID-19 vaccine?

______________________ (DD/MM/YYYY)
(If you do not recall the exact date, please provide an estimate. Enter "01-01-1900" if you would prefer not to answer.)

**13.11** Which vaccine did you receive? (fourth dose)

- ◯ Pfizer and BioNTech mRNA vaccine
- ◯ Moderna mRNA vaccine
- ◯ AstraZeneca Oxford vaccine
- ◯ Janssen (Johnson & Johnson) vaccine
- ◯ Other, specify______________________
  (Enter "555" if you would prefer not to answer.)
- ◯ Don't Know
- ◯ Prefer not to answer

(Go to Question 14)

**13.12** When a COVID-19 vaccine becomes available, how likely is it that you will choose to get it?

- ◯ Very Likely (Go to Question 14)
- ◯ Somewhat Likely (Go to Question 14)
- ◯ Somewhat Unlikely
- ◯ Very Unlikely
- ◯ Don't Know
- ◯ Prefer not to answer (Go to Question 14)

**13.13** What are some of the reasons why you don't want to get the COVID-19 vaccine?

- ☐ Not confident in the safety of the vaccine
- ☐ Concern about risks and side effects
- ☐ Will wait until it seems safe to get the vaccine
- ☐ Do not consider it necessary to get the vaccine
- ☐ Do not believe in vaccination
- ☐ Have not yet decided
- ☐ Have a pre-existing medical condition
- ☐ Already had or think I have had COVID-19
- ☐ Other, Specify______________________
  (Enter "555" if you would prefer not to answer.)
- ☐ Prefer not to answer

(Check all that apply.)

**14.** Where do you normally look or go to for information on the COVID-19 vaccine? (select all that apply)

- ☐ Family members
- ☐ Family doctor/pediatrician
- ☐ Provincial government websites
- ☐ Government of Canada websites (example: PHAC, Health Canada)
- ☐ Other internet sites
- ☐ Social media/networking (examples: TikTok, Twitter, Facebook, Instagram)
- ☐ Religious leaders
- ☐ Friends
- ☐ Co-workers
- ☐ School/School Teachers
- ☐ Other, Specify ______________________________

- ☐ None of these places
- ☐ Prefer not to answer

| | Never | Rarely | Occasionally | Often | Always | Don't Know | Prefer not to answer |
|---|---|---|---|---|---|---|---|
| **15.** How often have you worn a mask in public places since you completed your third survey on ____________________ ? (DD/MM/YYYY) | ○ | ○ | ○ | ○ | ○ | ○ | ○ |
| **16.** How often have you practiced physical distancing in public places since you completed your third survey on ____________________? (DD/MM/YYYY) | ○ | ○ | ○ | ○ | ○ | ○ | ○ |
| **17.** How often have you avoided crowded places/gatherings since you completed your third survey on ____________________? (DD/MM/YYYY) | ○ | ○ | ○ | ○ | ○ | ○ | ○ |
| **18.** How often have you avoided common greetings (e.g handshakes, hugs etc) since you completed your third survey on ____________________? (DD/MM/YYYY) | ○ | ○ | ○ | ○ | ○ | ○ | ○ |
| **19.** How often have you limited contact with people at higher risk (e.g. elderly relative) since you completed your third survey on ____________________ ? (DD/MM/YYYY) | ○ | ○ | ○ | ○ | ○ | ○ | ○ |

**20.** How often have you self-Isolated because someone thought you were infected with the virus that causes COVID-19 since you completed your third survey on ____________________ ?
(DD/MM/YYYY)

○ ○ ○ ○ ○ ○ ○

---

**21.** How often have you quarantined because you may have been exposed to the virus that causes COVID-19, but did not show symptoms since you completed your third survey on ________________________ ?
(DD/MM/YYYY)

○ ○ ○ ○ ○ ○ ○

---

**22.** Have you participated in any organized sports or recreational activities, in person, since you completed the third survey on ____________________? (DD/MM/YYYY)

- ○ Yes
- ○ No (Go to Question 23)
- ○ Prefer not to answer (Go to Question 23)

---

**22.1** If yes, how many people were in the activity?

- ○ 0-9
- ○ 10-19
- ○ 20-29
- ○ 30+
- ○ Prefer not to answer

---

**23.** How many times have you been in a gathering of > 10 or more people (excluding school or work), since you completed the third survey on ____________________?
(DD/MM/YYYY)

_______________________________
(A gathering in this context is defined as any assembly or meeting of more than 10 persons, in open air or in a building or premises. These can include gatherings with or without masks or social distancing. Gatherings include a range of interpersonal gatherings/events of varying purposes, from gatherings/events of family and friends (e.g., sports, weddings, funerals, baptisms, birthday parties and cultural ceremonies) to community gathering spaces (e.g., places of worship, places of traditional gatherings, libraries, community and recreation centres, camps, drop-in centres, food banks and communal kitchens, museums, theatres, cinemas, and tourist attractions) to larger planned or structured gatherings/events, including mass gatherings (e.g., large meetings or conferences, national sporting events, large cultural/religious events, festivals). Enter "555" if you would prefer not to answer.)

**Study ID: _ _ _ _ _ C _**

**24. The following are a number of mental health services that you may have accessed. Please check the relevant boxes for each service indicating if you have accessed the service since you completed the third survey on ____________________ or if you would be interested in accessing the service.**
(DD/MM/YYYY)

| | Have accessed this service since completing the third survey on ________________ (DD/MM/YYYY) | Would be interested in accessing this service | Not accessed, not interested | Prefer not to answer |
|---|---|---|---|---|
| Individual therapy (you alone) with a professional (in person or virtual) | ○ | ○ | ○ | ○ |
| Group therapy (you and others) with a professional (in person or virtual) | ○ | ○ | ○ | ○ |
| Couples therapy (you and your partner) with a professional (in person or virtual) | ○ | ○ | ○ | ○ |
| Family therapy (you and your family members) with a professional (in person or virtual) | ○ | ○ | ○ | ○ |
| Mental health mobile app (general) | ○ | ○ | ○ | ○ |
| Mental health mobile app to interact with a therapist | ○ | ○ | ○ | ○ |
| Phone helpline for mental health support (e.g. Crisis Services Canada) | ○ | ○ | ○ | ○ |
| Online mental health information you looked up yourself | ○ | ○ | ○ | ○ |

**Study ID: _ _ _ _ _ C _**

#### Worrying (PAST TWO WEEKS)

| | Not at all worried | Slightly worried | Moderately worried | Very worried | Extremely worried | Prefer not to answer |
|---|---|---|---|---|---|---|
| **25.** During the PAST TWO WEEKS, how worried have you been about being infected by the virus that causes COVID-19? | ○ | ○ | ○ | ○ | ○ | ○ |
| **26.** During the PAST TWO WEEKS, how worried have you been about friends or family being infected by the virus that causes COVID-19? | ○ | ○ | ○ | ○ | ○ | ○ |
| **27.** During the PAST TWO WEEKS, how worried have you been about your physical health being influenced by COVID-19? | ○ | ○ | ○ | ○ | ○ | ○ |
| **28.** During the PAST TWO WEEKS, how worried have you been about your mental/emotional health being influenced by COVID-19? | ○ | ○ | ○ | ○ | ○ | ○ |
| **29.** During the PAST TWO WEEKS, how worried were you generally? | ○ | ○ | ○ | ○ | ○ | ○ |

**30.** How much are you reading or talking about COVID-19, since you completed the third survey on ____________________?
(DD/MM/YYYY)

- ○ Never
- ○ A few times a month
- ○ Weekly
- ○ A few times a week
- ○ Daily
- ○ Prefer not to answer

#### LIFE CHANGES DUE TO THE COVID-19 PANDEMIC

| | |
|---|---|
| **31.** Based on what you know at this time, how safe do you think it is for students in your community to attend school in-person? | ◯ Very safe<br>◯ Somewhat safe<br>◯ Not very safe<br>◯ Not safe at all<br>◯ Don't Know<br>◯ Prefer not to answer |
| **32.** Since you completed the third survey on ____________________, (DD/MM/YYYY) how difficult has the cancellation of important events in your life (such as weddings, vacations, etc.) been for you? | ◯ Not difficult<br>◯ A little bit difficult<br>◯ Difficult<br>◯ Very difficult<br>◯ Extremely difficult<br>◯ There hasn't been any cancellation of important events<br>◯ Prefer not to answer |
| **33.** During the PAST TWO WEEKS, how many people (from outside of your household) have you had an in-person conversation with in an indoor environment?<br><br>This can include things like work, school or other activities with or without masks and/or social distancing. If you aren't sure, please estimate a number. | ◯ 0<br>◯ Less than 10<br>◯ 10 - 19<br>◯ 20 - 29<br>◯ 30 or more<br>◯ Prefer not to answer<br>(In-person means that the person is in the same place as you (NOT on the phone or video chat)) |
| **34.** During the PAST TWO WEEKS, how many people (from outside of your household) have you had an in-person conversation with in an outdoor environment?<br><br>This can include things like work, school or other activities with or without masks and/or social distancing. If you aren't sure, please estimate a number | ◯ 0<br>◯ Less than 10<br>◯ 10 - 19<br>◯ 20 - 29<br>◯ 30 or more<br>◯ Prefer not to answer<br>(In-person means that the person is in the same place as you (NOT on the phone or video chat)) |
| **35.** During the PAST TWO WEEKS, how many times did you go out outside of your home (e.g., going to stores, parks, etc.)? | ◯ Not at all<br>◯ 1-2 days per week or less<br>◯ 3-4 days per week<br>◯ 5-6 days per week<br>◯ Daily<br>◯ Prefer not to answer |
| **36.** During the PAST TWO WEEKS, how stressful have the restrictions on leaving home been for you? | ◯ Not stressful<br>◯ A little bit stressful<br>◯ Stressful<br>◯ Very stressful<br>◯ Extremely stressful<br>◯ There are no restrictions on leaving home<br>◯ Prefer not to answer |

| | |
|---|---|
| **37.** During the PAST TWO WEEKS, how difficult has it been for you to follow the recommendations for keeping away from close contact with people who you don't live with? | ◯ Not difficult<br>◯ A little bit difficult<br>◯ Difficult<br>◯ Very difficult<br>◯ Extremely difficult<br>◯ There are no recommendations for keeping away from close contact with people who I don't live with<br>◯ Prefer not to answer |
| **38.** During the PAST TWO WEEKS, to what degree have changes related to the COVID-19 pandemic in your area created financial problems for your family? | ◯ No financial problems<br>◯ A few financial problems<br>◯ Some financial problems<br>◯ A lot of financial problems<br>◯ Extreme financial problems<br>◯ Prefer not to answer |
| **39.** During the PAST TWO WEEKS, to what degree are you concerned about the stability of your living situation? | ◯ Not at all concerned<br>◯ A little concerned<br>◯ Concerned<br>◯ Very concerned<br>◯ Extremely concerned<br>◯ Prefer not to answer<br>(Stability of your living situation refers to forced moves that are the result of poor housing quality, unstable neighbourhood conditions and high costs of housing in relation to income. ) |
| **40.** During the PAST TWO WEEKS, did you worry your food would run out because of a lack of money? | ◯ Yes<br>◯ No<br>◯ Prefer not to answer |
| **41.** How hopeful are you that the COVID-19 pandemic in your area will end soon? | ◯ Extremely hopeful<br>◯ Very hopeful<br>◯ Hopeful<br>◯ A little bit hopeful<br>◯ Not at all hopeful<br>◯ There is no COVID-19 pandemic in my area<br>◯ Prefer not to answer |

#### DAILY BEHAVIORS (Past TWO WEEKS)

**42.** During the PAST TWO WEEKS, what time did you go to bed on WEEKDAYS (on average)?

- ◯ Before 8 pm
- ◯ 8 pm - 10 pm
- ◯ 10 pm - 12 am (Midnight)
- ◯ After 12 am (Midnight)
- ◯ I slept during daytime hours (example: shift or on-call work)
- ◯ Prefer not to answer

**43.** During the PAST TWO WEEKS, what time did you go to bed on WEEKENDS (on average)?

- ◯ Before 8 pm
- ◯ 8 pm - 10 pm
- ◯ 10 pm - 12 am (Midnight)
- ◯ After 12 am (Midnight)
- ◯ I slept during daytime hours (example: shift or on-call work)
- ◯ Prefer not to answer

**44.** During the PAST TWO WEEKS, how many hours per night did you sleep on WEEKDAYS (on average)?

- ◯ Less than 6 hours
- ◯ 6-8 hours
- ◯ 8-10 hours
- ◯ Greater than 10 hours
- ◯ Prefer not to answer

**45.** During the PAST TWO WEEKS, how many hours per night did you sleep on WEEKENDS (on average)?

- ◯ Less than 6 hours
- ◯ 6-8 hours
- ◯ 8-10 hours
- ◯ Greater than 10 hours
- ◯ Prefer not to answer

**46.** During the PAST TWO WEEKS, how many days per week did you exercise (e.g., increased heart rate, breathing)for at least 30 minutes over the entire day?

- ◯ Not at all
- ◯ 1-2 days per week or less
- ◯ 3-4 days per week
- ◯ 5-6 days per week
- ◯ Daily
- ◯ Prefer not to answer

**47.** During the PAST TWO WEEKS, how many days per week did you spend time outdoors?

- ◯ Not at all
- ◯ 1-2 days per week or less
- ◯ 3-4 days per week
- ◯ 5-6 days per week
- ◯ Daily
- ◯ Prefer not to answer

**48.** How has the COVID-19 outbreak affected your regular childcare, since you completed the third survey on ____________________? (DD/MM/YYYY)
(Select all that apply)

- ☐ I had difficulty arranging for childcare
- ☐ I had to pay more for childcare
- ☐ My spouse/partner or I had to change our work schedule to care for our children ourselves
- ☐ My spouse/partner or I had to reduce our paid work hours to care for our children ourselves
- ☐ My regular childcare has not been affected by the COVID-19 outbreak
- ☐ I do not have a child in childcare
- ☐ Prefer not to answer

#### EMOTIONS/WORRIES (PAST TWO WEEKS)

**49.** During the PAST TWO WEEKS, how happy versus sad were you?

- Very happy/cheerful
- Moderately happy/cheerful
- Neutral
- Moderately sad/depressed/unhappy
- Very sad/depressed/unhappy
- Prefer not to answer

**50.** During the PAST TWO WEEKS, how relaxed versus anxious were you?

- Very relaxed/calm
- Moderately relaxed/calm
- Neutral
- Moderately nervous/anxious
- Very nervous/anxious
- Prefer not to answer

**51.** During the PAST TWO WEEKS, how fidgety or restless were you?

- Not fidgety/restless at all
- Slightly fidgety/restless
- Moderately fidgety/restless
- Very fidgety/restless
- Extremely fidgety/restless
- Prefer not to answer

**52.** During the PAST TWO WEEKS, how tired were you?

- Not tired at all
- Slightly tired
- Moderately tired
- Very tired
- Extremely tired
- Prefer not to answer

**53.** During the PAST TWO WEEKS, how well were you able to concentrate or focus?

- Very focused/attentive
- Moderately focused/attentive
- Neutral
- Moderately unfocused/distracted
- Very unfocused/distracted
- Prefer not to answer

**54.** During the PAST TWO WEEKS, how irritable or easily angered were you?

- Not irritable or easily angered at all
- Slightly irritable or easily angered
- Moderately irritable or easily angered
- Very irritable or easily angered
- Extremely irritable or easily angered
- Prefer not to answer

**55.** During the PAST TWO WEEKS, how lonely were you?

- Not lonely at all
- Slightly lonely
- Moderately lonely
- Very lonely
- Extremely lonely
- Prefer not to answer

**56.** What have been your greatest sources of stress from the COVID-19 outbreak, since you completed the third survey on____________________? (Select all that apply) (DD/MM/YYYY)

- ☐ Health concerns
- ☐ Financial concerns
- ☐ Impact on work
- ☐ Impact on your child
- ☐ Impact on your community
- ☐ Impact on family members
- ☐ Impact on the economy
- ☐ Access to food
- ☐ Access to baby supplies (e.g., formula, diapers, wipes)
- ☐ Access to personal care products or household supplies
- ☐ Access to medical care, including mental health care
- ☐ Social distancing or being quarantined
- ☐ Other
- ☐ I am not stressed about the COVID-19 outbreak
- ☐ Prefer not to answer

**57.** What have you done to cope with your stress related to the COVID-19 outbreak, since you completed the third survey on____________________? (DD/MM/YYYY) (Select all that apply)

- ☐ Meditation and/or mindfulness practices
- ☐ Talking with friends and family (e.g., by phone, text, or video)
- ☐ Engaging in more family activities (e.g., games, sports)
- ☐ Increased television watching or other screen time activities (e.g., video games, social media)
- ☐ Eating more often, including snacking
- ☐ Increasing time reading books, or doing activities like puzzles and crosswords
- ☐ Drinking alcohol
- ☐ Using tobacco (e.g., smoking, vaping)
- ☐ Using marijuana / cannabis (e.g., vaping, smoking, eating) or cannabidiol (CBD)
- ☐ Talking to my healthcare providers more frequently, including mental healthcare provider (e.g., therapist, psychologist, counselor)
- ☐ Volunteer work
- ☐ Exercise
- ☐ Other
- ☐ I have not done any of these things to cope with the stress related to the COVID-19 outbreak
- ☐ I am not stressed about the COVID-19 outbreak
- ☐ Prefer not to answer

#### DEMOGRAPHIC INFORMATION

**58.** Has your living situation changed since you completed your third survey on ________________?
(DD/MM/YYYY)

- ☐ No, my living situation has not changed (Go to Question 59)
- ☐ Yes, I have moved (since the third survey)
- ☐ Yes, I regularly stay in more than one home (since the third survey)
- ☐ Yes, the number of people living in my home has changed (since the third survey) (Go to Question 58.2)
- ☐ Prefer not to answer (Go to Question 59)

**58.1** Please provide the postal code of your primary home

____________________________
(Enter "555" if you would prefer not to answer.)

**58.2** How many adults currently live at your primary home (excluding yourself)?

- ◯ 0
- ◯ 1
- ◯ 2
- ◯ 3
- ◯ 4
- ◯ 5
- ◯ 6
- ◯ 7
- ◯ 8
- ◯ 9
- ◯ 10
- ◯ Prefer not to answer

**58.3** How many children currently live at your primary home?

- ◯ 0
- ◯ 1
- ◯ 2
- ◯ 3
- ◯ 4
- ◯ 5
- ◯ 6
- ◯ 7
- ◯ 8
- ◯ 9
- ◯ 10
- ◯ Prefer not to answer

**58.4** How many bedrooms are in your primary home?

- ◯ 0
- ◯ 1
- ◯ 2
- ◯ 3
- ◯ 4
- ◯ 5
- ◯ 6
- ◯ 7
- ◯ 8
- ◯ 9
- ◯ 10
- ◯ Prefer not to answer

**58.5** How many toilets are in your primary home?

- ◯ 0
- ◯ 1
- ◯ 2
- ◯ 3
- ◯ 4
- ◯ 5
- ◯ 6
- ◯ 7
- ◯ 8
- ◯ 9
- ◯ 10
- ◯ Prefer not to answer

**58.6** Choose the type of dwelling that best describes your primary home

- ◯ Manufactured home/ mobile home or trailer
- ◯ Single family detached house
- ◯ Single family house attached to 1 or more houses (e.g townhouse/ semi-detached)
- ◯ High rise apartment/ condo/ co-op (4 or more floors)
- ◯ Low rise apartment/ condo/ co-op (1-3 floors)
- ◯ Multi-family home (more than one family in a converted single home)
- ◯ Other type
- ◯ Prefer not to answer

Do you regularly stay in more than one home (since the third survey)?

- ◯ Yes
- ◯ No (Go to Question 59)

**58.7** Please provide the postal code of your secondary home

______________________

(Enter "555" if you would prefer not to answer.)

**58.8** How many adults currently live at your secondary home (excluding yourself)?

- ◯ 0
- ◯ 1
- ◯ 2
- ◯ 3
- ◯ 4
- ◯ 5
- ◯ 6
- ◯ 7
- ◯ 8
- ◯ 9
- ◯ 10
- ◯ Prefer not to answer

**58.9** How many children currently live at yor secondary home?

- ◯ 0
- ◯ 1
- ◯ 2
- ◯ 3
- ◯ 4
- ◯ 5
- ◯ 6
- ◯ 7
- ◯ 8
- ◯ 9
- ◯ 10
- ◯ Prefer not to answer

**58.10** How many bedrooms are in your secondary home?

- ○ 0
- ○ 1
- ○ 2
- ○ 3
- ○ 4
- ○ 5
- ○ 6
- ○ 7
- ○ 8
- ○ 9
- ○ 10
- ○ Prefer not to answer

**58.11** How many toilets are in your secondary home?

- ○ 0
- ○ 1
- ○ 2
- ○ 3
- ○ 4
- ○ 5
- ○ 6
- ○ 7
- ○ 8
- ○ 9
- ○ 10
- ○ Prefer not to answer

**58.12** Choose the type of dwelling that best describes your secondary home

- ○ Manufactured home/ mobile home or trailer
- ○ Single family detached house
- ○ Single family house attached to 1 or more houses (e.g townhouse/ semi-detached)
- ○ High rise apartment/ condo/ co-op (4 or more floors)
- ○ Low rise apartment/ condo/ co-op (1-3 floors)
- ○ Multi-family home (more than one family in a converted single home)
- ○ Other type
- ○ Prefer not to answer

**59.** Please share any comments, suggestions or clarifications that you have related to this survey

______________________________

**60.** Did **you** complete this survey on your own?

Note: **You** refers to the adult participating in the study, who was assigned this survey

- ○ Yes - by myself
- ○ Yes - partially or with help from someone else
- ○ No - someone else completed this survey on my behalf

**61.** Today's Date

______________________________ (DD/MM/YYYY)
