## Appendix 2. for "Investigating SARS-CoV-2 infection and the health and psychosocial impact of the COVID-19 pandemic in the Canadian CHILD Cohort: study methodology and cohort profile"

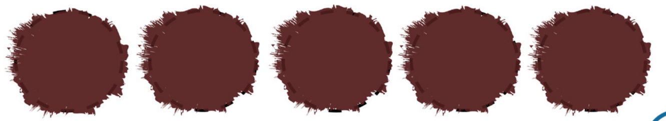

DO

Too little    Not Dry    Too much    Overlay

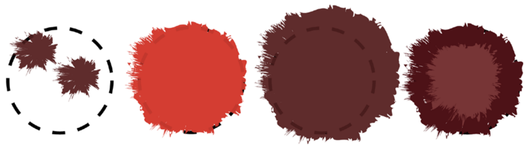

DO NOT

- Multiple small drops of blood can be dropped onto the same circle, but don't overlay them (drop them on top of the other).
- Avoid excessive "milking" or squeezing the area around the puncture site.
- Avoid touching or smearing spots.
- When completed, place a sterile gauze against your finger until the bleeding stops, then apply the bandage.

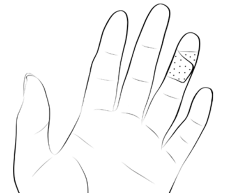

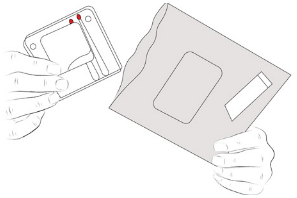

- Ensure the cartridge is closed. Insert cartridge into the silver specimen bag,containing the desiccant. Seal the bag.

**F. Dry the sample.**

- Let the blood spot card dry for at least **3 hours**: using the cover flap of the card, prop the blood spots up and place the card face up on a clean surface away from direct heat or sunlight; avoid touching it or knocking it over.
- Do not refrigerate.
- Once the card is dried, fold the paper flap to cover the spots. Place the dried card in the yellow specimen bag containing the desiccant and humidity card. Seal the bag.

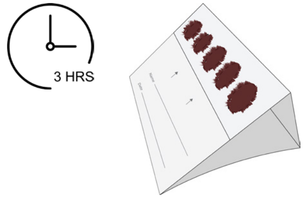


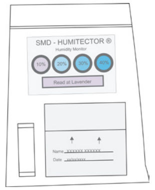


#### 4 Record Sample Information

On the card provided, record the date and time of sample collection, and any reasons a sample might not have been collected.
Visit https://childstudy.ca/covid-study/ (password = CHILDcv19!) to submit the sample collection information online.
**Return the card with your samples.**

#### 5 Return Samples

Place the safety bag (with the stool collection tube)and the 2 specimen bags (1 with the blood spot card, 1 with the Mitra cartridge) into the box you received (do not use the outer sleeve).

You do not need to recap used lancets. Return the lancets for proper disposal. Place them in the bag that the gauze and bandage came in and put that bag in the box with the samples. If you can, seal the box tightly with tape.

Ship the samples to the lab as soon as possible, ideally within **48 hours** of collection.

Using the pre-affixed and prepaid shipping label provided, schedule a FedEx pickup by calling 1-800-463-3339 or drop off your box at a FedEx center (https://fedex.com/locate). Do not cover or modify the shipping label or any other markings on the box.

If your sample kit DOES NOT HAVE a shipping label, please contact.

Thank you from the CHILD Cohort Study!

THE MICROSETTA INITIATIVE

Sample collection kits designed by The Microsetta Initiative. https://microsetta.ucsd.edu

**WARNING:**
**CHOKING HAZARD**
Small Parts, Adult Supervision Required.

FOR RESEARCH USE ONLY. NOT FOR USE IN DIAGNOSTIC PROCEDURES.

CHILD COHORT STUDY

### COVID-19 Add-on Study

Instructions for sample collection. This collection kit contains:

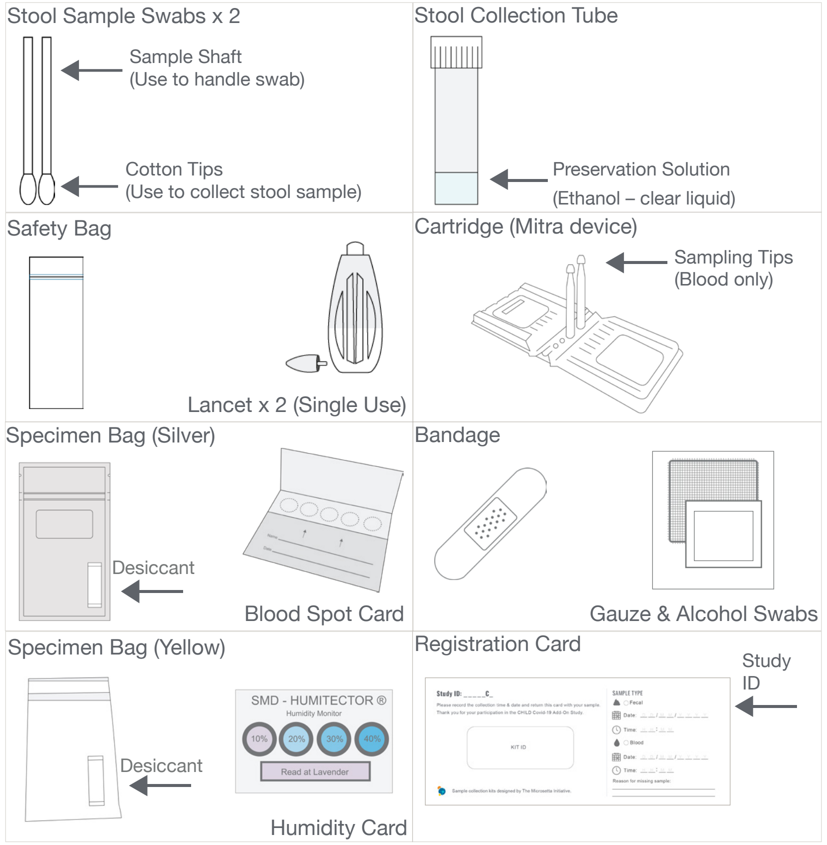


#### 1 Read Instructions and Watch Videos

Please read all the instructions (front to back) before collecting samples. Visit https://childstudy.ca/covid-study/ (password = CHILDcv19!) for more details and to watch a demonstration video. If you have any questions, please

Child-V3.indd 1    1/27/21 2:21 PM

#### 2 Select Sample Kit

Each kit has a unique ID and is for one person. Your household should receive one kit per participant. Each participant should select a kit and print, on the card provided, the unique study ID number they were assigned by research staff at recruitment.

#### 3 Collect Samples

This study requires three types of samples: stool and two types of blood samples. Collect your stool sample first.
Store samples at room temperature. Ideally, kits should be returned via FedEx within **48 hours** of sample collection.

##### 1. Stool Sample

Place the collection tube in the holder in the lower left-hand corner of the box. Open the sample swabs. Handle the swab by the shaft only; **avoid touching the cotton tips.**

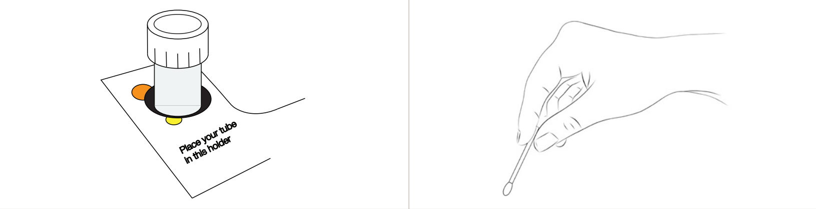


Collect your stool sample from a used piece of toilet paper (the first piece of paper used to wipe is best). Rub the cotton tip of the swab over the stool on the toilet paper. Repeat for the second swab.

**DO**

- Saturate at least half of the cotton tips. Do not oversaturate.
- If your stool is firm, rub enough on both sides of the cotton tips to get a light smear.
- If there is not enough stool to swab from toilet paper, deposit (poop) a small piece of stool directly onto the paper.

**DO NOT**

- Do not provide excess material.
- Do not collect any urine in your sample.
- Do not sample stool that has fallen into the toilet bowl.

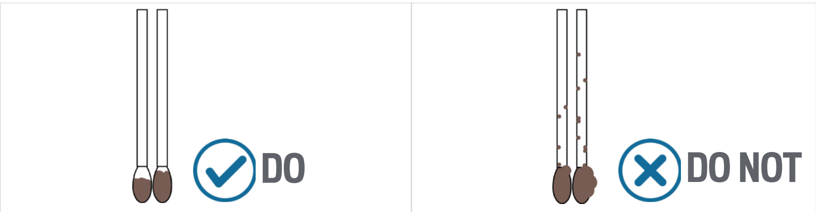


After collecting the samples, place the swabs (tips downward) into the collection tube with the preservation solution.
Tighten the cap onto the tube, ensuring no contents can leak out.
Remove the tube from the box, place it in the safety bag provided, and seal the bag. Wash your hands.

##### 2. Blood Samples

Blood samples are collected by two methods: cartridge and blood spot card. A single finger prick should give enough blood for both types of collection. If not, a second lancet is provided for a second finger prick

*** Do not leave any sample collection items around children**

**A. Select a puncture site.**

- Select a finger. Middle or ring finger is best. Do not use a thumb or pinky.
- You should use the side of your finger just below the tip.
- The puncture should occur across the fingerprints, not fall in a line parallel to them.

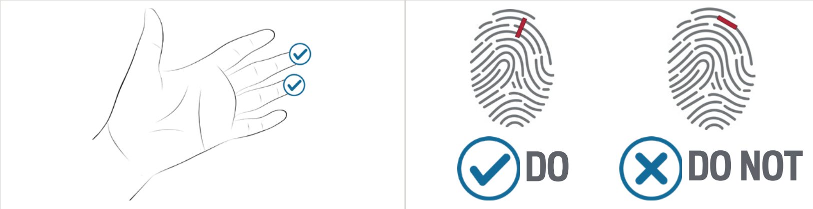


**B. Prepare to collect the blood samples.**

- Wash your hands thoroughly for at least 20 seconds. Dry completely.
- Sit comfortably with the kit in front of you.
- Warm your hands to increase blood flow by rubbing them together.
- Cleanse the selected site with an alcohol swab and air dry.

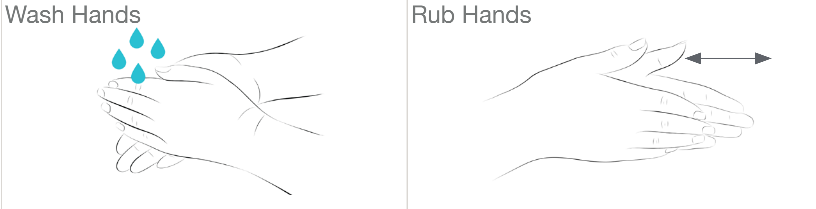


**C. Prick the finger.**

- Twist and pull the protective cap away from the lancet. Set the cap down.
- Hold the lancet between the fingers of one hand, positioning its tip on the puncture site of the other hand (parents/guardians: hold the tip of your child's finger to be pricked).
- Press the lancet firmly against the puncture site. Do not remove it until you hear a click.
- Wipe off the first drop of blood with sterile gauze.

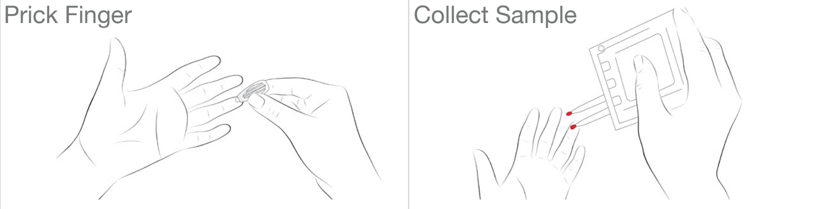


**D. Collect the sample using the cartridge.**

- With your finger facing up, touch the sampling tip to the blood drop. Let the tip turn fully red, count two seconds, and remove. Repeat for the second sampling tip. It is OK to apply tips to blood many times to fill.

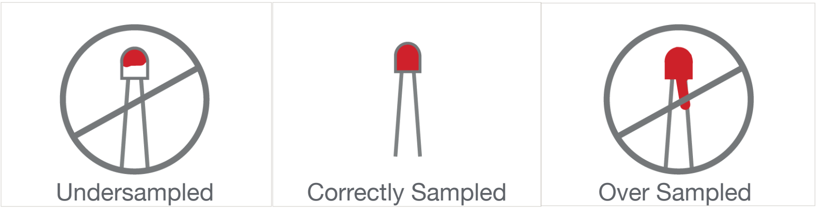


- Close the cartridge: lift both wings of the cartridge upwards to meet at the top. Press the tabs together until you hear a click.

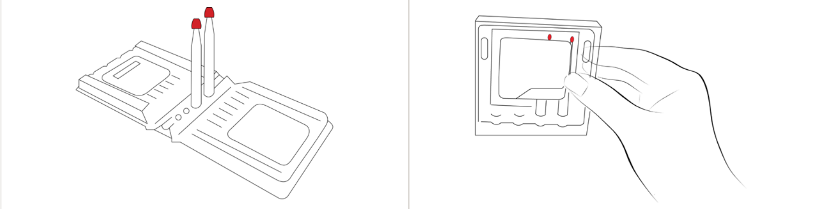

**E. Collect the sample on the blood spot card.**

- Allow a large blood drop to form. It may help to gently massage the base of the finger to stimulate blood flow.

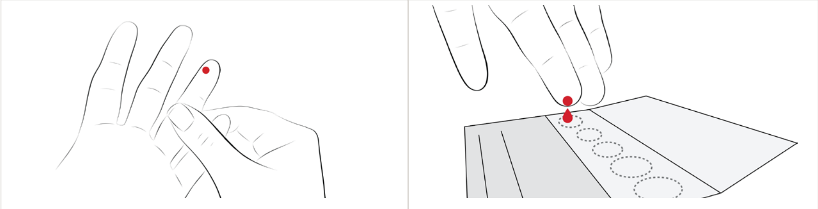

- Gently touch the blood drop to the center of the first circle on the card. Allow the blood to soak in and COMPLETELY fill the circle (do not let your finger touch the card).
- Move to the next circle and repeat until you have filled all 5 circles (or as many as you can). Three fully filled circles are better than 5 incomplete circles. If your blood stops flowing before at least 3 circles are filled, use the second lancet to prick a different finger.


Child-V3.indd 2 1/27/21 2:21 PM
